# Genome-Wide Meta-Analysis of Late-Onset Alzheimer’s Disease Using Rare Variant Imputation in 65,602 Subjects Identifies Novel Rare Variant Locus *NCK2*: The International Genomics of Alzheimer’s Project (IGAP)

**DOI:** 10.1101/2021.03.14.21253553

**Authors:** Adam C. Naj, Ganna Leonenko, Xueqiu Jian, Benjamin Grenier-Boley, Maria Carolina Dalmasso, Celine Bellenguez, Jin Sha, Yi Zhao, Sven J. van der Lee, Rebecca Sims, Vincent Chouraki, Joshua C. Bis, Brian W. Kunkle, Peter Holmans, Yuk Yee Leung, John J. Farrell, Alessandra Chesi, Hung-Hsin Chen, Badri Vardarajan, Penelope Benchek, Sandral Barral, Chien-Yueh Lee, Pavel Kuksa, Jacob Haut, Edward B. Lee, Mingyao Li, Yuanchao Zhang, Struan Grant, Jennifer E. Phillips-Cremins, Hata Comic, Achilleas Pitsillides, Rui Xia, Kara L. Hamilton-Nelson, Amanda Kuzma, Otto Valladares, Brian Fulton-Howard, Josee Dupuis, Will S. Bush, Li-San Wang, Jennifer E. Below, Lindsay A. Farrer, Cornelia van Duijn, Richard Mayeux, Jonathan L. Haines, Anita L. DeStefano, Margaret A. Pericak-Vance, Alfredo Ramirez, Sudha Seshadri, Philippe Amouyel, Julie Williams, Jean-Charles Lambert, Gerard D. Schellenberg

**Affiliations:** Department of Biostatistics, Epidemiology, and Informatics, Perelman School of Medicine, University of Pennsylvania, Philadelphia, PA, USA; Department of Pathology and Laboratory Medicine, Perelman School of Medicine, University of Pennsylvania, Philadelphia, PA, USA; Division of Psychological Medicine and Clinical Neurosciences, School of Medicine, Cardiff University, Wales, UK; Brown Foundation Institute of Molecular Medicine, University of Texas Health Sciences Center at Houston, Houston, TX, USA; Inserm, CHU Lille, Institut Pasteur de Lille, U1167-RID-AGE Facteurs de risque et déterminants moléculaires des maladies liées au vieillissement, University of Lille, Lille, France; Department of Psychiatry & Psychotherapy, University of Cologne, Cologne, Germany; Alzheimer Center, VU University Medical Center, Amsterdam, the Netherlands; Dementia Research Institute, School of Medicine, Cardiff University, Wales, UK; Cardiovascular Health Research Unit, Department of Medicine, University of Washington, Seattle, WA, USA; John P. Hussman Institute for Human Genomics, Miller School of Medicine, University of Miami, Miami, FL, USA; Dr. John T. Macdonald Foundation, Department of Human Genetics, Miller School of Medicine, University of Miami, Miami, FL, USA; Department of Medicine (Biomedical Genetics), Boston University School of Medicine, Boston, MA, USA; Vanderbilt Genetics Institute, Vanderbilt University Medical Center, Nashville, TN, USA; Taub Institute on Alzheimer’s Disease and the Aging Brain, Department of Neurology; Gertrude H. Sergievsky Center; and Department of Neurology, Columbia University, New York, NY, USA; Department of Population & Quantitative Health Sciences, School of Medicine, Case Western Reserve University, Cleveland, OH, USA; Translational Neuropathology Research Laboratory, Department of Pathology and Laboratory Medicine, Perelman School of Medicine, University of Pennsylvania, Philadelphia, PA, USA; Division of Human Genetics, The Children’s Hospital of Philadelphia, 3615 Civic Center Boulevard, Philadelphia, PA, USA; Department of Genetics, Perelman School of Medicine, University of Pennsylvania, 3615 Civic Center Boulevard, Philadelphia, PA, USA; Department of Bioengineering, University of Pennsylvania, Philadelphia, PA, USA; Department of Epidemiology, Erasmus Medical Center, Rotterdam, the Netherlands; Department of Biostatistics, School of Public Health, Boston University, Boston, MA, USA; Imm Center for Human Genetics, University of Texas Health Sciences Center at Houston, Houston, TX, USA; Ronald M. Loeb Center for Alzheimer’s Disease, Mt. Sinai Icahn School od Medicine, New York, NY, USA; Department of Neurology, School of Medicine, Boston University, Boston, MA, USA; Nuffield Department of Population Health, St. Cross College, Oxford University, Oxford, England, UK; Glenn Biggs Institute for Alzheimer’s & Neurodegenerative Diseases, University of Texas Health Sciences Center, San Antonio, TX, USA

**Author notes:** (*Group authorship currently under revision*.). On behalf of the International Genomics of Alzheimer’s Project (IGAP). Co-first authors (Adam C. Naj, Ganna Leonenko, Xueqiu Jian, Benjamin Grenier-Boley, Maria Carolina Dalmasso, Celine Bellenguez). Co-senior authors (Alfredo Ramirez, Sudha Seshadri, Philippe Amouyel, Julie Williams, Jean-Charles Lambert, Gerard D. Schellenberg). Corresponding author: **Address correspondence to:** Adam Naj, PhD, Assistant Professor of Epidemiology, Department of Biostatistics, Epidemiology, and Informatics (DBEI) and Department of Pathology and Laboratory Medicine, University of Pennsylvania Perelman School of Medicine, 229 Blockley Hall, 423 Guardian Drive, Philadelphia, PA 19107.

**Keywords:** Genome-wide association studies (GWAS), GWAS Meta-Analysis, Late-Onset Alzheimer Disease (LOAD), functional genomics

## Abstract

Risk for late-onset Alzheimer’s disease (LOAD) is driven by multiple loci primarily identified by genome-wide association studies, many of which are common variants with minor allele frequencies (MAF)> 0.01. To identify additional common and rare LOAD risk variants, we performed a GWAS on 25,170 LOAD subjects and 41,052 cognitively normal controls in 44 datasets from the International Genomics of Alzheimer’s Project (IGAP). Existing genotype data was imputed using the dense, high-resolution Haplotype Reference Consortium (HRC) r1.1 reference panel. Stage 1 associations of *P*<10^−5^ were meta-analyzed with the European Alzheimer’s Disease Biobank (EADB) (n=20,301 cases; 21,839 controls) (stage 2 combined IGAP and EADB). An expanded meta-analysis was performed using a GWAS of parental AD/dementia history in the UK Biobank (UKBB) (n=35,214 cases; 180,791 controls) (stage 3 combined IGAP, EADB, and UKBB). Common variant (MAF≥0.01) associations were identified for 29 loci in stage 2, including novel genome-wide significant associations at *TSPAN14* (*P*=2.33×10^−12^), *SHARPIN* (*P*=1.56×10^−9^), and *ATF5/SIGLEC11* (*P*=1.03×10^−8^), and newly significant associations without using AD proxy cases in *MTSS1L/IL34* (*P*=1.80×10^−8^), *APH1B* (*P*=2.10×10^−13^), and *CLNK* (*P*=2.24×10^−10^). Rare variant (MAF<0.01) associations with genome-wide significance in stage 2 included multiple variants in *APOE* and *TREM2*, and a novel association of a rare variant (rs143080277; MAF=0.0054; *P*=2.69×10^−9^) in *NCK2*, further strengthened with the inclusion of UKBB data in stage 3 (*P*=7.17×10^−13^). Single-nucleus sequence data shows that *NCK2* is highly expressed in amyloid-responsive microglial cells, suggesting a role in LOAD pathology.

## Introduction

Alzheimer’s disease (AD) is the most common neurodegenerative disease with an estimated 50 million persons affected globally. With the prevalence predicted to triple by 2050, AD is an intensifying public health crisis requiring insight into key genetic and environmental risk contributors to guide the development of therapeutic interventions to prevent and treat the disease. The common form of the disease, late-onset AD (LOAD) accounts for ∼ 95% of all cases and is highly heritable (*h*^2^∼60-80%)^1^ and polygenic^2^. Previous genetic studies of LOAD including genome-wide association studies^3–16^ and more recently sequencing studies^17–23^ yielded confirmed associations at nearly 30 distinct loci^24, 25^, including many common variants (minor allele frequency [MAF]>0.01) and more recently rare variants (MAF≤0.01) in *APP*^26^, *TREM2*^27, 28^, *PLCG2*^11^, *ABI3*^11^, *SORL1*^17, 29^, and *ABCA7*^30–32^. While whole genome sequencing (WGS) studies can identify novel rare variants and private mutations in families enriched for disease^33, 34^, current LOAD WGS study samples sizes are much smaller than available GWAS samples^34^, limiting statistical power to detect genome-wide significant associations at many rare variants^35^. High-resolution imputation of rare and low frequency variation in large GWAS studies is a cost-effective alternative to *de novo* sequencing for identifying rare LOAD loci^36^.

This multi-stage LOAD GWAS incorporated genotype data from 109,973 cases and 285,418 controls from the International Genomics of Alzheimer’s Project (IGAP; Stage 1), European Alzheimer’s Disease Biobank^37^ (EADB; IGAP+EADB, Stage 2), and the UK Biobank (UKBB: IGAP+EADB+UKBB, Stage 3), imputed using the Haplotype Reference Consortium (HRC) r1.1 (IGAP and UKBB) or the Trans-Omics for Precision Medicine (TOPMed) Freeze 5 panels (EADB). We also explored the association and potential functional roles of these imputed variants with function-informed analyses including genetically-regulated gene expression and co-expression analyses. Furthermore, we applied functional genomics approaches to characterize potential roles of identified variants at the chromatin level and on the cellular level in AD pathogenesis.

## Results

### Study Design and Participant Characteristics

We conducted a discovery meta-analysis (Stage 1) of 44 GWAS datasets of non-Hispanic Whites (NHW) subjects [n= 25,170 cases; 41,052 cognitively normal elders] in the IGAP. IGAP comprises data from five consortia including the Alzheimer’s Disease Genetics Consortium (ADGC), the Cohorts for Heart and Aging Research in Genomic Epidemiology Consortium (CHARGE), the European Alzheimer’s Disease Initiative (EADI), and Genetic and Environmental Risk in AD/Defining Genetic, Polygenic and Environmental Risk for Alzheimer’s Disease Consortium (GERAD/PERADES) and a dataset from the University of Bonn (BONN) (cohort details in the **Supplementary Note** and **Supplementary Table 1**). We validated Stage 1 suggestive associations of *P*<10^−5^ in IGAP by meta-analysis with association results from the EADB [n= 20,301 cases; 21,839 cognitively normal elders] (Stage 2). Additionally, we performed secondary replication in an AD/dementia family history GWAS in the UKBB using 35,214 ‘proxy cases’ (defined as individuals reporting a history of AD/dementia in one or both parents and with age≥65 years) and 180,791 ‘proxy controls’ (individuals reporting no history of AD/dementia in either biological parent and with age≥65 years). Stage 3 was a meta-analysis of IGAP, EADB, and UKBB findings for variants associated with *P*<10^−5^ in IGAP. Analytic methods for the EADB replication and the UKBB secondary replication dataset are also in the **Supplementary Note**. The structure of the analyses is depicted in **Figure 1**.

**Figure 1.**
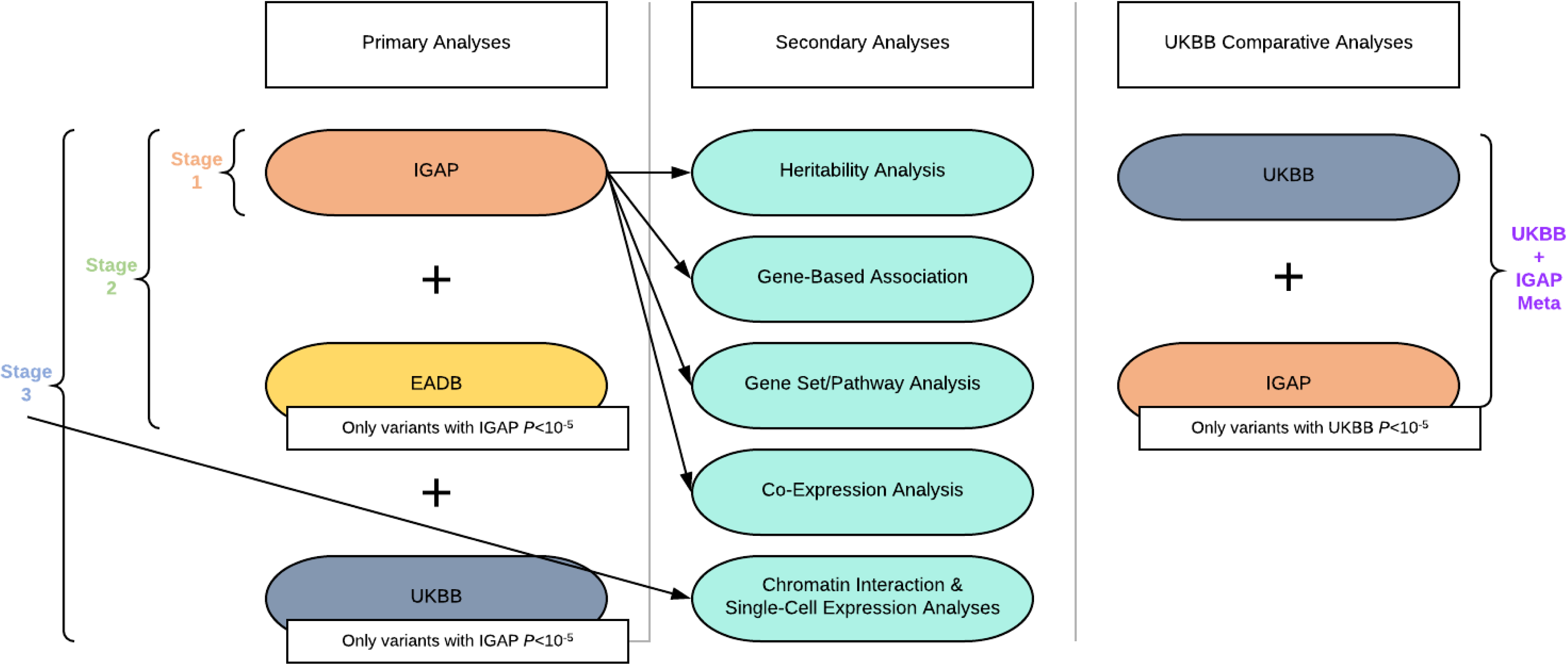
A diagram of the IGAP study design. Stage 1 included HRC-imputed data in ADGC, GERAD/PERADES, EADI, CHARGE, and Bonn cohorts. Stage 2 and Stage 3 variants were selected from variants with Stage 1 associations of *P*<10^−5^. Stage 2 genotyping was performed by EADB and collaborators and genotype data were imputed to the TOPMed Freeze 5 haplotype reference panel. Stage 3 genotyping was performed by the UKBB and genotype data were imputed to the HRC + UK10K reference panel. Secondary analysis depicted included multiple follow-up analyses of variants with GWS associations, as well as a discovery analysis of UKBB for comparison against discovery-level associations in IGAP.

IGAP, EADB, and UKBB all included a similar proportion of more females than males (IGAP, 60.3% female; EADB, 59.4%; UKBB, 55.2%). Mean ages-at-onset among LOAD cases were 74.6 and 71.1 years for IGAP and EADB, respectively, with a higher onset age for the CHARGE cohorts (mean ± SD: 83.6±1.1). Average ages-at-last-exam were later among IGAP cognitively normal controls (mean ± SD: 77.4±6.4) than EADB (mean ± SD: 66.9±14.3). *APOE* ε4 frequencies in IGAP were 35.1% and 13.2% among cases and controls, respectively, while in EADB, they were similarly 32.6% among cases and 13.2% among controls.

### Association Analysis of Common Variants (MAF>0.01)

In Stage 1 (IGAP analyses), the strongest genome-wide association was observed as expected in the region around *APOE* on chromosome 19q13.32 (rs7259620; *P*=2.25×10^−1^^99^; **Table 1**, **Supplementary Table 2**, **Supplementary Figures 1** and **4**). Fifteen loci outside of the *APOE* region yielded associations with genome-wide statistical significance (*P*<5×10^−8^; GWS) in Stage 1 analyses, including variants at loci observed in prior GWAS: *BIN1*, *PICALM*, *CR1*, the *MS4A* region, *CLU*, *SORL1*, *CD2AP*, *ABCA7*, *USP6NL/ECHDC3*, *SLC24A4*, *SPI1*, *PTK2B*, *IQCK*, and *NYAP1*. Two loci newly significant in IGAP were *TSPOASP1-AS1* on chromosome 17q22 (rs2526377, *P*=5.90×10^−9^) and *HBEGF* on chromosome 5q31.3 (rs11168036, *P*=1.95×10^−8^).

**Table 1.** Strongest regional common variant (MAF≥0.01) association results in IGAP (Stage 1), their associations in EADB, and combined analysis of IGAP and EADB (Stage 2)

| Variant RSID | Chr | Basepair (hg19) | Genomic Region | EFF | REF | Stage 1: IGAP |  |  | EADB |  |  | Stage 2: IGAP + EADB Meta-Analysis |  |  |
| --- | --- | --- | --- | --- | --- | --- | --- | --- | --- | --- | --- | --- | --- | --- |
|  |  |  |  |  |  | EAF | OR (95% CI) | P | EAF | OR (95% CI) | P | EAF | OR (95% CI) | P |
| rs7259620 | 19 | 45407788 | *APOE* Region* | A | G | 0.4397 | 0.67 (0.65, 0.69) | 2.25E-199 | 0.4197 | 0.64 (0.62, 0.66) | 1.20E-210 | 0.4308 | 0.66 (0.64, 0.67) | 1.15E-401 |
| rs6733839 | 2 | 127892810 | *BINI* * | T | C | 0.3836 | 1.18 (1.15, 1.21) | 2.36E-35 | 0.3991 | 1.21 (1.18, 1.25) | 5.12E-40 | 0.3906 | 1.19 (1.17, 1.22) | 3.06E-73 |
| rs7110631 | 11 | 85856187 | *PICALM* * | C | G | 0.318 | 0.88 (0.86, 0.90) | 1.42E-20 | 0.3065 | 0.88 (0.85, 0.91) | 2.77E-16 | 0.3130 | 0.88 (0.86, 0.90) | 3.33E-35 |
| rs679515 | 1 | 207750568 | *CR1* * | T | C | 0.1976 | 1.15 (1.12, 1.19) | 5.69E-19 | 0.2030 | 1.16 (1.12, 1.20) | 4.49E-17 | 0.2000 | 1.16 (1.13, 1.18) | 2.62E-34 |
| rs1582763 | 11 | 60021948 | *MS4A* Region* | A | G | 0.383 | 0.89 (0.87, 0.92) | 3.63E-18 | 0.3597 | 0.90 (0.87, 0.92) | 1.52E-13 | 0.3729 | 0.89 (0.88, 0.91) | 4.08E-30 |
| rs867230 | 8 | 27468503 | *CLU* * | C | A | 0.4052 | 0.89 (0.87, 0.92) | 1.34E-17 | 0.3875 | 0.90 (0.87, 0.92) | 2.75E-13 | 0.3973 | 0.90 (0.88, 0.91) | 3.23E-29 |
| rs11218343 | 11 | 121435587 | *SORL1* * | C | T | 0.0424 | 0.80 (0.75, 0.85) | 1.10E-11 | 0.0391 | 0.86 (0.80, 0.92) | 4.53E-5 | 0.0409 | 0.83 (0.79, 0.87) | 6.69E-15 |
| rs1385742 | 6 | 47595155 | *CD2AP* * | A | T | 0.3626 | 1.09 (1.06, 1.12) | 7.41E-11 | 0.3577 | 1.06 (1.03, 1.10) | 7.27E-5 | 0.3605 | 1.08 (1.06, 1.10) | 5.37E-14 |
| rs12151021 | 19 | 1050874 | *ABCA7* * | A | G | 0.3289 | 1.10 (1.07, 1.13) | 7.46E-11 | 0.3306 | 1.15 (1.12, 1.18) | 8.98E-20 | 0.3297 | 1.12 (1.10, 1.14) | 5.24E-28 |
| rs7920721 | 10 | 11720308 | *USP6NL/ ECHDC3* * | G | A | 0.3837 | 1.09 (1.06, 1.11) | 4.59E-10 | 0.3908 | 1.07 (1.03, 1.10) | 1.90E-5 | 0.3869 | 1.08 (1.06, 1.10) | 7.11E-14 |
| rs9323877 | 14 | 92934269 | *SLC24A4* * | G | A | 0.2422 | 1.09 (1.06, 1.13) | 1.01E-9 | 0.2531 | 1.07 (1.04, 1.11) | 1.70E-5 | 0.2470 | 1.08 (1.06, 1.11) | 1.28E-13 |
| rs7928419 | 11 | 47392114 | *SPII* * | G | A | 0.3399 | 0.92 (0.90, 0.95) | 1.74E-9 | 0.3330 | 0.95 (0.92, 0.98) | 0.00143 | 0.3369 | 0.94 (0.92, 0.95) | 3.52E-11 |
| rs2526377 | 17 | 56410041 | *BZRAP1-AS1 (TSPOAPI-AS1)* | G | A | 0.4479 | 0.93 (0.90, 0.95) | 5.90E-9 | 0.4465 | 0.94 (0.91, 0.96) | 1.07E-5 | 0.4473 | 0.93 (0.91, 0.95) | 3.87E-13 |
| rs11168036 | 5 | 139707439 | *HBEGF* | T | G | 0.4915 | 1.07 (1.05, 1.10) | 1.95E-8 | 0.4873 | 0.99 (0.96, 1.01) | 0.331 | 0.4996 | 0.95 (0.94, 0.97) | 8.96E-07 |
| rs73223431 | 8 | 27219987 | *PTK2B* * | T | C | 0.3596 | 1.08 (1.05, 1.10) | 2.31E-8 | 0.3682 | 1.07 (1.04, 1.11) | 1.198E-6 | 0.3634 | 1.08 (1.06, 1.10) | 1.44E-13 |
| rs114285994 | 16 | 19935763 | *IQCK* * | A | G | 0.1406 | 0.90 (0.87, 0.94) | 3.53E-8 | 0.1223 | 0.98 (0.94, 1.02) | 0.353 | 0.133 | 0.93 (0.91, 0.96) | 1.54E-6 |
| rs12539172 | 7 | 100091795 | *NYAP1* * | T | C | 0.3135 | 0.93 (0.90, 0.95) | 1.32E-7 | 0.3100 | 0.92 (0.89, 0.95) | 5.243E-7 | 0.312 | 0.93 (0.91, 0.95) | 3.594E-13 |
| rs3135348 | 6 | 32394098 | *HLA-DRB1/5* * | A | G | 0.4248 | 1.07 (1.04, 1.10) | 2.25E-7 | NA | NA | NA | 0.4248 | 1.07 (1.04, 1.10) | 2.11E-7 |
| rs10753507 | 1 | 21152380 | *EIF4G3* | T | A | 0.4038 | 0.93 (0.91, 0.96) | 2.72E-7 | 0.4013 | 0.97 (0.94, 1.00) | 0.0229 | 0.4027 | 0.95 (0.93, 0.97) | 9.47E-8 |
| rs11735125 | 4 | 66237551 | *EPHA5* | G | C | 0.0225 | 1.24 (1.14, 1.34) | 3.68E-7 | 0.0243 | 1.00 (0.91, 1.09) | 0.991 | 0.0233 | 1.12 (1.06, 1.19) | 1.51E-4 |
| rs17462136 | 20 | 54987216 | *CASS4* * | C | G | 0.0919 | 0.89 (0.85, 0.93) | 3.92E-7 | 0.0800 | 0.83 (0.78, 0.87) | 1.15E-12 | 0.0870 | 0.86 (0.83, 0.89) | 2.91E-17 |
| rs8107367 | 19 | 18564705 | *ELL* | G | A | 0.3337 | 1.07 (1.04, 1.10) | 5.55E-7 | 0.3319 | 1.01 (0.98, 1.04) | 0.483 | 0.3329 | 1.04 (1.02, 1.07) | 2.15E-5 |
| rs1943782 | 11 | 102357848 | *LOC102723838* | A | G | 0.0845 | 1.12 (1.07, 1.17) | 7.39E-7 | 0.0860 | 1.02 (0.97, 1.08) | 0.362 | 0.0851 | 1.08 (1.04, 1.12) | 1.50E-5 |
| rs140016620 | 16 | 70713787 | *MTSSL1/IL34* | G | A | 0.0653 | 1.14 (1.08, 1.19) | 9.31E-7 | 0.0628 | 1.09 (1.03, 1.16) | 0.00345 | 0.0642 | 1.12 (1.07, 1.16) | 1.80E-8 |
| rs56402156 | 7 | 143103481 | *EPHA1* * | A | G | 0.2002 | 0.93 (0.90, 0.95) | 1.26E-6 | 0.1838 | 0.92 (0.89, 0.96) | 9.71E-6 | 0.1932 | 0.92 (0.90, 0.95) | 5.72E-11 |
| rs1001158 | 4 | 11038456 | *CLNK* | G | A | 0.2825 | 1.07 (1.04, 1.10) | 2.03E-6 | 0.2841 | 1.07 (1.04, 1.10) | 2.63E-5 | 0.2832 | 1.07 (1.05, 1.09) | 2.24E-10 |
| rs10858815 | 12 | 89391846 | *LOC728084/ LINC02458* | C | A | 0.0411 | 0.85 (0.80, 0.91) | 2.07E-6 | 0.0424 | 0.99 (0.92, 1.06) | 0.766 | 0.0417 | 0.91 (0.87, 0.96) | 2.28E-4 |
| rs3896609 | 15 | 51057868 | *SPPL2A* * | A | C | 0.1855 | 0.92 (0.89, 0.95) | 2.11E-6 | 0.1863 | 0.98 (0.94, 1.01) | 0.181 | 0.1859 | 0.95 (0.92, 0.97) | 1.02E-5 |
| rs72985631 | 2 | 232829502 | *DIS3L2/ INPP5D* * | A | G | 0.0242 | 0.82 (0.76, 0.89) | 3.01E-6 | 0.0211 | 0.98 (0.89, 1.08) | 0.694 | 0.0229 | 0.88 (0.83, 0.94) | 1.34E-5 |
| rs12580654 | 12 | 52268547 | *ANKRD33* | C | G | 0.1061 | 1.10 (1.06, 1.15) | 3.48E-6 | 0.1034 | 1.02 (0.98, 1.07) | 0.352 | 0.1049 | 1.07 (1.03, 1.10) | 4.56E-5 |
| rs143867193 | 17 | 61503610 | *TANC2/ACE* * | T | C | 0.0163 | 1.27 (1.15, 1.40) | 3.56E-6 | 0.0146 | 1.10 (0.98, 1.24) | 0.0923 | 0.0156 | 1.19 (1.11, 1.29) | 3.91E-6 |
| rs6966331 | 7 | 37883793 | *NME8* * | T | C | 0.3523 | 0.94 (0.92, 0.97) | 6.09E-6 | 0.3480 | 0.94 (0.91, 0.97) | 2.88E-5 | 0.3504 | 0.94 (0.92, 0.96) | 7.14E-10 |
| rs11028038 | 11 | 24605495 | *LUZP2* | C | T | 0.2165 | 1.07 (1.04, 1.10) | 7.29E-6 | 0.2143 | 0.97 (0.94, 1.00) | 0.0828 | 0.2156 | 1.03 (1.00, 1.05) | 0.0270 |
| rs11236918 | 11 | 76448286 | *GUCY2EP/ TSKU* | A | C | 0.1122 | 1.09 (1.05, 1.13) | 8.13E-6 | 0.1150 | 1.01 (0.96, 1.05) | 0.78793 | 0.1134 | 1.05 (1.02, 1.09) | 3.97E-4 |
| rs9960448 | 18 | 57986377 | *MC4R* | T | G | 0.2904 | 0.94 (0.91, 0.97) | 8.24E-6 | 0.2773 | 1.00 (0.97, 1.03) | 0.9519 | 0.2848 | 0.96 (0.94, 0.98) | 6.46E-4 |

Abbreviations: Chr, chromosome; EFF, Effect allele; REF, reference allele; EAF, effect allele frequency; OR, Odds ratio; CI, Confidence interval; *P*, P-value
Shaded box indicates genome-wide significance within analysis stage
\* Loci with prior associations in IGAP [Lambert et al. (2013) or Kunkle et al. (2019)]

For Stage 2, we meta-analyzed data on all variants in IGAP meeting our discovery threshold (*P*<10^−5^; 3,345 variants) in Stage 1 with associations of those variants in EADB. Although the most significantly associated variants changed at many loci, Stage 2 GWS associations (**Table 2**, **Supplementary Table 2**) included all Stage 1 GWS loci. GWS associations were observed in Stage 2 at known AD loci *ADAMTS1*, *EPHA1*, *HLA-DRB1/5*, *SPI1*, *SORL1*, *IQCK*, *NME8*, *ACE*, and *UNC5CL/OARD1*, and at several novel loci, including *TSPAN14* (rs6586028, 2.33×10^−12^), *SHARPIN* (rs34173062, *P*=1.56×10^−9^), and *ATF5*/*SIGLEC11* (rs875121, *P*=1.03×10^−8^). Several loci that had been observed in studies mixing proxy cases and controls with clinically- and pathologically-defined cases and controls were now observed to be associated in data without proxy cases/controls, including *APH1B* (rs117618017, *P*=2.10×10^−13^), *CLNK* (rs2904297, *P*=1.20×10^−10^), and *MTSS1L/IL34* (rs11538963, *P*=7.80×10^−9^),

**Table 2.**
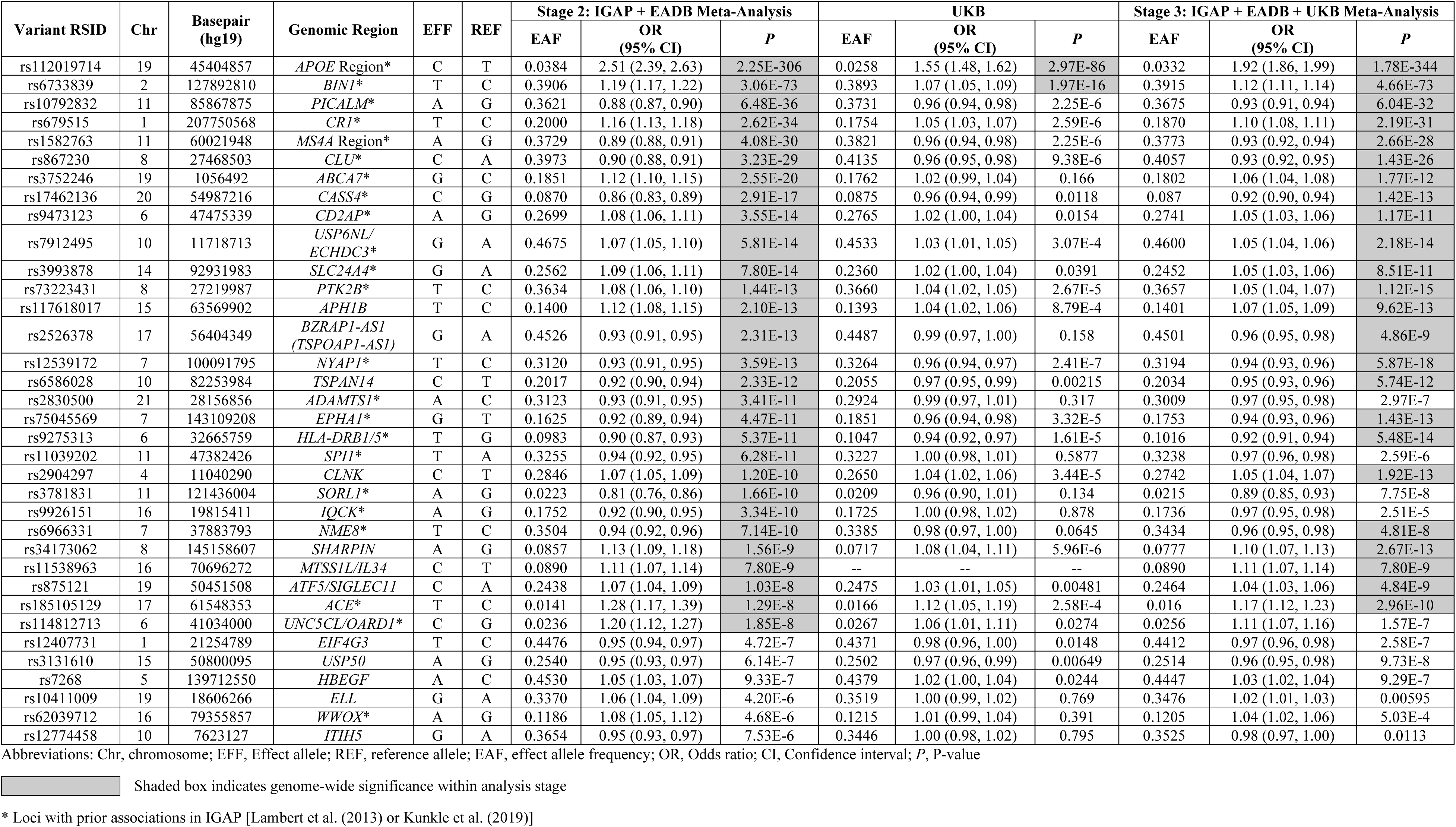
Strongest common variant (MAF≥0.01) association results by genomic region in Stage 2 and Stage 3. Variants depicted had *P*<10^−5^ in IGAP (Stage 1) analysis (n=3,345 variants) and had the strongest regional association in meta-analysis of IGAP+EADB (Stage 2). UKBB AD-proxy association results and meta-analysis of IGAP, EADB, and UKBB (Stage 3) are also shown.

For stage 3, we meta-analyzed IGAP and EADB with the UKBB parental history GWAS (**Table 2**, **Supplementary Table 2**). Although parental history of dementia/AD is predictive of dementia in later life^38–40^, the lower precision of this phenotype as a proxy for the clinical/neuropathologic AD phenotypes in IGAP and EADB likely resulted in more heterogeneity and smaller effect sizes among the associations observed in UKBB compared to IGAP and EADB without applying any corrective factors. We considered an association replicated in the UKBB data if there was nominal statistical significance (*P*<0.05) and the effect direction was consistent with Stage 1 and 2 results. Replication was obtained for associations in the *APOE* Region, *BIN1*, *PICALM*, *CR1*, the *MS4A* region, *CLU*, *CD2AP*, *ABCA7*, *USP6NL/ECHDC3*, *SLC24A4*, *TSPOAP1-AS1*, *PTK2B*, *NYAP1*, *CASS4*, *EPHA1*, *NME8*, *ACE*, and *UNC5CL*/*OARD1*. Several of the novel loci demonstrated GWS association in Stage 3, including *TSPAN14* (rs6586028, *P*=5.74×10^−12^), *CLNK* (rs2904297, *P*=1.92×10^−13^), *SHARPIN* (rs34173062, *P*=2.67×10^−13^), *APH1B* (rs117618017, *P*=9.62×10^−13^), and *ATF5/SIGLEC11* (rs3896609, *P*=4.84×10^−9^).

We also analyzed the UKBB data alone as a GWAS of parental history of AD/dementia to compare patterns of association between AD and AD proxy phenotypes. GWS common variant associations (**Supplementary Tables 3** and **4**) included signals in or around known susceptibility loci such as *APOE*, *BIN1*, *NYAP1*/*PILRB*, and *HLA-DRB1*, as well as an association near *VKORC1*/*BCKDK* (*P*=3.82×10^−8^), which was observed in the Schwarzentruber AD family history analysis^41^ and reported as *KAT8* in the Jansen et al. study^13^ which also included UKBB AD/dementia family history data. Discovery-level associations (*P*<10^−5^) were observed for several known AD susceptibility loci, including *ABCA7*, *PICALM*, *MS4A* Region, *ADAM10*/*MINDY2*, *EPHA1*, and *CR1*. We followed up UKBB associations with *P*<10^−5^ by meta-analysis with IGAP data and identified a GWS association at *EPHX2* (rs7341557, *P*=5.33×10^−11^) (**Supplementary Tables 3** and **4**), although it is unclear if this is independent of common variant associations at proximal loci *CLU* and *PTK2B*.

### Association Analysis of Rare Variants (MAF≤0.01)

Stage 1 analysis of rare variants (**Table 3**, **Supplementary Table 5**, **Supplementary Figures 2** and **5**) identified 22 variants at 12 distinct genomic regions with associations of discovery-level significance (*P*<10^−5^) for further follow-up. In addition to multiple associations in the *APOE* region and in/around *TREM2*, novel signals attaining discovery-level significance were present in/around the genes *RORA*, *NCK2*, *LINC02033/TRANK1*, *DYTN/MDH1B*, *ZBTB10/ZNF704*, *CNTNAP4*, *ODC1/NOL10*, *LINC00320*, *EPHA3*, and *CCDC102B*, though only *APOE* region and *TREM2* were GWS.

**Figure 2.**
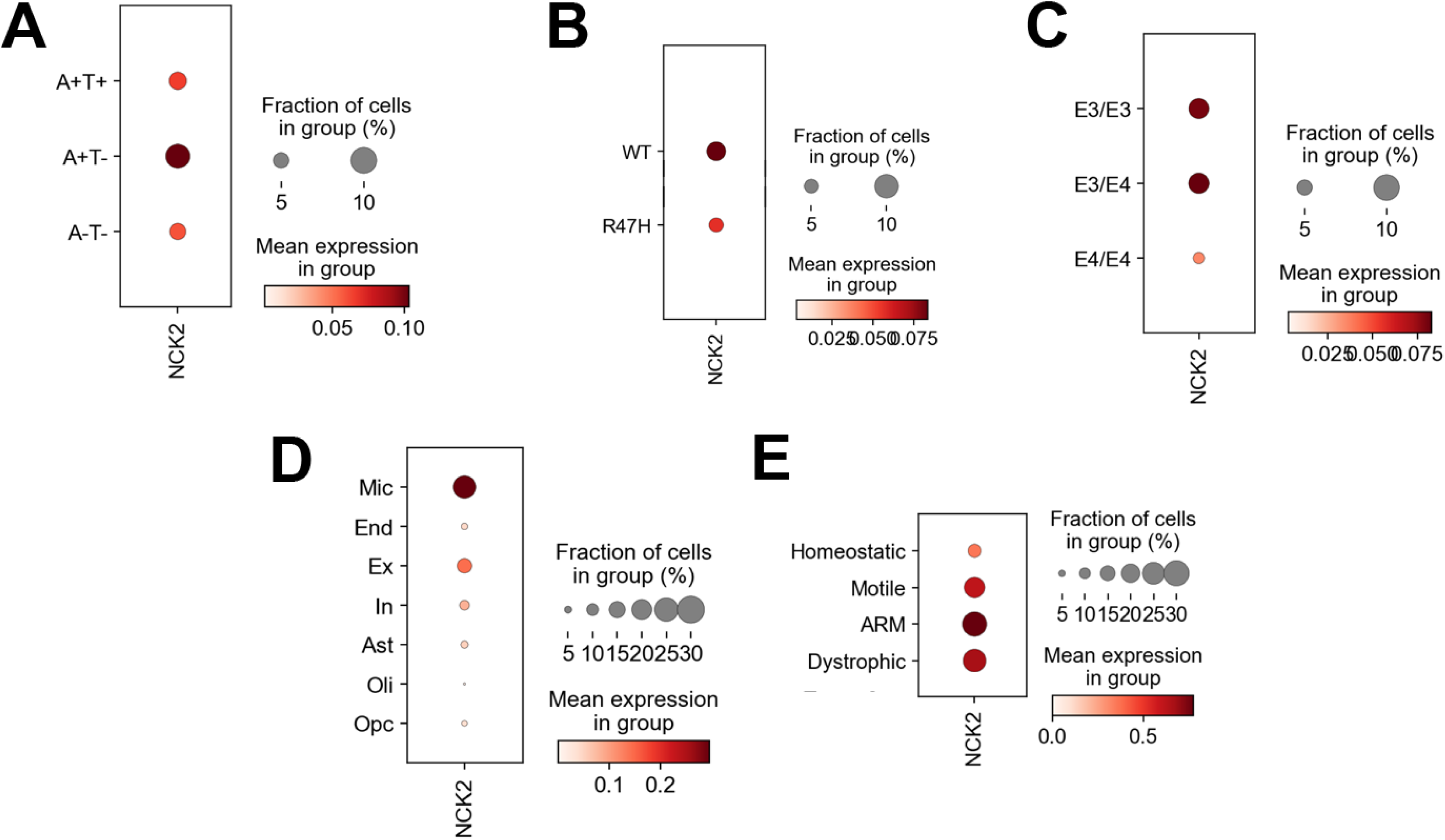
*NCK2* expression was analyzed using a single nucleus RNA-sequencing dataset consisting of human cortex samples that included normal tissue without amyloid or tau aggregates (A-T-), tissue with amyloid only (A+T-) and tissue with both amyloid and tau aggregates (A+T+). *NCK2* expression was highest in amyloid only brain tissues (panel **A**). Moreover, *NCK2* expression was reduced in cases with *TREM2* R47H and *APOE* ε4 risk genotypes (panels **B** and **C**), reminiscent of amyloid-responsive microglia. Expression analysis grouped by cell type showed that *NCK2* expression was highest in microglia (Mic, panel **D**) compared to endothelial cells (End), excitatory neurons (Ex), inhibitory neurons (In), astrocytes (Ast), oligodendrocytes (Oli) and oligodendroglial precursor cells (Opc). Within microglial subclusters, *NCK2* expression was highest in amyloid responsive microglia (ARM, panel **E**).

**Table 3.** All strongest rare variant (MAF<0.01) association results in IGAP (Stage 1) [no filtering on LD], their associations in EADB, and combined analysis of IGAP and EADB (Stage 2)

| Variant RSID | Chr | Basepair (hg19) | Genomic Region | EFF | REF | Stage 1: IGAP |  |  | EADB |  |  | Stage 2: IGAP + EADB Meta-Analysis |  |  |
| --- | --- | --- | --- | --- | --- | --- | --- | --- | --- | --- | --- | --- | --- | --- |
|  |  |  |  |  |  | EAF | OR (95% CI) | P | EAF | OR (95% CI) | P | EAF | OR (95% CI) | P |
| rs547509922 | 19 | 45331261 | *APOE* Region (intergenic) | T | C | 0.0048 | 2.80 (2.31, 3.40) | 1.12E-25 | 0.0053 | 2.26 (1.83, 2.78) | 2.01E-15 | 0.0051 | 2.54 (2.2, 2.92) | 5.04E-38 |
| rs559118614 | 19 | 45554058 | *APOE* Region (*CLASRP*) | G | T | 0.0094 | 2.04 (1.77, 2.35) | 1.88E-22 | 0.0097 | 2.42 (2.06, 2.84) | 5.64E-29 | 0.0095 | 2.2 (1.98, 2.44) | 1.83E-47 |
| rs75932628 | 6 | 41129252 | *TREM2* | T | C | 0.0043 | 2.45 (1.99, 3.02) | 3.72E-17 | 0.0028 | 2.14 (1.58, 2.89) | 2.97E-07 | 0.0038 | 2.34 (1.97, 2.78) | 2.15E-22 |
| rs187370608 | 6 | 40942196 | *TREM2* (upstream) | A | G | 0.0042 | 2.19 (1.78, 2.70) | 2.34E-13 | 0.0029 | 2.10 (1.57, 2.81) | 2.77E-07 | 0.0038 | 2.16 (1.82, 2.56) | 8.70E-19 |
| rs143202663 | 6 | 40865240 | *TREM2* (upstream) | C | T | 0.0040 | 2.11 (1.71, 2.61) | 2.59E-12 | 0.0027 | 2.18 (1.63, 2.90) | 4.41E-08 | 0.0036 | 2.14 (1.80, 2.53) | 1.83E-18 |
| rs145342536 | 6 | 40911888 | *TREM2* (upstream) | A | T | 0.0046 | 1.99 (1.64, 2.41) | 4.13E-12 | 0.0029 | 2.07 (1.56, 2.75) | 2.14E-07 | 0.0040 | 2.01 (1.71, 2.36) | 1.14E-17 |
| rs12664332 | 6 | 40904030 | *TREM2* (upstream) | A | G | 0.0045 | 1.95 (1.61, 2.37) | 1.57E-11 | 0.0029 | 2.10 (1.58, 2.79) | 1.31E-07 | 0.0040 | 2.00 (1.70, 2.34) | 2.98E-17 |
| rs112952132 | 19 | 45198060 | *APOE* Region (intergenic) | T | C | 0.0084 | 0.60 (0.52, 0.70) | 7.41E-11 | 0.0085 | 0.74 (0.63, 0.87) | 2.93E-4 | 0.0084 | 0.66 (0.59, 0.74) | 5.40E-13 |
| rs78905796 | 19 | 45132679 | *APOE* Region (*IGSF23*) | A | G | 0.0052 | 1.78 (1.48, 2.14) | 1.01E-9 | 0.0049 | 1.79 (1.44, 2.23) | 1.47E-7 | 0.005 | 1.78 (1.55, 2.05) | 1.13E-15 |
| rs540038005 | 15 | 61371352 | *RORA* | T | C | 0.00084 | 3.60 (2.27, 5.71) | 5.40E-8 | 0.00087 | 0.80 (0.49, 1.31) | 0.370 | 0.0009 | 1.79 (1.28, 2.51) | 7.25E-4 |
| rs551336410 | 15 | 61373788 | *RORA* | T | C | 0.00091 | 3.34 (2.14, 5.23) | 1.29E-7 | 0.00090 | 0.77 (0.47, 1.26) | 0.297 | 0.0009 | 1.71 (1.23, 2.38) | 0.00141 |
| rs192675224 | 6 | 40706366 | *TREM2* (upstream) | A | G | 0.0033 | 1.85 (1.46, 2.35) | 3.15E-7 | 0.0023 | 1.70 (1.26, 2.29) | 4.65E-4 | 0.0029 | 1.79 (1.49, 2.16) | 7.57E-10 |
| rs150085726 | 6 | 40729544 | *TREM2* (upstream) | A | G | 0.0033 | 1.86 (1.47, 2.37) | 3.24E-7 | 0.0022 | 1.76 (1.29, 2.40) | 2.91E-4 | 0.0029 | 1.82 (1.51, 2.20) | 4.84E-10 |
| rs143080277 | 2 | 106366056 | *NCK2* | C | T | 0.0052 | 1.65 (1.36, 2.01) | 3.66E-7 | 0.0056 | 1.32 (1.08, 1.61) | 0.00576 | 0.0054 | 1.48 (1.29, 1.70) | 2.69E-8 |
| rs17035636 | 3 | 36859000 | *LINC02033/TRANK1* (downstream) | C | T | 0.0016 | 0.43 (0.31, 0.62) | 2.64E-6 | 0.0026 | 0.85 (0.62, 1.16) | 0.302 | 0.0021 | 0.63 (0.5, 0.79) | 8.62E-5 |
| rs182464045 | 2 | 207595819 | *DYTN/MDH1B* | T | C | 0.0013 | 2.40 (1.66, 3.47) | 3.14E-6 | 0.0022 | 1.08 (0.79, 1.46) | 0.635 | 0.0018 | 1.49 (1.18, 1.89) | 8.55E-4 |
| rs78774825 | 8 | 81473635 | *ZBTB10/ZNF704* | T | G | 0.0025 | 1.90 (1.45, 2.50) | 3.81E-6 | 0.0035 | 0.97 (0.74, 1.27) | 0.831 | 0.003 | 1.36 (1.12, 1.65) | 0.00189 |
| rs114999466 | 16 | 76230037 | *CNTNAP4* (upstream) | G | A | 0.0069 | 0.69 (0.59, 0.81) | 3.91E-6 | 0.0085 | 0.82 (0.70, 0.96) | 0.0114 | 0.0077 | 0.75 (0.67, 0.84) | 4.51E-7 |
| rs552075630 | 2 | 10693329 | *ODCI/NOL10* | T | G | 0.0051 | 1.57 (1.29, 1.91) | 5.82E-6 | 0.0051 | 1.07 (0.86, 1.33) | 0.525 | 0.0051 | 1.33 (1.15, 1.53) | 1.44E-4 |
| rs112120395 | 21 | 21159556 | *LINC00320* | G | T | 0.0055 | 1.52 (1.27, 1.83) | 6.09E-6 | 0.0054 | 0.90 (0.74, 1.09) | 0.267 | 0.0055 | 1.19 (1.04, 1.36) | 0.0107 |
| rs527488596 | 3 | 90428380 | *EPHA3* (downstream) | A | T | 0.0055 | 0.65 (0.53, 0.78) | 6.32E-6 | NA | NA | NA | 0.0055 | 0.65 (0.53, 0.78) | 6.32E-6 |
| rs145472714 | 18 | 66411495 | *CCDC102B* | G | T | 0.0077 | 0.70 (0.60, 0.82) | 9.24E-6 | 0.0047 | 0.92 (0.73, 1.17) | 0.506 | 0.0068 | 0.76 (0.67, 0.87) | 5.46E-5 |

Abbreviations: Chr, chromosome; EFF, Effect allele; REF, reference allele; EAF, effect allele frequency; OR, Odds ratio; CI, Confidence interval; *P*, P-value
Shaded box indicates genome-wide significance within analysis stage

Stage 2 associations (**Table 4**, **Supplementary Table 5**) meta-analyzing IGAP and EADB revealed one novel GWS association at the chromosome 2q12.2 gene *NCK2* (rs143080277; Stage 2: MAF=0.0054, OR[95% CI]: 1.48 [1.29, 1.70], *P*=2.69×10^−8^; IGAP: MAF=0.0052, OR[95% CI]: 1.65 [1.36, 2.01], *P*=3.66×10^−7^; EADB: MAF=0.0056, OR[95% CI]: 1.32 [1.08, 1.61], *P*=0.00576).

**Table 4.** Strongest rare variant (MAF<0.01) association results by genomic region in Stage 2 and Stage 3. Variants depicted had *P*<10^−5^ in IGAP (Stage 1) analysis (n=372 variants) and had the strongest regional association in meta-analysis of IGAP+EADB (Stage 2). Variants in close proximity with similar allele frequencies were not included. UKBB AD-proxy association results and meta-analysis of IGAP, EADB, and UKBB (Stage 3) are also shown.

| Variant RSID | Chr | Basepair (hg19) | Genomic Region | EFF | REF | Stage 2: IGAP + EADB Meta-Analysis |  |  | UKBB |  |  | Stage 3: IGAP + EADB + UKBB Meta-Analysis |  |  |
| --- | --- | --- | --- | --- | --- | --- | --- | --- | --- | --- | --- | --- | --- | --- |
|  |  |  |  |  |  | EAF | OR (95% CI) | P | EAF | OR (95% CI) | P | EAF | OR (95% CI) | P |
| rs183427010 | 19 | 45366498 | *APOE* Region (*PVRL2*) | A | G | 0.0093 | 2.81 (2.51, 3.13) | 5.32E-75 | 0.0055 | 1.53 (1.39, 1.69) | 1.82E-17 | 0.0075 | 2.00 (1.86, 2.15) | 1.53E-76 |
| rs75932628 | 6 | 41129252 | *TREM2* | T | C | 0.0038 | 2.34 (1.97, 2.78) | 2.15E-22 | 0.0025 | 1.45 (1.25, 1.69) | 9.00E-7 | 0.0032 | 1.79 (1.60, 2.00) | 5.59E-24 |
| rs143080277 | 2 | 106366056 | *NCK2* | C | T | 0.0054 | 1.48 (1.29, 1.70) | 2.69E-08 | 0.0041 | 1.34 (1.18, 1.52) | 3.22E-6 | 0.0048 | 1.40 (1.28, 1.54) | 7.17E-13 |
| rs921719566 | 16 | 76230037 | *CNTNAP4* (upstream) | G | A | 0.0077 | 0.75 (0.67, 0.84) | 4.51E-7 | 0.0066 | 1.02 (0.93, 1.13) | 0.651 | 0.0071 | 0.89 (0.83, 0.96) | 0.00252 |
| rs1032524620 | 5 | 140854118 | *PCDHGA/B* (gene family cluster) | G | C | 0.0003 | 5.14 (2.66, 9.96) | 1.17E-6 | -- | -- | -- | 0.0003 | 5.14 (2.66, 9.96) | 1.17E-6 |
| rs778473798 | 5 | 66025094 | *MAST4* | T | G | 0.0003 | 13.8 (4.72, 40.4) | 1.65E-6 | -- | -- | -- | 0.0003 | 13.8 (4.72, 40.4) | 1.65E-6 |
| rs779452860 | 19 | 24444215 | *ZNF254* (downstream) | G | T | 0.0006 | 3.62 (2.14, 6.15) | 1.79E-6 | -- | -- | -- | 0.0006 | 3.62 (2.14, 6.15) | 1.79E-6 |
| rs1004589836 | 18 | 59553737 | *RNF152* | C | A | 0.0004 | 3.93 (2.20, 7.02) | 3.81E-6 | -- | -- | -- | 0.0004 | 3.93 (2.20, 7.02) | 3.81E-6 |
| rs570907469 | 2 | 158285821 | *CYTIP* | A | G | 0.0026 | 1.79 (1.40, 2.29) | 3.88E-6 | 0.0025 | 1.04 (0.88, 1.22) | 0.677 | 0.0025 | 1.23 (1.07, 1.41) | 0.00338 |
| rs749016100 | 7 | 91966514 | *ANKIB1* | G | A | 0.0004 | 4.39 (2.34, 8.22) | 3.93E-6 | -- | -- | -- | 0.0004 | 4.39 (2.34, 8.22) | 3.93E-6 |
| rs527488596 | 3 | 90428380 | *EPHA3* (downstream) | A | T | 0.0055 | 0.65 (0.53, 0.78) | 6.32E-6 | 0.0057 | 0.98 (0.88, 1.1) | 0.784 | 0.0056 | 0.88 (0.80, 0.97) | 0.0113 |
| rs776752144 | 10 | 36231917 | *PCAT5* (downstream) | G | T | 0.0002 | 13.1 (4.28, 40.0) | 6.43E-6 | -- | -- | -- | 0.0002 | 13.1 (4.28, 40.0) | 6.43E-6 |
| rs958893222 | 3 | 104387013 | *ALCAM* (upstream) | A | G | 0.0003 | 7.51 (3.13, 18.0) | 6.51E-6 | -- | -- | -- | 0.0003 | 7.51 (3.13, 18.0) | 6.51E-6 |
| rs779246720 | 10 | 92032063 | *HTR7* (downstream) | A | G | 0.0013 | 0.46 (0.33, 0.65) | 6.73E-6 | -- | -- | -- | 0.0013 | 0.46 (0.33, 0.65) | 6.73E-6 |
| rs536943899 | 3 | 28624836 | *ZCWPW2* (downstream) | C | T | 0.0017 | 1.79 (1.39, 2.31) | 6.98E-6 | 0.0025 | 1.00 (0.85, 1.19) | 0.955 | 0.0022 | 1.20 (1.04, 1.38) | 0.0109 |
| rs190394847 | 21 | 45550148 | *PWP2* | A | G | 0.001 | 2.90 (1.82, 4.61) | 7.61E-6 | 0.0012 | 0.90 (0.70, 1.17) | 0.440 | 0.0011 | 1.19 (0.95, 1.50) | 0.130 |
| rs146698767 | 21 | 45544573 | *PWP2* | A | G | 0.0013 | 2.49 (1.67, 3.71) | 8.21E-6 | 0.0015 | 0.91 (0.72, 1.14) | 0.397 | 0.0014 | 1.16 (0.95, 1.42) | 0.135 |
| rs559275238 | 19 | 45939260 | *ERCC1* | G | T | 0.0067 | 1.54 (1.27, 1.85) | 8.45E-6 | 0.0080 | 1.06 (0.96, 1.16) | 0.263 | 0.0078 | 1.14 (1.05, 1.24) | 0.00282 |
| rs138181035 | 19 | 44863163 | *ZFP112* | T | C | 0.0037 | 0.67 (0.57, 0.80) | 8.60E-6 | 0.0043 | 0.98 (0.86, 1.13) | 0.806 | 0.0041 | 0.85 (0.76, 0.95) | 0.00329 |

Abbreviations: Chr, chromosome; EFF, Effect allele; REF, reference allele; EAF, effect allele frequency; OR, Odds ratio; CI, Confidence interval; *P*, P-value
Shaded box indicates genome-wide significance within analysis stage

Stage 3 meta-analyses (**Supplementary Table 5**) greatly strengthened the association at the *NCK2* variant rs143080277, (Stage 3: MAF=0.0048, OR[95% CI]: 1.40 [1.28, 1.54], *P*=7.17×10^−13^; UKBB: MAF=0.0041, OR[95% CI]: 1.34 [1.18, 1.52], *P*=3.22×10^−6^). Since these *NCK2* associations were inferred from imputed genotypes, we used Sanger sequencing to independently validate imputed genotypes. Assigning genotypes where imputation probabilities were >0.9, we observed 98% agreement (98% sensitivity and specificity for the minor allele) between imputed and directly observed genotypes (n=127 samples sequenced, 50% predicted referent homozygotes and 50% predicted heterozygous carriers).

Rare variant association analysis in the UKBB parental history of dementia/AD GWAS identified novel associations in the chromosome 16p13.12 gene *MRTFB* (rs149416930, *P*=1.94×10^−8^) and on chromosome 12p12.1 near the genes *AC019209.2*/*MIR4302*/*RASSF8* (rs148024771, *P*=3.89×10^−8^) (**Supplementary Tables 6** and **7**).

We performed several follow-up analyses in the ADGC data on all *APOE*, *TREM2*, and *NCK2* associations to determine if they were independent from known associations in *APOE* (the common risk-increasing ε4 and rarer protective ε2 haplotypes) and in *TREM2* (R47H and R62H). For rare *APOE* region variants, adjustment for only *APOE* ε2 did not substantially reduce the association observed for any of the variants but after adjustment for both *APOE* ε2 and ε4, main effects were no longer statistically significant (**Supplementary Table 8a**), suggesting the association is driven by correlation with *APOE* ε4. Similarly, for *TREM2*, variants adjustment for R47H and R62H saw a reduction in the association of all rare variants upstream of *TREM2*, suggesting those associations are correlated with existing signals at R47H and R62H (**Supplementary Table 8b**). Separately modeling covariate adjustment for *APOE* ε2 and *TREM2* R47H, the strength of the association *NCK2* SNP rs143080277 was not substantially changed. We also tested statistical interaction between rs143080277 and *TREM2* R47H and also between rs143080277 and *APOE* ε2; the main effects of the *NCK2* variant in these models were not substantially changed, suggesting the *NCK2* variant remains largely independent of *APOE* ε2 and *TREM2* R47H (**Supplementary Table 8c**). We also examined *NCK2* for common variant and other rare variant associations, and though a discovery-level association was observed in the UKBB for a variant of MAF=0.022 (rs17269688, *P*=1.52×10^−6^), no *NCK2* variants other than rs143080277 had discovery-level associations in IGAP (Stage 1) (data not shown).

### Re-analysis with weighted AD/dementia parental history GWAS statistics

In prior analyses of the AD/dementia family history phenotype, the *β* and SE were doubled in order to account for the proxy phenotype being a measure of the reported AD/dementia affection status of two parents. We re-analyzed the Stage 1-3 common variant and rare variant analyses using a doubled *β* and SE to examine potential strengthened associations. For common variants, several loci which attained GWS in Stage 2 but not in Stage 3 were GWS in Stage 3 re-analysis (**Supplementary Table 9a**), including *ADAMTS1* (rs2830500, *P*=3.98×10^−10^), *SPI1* (rs11039202, *P*=2.88×10^−9^), *SORL1* (rs3781831, *P*=2.85×10^−10^), *USP50* [near *SPPL2A*] (rs3131610, *P*=1.35×10^−8^) and *IQCK* (rs9926151, *P*=3.61×10^−8^). In addition, a newly GWS association was observed in Stage 3 for a variant in *EIF4G3* (rs12407731, *P*=2.36×10^−8^). For rare variants, while the level of significance for most variants increased, no additional loci demonstrated GWS in Stage 3 re-analysis (**Supplementary Table 9b**), only the rare variants at *APOE*, *TREM2*, and *NCK2*. Revisiting the UKBB common variant and rare variant discovery analyses (**Supplementary Tables 9c** and **9d**, respectively), no additional variants demonstrated GWS in the UKBB + IGAP meta-analysis.

### Gene-Based Association Analysis

Performing gene-based association testing on all rare variants (MAF≤0.01) (**Supplementary Table 10**), we observed only Bonferroni-corrected statistically significant associations at *APOE* and *TREM2*. After applying additional filtering on CADD *C*-score predicted deleteriousness (**Supplementary Table 11**), again only *APOE* and *TREM2* demonstrated association at Bonferroni-corrected significance thresholds at all three deleteriousness thresholds, *C*≥5, *C*≥10, and *C*≥15 and including variants with imputation *R^2^*≥0.3,

We also examined gene-based association with MAGMA using common variants (MAF>0.01) within a 35kb upstream (5’) and 10kb downstream (3’) window around gene boundaries. Using a gene-based testing approach aggregating *P*-values for the most associated SNP and the mean SNP association (“*P*_multi_”), we identified several gene associations meeting a Bonferroni-corrected threshold for significance for 19,427 genes tested (*α*=2.57×10^−6^) (**Supplementary Table 12**). In addition to associations at multiple genes in the *APOE* region, many genes containing GWS-associated SNPs from single-variant analyses were also strongly associated in gene-based analyses, including *CR1/CR1L*, *BIN1*, multiple *MS4A* region genes, *CLU*, *PICALM*, *TREM2*, *CD2AP*, *SPI1*, *SORL1*, *IQCK*, *PTK2B*, and *HBEGF*. Notably, multiple genes in the immediate vicinity of *SPI1* on chromosome 11p11.2 demonstrated highly significant associations exceeding both GWS and Bonferroni-corrected thresholds in both single-variant and gene-based analyses, including the genes *SLC39A13*, *MTCH2*, *PSMC3*, *FNBP4*, *NDUFS3*, *NUP160*, *RAPSN*, *AGBL2*, *PTPMT1*, and *MYBPC3*.

### Pathway Analysis

MAGMA-based pathway analyses using multiple annotation sources identified 30 distinct pathways with FDR-corrected associations of *Q*<0.05 when using gene-wide statistics from the MAGMA “multi” association model and a 35kb upstream/10kb downstream window to assign SNPs to genes (**Supplementary Table 13**). These included more significant associations for LOAD pathologic processes identified in previous IGAP analyses^12^, such as both negative and positive regulation of APP protein catabolism (GO:1902991, GO:1902993), tau protein binding (GO:0048156), Aβ formation (GO:1902003); pathways capturing cognitive processes, such as learning or memory (GO:0007611) and cognition (GO:0050890); and immune-related pathways like those involving the MHC protein complex (GO:0042611). Examining the strongest gene-based associations among genes assigned to the 30 pathways with strongest associations (*Q*<0.05) in the MAGMA pathway analysis (**Supplementary Table 14**) identified multiple genes containing confirmed loci, including *APOE*, *CLU*, *BIN1*, *PICALM*, *SORL1*, *PTK2B*, and *ACE*, as well as several biological candidates including *SCARA3*, and *SPI1* region genes such as *NUP160* and *PTPMT1*.

### Co-expression Analysis

MAGMA analyses (“multi” gene association module with a 35kb 5’/10kb 3’ window) using co-expression modules from multiple sources (**Supplementary Table 15**) identified strong associations in modules from multiple sources that are enriched for immune response-related genes, some of which have considerable overlap with others (Gibbs:34, Gandal:CD11, Neueder:28, and Neueder:54) while others included distinct gene sets (Zhang:25). Two of the most strongly associated modules (Gibbs:34 and Gibbs:56), both previously correlated with AD^42^, were enriched for genes in the *TYROBP* causal network which relates to immune receptor binding on microglia, however none of the overlapping genes included GWAS signals or other known AD susceptibility loci.

The genes most strongly associated in the top co-expression modules (Neueder:28, Gibbs:34, Zhang:37, and Gandal:CD11) (**Supplementary Table 16**) include many genes containing top LOAD susceptibility loci from GWAS (several *APOE* region genes, *MS4A4A*/*MS4A6A*, *PICALM*, *SPI1*, *TREM2*, *SORL1*, *SLC24A4*, and *ACE*) as well as genes (*RERE*, *PURA*) implicated in AD pathology in functional or transcriptomic analyses^43^.

### Analysis of Genetically Regulated Expression Using PrediXcan

We used PrediXcan to assess whether the variants (**Supplementary Table 17a-b**) and genes (**Supplementary Table 17c-d**) with discovery-level association (*P*<10^−5^ for variants; Bonferroni-corrected α thresholds for genes) were associated with expression patterns imputed using whole-body tissue transcriptome reference data from the GTEx Consortium (version 8). Looking at genes containing GWS variants, we observed 137 genes associated with FDR-corrected *Q*<0.05 (**Supplementary Table 17a**) across a broad range of tissues, including brain region-specific associations with *CR1* and *CLU* as well as multiple *SPI1*-region genes including *MTCH2* and *SLC39A13*. Collapsing expression patterns across brain tissues (“cross-brain”), we observed brain-specific associations (*Q*<0.05) in *APOE* region genes, *CLU*, *CR1*, and *MS4A* region genes (**Supplementary Table 17b**). Following up on 20 genes of interest from gene-based analyses, five genes met FDR-corrected *Q*<0.05 in preliminary analyses of expression of individual tissue types (**Supplementary Table 17c**), while six of the 20 genes had at least one significant association of FDR-corrected *Q*<0.05 with collapsed expression patterns across all tissues (“cross-all”) or “cross-brain” (**Supplementary Table 17d**): *TOMM40*, *BCAM*, *PPP1R37*, and *CLASRP* in the *APOE* region; *PSMC3* in the vicinity of *SPI1*; and *TREM2*.

### NCK2 Tissue-Specific Expression

The GWS variant rs143080277 is in the first intron of *NCK2* (encoding the NCK adaptor protein 2; MIM: 604930). This gene is in a 743 kb topologically-associating domain (TAD)^44^ containing only three other genes, *C2orf49*, *FHL2*, and *ECRG4* (*C2orf40*). Of these genes, only *C2orf49* and *NCK2* are expressed in multiple brain regions. *NCK2* is also highly expressed in a variety of cell types including CD19+ B-Cells, CD4+ T cells, and CD8+ B cells (**Supplementary Figure 6**, data from BioGPS^45^). Single nuclei RNA-Seq of these genes show that NCK2 is selectively expressed in the AD-relevant cell type, microglia (**Figure 2**). We examined human cortex samples^46^ that included normal tissue without amyloid or tau aggregates (A-T-), tissue with amyloid only (A+T-) and tissue with both amyloid and tau aggregates (A+T+). The highest expression was observed is in amyloid-responsive and homeostatic microglia in tau-negative regions (A+T-). *NCK2* expression in microglia is also influenced by *TREM2* genotypes and is reduced in R47H *TREM2* carriers compared to the wildtype genotype. Also, *NCK2* expression is reduced in *APOE* ε4/ε4 subjects compared to *APOE* ε3/ε3 or ε3/ε4. Both *TREM2* and *APOE* are part of the microglial response triggered in AD.

### Mapping Regulatory Roles and Chromatin Interactions of rs143080277/NCK2

The *NCK2* variant rs143080277 falls within enhancer regions in 47 ROADMAP^47^ enhancer tracks, including tissues in four brain regions (dorsolateral prefrontal cortex, inferior temporal lobe, substantia nigra, and anterior caudate nucleus) and cell types including monocytes and T cells, among others. Analyzing Hi-C data^48^ from six human cell lines with ENCODE/ROADMAP functional genomic data, we found interactions between the rs143080277 location and multiple other genomic regions, but not the promoter of *NCK2* (**Supplementary Figure 7**). Using our high-resolution promoter-focused Capture C and ATAC-Seq database that includes >40 cell types and conditions^49^, we observed two enhancer-promoter interactions in follicular B helper T (TFH) cells^50^. One is a loop (∼390 kb) from this SNP to an open alternative promoter of a short isoform (NM_001253876.1) of *UXS1* in TFH cells (**Supplementary Figure 8a**). The other contacted a closed alternative promoter of a short isoform (NM_001004720.3) of *NCK2*.

We also performed a more relaxed search in neural cell types only without requiring the SNP or the gene promoter to reside in open chromatin and investigating both the index SNP (rs143080277) and a proxy SNP (rs144636993; *r^2^*∼0.5 (*D’*=1) in the 1000 Genomes European [EUR] populations). One loop was observed between one loop between rs144636993 and an alternative promoter of the same short isoform (NM_001004720.3) of *NCK2* in ESC-derived hypothalamic neurons^51^, however neither the SNP nor the promoter were found to be open (**Supplementary Figure 8b**).

## Discussion

In this study, we imputed rare variants using a high-density haplotype reference panel in a large collection of LOAD GWAS to identify novel associations with individual variants, genes, and gene sets, with the goal of further elucidating the genetic contributors to the pathology of this highly prevalent disease. We extended our genomic interrogation with functional genomics annotation and AD-relevant functional genomics resources including expression data and chromatin structure data to better characterize the role of novel variants we identified. This strategy proved effective in identifying a novel rare variant association with LOAD in the gene *NCK2*, rs143080277 (MAF=0.0054), which appears to alter expression patterns in amyloid-responsive microglia. Although more functional characterization is needed, this suggests a potential role in AD pathology worth additional exploration.

While little is understood about the specific role or roles *NCK2* (encoding the NCK adaptor protein 2; MIM: 604930) may play in neurodegenerative processes, it is known to interact with a variety of genes/proteins that have already been implicated in AD (**Supplementary Figure 9**), including ephrins (*EFNB1*, *EFNB2*, *EFNB3*) [genes encoding ephrin receptors include known AD gene *EPHA1*]; *VLDLR*, which encodes an ApoE receptor; and *IGF1R*^12^.

Our exploratory functional work noted that the associated rs143080277 variant may perform a regulatory role in the expression of *NCK2* in brain tissues, and in particular amyloid-responsive microglia. Furthermore, our examination of differential expression patterns of *NCK2* against *APOE* ε2/ε3/ε4 and *TREM2* R47H backgrounds suggests that there may be some functional interactions with these major contributors to AD risk.

In addition to the novel rare variant association, we observed several novel common variant associations at *APH1B*, *TSPAN14*, *CLNK*, *SHARPIN*, *MTSS1L/IL34*, and *ATF5/SIGLEC11* as well as the first ancestry-specific confirmatory GWS associations of *TSPOASP1-AS1* and *HBEGF*, which were previously observed in a multi-ethnic GWAS of LOAD^52^. Among the novel loci, *TSPAN14*, encoding tetraspanin 14, is one of a family of proteins involved in proteolytic processing of the amyloid precursor protein (APP)^53^ through physical interactions with APP secretases, and appears to regulate ADAM10 cleavage of substrates^54^, which may alter levels of Aβ production^55^. *SHARPIN*, which encodes Shank-associated RH domain-interacting protein, is a key regulator of neuroinflammatory response, stimulating macrophage activity and promoting Aβ phagocytosis in the presence of circulating Aβ^56^. A rare variant in *SHARPIN* have been shown to limit immune response^57^, which may promote LOAD development. *ATF5*, encoding activating transcription factor 5, and plays a critical regulatory role in neuronal development, downregulating differentiation of neuroprogenitor cells^58^. *SIGLEC11* is a member of the sialic acid-binding, immunoglobulin-like lectin family, and Siglec-11 mediates anti-inflammatory activity and acts as an inhibitory receptor of inflammatory responses of microglia, potentially regulating microglial neurotoxivity^59, 60^. Among the loci identified in previous studies which used proxy cases/controls, *APH1B*, which encodes the Aph1B subunit of the γ-secretase complex, is critical to processing APP and inhibition or deletion of the subunit leads to reduced clearance of Aβ^61^ and memory issues^62^. *MTSS1L*, encoding metastasis suppressor 2 (formerly metastasis suppressor 1-like), is involved in plasma membrane dynamics. It is highly expressed in brain with modest or little expression in other tissues^63^. Though there is limited information on its potential role in Alzheimer’s disease, whole exome sequencing in 143 consanguineous families identified variation in this gene associated with neurodegeneration^64^. Variants of *MTSS1L* have also been implicated in a spectrum of neurodevelopment-related phenotypes including global development delay^65^ and educational attainment^66^. IL34 encodes the cytokine Interleukin-34^67^ which promotes the proliferation of microglia and clearance of soluble oligomeric Aβ, reducing neuronal damage in AD^68^. *CLNK* encodes cytokine-dependent hematopoietic cell linker and is known to play a role in immunoreceptor-mediated signaling events. *CLNK* has been shown to interact, when phosphorylated, with *PLCG2*^69^. *PLCG2* is a transmembrane signaling enzyme with critical regulatory functions in the immune system^70^ in which variation has been shown previously to be associated with AD^11^. While *CLNK* has previously been identified in GWAS of family history of AD and studies mixing AD status with AD/dementia family history, this is first time GWS association with a pure AD phenotype has been observed at this locus.

The emerging LOAD loci confirmed in this analysis, *TSPOAP1* and *HBEGF*, were both observed with GWS in a prior GWAS meta-analysis^52^ of 33,269 ADGC subjects, including 26,320 individuals of European ancestry, 4,983 African Americans, 1,845 Japanese subjects, and 115 Israeli-Arab participants). It should be noted that in a multi-ethnic study, allele frequencies and strength of association may be variable across ancestries; this study is the first time that these associations have been observed to attain genome-wide significance within ancestry among subjects of European ancestry, strengthening support for true association. That these loci were only observed as GWS in a single-ancestry sample of 80,685 cases and 243,682 controls of European ancestry, a study nearly 10 times larger than the multi-ethnic sample, reflects the power and utility of trans-ethnic meta-analyses in identifying novel loci over much larger ancestrally homogeneous data collections.

*TSPOAP1* (formerly *BZRAP1*) encodes benzodiazepine-associated protein 1, a subunit of the benzodiazepine receptor complex in mitochondria, of which increased binding activity may indicate brain inflammation^71–73^. Additionally, benzodiazepine use has been shown to increase risk of dementia^74^. *HBEGF*, which encodes heparin EGF-like growth factor, has varied roles including cardiac hypertrophy and development^75^. Some functional work suggests that *HBEGF* may have a role in AD: a murine model with *HBEGF* expression knocked out in cortex and hippocampus demonstrated the impaired psychiatric and cognitive functions expected with down-regulation of NMDA receptors^76^.

While our analyses benefitted from the combined statistical power of large, well-characterized AD GWAS collections with biobank GWAS data, we had several concerns about the different phenotype (AD/dementia parental history) we used in the UKBB. Following up discovery-level associations first in EADB and then in UKBB, while we expected and observed reduced statistical significance in those data because of the “winner’s curse,” we noticed lower strength of association (ORs closer to 1) across variants in UKBB compared to IGAP and EADB, which had similar strength of association, although the directions of association and minor allele frequencies were similar for most variants across all three datasets/collections. We surmise that this may indicate the effect of two forms of “misclassification” in the AD/dementia parental history phenotype relative to LOAD affection status: (1) as subject ages-at-exam in UKBB are mostly below the LOAD average age-of-onset in IGAP and EADB, the parental history phenotype misclassifies true future AD status, as some persons with parental history will not develop AD and some persons without parental history will develop AD; (2) as the phenotype includes parental history of dementia, some subjects are likely at elevated risk of alternate causes of dementia (e.g., Parkinsonian dementia, Frontotemporal Dementia, and Lewy Body Disease, among others) relative to their risk for AD. While we expect that (1) might explain reduced associations in UKBB, we performed a discovery GWAS analysis using UKBB and the parental history phenotype to determine whether a large set of non-AD loci would be identified. If this were the case, this would suggest that the more concerning form of misclassification might be (2). In comparing the top hits of the two analyses, a substantial overlap was observed among the strongest associations, which included GWS associations in the *APOE* region, *BIN1*, *HLA-DRB1*, *ABCA7*, *PICALM*, *ADAM10*, the *MS4A* region, and *CR1*. Several of these top associations are shared across multiple neurodegenerative phenotypes (e.g., *HLA-DRB1* in Parkinson’s disease (PD)). One association with GWS after combining UKBB and IGAP was observed in *EPHX2*, which has some prior evidence of association with AD^77^ and upregulation of its hydrolase activity leads to increased neuronal cell death^78^, though it is unclear if this association is capturing the effects of variation at the proximal genes *CLU* and *PTK2B*. A discovery-level association (rs575683378, *P*=4.85×10^−6^) was observed for a rare variant (MAF=0.0014) in the gene encoding α-synuclein (*SNCA*), as established PD locus, however a similar rare variant association was not observed at or near this variant in IGAP. While this does not rule out possible misclassification effects from persons with higher risk of dementia from non-AD causes, it suggests that the higher prevalence of AD relative to other forms of dementia likely exerts minimal effect on the most strongly associated AD loci. It remains unclear if combining LOAD GWAS studies in IGAP and EADB with less rigorously defined case definitions like in UKBB would increase the possibility of misclassification. Therefore, the increased sample size available from the integration of studies with “silver standard” phenotypes (such as UKBB) may be useful if evaluated with appropriate caution regarding study conclusions^79^.

This study has several potential limitations, including modest increases in sample sizes from prior AD GWAS meta-analyses and potentially reduced power to detect novel AD loci by excluding the UKBB GWAS from discovery-level analyses. While the modest growth in sample size may have provided limited additional power to detect several additional novel common variant loci, we believe the strength of the analysis lie in the ability to robustly identify rare variant associations by combining high-quality rare variant imputation and a score-based regression approach that stabilized measures of effect size and standard error estimates across individual studies. Without this strategy, it is unlikely that rare variant associations could have been examined in small datasets due to wide variability in effect betas and standard errors, which would not be distinguishable from other sources of heterogeneity between studies. Regarding the decision to not include UKBB in our IGAP discovery analyses, our secondary analysis of UKBB showed that while combining IGAP and UKBB may have identified additional AD GWAS loci, it may have also led to discovery-level significance (*P*<10^−5^) of risk variants for other dementia causes being potentially misreported as AD susceptibility loci if those loci have strong effects in UKBB alone. Those loci might yield valuable insights into AD pathogenesis, but would require additional scrutiny with pleiotropy analyses and functional genomics studies to confirm that their roles in neurodegeneration are important to AD.

## Methods

### Variant- and sample-level quality control (QC) on Stage 1 IGAP data

Standard QC was performed on individual IGAP datasets using PLINK v1.9^80–82^ and including filtering and re-estimating all quality metrics after excluding variants with a missingness rate of >10% of genotype calls. QC filters included exclusions on SNPs with call rates below 98% for Illumina and 95% for Affymetrix panels; SNPs with departure from Hardy-Weinberg Equilibrium (HWE) of *P*<10^−6^ among cognitively normal elders (CNEs, either non-cases or controls) for variants of MAF>0.01; and SNPs with informative missingness by case-CNE status of *P*<10^−6^. Samples were dropped if the individual call rate was <95%; if X chromosome heterozygosity indicated inconsistency between predicted and reported sex; or if population substructure analyses (described below) indicated the sample did not cluster with 1000 Genomes Phase 3 populations of European ancestry. More details including differences in QC approach for EADB and UKBB are provided in the Methods section of the **Supplemental Note**.

### Relatedness Checks in Stage 1 IGAP data

Relatedness was assessed using the “--genome” function of PLINK v1.9. Using ∼20,000 linkage disequilibrium (LD)-pruned SNPs sampled from among genotyped variants, 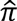 (the proportion of alleles shared IBD) was estimated across all pairs of subjects across all ADGC datasets. Among pairs of subjects with no known familial relationships, one sample was excluded among pairs with 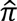 >0.95 if phenotype and covariate data matched, otherwise both samples were excluded; among all pairs with 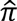 >0.4 but less than 0.95, one sample was kept giving preference to cases over CNEs, age (earlier age-at-onset among case pairs, later age-at-exam among CNE pairs). Pairs of relatives were dropped from family datasets if 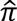 differed substantially from expectation based on their reported relationships.

### Population substructure in Stage 1 IGAP data

To identify samples of non-European ancestry, we performed a principal components (PCs) analysis using ‘smartpca’ in EIGENSOFT^83, 84^ on the subset of ∼20,000 LD-pruned SNPs used for relatedness checks on genotypes from all samples within each individual dataset and from the 1000 Genomes Phase III reference panels. Subjects not clustering with European ancestry groups were excluded from analysis. To account for the effects of population substructure in our analysis, a second PC analysis was performed using only the remaining subjects in each dataset. PCs 1-10 were examined for association with AD case-control status and eigenvector loading, and only PCs showing nominal association with AD (*P*<0.05) and eigenvector loadings >3 were used in covariate adjustment for populations substructure, (average number of PCs used is 3; range: 2-4).

### Imputation of Stage 1 IGAP data

For each dataset, SNPs not directly genotyped were imputed on the Michigan Imputation Server (MIS)^85^ using samples of all ancestries available on the Haplotype Reference Consortium (HRC) 1.1 reference panel^86^, which includes 39,235,157 SNPs observed on 64,976 haplotypes (from 32,488 subjects), all with an estimated minor allele count (MAC)≥5 and observed in samples from at least two separately-ascertained data sources. Phasing on the MIS was done with EAGLE^87^ while imputation was performed using Minimac3^85^. Quality of imputation for all variants was assessed using *R^2^* for imputation quality, although all variants were retained and not filtered prior to analysis. Following imputation and analysis, variants were filtered using two quality thresholds, a conservative *R^2^*≥0.8, suggested to assure high quality of rare variants with MAF≤0.01, and a more liberal *R^2^*≥0.3 to evaluate common variants (MAF>0.01). Analysis of SNP imputation quality by bin of MAF revealed that more than 80% of variants of MAF>0.0005 had *R^2^*≥0.8; among variants with MAF≤0.0005, approximately 50% of variants had *R^2^*≥0.8 in most datasets with n>1,000 subjects. Final Stage 1 (IGAP) analyses include only variants of *R^2^*≥0.8, using a global average of *R^2^* across all datasets weighted by sample size, to minimize potential bias from differences in imputation quality across datasets, while imputation quality were relaxed to *R^2^*>0.4 for the same variants followed up in EADB and UKBB. Genotyped and imputed variants were all mapped to the mapped to the GRCh137/hg19 human genome build.

### Single-variant Association Analysis and Meta-analysis for Common Variants (MAF>0.01)

Single variant-based association analysis on datasets of unrelated cases and CNEs were performed in SNPTEST^88^ using score-based logistic regression under an additive model with minimal covariate adjustment for PCs and study-specific indicator variables only. Additional analyses with adjustment for age (defined as age-at-onset for cases and age-at-last exam for CNEs), sex, and dosages of *APOE* ε4 (0/1/2 copies) and *APOE* ε2 were evaluated and we observed similar findings to those reported for the minimally adjusted model, however those results are not shown. Family-based datasets were analyzed using the GWAF^89^ package in R^90^ which implemented a generalized estimating equation (GEE) approach to account for correlation between subjects. After association analysis on imputed data, variants with regression coefficient of |β| > 5, negative standard errors, or *P*-values equal to 0 or 1 were excluded from further analysis. Within-study association results for common variants (MAF>0.01) were meta-analyzed using a fixed-effects approach with inverse variance-weighting and with genomic control using METAL^91^.

### Single-variant Association Analysis and Meta-analysis for Rare Variants (MAF≤0.01)

Rare variant association and meta-analysis was performed for individual variants using the SeqMeta^92^ package in R^90^. SeqMeta performs a score-based logistic regression, estimating scores in individuals using ‘prepScores()’ and performing meta-analysis using ‘singleSNPMeta()’. Family-based datasets were analyzed by selecting a maximally informative subset of unrelated individuals for analysis, and no datasets with fewer than 100 cases and/or CNEs were analyzed (including the CSDC which demonstrated extreme association patterns and genomic inflation suggesting potential bias). As in common variant analyses, models evaluated included covariate adjustment for PCs. After meta-analysis of imputed rare variants, any SNPs with a regression coefficient of |β| > 5, negative standard errors, or *P*-values equal to 0 or 1 were excluded from further analysis.

### MAGMA Gene-based Association and Pathway Analyses

We performed gene-based analyses and multiple pathway analyses of the IGAP GWAS using common variants (MAF>0.01) with MAGMA v1.08^93^. For gene-based analysis, MAGMA uses GWAS summary statistics to perform a SNP-wise analysis within a gene using either (a) the mean association statistic across all SNPs in a gene, denoted by “mean” in **Supplementary Tables 12-14** and **16** or (b) a combination of the mean statistic and that of the most significant single SNP, denoted by “multi”. MAGMA uses a numerical integration procedure combined with simulation to produce an accurate *P*-value that corrects for LD structure between variants.

Pathway/gene-set analyses perform a joint test of association on multiple genes to determine strength of association with the phenotype of interest compared to the association of all other genes in the genome (a “competitive” analysis), correcting for LD between genes. The version of MAGMA used (v1.08) was recently updated to fix an elevated rate of false-positive associations observed in prior analyses^94^. Gene sets used in the analyses were from GO^95, 96^, KEGG^97, 98^, REACTOME^99, 100^, BIOCARTA^101^, and MGI^102^ pathways. Analyses examined 17,574 genes with 7,565,152 common variants (MAF>0.01), in two subsets of analyses: (a) using variants within the annotated 5’ and 3’ boundaries of the genes and (b) using a 35-kb upstream/10-kb downstream window around each gene to incorporate potential regulatory variants^103^. LD structure from MAGMA was determined using a combined ADGC reference panel using HRC imputation and genotype assignment (based on genotype probability from imputation exceeding 0.9) including data on 31,876 subjects and 8,299,079 variants. GO pathway analysis was restricted to GO terms containing between 10 and 2000 genes. No size restrictions were placed on the other gene sets, since there were many fewer of them. This resulted in a total of 10,042 gene sets for analysis. Multiple testing correction for the number of gene sets tested was applied by calculating *Q*-values using the R package ‘qvalue’^104^.

### MAGMA Co-Expression Analysis

While pathway analyses examine gene sets defined by their shared role in known biological pathways, co-expression analyses examine gene sets (modules) defined by correlated expression patterns in the same or related tissues, on the grounds that such genes are likely to have similar biological functions. Here we analyzed co-expression data from multiple annotation sources to identify modules of co-expressed genes enriched for the current GWAS signals using an approach implemented in previous IGAP studies^12, 42^. We collated a large set of co-expression modules derived from brain tissue from the following studies: BRAINEAC^105^, the Common Mind Consortium^106^, Gandal et al. (2018)^107^, Gibbs et al. (2010)^108^, Neueder & Bates (2014)^109^, ROSMAP^110^, PsychENCODE^111^, and Zhang et al. (2013)^112^. Since immunity plays an important role in genetic susceptibility to AD, we also used WGCNA^113^ to derive co-expression modules from the monocyte gene expression data of Fairfax et al. (2014)^114^. A complete description of the datasets, along with the number and size of modules, is given in **Supplementary Table 19**. These modules were used as gene sets for testing enrichment of association signals with MAGMA v1.08^93^. To facilitate biological interpretation of modules showing statistically significant enrichment, annotations of the modules were performed using gProfiler^115^.

### Gene-based Association Analysis of Rare Variants

We performed gene-based testing on rare variants (MAF≤0.01) using two approaches: (a) burden tests^116, 117^; and (b) SKAT-O^118^, which optimally combines the features of the sequence kernel association test (SKAT)^119^ and burden tests. SKAT testing pre-specified beta distribution weighting of β(1, 25) (“Wu weights”). As we examined variants with frequencies as low as MAF∼0.0008 in IGAP datasets of varying sample size, we used the meta-analytic framework of SeqMeta in R to perform gene-based testing across datasets to limit asymptotic effects on the estimation of betas and standard errors due to small cell sizes. Using subject-level scores in individuals previously estimated using ‘prepScores()’ for single variant analyses, we performed gene-based testing of 29,138,970 variants assigned to 22,650 genes with RefSeq^120^ release 85 using the ‘skatOmeta()’ argument for SKAT-O testing and ‘burdenMeta()’ for burden testing. Pre-analysis filtering of variants excluded variants at two imputation quality thresholds, *R^2^*>0.3 and *R^2^*>0.8, with gene-based analysis performed in parallel on variant sets at both thresholds. Post-analysis filtering of gene-based results excluded genes with fewer than two variants of MAF≤0.01, leaving a final set of 21,198 genes in the *R^2^*>0.3 analysis and 20,377 genes in the *R^2^*>0.8 analysis.

### Gene-based Association Analysis of Rare Variants Using Predicted Deleteriousness

We further extended gene-based testing in SeqMeta/R by filtering on predicted deleteriousness using the Combined Annotation-Dependent Depletion (CADD)^121^ method. CADD^122^ systematically integrates multiple functional annotation metrics into the “*C*-score” to indicate predicted deleteriousness. We downloaded CADD *C*-score annotations (Development release v1.6) from the CADD server at the University of Washington for all ∼39.2M variants imputed by HRC and mapped these annotations to variants accordingly. After removing all variants of MAF>0.01 and variants of MAF≤0.01 to which no *C*-scores mapped, we applied imputation filters of *R^2^*>0.3 and *R^2^*>0.8 as before to generate two sets of variants, and then further filtered these into sets of variants with *C*≥5, *C*≥10, and *C*≥15 prior to gene-based testing. We used *C*≥15 as the highest threshold following CADD recommendations^123^ as this includes 5% of variants genome-wide and is the median C-score for all possible non-synonymous variants and canonical splice site changes. Following filtering on MAF, *R^2^*, and *C*-score, less than 25% of all genes in the *R^2^*>0.8/*C*≥10 and *R^2^*>0.8/*C*≥15 subsets had at least one variant, so the only subsets analyzed were *R^2^*>0.8/*C*≥5, *R^2^*>0.3/*C*≥10, *R^2^*>0.3/*C*≥15, and *R^2^*>0.8/*C*≥5.

### Functional Genomics Follow-Up at Loci with GWS Associations

To explore potential functional effects at the newly associated variants identified in this analysis, we performed several follow-up analyses and functionality assessments on both individual variants and genes such as (1) chromatin interaction analyses using annotation in whole-body and brain-specific tissue and cell types; (2) chromatin accessibility analyses using chromosome conformation capture and ATAC-Seq methods; (3) expression pattern differences between brain/non-brain tissues and cell types from existing resources; (4) genetically-regulated expression patterns using inferred expression patterns through PrediXcan; and (5) effects of variation on alternate splicing and on populations of microglia subtypes in postmortem AD and cognitively normal brains.

### PrediXcan and MultiXcan Analysis

These comprised two different analyses, a basic PrediXcan^124^ analysis examining individual tissues and a MultiXcan^125^ analysis aggregating expression across multiple tissue types. Tissue-specific genetic expression modules (GReX) in IGAP were imputed with publicly-available gene expression imputation models constructed in reference transcriptome datasets, including the whole blood model for DGN^126^, the dorsolateral prefrontal cortex (DLPFC) model from the CommonMind Consortium^127, 128^, and another 49 tissues models from the Genotype-Tissue Expression (GTEx) project (version v8)^129^. DGN and DLPFC models were trained using an elastic net approach, and the other models from GTEx v8 used multivariate adaptive shrinkage. S-PrediXcan was then run with summary statistics of qualifying GWS associations (*R^2^*>0.3 and MAF>0.01) with all 51 tissue-specific models described above and S-MultiXcan with two cross-tissue models, one model looking across all 49 GTEx tissues (“cross-all”) and one model looking across 13 GTEx brain tissues (“cross-brain”).

### Gene-based Phenome-wide Association Study

To investigate phenotypes and traits with shared genetic components, we conducted a phenome-wide association study in the DNA biobank at Vanderbilt University (BioVU). BioVU has collected DNA samples from over 280,000 participants and linked genetic data for over 70,000 to their electronic health records. To identify gene-trait associations phenome-wide, we used PredixVU, a PrediXcan-based application which estimates genetically regulated expression based on each individual’s genotype data using GTEx v8 models in a subset of 23,000 genotyped BioVU individuals of European ancestry. ICD-9 (International Classification of Disease, 9th Edition) codes from electronic health records were mapped to phenotype codes (“PheCodes”), defining cases and control for a set of diseases and symptoms^130, 131^; a total of 1,514 PheCodes were included. Then, association between imputed expression and PheCode was calculated.

All genes identified from single-variant and gene-based common and rare variant analyses were tested to characterize traits associated with their imputed expression in the electronic health records linked to BioVU samples. To identify traits with shared expression architecture, we first tallied significant (*P*<0.05) PredixVU association results for each trait with the selected genes. To obtain the *P*-value for each phenotype, we created an empirical distribution for each trait by first identifying the subset of selected genes expressed in a given tissue and then randomly selecting the same number of genes and assessing the number of genes in the random set that were associated with each trait in PredixVU. The process was repeated 100,000 times to create a null distribution, and the empirical *P*-value was estimated based on this permutation procedure. We performed a Bonferroni correction for 1,514 PheCodes, making the significance level *P*<3.3×10^−5^ for determining selected gene enrichment in GReX-trait association.

### Chromatin Interaction and Regulatory Variant Modeling Database Interrogation

To determine whether variants of interest may be involved in physical interactions with elements such as enhancers, insulators, or promoters, we applied HIPPIE2^48^ to high-throughput chromosome conformation capture (Hi-C) data from six human cell lines (GM12878, IMR90, K562, HMEC, HUVEC, NHEK) with matched Encyclopedia of DNA Elements (ENCODE) project^132, 133^ and Roadmap Epigenomics Project^47^ functional genomic data to infer physically-interacting regions (PIRs). We examined the chromosomal region ±50kb flanking the variant to identify a potential enhancer/promoter interaction pair. We also explored chromatin interactions using the high-resolution (1 kb) haploid and diploid three-dimensional (3D) genome maps constructed by Rao and colleagues^134^ using Hi-C to characterize chromatin ‘loop anchors’, stretches of sequence motifs that typically bind to the CTCF protein in a paired fashion at the ends of a chromatin loop. To determine potential regulatory effects of variants of interest, we looked at enhancer mapping from ChromHMM^135^, which integrates epigenomic states measured through chromatin immunoprecipitation and sequencing (ChIP-Seq) of histone modifications to mark active enhancers^136, 137^ using 127 tissues and cell types from ENCODE and Roadmap data.

### Examining Chromatin Accessibility and Looping using high resolution promoter-focused Capture C and ATAC-Seq

To examine chromatin looping and accessibility at the variants of interest, we queried our ATAC-Seq (Assay for Transposase-Accessible Chromatin using Sequencing) and high-resolution Capture C database comprising multiple cell types and conditions. Our Capture C library design utilizes custom Agilent SureSelect capture baits targeting both ends of DpnII restriction fragments (mean size ∼450bp) encompassing promoters (including alternative promoters) of all human coding genes, noncoding RNA, antisense RNA, snRNA, miRNA, snoRNA, and lincRNA transcripts, totaling 36,691 RNA baited fragments. Protocols for ATAC-Seq and Capture C have been described previously^138^.

We also investigated putative active promoters-enhancer interactions in epigenomic data from multiple brain cell types from the Nott et al. (2019)^49^ study. This dataset comprises ATAC-Seq, H3K4me3 and H3K27ac for neurons, astrocytes, microglia and oligodendrocytes from human brain biopsies, as well as cell specific-enhancers and chromatin looping data (PLAC-Seq) from microglia, neurons and oligodendrocytes.

### Candidate Gene Expression Patterns across Tissue Types

We queried two publicly available databases for expression information on genes of interest, BioGPS^139^ and Brain RNA-Seq^140^. BioGPS^45^, an online portal which combines extensive gene annotation resources to provide comprehensive information on genes of interest, includes expression data from a multitude of sources including multi-tissue reference expression patterns from the Gene Atlas data sets^141^, mouse brain expression in the Allen Brain Atlas^142^, multiple expression quantitative trait loci studies^143^ among others. Brain RNA-Seq^144, 145^ is an on-line a transcriptomics database with single-cell RNA-Seq data on multiple cell types including neurons, astrocytes, oligodendrocyte precursor cells, newly formed oligodendrocytes, myelinating oligodendrocytes, microglia, endothelial cells, and pericytes from mouse and human brains.

### Examining effects of candidate variants on microglia subpopulations in postmortem brains

We examined the effects of variants of interest on levels of microglia subpopulations originally identified using single-nucleus RNA sequencing applied to postmortem human brains from individuals with varied neuropathology and varied *APOE* and *TREM2* genotypes. This approach identified a subpopulation of CD163-positive microglia responsive to amyloid deposition that were significantly attenuated in individuals carrying *APOE* and *TREM2* risk haplotypes/genotypes^46^.

## Data Availability

Summary statistics will be made available upon publication. Original genotype and phenotype data may be made available upon request through study principal investigators and/or controlled-access genetics data repositories- see individual dataset descriptions for additional details.
All the data used in the gene prioritization are publically available:
GTEx pipeline: https://github.com/broadinstitute/gtex-pipeline
GTEx v8 eQTL and sQTL catalogues: https://www.gtexportal.org/
GTEx v8 expression and splicing prediction models: http://predictdb.org/

https://www.niagads.org/home

## Acknowledgements

*(Currently under revision)*

## Author Contributions

*(Currently under revision)*

## REFERENCES

1. Gatz, M. et al. Role of genes and environments for explaining Alzheimer disease. Arch Gen Psychiatry 63, 168–74 (2006).

2. Escott-Price, V. et al. Common polygenic variation enhances risk prediction for Alzheimer’s disease. Brain 138, 3673–84 (2015).

3. Harold, D. et al. Genome-wide association study identifies variants at CLU and PICALM associated with Alzheimer’s disease. Nat Genet 41, 1088–93 (2009).

4. Lambert, J.C. et al. Genome-wide association study identifies variants at CLU and CR1 associated with Alzheimer’s disease. Nat Genet 41, 1094–9 (2009).

5. Naj, A.C. et al. Common variants at MS4A4/MS4A6E, CD2AP, CD33 and EPHA1 are associated with late-onset Alzheimer’s disease. Nat Genet 43, 436–41 (2011).

6. Seshadri, S. et al. Genome-wide analysis of genetic loci associated with Alzheimer disease. JAMA 303, 1832–40 (2010).

7. Hollingworth, P. et al. Common variants at ABCA7, MS4A6A/MS4A4E, EPHA1, CD33 and CD2AP are associated with Alzheimer’s disease. Nat Genet 43, 429–35 (2011).

8. Lambert, J.C. et al. Meta-analysis of 74,046 individuals identifies 11 new susceptibility loci for Alzheimer’s disease. Nat Genet 45, 1452–8 (2013).

9. Escott-Price, V. et al. Gene-wide analysis detects two new susceptibility genes for Alzheimer’s disease. PLoS One 9, e94661 (2014).

10. Ruiz, A. et al. Follow-up of loci from the International Genomics of Alzheimer’s Disease Project identifies TRIP4 as a novel susceptibility gene. Transl Psychiatry 4, e358 (2014).

11. Sims, R. et al. Rare coding variants in PLCG2, ABI3, and TREM2 implicate microglial-mediated innate immunity in Alzheimer’s disease. Nat Genet 49, 1373–1384 (2017).

12. Kunkle, B.W. et al. Genetic meta-analysis of diagnosed Alzheimer’s disease identifies new risk loci and implicates Abeta, tau, immunity and lipid processing. Nat Genet 51, 414–430 (2019).

13. Jansen, I.E. et al. Genome-wide meta-analysis identifies new loci and functional pathways influencing Alzheimer’s disease risk. Nat Genet 51, 404–413 (2019).

14. Zhang, X. et al. A rare missense variant of CASP7 is associated with familial late-onset Alzheimer’s disease. Alzheimers Dement 15, 441–452 (2019).

15. Raghavan, N.S. et al. Whole-exome sequencing in 20,197 persons for rare variants in Alzheimer’s disease. Ann Clin Transl Neurol 5, 832–842 (2018).

16. Patel, D. et al. Association of Rare Coding Mutations With Alzheimer Disease and Other Dementias Among Adults of European Ancestry. JAMA Netw Open 2, e191350 (2019).

17. Vardarajan, B.N. et al. Coding mutations in SORL1 and Alzheimer disease. Ann Neurol 77, 215–27 (2015).

18. Vardarajan, B.N. et al. Rare coding mutations identified by sequencing of Alzheimer disease genome-wide association studies loci. Ann Neurol 78, 487–98 (2015).

19. Steinberg, S. et al. Loss-of-function variants in ABCA7 confer risk of Alzheimer’s disease. Nat Genet 47, 445–7 (2015).

20. Logue, M.W. et al. Two rare AKAP9 variants are associated with Alzheimer’s disease in African Americans. Alzheimers Dement 10, 609–618 e11 (2014).

21. Jun, G. et al. PLXNA4 is associated with Alzheimer disease and modulates tau phosphorylation. Ann Neurol 76, 379–92 (2014).

22. Wetzel-Smith, M.K. et al. A rare mutation in UNC5C predisposes to late-onset Alzheimer’s disease and increases neuronal cell death. Nat Med 20, 1452–7 (2014).

23. Blue, E.E. et al. Genetic Variation in Genes Underlying Diverse Dementias May Explain a Small Proportion of Cases in the Alzheimer’s Disease Sequencing Project. Dement Geriatr Cogn Disord 45, 1–17 (2018).

24. Tosto, G. & Reitz, C. Genomics of Alzheimer’s disease: Value of high-throughput genomic technologies to dissect its etiology. Mol Cell Probes 30, 397–403 (2016).

25. Naj, A.C., Schellenberg, G.D. & Alzheimer’s Disease Genetics, C. Genomic variants, genes, and pathways of Alzheimer’s disease: An overview. Am J Med Genet B Neuropsychiatr Genet 174, 5–26 (2017).

26. Jonsson, T. et al. A mutation in APP protects against Alzheimer’s disease and age-related cognitive decline. Nature 488, 96–9 (2012).

27. Jonsson, T. et al. Variant of TREM2 associated with the risk of Alzheimer’s disease. N Engl J Med 368, 107–16 (2013).

28. Guerreiro, R. et al. TREM2 variants in Alzheimer’s disease. N Engl J Med 368, 117–27 (2013).

29. Pottier, C. et al. High frequency of potentially pathogenic SORL1 mutations in autosomal dominant early-onset Alzheimer disease. Mol Psychiatry 17, 875–9 (2012).

30. Logue, M.W. et al. Targeted Sequencing of Alzheimer Disease Genes in African Americans Implicates Novel Risk Variants. Front Neurosci 12, 592 (2018).

31. Bellenguez, C. et al. Contribution to Alzheimer’s disease risk of rare variants in TREM2, SORL1, and ABCA7 in 1779 cases and 1273 controls. Neurobiol Aging 59, 220 e1–220 e9 (2017).

32. De Roeck, A. et al. An intronic VNTR affects splicing of ABCA7 and increases risk of Alzheimer’s disease. Acta Neuropathol 135, 827–837 (2018).

33. Wu, Y., Zheng, Z., Visscher, P.M. & Yang, J. Quantifying the mapping precision of genome-wide association studies using whole-genome sequencing data. Genome Biol 18, 86 (2017).

34. Panoutsopoulou, K., Tachmazidou, I. & Zeggini, E. In search of low-frequency and rare variants affecting complex traits. Hum Mol Genet 22, R16–21 (2013).

35. Quick, C. et al. Sequencing and imputation in GWAS: Cost-effective strategies to increase power and genomic coverage across diverse populations. Genet Epidemiol 44, 537–549 (2020).

36. Surakka, I. et al. The impact of low-frequency and rare variants on lipid levels. Nat Genet 47, 589–97 (2015).

37. Bellenguez, C. et al. New insights on the genetic etiology of Alzheimer’s and related dementia. medRxiv, 2020.10.01.20200659 (2020).

38. McDowell, I. Alzheimer’s disease: insights from epidemiology. Aging (Milano) 13, 143–62 (2001).

39. Jarvik, G., Larson, E.B., Goddard, K., Schellenberg, G.D. & Wijsman, E.M. Influence of apolipoprotein E genotype on the transmission of Alzheimer disease in a community-based sample. Am J Hum Genet 58, 191–200 (1996).

40. Slooter, A.J. et al. Risk estimates of dementia by apolipoprotein E genotypes from a population-based incidence study: the Rotterdam Study. Arch Neurol 55, 964–8 (1998).

41. Schwartzentruber, J. et al. Genome-wide meta-analysis, fine-mapping, and integrative prioritization identify new Alzheimer’s disease risk genes. medRxiv, 2020.01.22.20018424 (2020).

42. International Genomics of Alzheimer’s Disease, C. Convergent genetic and expression data implicate immunity in Alzheimer’s disease. Alzheimers Dement 11, 658–71 (2015).

43. Patel, H., Dobson, R.J.B. & Newhouse, S.J. A Meta-Analysis of Alzheimer’s Disease Brain Transcriptomic Data. J Alzheimers Dis 68, 1635–1656 (2019).

44. Beagan, J.A. & Phillips-Cremins, J.E. On the existence and functionality of topologically associating domains. Nat Genet 52, 8–16 (2020).

45. Wu, C. et al. BioGPS: an extensible and customizable portal for querying and organizing gene annotation resources. Genome Biol 10, R130 (2009).

46. Nguyen, A.T. et al. APOE and TREM2 regulate amyloid-responsive microglia in Alzheimer’s disease. Acta Neuropathol 140, 477–493 (2020).

47. Roadmap Epigenomics, C. et al. Integrative analysis of 111 reference human epigenomes. Nature 518, 317–30 (2015).

48. Kuksa, P.P., et al. HIPPIE2: a method for fine-scale identification of physically interacting chromatin regions. NAR Genom Bioinform 2, lqaa022 (2020).

49. Nott, A. et al. Brain cell type-specific enhancer-promoter interactome maps and disease-risk association. Science 366, 1134–1139 (2019).

50. Su, C. et al. Mapping effector genes at lupus GWAS loci using promoter Capture-C in follicular helper T cells. Nat Commun 11, 3294 (2020).

51. Lasconi, C. et al. Variant-to-Gene-Mapping Analyses Reveal a Role for the Hypothalamus in Genetic Susceptibility to Inflammatory Bowel Disease. Cell Mol Gastroenterol Hepatol (2020).

52. Jun, G.R. et al. Transethnic genome-wide scan identifies novel Alzheimer’s disease loci. Alzheimers Dement 13, 727–738 (2017).

53. Seipold, L. & Saftig, P. The Emerging Role of Tetraspanins in the Proteolytic Processing of the Amyloid Precursor Protein. Front Mol Neurosci 9, 149 (2016).

54. Jouannet, S. et al. TspanC8 tetraspanins differentially regulate the cleavage of ADAM10 substrates, Notch activation and ADAM10 membrane compartmentalization. Cell Mol Life Sci 73, 1895–915 (2016).

55. Yuan, X.Z., Sun, S., Tan, C.C., Yu, J.T. & Tan, L. The Role of ADAM10 in Alzheimer’s Disease. J Alzheimers Dis 58, 303–322 (2017).

56. Krishnan, D., Menon, R.N., Mathuranath, P.S. & Gopala, S. A novel role for SHARPIN in amyloid-beta phagocytosis and inflammation by peripheral blood-derived macrophages in Alzheimer’s disease. Neurobiol Aging 93, 131–141 (2020).

57. Asanomi, Y. et al. A rare functional variant of SHARPIN attenuates the inflammatory response and associates with increased risk of late-onset Alzheimer’s disease. Mol Med 25, 20 (2019).

58. Angelastro, J.M. et al. Regulated expression of ATF5 is required for the progression of neural progenitor cells to neurons. J Neurosci 23, 4590–600 (2003).

59. Angata, T. et al. Cloning and characterization of human Siglec-11. A recently evolved signaling molecule that can interact with SHP-1 and SHP-2 and is expressed by tissue macrophages, including brain microglia. J Biol Chem 277, 24466–74 (2002).

60. Wang, Y. & Neumann, H. Alleviation of neurotoxicity by microglial human Siglec-11. J Neurosci 30, 3482–8 (2010).

61. Serneels, L. et al. Differential contribution of the three Aph1 genes to gamma-secretase activity in vivo. Proc Natl Acad Sci U S A 102, 1719–24 (2005).

62. Biundo, F., Ishiwari, K., Del Prete, D. & D’Adamio, L. Deletion of the gamma-secretase subunits Aph1B/C impairs memory and worsens the deficits of knock-in mice modeling the Alzheimer-like familial Danish dementia. Oncotarget 7, 11923–44 (2016).

63. Fagerberg, L. et al. Analysis of the human tissue-specific expression by genome-wide integration of transcriptomics and antibody-based proteomics. Mol Cell Proteomics 13, 397–406 (2014).

64. Alazami, A.M. et al. Accelerating novel candidate gene discovery in neurogenetic disorders via whole-exome sequencing of prescreened multiplex consanguineous families. Cell Rep 10, 148–61 (2015).

65. Michelson, D.J. et al. Evidence report: Genetic and metabolic testing on children with global developmental delay: report of the Quality Standards Subcommittee of the American Academy of Neurology and the Practice Committee of the Child Neurology Society. Neurology 77, 1629–35 (2011).

66. Lee, J.J. et al. Gene discovery and polygenic prediction from a genome-wide association study of educational attainment in 1.1 million individuals. Nat Genet 50, 1112–1121 (2018).

67. Zuroff, L.R. et al. Effects of IL-34 on Macrophage Immunological Profile in Response to Alzheimer’s-Related Abeta42 Assemblies. Front Immunol 11, 1449 (2020).

68. Mizuno, T. et al. Interleukin-34 selectively enhances the neuroprotective effects of microglia to attenuate oligomeric amyloid-beta neurotoxicity. Am J Pathol 179, 2016–27 (2011).

69. Goitsuka, R. et al. A BASH/SLP-76-related adaptor protein MIST/Clnk involved in IgE receptor-mediated mast cell degranulation. Int Immunol 12, 573–80 (2000).

70. Lehman, H.K. Autoimmunity and Immune Dysregulation in Primary Immune Deficiency Disorders. Curr Allergy Asthma Rep 15, 53 (2015).

71. Cartier, L., Hartley, O., Dubois-Dauphin, M. & Krause, K.H. Chemokine receptors in the central nervous system: role in brain inflammation and neurodegenerative diseases. Brain Res Brain Res Rev 48, 16–42 (2005).

72. Versijpt, J.J. et al. Assessment of neuroinflammation and microglial activation in Alzheimer’s disease with radiolabelled PK11195 and single photon emission computed tomography. A pilot study. Eur Neurol 50, 39–47 (2003).

73. Galiegue, S. et al. Cloning and characterization of PRAX-1. A new protein that specifically interacts with the peripheral benzodiazepine receptor. J Biol Chem 274, 2938–52 (1999).

74. Shash, D. et al. Benzodiazepine, psychotropic medication, and dementia: A population-based cohort study. Alzheimers Dement 12, 604–13 (2016).

75. Nanba, D. & Higashiyama, S. Dual intracellular signaling by proteolytic cleavage of membrane-anchored heparin-binding EGF-like growth factor. Cytokine Growth Factor Rev 15, 13–9 (2004).

76. Sasaki, K., Omotuyi, O.I., Ueda, M., Shinohara, K. & Ueda, H. NMDA receptor agonists reverse impaired psychomotor and cognitive functions associated with hippocampal Hbegf-deficiency in mice. Mol Brain 8, 83 (2015).

77. Chapuis, J. et al. Genome-wide, high-content siRNA screening identifies the Alzheimer’s genetic risk factor FERMT2 as a major modulator of APP metabolism. Acta Neuropathol 133, 955–966 (2017).

78. Koerner, I.P. et al. Polymorphisms in the human soluble epoxide hydrolase gene EPHX2 linked to neuronal survival after ischemic injury. J Neurosci 27, 4642–9 (2007).

79. Huffman, J.E. Examining the current standards for genetic discovery and replication in the era of mega-biobanks. Nat Commun 9, 5054 (2018).

80. Purcell, S. et al. PLINK: a tool set for whole-genome association and population-based linkage analyses. Am J Hum Genet 81, 559–75 (2007).

81. Chang, C.C. et al. Second-generation PLINK: rising to the challenge of larger and richer datasets. Gigascience 4, 7 (2015).

82. Chang, C.C. Data Management and Summary Statistics with PLINK. Methods Mol Biol 2090, 49–65 (2020).

83. Price, A.L. et al. Principal components analysis corrects for stratification in genome-wide association studies. Nat Genet 38, 904–9 (2006).

84. Patterson, N., Price, A.L. & Reich, D. Population structure and eigenanalysis. PLoS Genet 2, e190 (2006).

85. Das, S. et al. Next-generation genotype imputation service and methods. Nat Genet 48, 1284–1287 (2016).

86. McCarthy, S. et al. A reference panel of 64,976 haplotypes for genotype imputation. Nat Genet 48, 1279–83 (2016).

87. Loh, P.R. et al. Reference-based phasing using the Haplotype Reference Consortium panel. Nat Genet 48, 1443–1448 (2016).

88. Marchini, J., Howie, B., Myers, S., McVean, G. & Donnelly, P. A new multipoint method for genome-wide association studies by imputation of genotypes. Nat Genet 39, 906–13 (2007).

89. Chen, M.H. & Yang, Q. GWAF: an R package for genome-wide association analyses with family data. Bioinformatics 26, 580–1 (2010).

90. Team., R.D.C. R: a language and environment for statistical computing.

91. Willer, C.J., Li, Y. & Abecasis, G.R. METAL: fast and efficient meta-analysis of genomewide association scans. Bioinformatics 26, 2190–1 (2010).

92. Voorman, A., Brody, J., Chen, H., Lumley, T. & Davis, B. seqMeta: an R package for meta-analyzing region-based tests of rare DNA variants. (2013).

93. de Leeuw, C.A., Mooij, J.M., Heskes, T. & Posthuma, D. MAGMA: generalized gene-set analysis of GWAS data. PLoS Comput Biol 11, e1004219 (2015).

94. Yurko, R., Roeder, K., Devlin, B. & G’Sell, M. H-MAGMA, inheriting a shaky statistical foundation, yields excess false positives. bioRxiv, 2020.08.20.260224 (2020).

95. Ashburner, M. et al. Gene ontology: tool for the unification of biology. The Gene Ontology Consortium. Nat Genet 25, 25–9 (2000).

96. Gene Ontology, C. Gene Ontology Consortium: going forward. Nucleic Acids Res 43, D1049–56 (2015).

97. Kanehisa, M., Sato, Y., Kawashima, M., Furumichi, M. & Tanabe, M. KEGG as a reference resource for gene and protein annotation. Nucleic Acids Res 44, D457–62 (2016).

98. Ogata, H. et al. KEGG: Kyoto Encyclopedia of Genes and Genomes. Nucleic Acids Res 27, 29–34 (1999).

99. Fabregat, A. et al. The Reactome pathway Knowledgebase. Nucleic Acids Res 44, D481–7 (2016).

100. Croft, D. et al. Reactome: a database of reactions, pathways and biological processes. Nucleic Acids Res 39, D691–7 (2011).

101. Nishimura, D. BioCarta. Biotech Software and Internet Report 2, 117–120 (2001).

102. Eppig, J.T. et al. The Mouse Genome Database (MGD): facilitating mouse as a model for human biology and disease. Nucleic Acids Res 43, D726–36 (2015).

103. Pathway Analysis Subgroup of Psychiatric Genomics Consortium. Psychiatric genome-wide association study analyses implicate neuronal, immune and histone pathways. Nat Neurosci 18, 199–209 (2015).

104. Storey, J.D., Bass, A.J., Dabney, A. & Robinson, D. qvalue: Q-value estimation for false discovery rate control. R package version 2.22.0 edn (2020).

105. Ramasamy, A. et al. Genetic variability in the regulation of gene expression in ten regions of the human brain. Nat Neurosci 17, 1418–1428 (2014).

106. Fromer, M. et al. Gene expression elucidates functional impact of polygenic risk for schizophrenia. Nat Neurosci 19, 1442–1453 (2016).

107. Gandal, M.J. et al. Shared molecular neuropathology across major psychiatric disorders parallels polygenic overlap. Science 359, 693–697 (2018).

108. Gibbs, J.R. et al. Abundant quantitative trait loci exist for DNA methylation and gene expression in human brain. PLoS Genet 6, e1000952 (2010).

109. Neueder, A. & Bates, G.P. A common gene expression signature in Huntington’s disease patient brain regions. BMC Med Genomics 7, 60 (2014).

110. Mostafavi, S. et al. A molecular network of the aging human brain provides insights into the pathology and cognitive decline of Alzheimer’s disease. Nat Neurosci 21, 811–819 (2018).

111. Gandal, M.J. et al. Transcriptome-wide isoform-level dysregulation in ASD, schizophrenia, and bipolar disorder. Science 362(2018).

112. Zhang, B. et al. Integrated systems approach identifies genetic nodes and networks in late-onset Alzheimer’s disease. Cell 153, 707–20 (2013).

113. Langfelder, P. & Horvath, S. WGCNA: an R package for weighted correlation network analysis. BMC Bioinformatics 9, 559 (2008).

114. Fairfax, B.P. et al. Innate immune activity conditions the effect of regulatory variants upon monocyte gene expression. Science 343, 1246949 (2014).

115. Raudvere, U. et al. g:Profiler: a web server for functional enrichment analysis and conversions of gene lists (2019 update). Nucleic Acids Res 47, W191–W198 (2019).

116. Madsen, B.E. & Browning, S.R. A groupwise association test for rare mutations using a weighted sum statistic. PLoS Genet 5, e1000384 (2009).

117. Han, F. & Pan, W. A data-adaptive sum test for disease association with multiple common or rare variants. Hum Hered 70, 42–54 (2010).

118. Lee, S. et al. Optimal unified approach for rare-variant association testing with application to small-sample case-control whole-exome sequencing studies. Am J Hum Genet 91, 224–37 (2012).

119. Wu, M.C. et al. Rare-variant association testing for sequencing data with the sequence kernel association test. Am J Hum Genet 89, 82–93 (2011).

120. Pruitt, K.D. et al. RefSeq: an update on mammalian reference sequences. Nucleic Acids Res 42, D756–63 (2014).

121. Kircher, M. et al. A general framework for estimating the relative pathogenicity of human genetic variants. Nat Genet 46, 310–5 (2014).

122. Rentzsch, P., Witten, D., Cooper, G.M., Shendure, J. & Kircher, M. CADD: predicting the deleteriousness of variants throughout the human genome. Nucleic Acids Res 47, D886–D894 (2019).

123. Kircher, M., Rentzsch, P., Witten, D.M., Cooper, G.M. & Shendure, J. Combined Annotation-Dependent Depletion (CADD) Annotation Resource.

124. Gamazon, E.R. et al. A gene-based association method for mapping traits using reference transcriptome data. Nat Genet 47, 1091–8 (2015).

125. Barbeira, A.N. et al. Integrating predicted transcriptome from multiple tissues improves association detection. PLoS Genet 15, e1007889 (2019).

126. Battle, A. et al. Characterizing the genetic basis of transcriptome diversity through RNA-sequencing of 922 individuals. Genome Res 24, 14–24 (2014).

127. Huckins, L.M. et al. Gene expression imputation across multiple brain regions provides insights into schizophrenia risk. Nat Genet 51, 659–674 (2019).

128. Hoffman, G.E. et al. CommonMind Consortium provides transcriptomic and epigenomic data for Schizophrenia and Bipolar Disorder. Sci Data 6, 180 (2019).

129. Consortium, G.T. The GTEx Consortium atlas of genetic regulatory effects across human tissues. Science 369, 1318–1330 (2020).

130. Denny, J.C. et al. PheWAS: demonstrating the feasibility of a phenome-wide scan to discover gene-disease associations. Bioinformatics 26, 1205–10 (2010).

131. Wei, W.Q. et al. Evaluating phecodes, clinical classification software, and ICD-9-CM codes for phenome-wide association studies in the electronic health record. PLoS One 12, e0175508 (2017).

132. Consortium, E.P. An integrated encyclopedia of DNA elements in the human genome. Nature 489, 57–74 (2012).

133. Ernst, J. et al. Mapping and analysis of chromatin state dynamics in nine human cell types. Nature 473, 43–9 (2011).

134. Rao, S.S. et al. A 3D map of the human genome at kilobase resolution reveals principles of chromatin looping. Cell 159, 1665–80 (2014).

135. Ernst, J. & Kellis, M. Chromatin-state discovery and genome annotation with ChromHMM. Nat Protoc 12, 2478–2492 (2017).

136. Heinz, S., Romanoski, C.E., Benner, C. & Glass, C.K. The selection and function of cell type-specific enhancers. Nat Rev Mol Cell Biol 16, 144–54 (2015).

137. Zentner, G.E. & Scacheri, P.C. The chromatin fingerprint of gene enhancer elements. J Biol Chem 287, 30888–96 (2012).

138. Chesi, A. et al. Genome-scale Capture C promoter interactions implicate effector genes at GWAS loci for bone mineral density. Nat Commun 10, 1260 (2019).

139. BioGPS Portal.

140. Brain RNA-Seq Database.

141. Su, A.I. et al. A gene atlas of the mouse and human protein-encoding transcriptomes. Proc Natl Acad Sci U S A 101, 6062–7 (2004).

142. Lein, E.S. et al. Genome-wide atlas of gene expression in the adult mouse brain. Nature 445, 168–76 (2007).

143. Wu, C. et al. Gene set enrichment in eQTL data identifies novel annotations and pathway regulators. PLoS Genet 4, e1000070 (2008).

144. Zhang, Y. et al. An RNA-sequencing transcriptome and splicing database of glia, neurons, and vascular cells of the cerebral cortex. J Neurosci 34, 11929–47 (2014).

145. Zhang, Y. et al. Purification and Characterization of Progenitor and Mature Human Astrocytes Reveals Transcriptional and Functional Differences with Mouse. Neuron 89, 37–53 (2016).

