## Supplementary Files for "Genome-Wide Meta-Analysis of Late-Onset Alzheimer’s Disease Using Rare Variant Imputation in 65,602 Subjects Identifies Novel Rare Variant Locus *NCK2*: The International Genomics of Alzheimer’s Project (IGAP)"

### Supplementary Materials

**Supplementary Note** . . . . . . . . . . . . . . . . . . . . . . . . . . . . . . . . . . . . . . . . . . . . . 3
Supplementary Methods and Dataset Descriptions

**Supplementary Table 1** . . . . . . . . . . . . . . . . . . . . . . . . . . . . . . . . . . . . . . . . . . . Excel
Basic Characteristics of IGAP, EADB, and UK Biobank

**Supplementary Table 2** . . . . . . . . . . . . . . . . . . . . . . . . . . . . . . . . . . . . . . . . . . . Excel
Stage 1-3 All Association Results for Common Variants (MAF≥0.01)

**Supplementary Table 3** . . . . . . . . . . . . . . . . . . . . . . . . . . . . . . . . . . . . . . . . . . . 51
Strongest Regional Common Variant (MAF≥0.01) Association Results in UKBB

**Supplementary Table 4** . . . . . . . . . . . . . . . . . . . . . . . . . . . . . . . . . . . . . . . . . . . Excel
UKBB All Association Results for Common Variants (MAF≥0.01)

**Supplementary Table 5** . . . . . . . . . . . . . . . . . . . . . . . . . . . . . . . . . . . . . . . . . . . Excel
Stage 1-3 Association Results for Rare Variants (MAF<0.01)

**Supplementary Table 6** . . . . . . . . . . . . . . . . . . . . . . . . . . . . . . . . . . . . . . . . . . . 52
Strongest Regional Rare Variant (MAF≤0.01) Association Results in UKBB

**Supplementary Table 7** . . . . . . . . . . . . . . . . . . . . . . . . . . . . . . . . . . . . . . . . . . . Excel
UKBB All Association Results for Rare Variants (MAF≤0.01)

**Supplementary Table 8** . . . . . . . . . . . . . . . . . . . . . . . . . . . . . . . . . . . . . . . . . . . 53
Conditional and Interaction Analysis on GWS-Associated Rare Variants

**Supplementary Table 9** . . . . . . . . . . . . . . . . . . . . . . . . . . . . . . . . . . . . . . . . . . . Excel
Common and Rare Variant Analysis with Weighted UKBB Association Statistics

**Supplementary Table 10** . . . . . . . . . . . . . . . . . . . . . . . . . . . . . . . . . . . . . . . . . . 54
Rare Variant (MAF<0.01) Gene-Based Association Results

**Supplementary Table 11** . . . . . . . . . . . . . . . . . . . . . . . . . . . . . . . . . . . . . . . . . . 56
Rare Variant (MAF<0.01) Gene-Based Association Results

**Supplementary Table 12** . . . . . . . . . . . . . . . . . . . . . . . . . . . . . . . . . . . . . . . . . . Excel
MAGMA Gene-based Association with Common Variants (MAF>0.01) Results

**Supplementary Table 13** . . . . . . . . . . . . . . . . . . . . . . . . . . . . . . . . . . . . . . . . . . Excel
MAGMA Pathway Association with Common Variants (MAF>0.01) Results

**Supplementary Table 14** . . . . . . . . . . . . . . . . . . . . . . . . . . . . . . . . . . . . . . . . . . Excel
MAGMA Top 30 Gene-based Common Variant Association Results (MAF>0.01)

**Supplementary Table 15** . . . . . . . . . . . . . . . . . . . . . . . . . . . . . . . . . . . . . . . . . . Excel
MAGMA Gene-Set Analysis of Co-Expression Modules

**Supplementary Table 16** . . . . . . . . . . Excel
Gene-based Associations for Genes in the Four Most-Associated Co-Expression Modules (Implicating 'Immune Response')

**Supplementary Table 17** . . . . . . . . . . Excel
PrediXcan and MultiXcan Analysis of Genetically-Regulated Gene Expression using GTEx Whole-Body and Brain-Specific Tissue Reference Data

**Supplementary Table 18** . . . . . . . . . . Excel
Variant Effect Predictor (VEP) Annotations for Significantly Associated Variants

### Supplementary Note

#### *Dataset Descriptions*

#### The Alzheimer's Disease Genetics Consortium (ADGC)

The ADGC dataset comprises subjects from 35 datasets including two waves of the Adult Changes in Thought (ACT) cohort study [ACT1/ACT2]; ten waves of cases and cognitively normal controls from the National Institute on Aging (NIA) Alzheimer Disease Centers (ADCs); the Alzheimer Disease Neuroimaging Initiative (ADNI); the Biomarkers of Cognitive Decline Among Normal Individuals (BIOCARD) Cohort; two waves of the Religious Orders Study/Memory and Aging Project (ROSMAP1-2) and the Chicago Health and Aging Project (CHAP) cohort studies at Rush University; the Einstein Aging Study (EAS); the Multi-Site Collaborative Study for Genotype-Phenotype Associations in Alzheimer's Disease (GenADA) Study by GlaxoSmithKline; Mayo Clinic Jacksonville (MAYO) and Rochester (RMAYO) case-control datasets; the Multi-Institutional Research in Alzheimer's Genetic Epidemiology (MIRAGE) study; the NIA Late-Onset Alzheimer's Disease (LOAD) Family Study (NIA-LOAD); the Netherlands Brain Bank (NBB) case-control dataset; the Oregon Health and Science University (OHSU) case-control dataset; the Pfizer case-control dataset; the Texas Alzheimer's Research and Care Consortium (TARCC) dataset; the Translational Genomics Research Institute series 2 (TGEN2) dataset; the University of Miami (UM)/ Case Western Reserve University (CWRU)/ Mt. Sinai School of Medicine (MSSM) and UM/CWRU/TARCC wave 2 datasets [UMCWRUMSSM and UMCWRUTARCC2]; the Universitatsklinikum Saarlandes (UKS) case-control dataset; the University of Pittsburgh (UPITT) case-control dataset; Washington University (WASHU) wave 1 and 2 case-control datasets [WASHU1/WASHU2]; and the Washington Heights-Inwood Community Aging Project (WHICAP) study datasets.

Descriptions of the ACT1, ADC waves 1-7, ADNI, BIOCARD, CHAP, EAS, GenADA, MAYO, MIRAGE, NBB, NIA-LOAD, OHSU, PFIZER, RMAYO, ROSMAP1, ROSMAP2, TARCC, TGEN2, UKS, UM/CWRU/MSSM, UM/CWRU/TARCC2, UPITT, WASHU1, WASHU2, and WHICAP cohorts have been provided in previous ADGC and IGAP studies[1-6]. Here we update descriptions of these studies, where applicable, and provide descriptions for ACT2, ADC wave 8-10, as well as the Combined Small Datasets Collection (CSDC). The CSDC is a harmonized collection of merged small datasets (comprising the existing datasets of ACT2, BIOCARD, CHAP2, EAS, NBB, RMAYO, ROSMAP2, and WASHU2) used in common variant analyses to deal with small case and control counts within the individual studies. All analyses were restricted to individuals of European ancestry. All subjects were recruited under protocols approved by the appropriate Institutional Review Boards (IRBs).

*ACT1/ACT2*: The ACT cohort is an urban and suburban elderly population from a stable HMO that includes 2,581 cognitively intact subjects age ≥ 65 years who were enrolled between 1994 and 1998[7,8]. An additional 811 subjects were enrolled in 2000-2002 using the same methods except oversampling clinics with more minorities. More recently, a Continuous Enrollment strategy was initiated in which new subjects are contacted, screened, and enrolled to keep 2,000 active at-risk person-years accruing in each calendar year. This resulted in an enrollment of 4,146 participants as of May 2009. All clinical data are reviewed at a consensus conference. Dementia onset is assigned half-way between the prior biennial and the exam that diagnosed dementia. A waiver of consent was obtained from the IRB to enroll deceased ACT participants. In total, ACT contributed data on 553 individuals with probable or possible Alzheimer's disease (70 with autopsy-confirmation) and on 1,579 CNEs (155 with autopsy-confirmation) who were included in the analyses, with 2,103 cases/1,571 CNEs in the first wave (ACT1) and 29 cases/8 CNEs in the second wave (ACT2).

*NIA ADC Samples (ADC1-10)*: The NIA ADC cohort included subjects ascertained and evaluated by the clinical and neuropathology cores of the 32 NIA-funded ADCs. Data collection is coordinated by the National Alzheimer's Coordinating Center (NACC). NACC coordinates collection of phenotype data from the 32 ADCs, cleans all data, coordinates implementation of definitions of Alzheimer's disease cases and controls, and coordinates collection of samples. The ADC cohort consists of 3,311 autopsy-confirmed and 2,889 clinically-confirmed Alzheimer's disease cases, and 247 cognitively normal elders (CNEs) with complete neuropathology data who were older than 60 years at age of death, and 3,687 living CNEs evaluated using the Uniform dataset (UDS) protocol[9,10] who were documented to not have mild cognitive impairment (MCI) and were between 60 and 100 years of age at assessment. Based on the data collected by NACC, the ADGC Neuropathology Core Leaders Subcommittee derived inclusion and exclusion criteria for Alzheimer's disease and control samples. All autopsied subjects were age ≥ 60 years at death. Based on the data collected by NACC, the ADGC Neuropathology Core Leaders Subcommittee derived inclusion and exclusion criteria for Alzheimer's disease and control samples. All autopsied subjects were age ≥ 60 years at death. Alzheimer's disease cases were demented according to NINCDS-ADRDA/DSMIV-V or more recent NIA-AA criteria[11,12] or Clinical Dementia Rating (CDR) ≥ 137. Neuropathologic stratification of cases followed NIA/Reagan criteria explicitly or used a similar approach when NIA/Reagan criteria4 were coded as not done, missing, or unknown. Cases were intermediate or high likelihood by NIA/Reagan criteria with moderate to frequent amyloid plaques[13] and neurofibrillary tangle

(NFT) Braak stage of III-VI[14,15]. Persons with Down's syndrome, non-Alzheimer's disease tauopathies and synucleinopathies were excluded. All autopsied controls had a clinical evaluation within two years of death. Controls did not meet NINCDS-ADRDA/DSMIV-V criteria for dementia, did not have a diagnosis of mild cognitive impairment (MCI), and had a CDR of 0, if performed. Controls did not meet or were low-likelihood Alzheimer's disease by NIA/Reagan criteria, had sparse or no amyloid plaques, and a Braak NFT stage of 0 – II. ADCs sent frozen tissue from autopsied subjects and DNA samples from some autopsied subjects and from living subjects to the ADCs to the National Cell Repository for Alzheimer's Disease (NCRAD). DNA was prepared by NCRAD for genotyping and sent to the genotyping site at Children's Hospital of Philadelphia. ADC samples were genotyped and analyzed in separate batches (waves 1-10). The ADC data used in IGAP discovery analyses (ADC1-10) consist of 6,292 cases and 4,980 CNEs in total.

*ADNI*: ADNI is a longitudinal, multi-site observational study including Alzheimer's disease, mild cognitive impairment (MCI), and elderly individuals with normal cognition assessing clinical and cognitive measures, MRI and PET scans (FDG and 11C PIB) and blood and CNS biomarkers. For this study, ADNI contributed data on 268 Alzheimer's disease cases with MRI confirmation of Alzheimer's disease diagnosis and 173 healthy controls with Alzheimer's disease-free status confirmed as of most recent follow-up. Alzheimer's disease subjects were between the ages of 55–90, had an MMSE score of 20–26 inclusive, met NINCDS-ADRDA/ NIA-AA criteria for probable Alzheimer's disease[11,12], and had an MRI consistent with the diagnosis of Alzheimer's disease. Control subjects had MMSE scores between 28 and 30 and a Clinical Dementia Rating of 0 without symptoms of depression, MCI or other dementia and no current use of psychoactive medications. According to the ADNI protocol, subjects were ascertained at regular intervals over 3 years, but for the purpose of our analysis we only used the final ascertainment status to classify case-control status. Additional details of the study design are available elsewhere[5,16,17].

*BIOCARD*: The BIOCARD study is supported by a grant jointly funded by the National Institute on Aging (NIA) and the National Institute of Mental Health (NIMH). The overarching goal of the BIOCARD Study is to identify biomarkers associated with progression from normal cognitive status to cognitive impairment or dementia, with a particular focus on Alzheimer's Disease. Please see Albert et al. 2014[18] for a detailed description of the study. A total of 354 individuals were initially enrolled in the study. Recruitment was conducted by the staff of the Geriatric Psychiatry Branch (GPB) of the intramural program of the NIMH, beginning in 1995 and ending in 2005. The domains of information collected as part of the study include: cognitive testing, magnetic resonance imaging (MRI), cerebrospinal fluid (CSF), amyloid imaging (using PET-PiB), and blood specimens. Investigators at the Johns Hopkins University School of Medicine began evaluating participants in 2009, and subjects are seen annually. At each visit there are assessments of medical and cognitive status, as well as acquisition of MRI, CSF, PET-PiB, and blood. Each subject in the analyses received a consensus diagnosis by a team of neurologists, neuropsychologists, research nurses and research assistants of the BIOCARD Clinical Core at Johns Hopkins with diagnoses based on evidence of clinical or cognitive dysfunction (i.e., individuals with a CDR score > 0 and/or evidence of decline on cognitive testing). To the extent possible, this diagnosis did not use the cognitive test scores. In brief, (1) clinical data relating to the medical, neurologic and psychiatric status of the subject were examined, (2) reports of changes in cognition by the subject and other sources were examined, and (3) decline in cognitive performance was established. Cognitive test scores were used to: (1) determine whether the subject had become cognitively impaired, and (2) determine the likely etiology of such impairment. These diagnostic procedures are comparable to those implemented in the Alzheimer's Disease Centers (ADC) program, supported by the NIA. For this study, BIOCARD contributed data on 6 Alzheimer's disease cases and 112 healthy controls with Alzheimer's disease-free status confirmed as of most recent follow-up.

*CHAP*: CHAP is an on-going community based study of individuals from a geographically defined community of 3 neighborhoods in Chicago, Illinois (Morgan Park, Washington Heights, and Beverly), with 6,158 participants in the first phase of the study[19] (78.7% overall; 80.5% of the blacks, 74.6% of the whites). Data were collected in cycles of approximately 3 years; each consisting of an in-home interview of all participants and clinical evaluation of a random, stratified sample. The baseline cycle measured disease prevalence and provided risk factor data prior to incident disease onset. A cohort of 3,838 persons free of Alzheimer's disease was identified; 729 persons were sampled for baseline clinical evaluation. Persons in the disease-free cohort had either good cognitive function at baseline, or if cognitive function was intermediate or poor, were free from Alzheimer's disease at the baseline clinical evaluation. This disease-free cohort was evaluated for incident disease after an average of 4.1 years. Sampling for incident clinical evaluation was based on age, sex, race, and change in cognitive function (i.e., stable or improved, small decline, or large decline). The sample set available in the ADGC for genetic analyses included 27 Alzheimer's disease cases and 144 persons free of Alzheimer's disease at time of last assessment. All subjects were age 65 years or older at last assessment.

*EAS*: Based at the Albert Einstein College of Medicine, the EAS is an ongoing community based cohort study of cognitive aging and Alzheimer's disease in the elderly which began over four decades ago. Please see Barzilai et al. 2004[20] and Katz et al. 2012[21] for details. The EAS cohort has employed systematic recruiting methods to reduce the selection biases that arise from clinic-based samples and to capture the racial diversity within the Bronx community. Since 1993, a total of 1,944 participants have been enrolled. Between 1993 and 2004, Health Care Financing Administration/Centers for Medicaid and Medicare Services (HCFA/CMS) rosters of Medicare eligible persons aged 70 and above were used to develop sampling frames of community residing participants in Bronx County. Since 2004, New York City Board of Elections registered voter lists for the Bronx have been used due to changes in policies for release of HCFA/CMS rosters. Individuals were mailed introductory letters regarding the study and were then telephoned to complete a brief screening interview. Eligible participants were at least 70 years of age, Bronx residents, non-institutionalized, and English speaking. Exclusion criteria included visual or auditory impairments that preclude neuropsychological testing, active psychiatric symptomatology that interfered with the ability to complete assessments, and non-ambulatory status. Written informed consent was obtained at the initial clinic visit. In-person evaluations were completed at baseline and at subsequent 12-month intervals. Functional status was assessed by the self-administered CERAD C1-ALT, a cognitive/functional impairment instrument, and the Instrumental Activities of Daily Living scale (IADL), a subscale on the Lawton Brody Activities of Daily Living Scale. The score on the IADL was based on 5 domains of function that were common to both elderly men and women. Scores for each domain were dichotomized as impaired vs. not impaired and then the domain scores were summed. If the participant agreed, an informant completed the CERAD C2-ALT, a cognitive/functional impairment instrument, and the Informant Questionnaire on Cognitive Decline in the Elderly (IQ-CODE) 14 forms. The standard neurological physical examination was adapted from the Unified Parkinson's Disease Rating Scale. The evaluation assessed the participant's memory for significant recent events in the news and personal events. The coherence and focus of responses, repetitiveness, and language were determined. When possible, informants were interviewed to ascertain whether they noted any cognitive changes in the participant, and to assess accuracy of the participant's responses. The neurologist also assessed each participant for abnormal behaviors, fluctuation in cognition, and history of sleep disturbance and visual/auditory hallucinations. The neurologist assigned an Hachinski Ischemic Score (HIS), the Clinical Dementia Rating (CDR), and provided a clinical impression of presence or absence of dementia. A diagnosis of dementia was based on standardized clinical criteria from the Diagnostic and Statistical Manual, Fourth Edition (DSM-IV) and required impairment in memory plus at least one additional cognitive domain, accompanied by evidence of functional decline. Diagnoses were assigned at consensus case conferences, which included comprehensive review of cognitive test results, relevant neurological signs and symptoms, and functional status. Memory impairment was defined as scores in the impaired range on any of the memory tests in the neuropsychological battery. (FCSRT $\leq$ 2430 or 1.5 standard deviations (SD) below the age-adjusted mean on Logical Memory) Functional decline was determined at case conference based on information from self or informant report, impairment score on the IADL Lawton Brody Scale, clinical evaluation, and informant questionnaires. Alzheimer's disease was diagnosed in participants with dementia meeting clinical criteria for probable or possible disease established by the NINCDS-ADRDA/ NIA-AA[11,12]. Incident dementia and Alzheimer's disease were diagnosed in persons free of dementia at baseline who met criteria at follow-up. A subset of individuals who participated in the clinical studies of the EAS came to autopsy, providing an important quality control for diagnostic accuracy. A clinical diagnosis of dementia had a positive predictive value (PPV) of 96% for significant pathology upon autopsy. A clinical diagnosis of possible or probable Alzheimer's disease had a PPV of 79% for the presence of NIA-Reagan intermediate or high likelihood Alzheimer type pathology based on an autopsy sample of 175. For this study, EAS contributed data on 9 Alzheimer's disease cases and 141 healthy controls with Alzheimer's disease-free status confirmed as of most recent follow-up.

*GenADA*: GenADA study data analyzed included 666 Alzheimer's disease cases and 712 CNEs ascertained from nine memory referral clinics in Canada between 2002 and 2005. Patients and CNEs were of non-Hispanic White (NHW) ancestry from Northern Europe. All patients with Alzheimer's disease satisfied NINCDS-ADRDA/NIA-AA[11,12] and DSM-IV criteria for probable Alzheimer's disease with Global Deterioration Scale scores of 3-7. CNEs had MMSE test scores higher than 25 (mean 29.2 ± 1.1), a Mattis Dementia Rating Scale score of ≥ 136, a Clock Test without error,

and no impairments on seven instrumental activities of daily living questions from the Duke Older American Resources and Services Procedures test. Data were collected under an academic-industrial grant from Glaxo-Smith-Kline, Canada by Principal Investigator P. St George-Hyslop. Detailed characteristics of this cohort have been described previously[22].

*MAYO*/*RMAYO*: All 671 cases and 1,279 controls consisted of NHW subjects from the United States ascertained at the Mayo Clinic. All subjects were diagnosed by a neurologist at the Mayo

Clinic in Jacksonville, Florida or Rochester, Minnesota. The neurologist confirmed a Clinical Dementia Rating score of 0 for all controls; cases had diagnoses of possible or probable Alzheimer's disease made according to NINCDS-ADRDA/NIA-AA[11,12] criteria. Autopsy-confirmed samples (221 cases, 216 CNEs) came from the brain bank at the Mayo Clinic in Jacksonville, FL and were evaluated by a single neuropathologist. In clinically-identified cases, the diagnosis of definite Alzheimer's disease was made according to NINCDS-ADRDA criteria. All Alzheimer's disease brains analyzed in the study had a Braak score of 4.0 or greater. Brains employed as controls had a Braak score of 2.5 or lower but often had brain pathology unrelated to Alzheimer's disease and pathological diagnoses that included vascular dementia, frontotemporal dementia, dementia with Lewy bodies, multi-system atrophy, amyotrophic lateral sclerosis, and progressive supranuclear palsy.

*MIRAGE:* The MIRAGE study is a family-based genetic epidemiology study of Alzheimer's disease that enrolled Alzheimer's disease cases and unaffected sibling controls at 17 clinical centers in the United States, Canada, Germany, and Greece (details elsewhere[23]), and contributed 1,229 subjects (491 Alzheimer's disease cases and 738 CNEs), a subset of the cases and controls that were incorporated into our prior studies[1,5] which met more stringent QC criteria for this study. Briefly, families were ascertained through a proband meeting the NINCDS-ADRDA/ NIA-AA[11,12] criteria for definite or probable Alzheimer's disease. Unaffected sibling controls were verified as cognitively healthy based on a Modified Telephone Interview of Cognitive Status score ≥ 86[24].

*UMCWRUTARCC2:* The UMCWRUTARCC2 sample included 256 cases and 189 controls from the University of Miami, Case Western Reserve University, and the Texas Alzheimer's Research Care Consortium (wave 2). All Alzheimer's disease cases had onset of disease symptoms after age 65 years and met NINCDS-ADRDA/NIA-AA[11,12] criteria for probable or possible Alzheimer's disease. Controls were adjudicated to have MMSE scores greater than 28 and no clinically identified signs of cognitive impairment. Additional details of subject recruitment at these sites are described in the UM/CWRU/MSSM (formerly UM/VU/MSSM) and TARCC cohort descriptions in this supplement and elsewhere[1,3,4].

*NIA-LOAD*: The NIA LOAD Family Study[25] recruited families with two or more affected siblings with LOAD and unrelated, CNEs similar in age and ethnic background. A total of 1,819 cases and 1,969 CNEs from 1,802 families were recruited through the NIA LOAD study, NCRAD, and the University of Kentucky, with 1,798 cases and 1,568 CNEs included for analysis. One case per family was selected after determining the individual with the strictest diagnosis (definite > probable > possible LOAD). If there were multiple individuals with the strictest diagnosis, then the individual with the earliest age of onset was selected. The controls included only those samples that were neurologically evaluated to be normal and were not related to a study participant.

*NBB*: The Netherlands Brain Bank, which has been previously described elsewhere[3], is a department of the Netherlands Institute for Neuroscience, an institute of the Royal Netherlands Academy of Arts and Sciences. The NBB is a non-profit organization that collects human brain tissue from donors with a variety of neurological and psychiatric disorders and brain tissue from non-diseased donors, as well as anonymized summaries of donors' medical records to be made available for neuroscience research[26]. The sample set available in the ADGC for genetic analyses included 80 pathologically-confirmed Alzheimer's disease cases and 48 subjects free of Alzheimer's pathology at autopsy. All cases were age 65 years or older at time of diagnosis, and all controls were age 65 years or older at time of death.

*OHSU*: The OHSU dataset includes 132 autopsy-confirmed Alzheimer's disease cases and 153 deceased controls that were evaluated for dementia within 12 months prior to death (age at death > 65 years), which are a subset of the 193 cases and 451 controls examined in our previous study[5] meeting more stringent QC criteria in this study. Subjects were recruited from aging research cohorts at 10 NIA-funded ADC, and did not overlap other samples assembled by the ADGC. A more extensive description of control samples can be found elsewhere[27].

*Pfizer:* The Pfizer sample collection comprises Alzheimer's disease cases taken from the Lipitor's Effect in Alzheimer's Disease (LEADe) trial, including subjects who converted to Alzheimer's disease after ascertainment as MCI, as well as 216 probable Alzheimer's disease subjects enrolled by PrecisionMed for a case-control study and 149 subjects from a Phase II trial (#A3041005) of CP-457920 (a selective α5 GABAA receptor inverse agonist) in Alzheimer's disease. Samples were collected from multiple clinical sites, and with appropriate IRB/ethics committee approvals at each individual site, with written and informed consent given by subjects for use in follow-up studies. All subjects were diagnosed with probable or possible Alzheimer's disease if they met NINCDS-ADRDA/NIA-AA[11,12] and/or DSM-IV criteria, and had Mini-Mental Status Exam (MMSE) scores < 25 at baseline. The control group included subjects from two studies: 1) the PrecisionMed case-control study (#A9010012), which recruited elderly subjects free of neurological or psychiatric conditions, and 2) 999-GEN-0583-001, which obtained a reference population of cognitively, neurologically, and psychiatrically normal subjects. Controls have no neuropsychiatric conditions or diseases and had MMSE>27 at the time of enrollment. For Alzheimer's disease analysis, all cases with age-at-onset (AAO) less than 65 years were removed to exclude early-onset Alzheimer's disease subjects. All controls were re-matched with remaining cases according to gender, age (all controls are older than cases), and ethnicity (only individuals with NHW background were analyzed). The final Pfizer Alzheimer's disease case-control GWAS dataset included 696 cases and 762 controls. Cases from the PrecisionMed/ A3041005 and LEADe studies and age-matched controls were genotyped using the Illumina HumanHap550 array. *APOE* genotypes were determined from genotypes for rs429358 and rs7412 obtained using Taqman assays.

*ROSMAP*: ROSMAP are two community-based cohort studies. The ROS has been ongoing since 1993, with a rolling admission. Through July of 2010, 1,139 older nuns, priests, and brothers from across the United States initially free of dementia who agreed to annual clinical evaluation and brain donation at the time of death completed their baseline evaluation. The MAP has been on-going since 1997, also with a rolling admission. Through July of 2010, 1,356 older persons from across northeastern Illinois initially free of dementia who agreed to annual clinical evaluation and organ donation at the time of death completed their baseline evaluation. Details of the clinical and neuropathologic evaluations have been previously reported[28-30]. A total of 1,064 persons passed genotyping QC. Of these, 295 met clinical criteria for Alzheimer's disease at the time of their last clinical evaluation or time of death and met neuropathologic criteria for

Alzheimer's disease for those on whom neuropathologic data were available, and 769 were without dementia or MCI at the time of their last clinical evaluation or time of death and did not meet neuropathologic criteria for Alzheimer's disease for those on whom neuropathologic data were available. A second wave of ROSMAP (referred to as ROSMAP2 in this study) included 59 persons who met clinical criteria for Alzheimer's disease at the time of their last clinical evaluation or time of death and met neuropathologic criteria for Alzheimer's disease for those on whom neuropathologic data were available, and 217 persons who were without dementia or MCI at the time of their last clinical evaluation or time of death and did not meet neuropathologic criteria for Alzheimer's disease for those on whom neuropathologic data were available.

*TARCC*: The TARCC is a collaborative Alzheimer's research effort directed and funded by the Texas Council on Alzheimer's Disease and Related Disorders (the Council), as part of the Darrell K Royal Texas Alzheimer's Initiative. Composed of Baylor College of Medicine (BCM), Texas Tech University Health Sciences Center (TTUHSC), University of North Texas Health Science Center (UNTHSC), the UT Southwestern Medical Center at Dallas (UTSW), University of Texas Health Science Center at San Antonio (UTHSCSA), Texas A&M Health Science Center (TAMHSC), and the University of Texas at Austin (UTA), this consortium was created to establish a comprehensive research cohort of well characterized subjects to address better diagnosis, treatment, and ultimately prevention of Alzheimer's disease[31]. The resulting prospective cohort, the Texas Harris Alzheimer's Research Study, contains clinical, neuropsychiatric, genetic, and blood biomarker data on more than 3,000 participants diagnosed with Alzheimer's disease, mild cognitive impairment (MCI), and cognitively normal individuals. Longitudinal data/sample collection and follow-up on participants occurs on an annual basis.

Two waves of case-control data from TARCC were examined as part of genetic analyses in the ADGC. Data from the TARCC included 323 cases and 181 controls in the first wave (included in the TARCC1 cohort), with 84 cases and 115 controls in the second wave (included in the MTC cohort). All TARCC subjects were greater than 65 years of age at disease onset (cases) or at last disease-free exam (non-cases).

*TGEN2*: Among the TGEN2 data analyzed were 668 clinically- and neuropathologically-characterized brain donors, and 365 CNEs without dementia or significant Alzheimer's disease pathology. Of these cases and CNEs, 667 were genotyped as a part of the TGEN1 series[32]. Samples were obtained from twenty-one different National Institute on Aging-supported Alzheimer's disease Center brain banks and from the Miami Brain Bank as previously described[32-35]. Additional individual samples from other brain banks in the United States, United Kingdom, and the Netherlands were also obtained in the same manner. The criteria for inclusion were as follows: self-defined ethnicity of European descent, neuropathologically confirmed Alzheimer's disease or neuropathology present at levels consistent with status as a control, and age of death greater than 65. Autopsy diagnosis was performed by board-certified neuropathologists and was based on the presence or absence of the characterization of probable or possible Alzheimer's disease. Where possible, Braak staging and/or CERAD classification were employed. Samples derived from subjects with a clinical history of stroke, cerebrovascular disease, comorbidity with any other known neurological disease, or with the neuropathological finding of Lewy bodies were excluded.

*UKS*: The UKS cohort is a thoroughly diagnosed case-control cohort from Universitatsklinikum

Saarlandes, consisting of individuals clinically diagnosed with sporadic Alzheimer's disease (N = 596; mean age onset, 72.2 ± 6.6 years) and cognitively healthy, age-, gender-, and ethnicity-matched population-based controls (N = 170; 64.1 ± 3.0 years).

*UM/CWRU/MSSM*: The UM/CWRU/MSSM dataset (formerly UM/VU/MSSM[36-39]) contains 1,177 cases and 1,126 CNEs ascertained at the University of Miami, Case Western Reserve University and Mt. Sinai School of Medicine, including 409 autopsy-confirmed cases and 136 controls, primarily from the Mt. Sinai School of Medicine[40]. An additional 16 cases were included and 34 controls excluded from the data analyzed in the Jun et al. 2010 study[5]. Each affected individual met NINCDS-ADRDA/NIA-AA[11,12] criteria for probably or definite Alzheimer's disease with age at onset greater than 60 years as determined from specific probe questions within the clinical history provided by a reliable family informant or from documentation of significant cognitive impairment in the medical record. Cognitively healthy controls were unrelated individuals from the same catchment areas and frequency matched by age and gender, and had a documented MMSE or 3MS score in the normal range. Cases and controls had similar demographics: both had similar ages-at-onset/ages-at-exam of 71.1 (±17.4 SD) for cases and 73.5 (±10.6 SD) for controls, and cases and controls were 64.5% and 61.3% female, respectively.

*UPITT*: The University of Pittsburgh dataset contains 1,255 NHW Alzheimer's disease cases (of which 277 were autopsy-confirmed) recruited by the University of Pittsburgh Alzheimer's Disease Research Center, and 829 NHW, CNEs ages 60 and older (2 were autopsy-confirmed). All Alzheimer's disease cases met NINCDS-ADRDA/NIA-AA[11,12] criteria for probable or definite Alzheimer's disease. Additional details of the cohort used for GWAS have been previously published[41].

*WASHU*: An NHW LOAD case-control dataset consisting of 377 cases and 281 healthy elderly controls was used in analyses for this study. This dataset was split between two analysis datasets (WASHU1 and WASHU2). Participants were recruited as part of a longitudinal study of healthy aging and dementia. Diagnosis of dementia etiology was made in accordance with standard criteria and methods[10]. Severity of dementia was assessed using the Clinical Dementia Rating scale[42].

*WHICAP*: WHICAP[43,44] is a community-based longitudinal study of aging and dementia among elderly, urban-dwelling residents. Beginning enrolment in 1989, WHICAP has followed more than 5,900 residents over 65 years of age, including white, African American, and Hispanic participants. Detailed clinical assessments were performed at approximately 24-month intervals over the 7 years of the initial study. All interviews were conducted in either English or Spanish. The choice of language was decided by the subject to ensure the best performance, and the majority of assessments were performed in the subject's home, which included medical, neurological, and neuropsychological evaluations. Results of the neurological, psychiatric, and neuropsychological assessments were reviewed in a consensus conference comprised of neurologists, psychiatrists, and neuropsychologists. Based on this review all participants were assigned to one of three categories: dementia, cognitive impairment, or normal cognitive function. The sample set available in the ADGC for genetic analyses included 73 Alzheimer's disease cases and 560 subjects with normal cognitive function.

---

*CSDC*: The Combined Small Datasets Collection is a harmonized dataset including all data from eight separately ascertained datasets or waves of datasets already described above (ACT2, BIOCARD, CHAP2, EAS, NBB, RMAYO, ROSMAP2, and WASHU2). None of these datasets were separately incorporated into IGAP analyses, and were only analyzed in the CSDC. Datasets or waves were incorporated into the CSDC if they included fewer than 100 cases and/or 100 CNEs. As all datasets were genotyped separately but imputed to the same dataset, genotyped SNPs overlapping all of the high-density genotyping platforms used were extracted and a set of ~20,000 variants were used to estimate both population substructure and explore potential heterogeneity between datasets using principal components analysis (PCA) in the 'smartpca.pl' program in EIGENSTRAT/EIGENSOFT[45,46]. Data-set level association analyses similar to those described for all other cohorts and datasets were performed, though covariate adjustment additionally included indicator variables for study to adjust for residual batch effects not captured in PCs. Association results among common variants (minor allele frequency [MAF]>0.01) in the CSDC were similar to other datasets in the ADGC with very modest inflation ($\lambda$=1.01) while rare variant associations demonstrated extreme patterns of association with a high degree of genomic inflation ($\lambda$=1.22). For this reason, only common variant association results from the CSDC were incorporated common variant meta-analysis, while rare variant association results were excluded from rare variant meta-analysis (none of the eight individual datasets composing the CSDC were used in rare variant meta-analysis).

### The Cohort for Heart and Ageing Research in Genomic Epidemiology (CHARGE) Consortium

The CHARGE consortium currently includes several large, prospective, community-based cohort studies that have genome-wide variation data coupled with extensive data on multiple phenotypes[47]. A neurology working-group arrived at a consensus on phenotype harmonization, covariate selection and analytic plans for within-study analyses and meta-analysis of results[48]. Consent procedures, examination and surveillance components, data security, genotyping protocols and study design at each study were approved by a local Institutional Review Board, details are provided below. Three of the studies, the Cardiovascular Health Study (CHS), the Framingham Heart Study (FHS) and the Rotterdam Study had data on both the baseline prevalence and on incident Alzheimer's disease, whereas the Atherosclerosis Risk in Communities Study (ARIC) only had data on prevalent Alzheimer's disease.

*ARIC.* The ARIC study is a population-based cohort study of atherosclerosis and clinical atherosclerotic diseases[49]. Starting with a baseline data collection exam over 1987-1989, 15,792 individuals aged 45-64 years were recruited from four U.S. communities: suburban Minneapolis, MN; Washington County, MD; Forsyth County, NC; and Jackson, MS. Since the baseline exam, there have been five subsequent follow-up visits with neurocognitive functioning assessed in visit 2 (1990-1992), visit 4 (1996-1998), visit 5 (2011-2013), and visit 6 (2016 – 2017).

At baseline, blood was drawn for DNA extraction and participants consented to genetic testing. Genome-wide genotyping was conducted at the Broad Institute using the Affymetrix 6.0 SNP Array. Genotyping calling was performed using Birdseed for 9,747 European Americans. Imputation was performed on the QCed data on the Michigan Imputation Server in two steps:

pre-phasing with EAGLE and imputation with MiniMac3, using the HRC r1.1 data reference panel.

Cognitive diagnoses were adjudicated at the fifth visit using cognitive, neurologic, and brain imaging assessments (comprehensive diagnostic details are available elsewhere[49]). Cognitive status was not available on the whole sample at the third visit. The ARIC study has been approved by the Institutional Review Board at each field center, including Wake Forest Baptist Medical Center (Forsyth County, NC), University of Mississippi Medical Center (Jackson, MS), University of Minnesota (suburban Minneapolis, MN), and Johns Hopkins University (Washington County, MD). Participants provided written informed consent prior to each examination.

*CHS.* The CHS is a prospective population-based cohort study of risk factors for vascular and metabolic disease that in 1989-90, enrolled adults aged ≥65 years, at four field centers located in North Carolina, California, Maryland, and Pennsylvania[50]. The original predominantly NHW ancestry cohort of 5,201 persons was recruited from a random sample of people on Medicare eligibility lists and an additional 687 African Americans were enrolled subsequently for a total sample of 5,888[50]. Deoxyribonucleic acid (DNA) was extracted from blood samples drawn on all persons who consented to genetic testing at their baseline examination in 1989-90. In 2007-2008, genotyping was performed at the General Clinical Research Center's Phenotyping/Genotyping Laboratory at Cedars-Sinai using the Illumina 370CNV Duo® BeadChip system on 3,980 CHS participants who were free of cardiovascular disease (CVD) at baseline. The 1,908 persons excluded for prevalent CVD had prevalent coronary heart disease (n=1,195), congestive heart failure (n=86), peripheral vascular disease (n=93), valvular heart disease (n=20), stroke (n=166) or transient ischemic attack (n=56). Some persons had more than one reason to be excluded and for these individuals only the initial exclusionary event is listed. Because the other cohorts were predominantly white, the African American CHS participants were excluded from this analysis to limit errors secondary to population stratification. Among white participants genotyping was attempted in 3,397 participants and was successful in 3,295 persons. A total of 742 persons were excluded as they either died prior to the start of the CHS cognition study in 1992 (see section 3 for details), or could not be evaluated completely for baseline cognitive status, leaving a baseline sample of 2,553 persons; an additional 31 persons were excluded for having dementia other than Alzheimer's disease leaving a study sample of 2,522 persons. The CHS study protocols were approved by the Institutional review boards at the individual participating centers.

The Alzheimer's disease sample for this study included all prevalent cases identified in 1992 and incident events identified between 1992 and December 2006[51]. Briefly, persons were examined annually from enrollment to 1999, and the examination included a 30-minute screening cognitive battery. In 1992-94 and again in 1997-99, participants were invited to undergo brain MRI and detailed cognitive and neurological assessment as part of the CHS Cognition Study. Persons with prevalent dementia were identified, and all others were followed until 1999 for the development of incident dementia and Alzheimer's disease. Since then, CHS participants at the Maryland and Pennsylvania centers have remained under ongoing dementia surveillance[52].

Beginning in 1988/89, all participants completed the Modified Mini-Mental State Examination (3MSE) and the DSST at their annual visits, and the Benton Visual Retention Test (BVRT) from 1994 to 1998. The Telephone Interview for Cognitive Status (TICS) was used when participants did not come to the clinic. Further information on cognition was obtained from proxies using the Informant Questionnaire for Cognitive Decline in the Elderly (IQCODE), and the dementia questionnaire (DQ). Symptoms of depression were measured with the modified version of the Center for Epidemiology Studies Depression Scale (CES-D). In 1991-94, 3608 participants had an MRI of the brain and this was repeated in 1997-98. The CHS staff also obtained information from participants and next-of-kin regarding vision and hearing, the circumstances of the illness, history of dementia, functional status, pharmaceutical drug use, and alcohol consumption. Data on instrumental activities of daily living (IADL), and activities of daily living (ADL) were also collected.

Persons suspected to have cognitive impairment based on the screening tests listed above underwent a neuropsychological and a neurological evaluation. The neuropsychological battery included the following tests: the American version of the National Reading test (AMNART), Raven's Coloured Progressive Matrices, California Verbal Learning Test (CVLT), a modified Rey-Osterreith figure, the Boston Naming test, the Verbal fluency test, the Block design test, the Trails A and B tests, the Baddeley & Papagno Divided Attention Task, the Stroop, Digit Span and Grooved Pegboard Tests. The results of the neuropsychological battery were classified as normal or abnormal (>1.5 standard deviations below individuals of comparable age and education) based on normative data collected from a sample of 250 unimpaired subjects. The neurological exam included a brief mental status examination, as well as a complete examination of other systems. The examiner also completed the Unified Parkinson's Disease Rating Scale (UPDRS) and the Hachinski Ischemic Scale. After completing the neurological exam, the neurologist classified the participant as normal, having mild cognitive impairment (MCI), or dementia. International diagnostic guidelines, including the NINCDS-ADRDA criteria for probable and possible Alzheimer's disease and the ADDTC's State of California criteria for probable and possible vascular dementia (VaD) with or without Alzheimer's disease, were followed. CHS identified 3 subtypes: possible/probable Alzheimer's disease without VaD (categorized as pure Alzheimer's disease, included in all Alzheimer's disease) and mixed Alzheimer's disease (for cases that met criteria for both Alzheimer's disease and VaD, included in all-Alzheimer's disease), and, possible/probable VaD without Alzheimer's disease (excluded from current study). For this study, CHS contributed data on 450 Alzheimer's disease cases and 1,702 healthy controls with Alzheimer's disease-free status confirmed as of most recent follow-up.

*FHS.* The FHS is a three-generation, single-site, community-based, ongoing cohort study that was initiated in 1948. It now comprises three generations of participants including the Original cohort followed since 1948 (n=5,209)[53], their Offspring and spouses of the offspring (n=5,216) followed since 1971[54]; and children from the largest Offspring families enrolled in 2000 (Gen 3)[55].

Participants in the Original and Offspring cohorts are used in these analyses, but Gen 3 participants were not included since they are young (mean age 40±9 years in 2000) and none had developed Alzheimer's disease. The Original cohort enrolled 5,209 men and women who comprised two-thirds of the adult population then residing in Framingham, Massachusetts. Survivors continue to receive biennial examinations. The Offspring cohort comprises 5,124 persons (including 3,514 biological offspring) who have been examined approximately once every 4 years. Almost all the FHS Original and Offspring participants are NHW. FHS participants had DNA extracted and provided consent for genotyping in the 1990s. All available eligible participants were genotyped at Affymetrix (Santa Clara, CA) through an NHLBI funded

SNP-Health Association Resource (SHARe) project using the Affymetrix GeneChip® Human Mapping 500K Array Set and 50K Human Gene Focused Panel®. In 272 persons, small amounts of DNA were extracted from stored whole blood and required whole genome amplification prior to genotyping. Cell lines were available for most of the remaining participants. Genotyping was attempted in 5,293 Original and Offspring cohort participants, and 4,425 persons met QC criteria. Failures (call rate<97%, extreme heterozygosity or high Mendelian error rate) were largely restricted to persons with whole-genome amplified DNA and DNA extracted from stored serum samples. In addition, since the persons with whole genome amplified DNA represent a group of survivors who may differ from the others we included whole genome amplified status as a covariate in FHS analyses. For the prevalent analyses, we also excluded 2,268 participants who were less than 65 years old at the time of the DNA draw and 14 persons with dementia other than Alzheimer's disease; the remaining 2,143 subjects constitute the FHS sample for the prevalent study. A total of 806 well-genotyped persons from the Original cohort (which has been under ongoing surveillance for incident dementia since 1975) were included in the incident Alzheimer's disease analyses. The FHS component of this study was approved by the Institutional Review Board of the Boston Medical Center.

The Original cohort of the FHS has been evaluated biennially since 1948, was screened for prevalent dementia and Alzheimer's disease in 1974-76 and has been under surveillance for incident dementia and Alzheimer's disease since then[56-58]. The Offspring have been examined once every 4 years and have been screened for prevalent dementia with a neuropsychological battery and brain MRI[59,60]. In order to be consistent with the sampling frame for the AGES and CHS samples, we excluded FHS subjects with a baseline age <65 yrs at the time of DNA draw which was in the 1990s. To minimize survival biases, Original cohort and Offspring participants who developed dementia prior to the date of DNA draw were treated as prevalent cases, and subsequent events in the Original cohort occurring prior to December 2006 were included in the incident analyses.

At each clinic exam, participants receive questionnaires, physical examinations and laboratory testing; between examinations they remain under surveillance (regardless of whether or not they live in the vicinity) via physician referrals, record linkage and annual telephone health history updates. Methods used for dementia screening and follow-up have been previously described[56,61]. Briefly, surviving cohort members who attended biennial examination cycles 14 and 15 (May 1975-November 1979) were administered a standardized neuropsychological test battery to establish a dementia-free cohort.

Beginning at examination cycle 17 (1982), the MMSE was administered biennially to the cohort. A MMSE score below the education-specific cutoff score, a decline of three or more points on subsequent administrations, a decline of more than five points compared with any previous examination, or a physician or family referral prompted further in-depth testing. The Offspring cohort that was enrolled in 1971 has undergone eight re-examinations, one approximately every four years. Starting at the second Offspring examination, participants were questioned regarding any subjective memory complaints and since the fifth Offspring examination participants have been administered the MMSE at each visit. In addition concurrent with the seventh and eighth Offspring examinations (between 1999 and 2004 and then again between 2005 and 2009) surviving Original cohort and all eligible and consenting Offspring participants have undergone volumetric brain MRI and neuropsychological testing[59,60]. The neuropsychological test battery included the Reading subtest of the Wide Range Achievement Test (WRAT-3), the Logical Memory and the Paired Associates Learning tests from the

Wechsler Memory Scale, the Visual Reproduction and Hooper Visual Organization Tests, Trails A and B, the Similarities subtest from the Wechsler Adult Intelligence test, the 30-iterm version of the Boston Naming Test and at the second assessment only, the Digit Span, Controlled Word Association and Clock Drawing Tests. Offspring participants suspected to have cognitive impairment based on their MMSE scores, participant, family or physician referral, hospital records or performance in the neuro-psychological test battery described above were referred for more detailed neuropsychological and neurological evaluation.

Each participant thus identified underwent baseline neurologic and neuropsychological examinations. Neurologists (trained in geriatric behavioral assessment) supplemented their clinical assessment with a few structured cognitive tests and administered the Clinical Dementia Rating (CDR). Persons were reassessed systematically for the onset of at least mild dementia. A panel consisting of at least 1 neurologist (S.A., P.A.W., or S.S.) and 1 neuropsychologist (R.A.) reviewed all available medical records to arrive at a final determination regarding the presence or absence of dementia, the date of onset of dementia, and the type of dementia. For this determination, we used data from the neurologist's examination, neuropsychological test performance, Framingham Study records, hospital records, information from primary care physicians, structured family interviews, computed tomography and magnetic resonance imaging records, and autopsy confirmation when available. All individuals identified as having dementia satisfied the DSM-IV criteria, had dementia severity equivalent to a CDR of 1 or greater, and had symptoms of dementia for at least 6 months. All individuals identified as having Alzheimer-related dementia met the NINCDS-ADRDA criteria for definite, probable, or possible Alzheimer's disease. Vascular Dementia was diagnosed using the ADDTC criteria, but the presence of vascular dementia did not disqualify a participant from obtaining a concomitant diagnosis of Alzheimer's disease if indicated. The recruitment of Original cohort participants at FHS had occurred long before the DNA collection with the result that the majority of dementia events in the FHS (although ascertained prospectively) were prevalent at the time of DNA collection or these persons had died prior to DNA draw and were thus excluded from analyses of incident disease. Due to the limited number of incident dementia and Alzheimer's disease events in the Framingham Offspring only the Original cohort were included in our analyses of incident events. For this study, FHS contributed data on 330 Alzheimer's disease cases and 3,910 healthy controls with Alzheimer's disease-free status confirmed as of most recent follow-up.

*RS1* and *RS2.* RS1 enrolled inhabitants from a district of Rotterdam (Ommoord) aged≥55 years (N=7,983, virtually all white) at the baseline examination in 1990-93 when blood was drawn for genotyping[62,63]. It aims to examine the determinants of disease and health in the elderly with a focus on neurogeriatric, cardiovascular, bone, and eye disease.8 All inhabitants of Ommoord aged≥55 years (n=10,275) were invited to participate and the participation rate was 78%. All participants gave written informed consent to retrieve information from treating physicians. Baseline measurements were obtained from 1990 to 1993 and consisted of an interview at home and two visits to the research center for physical examination. This cohort was extended with 3,011 participants who had become 55 years of age or had moved into the district since the start of the study (RS2).

Survivors have been re-examined three times: in 1993-1995, 1997-1999, and 2002-2004. All persons attending the baseline examination in 1990-93 consented to genotyping and had DNA extracted. This DNA was genotyped using the Illumina Infinium II HumanHap550chip v3·0® array in 2007-2008 according to the manufacturer's protocols. Genotyping was attempted in persons with high-quality extracted DNA (n=6,449). From these 6,449, samples with low call rate (<97.5%, n=209), with excess autosomal heterozygosity (> 0.336, n=21), with sex-mismatch (n=36), or if there were outliers identified by the IBS clustering analysis (>3 standard deviations from population mean, n=102 or IBS probabilities > 97%, n=129) were excluded from the study population with some persons meeting more than one exclusion criterion; in total, 5,974 samples were available with good quality genotyping data, 42 persons were excluded since they did not undergo cognitive screening at baseline, hence their cognitive status was uncertain. An additional 61 persons were excluded because they suffered from dementia other than Alzheimer's disease at baseline. Thus there were 5,871 persons included in the prevalent analysis and after exclusion of 171 persons with prevalent Alzheimer's disease, 5,700 persons were followed for incident Alzheimer's disease and other dementia. The Rotterdam Study (including its brain magnetic resonance imaging (MRI) and neurological components) has been approved by the institutional review board (Medical Ethics Committee) of the Erasmus Medical Center and the Netherlands Ministry of Health, Welfare and Sports.

Participants were screened for prevalent dementia in 1990-93 using a three-stage process; those free of dementia remained under surveillance for incident dementia, a determination made using records linkage and assessment at three subsequent re-examinations[64]. We included all prevalent cases and all incident events up to 31st December 2004[65,66].

Screening was done with the MMSE and GMS organic level for all persons. Screen-positives (MMSE <26 or Geriatric Mental Schedule (GMS) organic level >0) underwent the CAMDEX. Persons who were suspected of having dementia underwent more extensive neuropsychological testing. When available, imaging data were used. In addition, all participants have been continuously monitored for major events (including dementia) through automated linkage of the study database with digitized medical records from general practitioners, the Regional Institute for Outpatient Mental Health Care and the municipality. In addition, physician files from nursing homes and general practitioner records of participants who moved out of the Ommoord district were reviewed twice a year. For suspected dementia events, additional information (including neuroimaging) was obtained from hospital records and research physicians discussed available information with a neurologist experienced in dementia diagnosis and research to verify all diagnoses. Dementia was diagnosed in accordance with internationally accepted criteria for dementia (Diagnostic and Statistical Manual of Mental Disorders, Revised Third Edition (DSM- III-R), and Alzheimer's disease using the NINCDS-ADRDA criteria for possible, probable and definite Alzheimer's disease. The National Institute of Neurological Disorders and Stroke–Association Internationale pour la Recherche et l'Enseignement en Neurosciences (NINDS-AIREN) criteria were used to diagnose vascular dementia. The final diagnosis was determined by a panel of a neurologist, neurophysiologist, and research physician and the diagnoses of Alzheimer's disease and VaD were not mutually exclusive. Rotterdam contributed data on 985 Alzheimer's disease cases and 4,985 healthy controls with Alzheimer's disease-free status confirmed as of most recent follow-up.

**European Alzheimer's Disease Initiative (EADI) Consortium**

All the 2,328 Alzheimer's disease cases were ascertained by neurologists from Bordeaux, Dijon, Lille, Montpellier, Paris, and Rouen. Clinical diagnosis of probable Alzheimer's disease was established according to the DSM-III-R and NINCDS-ADRDA criteria. Controls were selected from the 3C Study[66]. This cohort is a population-based, prospective study of the relationship between vascular factors and dementia. It has been carried out in three French cities: Bordeaux (southwest France), Montpellier (southeast France) and Dijon (central eastern France). A sample of non-institutionalized subjects over 65 years of age was randomly selected from the electoral rolls of each city. Between January 1999 and March 2001, 9,686 subjects meeting the inclusion criteria agreed to participate. Following recruitment, 392 subjects withdrew from the study. Thus, 9,294 subjects were finally included in the study (2,104 in Bordeaux, 4,931 in Dijon and 2,259 in Montpellier). AD status was defined based on 12 years of follow up. The Alzheimer's disease cases from 3C were included as cases in the EADI discovery dataset and the other individuals were retained as controls. Genomic DNA samples of 7,200 individuals were transferred to the French Centre National de Génotypage (CNG). First stage samples that passed DNA quality control were genotyped with Illumina Human 610-Quad BeadChips.

**Genetic and Environmental Risk in AD (GERAD) Consortium / Defining Genetic, Polygenic, and Environmental Risk for Alzheimer's Disease (PERADES) Consortium**

The GERAD/PERADES sample comprises 3,177 Alzheimer's disease cases and 7,277 controls with available age and gender data[67]. Cases and elderly screened controls were recruited by the Medical Research Council (MRC) Genetic Resource for Alzheimer's disease (Cardiff University; Institute of Psychiatry, London; Cambridge University; Trinity College Dublin), the Alzheimer's Research Trust (ART) Collaboration (University of Nottingham; University of Manchester; University of Southampton; University of Bristol; Queen's University Belfast; the Oxford Project to Investigate Memory and Ageing (OPTIMA), Oxford University); Washington University, St Louis, United States; MRC PRION Unit, University College London; London and the South East Region Alzheimer's disease project (LASER-AD), University College London; Competence Network of Dementia (CND) and Department of Psychiatry, University of Bonn,

Germany; the National Institute of Mental Health (NIMH) Alzheimer's disease Genetics Initiative. 6129 population controls were drawn from large existing cohorts with available GWAS data, including the 1958 British Birth Cohort (1958BC) (http://www.b58cgene.sgul.ac.uk), the KORA F4 Study and the Heinz Nixdorf Recall Study. All Alzheimer's disease cases met criteria for either probable (NINCDS-ADRDA, DSM-IV) or definite (CERAD) Alzheimer's disease. All elderly controls were screened for dementia using the MMSE or ADAS-cog, were determined to be free from dementia at neuropathological examination or had a Braak score of 2.5 or lower. Genotypes from all cases and 4617 controls were previously included in the AD GWAS by Harold and colleagues[67]. Genotypes for the remaining 2,660 population controls were obtained from WTCCC2.

**The University of Bonn Study**

AD cases and cognitive controls from Bonn, Germany, where ascertained through the Competence Network of Dementia (CND), the Department of Psychiatry of the University of Bonn, and through the Heinz Nixdorf Recall Study independently of samples collected in other IGAP and EADB cohorts. All samples were genotyped using the Illumina Omni1-Quad chip. Samples with a genotype call rate worse than 99% or identified as outliers by PCA were excluded. We identified familial relationships and sample duplications using pairwise identity-by-state (IBS) estimation and removed one individual from each pair with an IBS that was 4 standard deviations higher than the average IBS of all pairs of individuals. AD patients overlapping those in the original GERAD study were excluded from analysis. AD diagnoses were made according to the NINCDS-ADRDA/NIA-AA criteria.

**The European Alzheimer's Disease Biobank (EADB)**

The European Alzheimer's Disease Biobank (EADB)[68] dataset consortium groups together AD cases and controls from 15 European countries (Belgium, Bulgaria, Czech Republic, Denmark, Finland, France, Germany, Greece, Italy, Portugal, Spain, Sweden, Switzerland, The Netherlands and the UK). These samples were genotyped in three independent centers (France, Germany and the Netherlands) leading to define three nodes: EADB-France, EADB-Germany and EADB-Netherlands. In addition, EADB also included Australian partners.

***EADB-France.*** In the EADB-France node, samples were collected from nine countries (39 centers/studies), and after quality controls (QCs), we obtained 13,867 AD cases and 15,310 controls. All these samples were genotyped at the Centre National de Recherche en Génomique Humaine (CNRGH, Evry, France).

***Belgium.*** The participants were part of a large prospective cohort[69] of Belgian AD patients and healthy elderly control individuals. The patients were ascertained at the memory clinic of Middelheim and Hoge Beuken (Hospital Network Antwerp, Belgium) and at the memory clinic of the University Hospitals of Leuven, Belgium. The control individuals were the partners of the patients or volunteers from the Belgian community. The study protocols were approved by the ethics committees of the Antwerp University Hospital and the participating neurological centers at the different hospitals of the BELNEU consortium and by the University of Antwerp.

***Czech Republic.*** The Czech Brain Aging Study (CBAS)[70] is a longitudinal memory-clinic–based study recruiting subjects at risk of dementia (subjects referred for cognitive complaints—SCD, MCI). The CBAS+ study is a cross-sectional study of patients in the early stages of dementia. All subjects signed informed consent and both studies were approved by the local ethics committee.

***Denmark.*** The Copenhagen General Population Study (CGPS) is a prospective study of the Danish general population initiated in 2003 and still recruiting. Individuals were selected randomly based on the national Danish Civil Registration System to reflect the adult Danish population aged 20-100. Data were obtained from a self-administered questionnaire reviewed together with an investigator at the day of attendance, a physical examination, and from blood samples including DNA extraction.

***Finland.*** *ADGEN.* The ADGEN cohort[71] is a clinic-based collection of AD patients from Eastern and Northern Finland examined in the Department of Neurology in Kuopio University Hospital and the Department of Neurology in Oulu University Hospital. All the patients were diagnosed with probable AD according to the criteria of the National Institute of Neurological and Communicative Disorders and Stroke and the Alzheimer's disease and Related Disorders Association (NINCDS–ADRDA). The study was approved by the ethics committee of Kuopio University Hospital, Finland (420/2016).

*The FINGER Study.* The FINGER Study[72] is a Finnish multi-domain lifestyle RCT enrolling 1,260 older adults with an increased risk of dementia from the general population. The intensive lifestyle intervention lasted for two years, and follow-up extends currently up to seven years. The FINGER study was approved by the coordinating ethics committee of the Hospital District of Helsinki and Uusimaa (94/13/03/00/2009 and HUS/1204/2017), and all the participants gave written informed consent.

***France.*** *The BALTAZAR multicenter prospective study.* BALTAZAR[73] includes 1,040 participants collected at 23 memory centers from September 2010 to April 2015. They were classified as AD cases (n = 501) according to DSM IV-TR and NINCDS–ADRDA criteria as well as amnestic mild cognitive impairment (MCI) cases (a MCI, n = 417) and non-amnestic MCI cases (na MCI, n = 122) according to Petersen's criteria. A comprehensive battery of cognitive tests was performed, including MMSE, verbal fluency, and FCSRT. All the participants or their legal guardians gave written informed consent. The study was approved by the Paris ethics committee (CPP Ile de France IV Saint Louis Hospital).

*MEMENTO*. MEMENTO is a clinic-based study[74] aimed at better understanding the natural history of AD, dementia, and related diseases. Between 2011 and 2014, 2,323 individuals presenting either recently diagnosed MCI or isolated cognitive complaints were enrolled in 26 memory centers in France. This study was performed in accordance with the guidelines of the Declaration of Helsinki. The MEMENTO study protocol has been approved by the local ethics committee (Comité de Protection des Personnes Sud-Ouest et Outre Mer III; approval number 2010-A01394-35). All the participants provided written informed consent.

*The CNRMAJ-Rouen Study*[75]. CNRMAJ-Rouen collects early-onset AD (EOAD) patients (n = 870). The patients or their legal guardians provided written informed consent. This study was approved by the ethics committee of CPP Ile de France II.

***Italy.*** The AD cases and controls were collated through Italy in different centers: Brescia, Cagliari, Florence, Milan, Rome, Pertugia, San Giovani Rotondo and Torino. AD cases were diagnosed according to DSM III-R,IV and NINCDS–ADRDA criteria. Controls were defined a minima as subjects without DMS-III-R dementia criteria and with integrity of their cognitive functions (MMS>25).

***Spain.*** *The Dementia Genetic Spanish Consortium (DEGESCO).* DEGESCO is a national consortium comprising 23 research centers and hospitals across the country, that holds the institutional coverage of The Network Center for Biomedical Research in Neurodegenerative Diseases (CIBERNED). Created in 2013, DEGESCO's objective is the promotion and conduction of genetic studies aimed at understanding the genetic architecture of neurodegenerative dementias in the Spanish population and participate in coordinated actions in national and international frameworks. All DNA samples are in compliance with the Law of Biomedical Research (Law 14/2007) and the Royal Decree on Biobanks (RD 1716/2011). Patients included in the present study met clinical criteria for probable or possible disease established by the NINCDS-ADRDA[11]. Cognitively healthy controls were unrelated individuals who had a documented MMSE in the normal range. Contributing centers in the France node genotyping were Centro de Biología Molecular Severo Ochoa (CSIC-UAM (Madrid), the Institute Biodonostia, University of Basque Contry (EHU-UPV, San Sebastián), Institut de Biomedicina de Valencia CSIC (València), and Sant Pau Biomedical Research Institute (Barcelona).

***Sweden.*** *Upsala.* The Swedish AD patients were ascertained at the Memory Disorder Unit at Uppsala University Hospital. For all patients, the diagnosis was established according to the NINCDS-ADRDA guidelines[11]. Healthy control subjects were recruited from the same geographic region following advertisements in local newspapers and displayed no signs of dementia upon Mini Mental State Examination (MMSE).

*Swedish National Study on Aging and Care in Kungsholmen (SNAC-K)*. The original SNAC-K population consisted of 4,590 living and eligible persons who lived on the island of Kungsholmen in Central Stockholm, belonged to pre-specified age strata, and were randomly selected to take part in the study. Between 2001 and 2004, 3363 persons participated in the baseline assessment. They belonged to the age cohorts 60, 66, 72, 78, 81, 84, 87, 90, 93, and 96 years and 99 years and older. The examination consists of three parts: a nurse interview, a medical examination, and a neuropsychological testing session. Altogether, the examination takes about six hours. The participants are reexamined each time they reach the next age cohort. All parts of the SNAC-K project have been approved by the ethical committee at Karolinska Institutet or the regional ethical review board. Informed consent was collected from all the participants or, if the person was severely cognitively impaired, from their next of kin.

***UK.*** *The UK Medical Research Council (MRC) cohort*. This sample set comprises individuals with Alzheimer's disease (AD) and healthy controls recruited across the MRC Centre for Neuropsychiatric Genetics and Genomics, Cardiff University, Cardiff, UK; Institute of Psychiatry, London, UK; University of Cambridge, Cambridge, UK. The collection of the samples was through multiple channels, including specialist NHS services and clinics, research registers and Join Dementia Research (JDR) platform. The participants were assessed at home or in research clinics along with an informant, usually a spouse, family member or close friend, who provided information about and on behalf of the individual with dementia. Established measures were used to ascertain the disease severity: Bristol activities of daily living (BADL), Clinical Dementia Rating scale (CDR), Neuropsychiatric Inventory (NPI) and Global Deterioration Scale (GlDS). Individuals with dementia completed the Addenbrooke's Cognitive Examination (ACE-r), Geriatric Depression Scale (GeDS), and the National Adult Reading Test (NART) too. Control participants were recruited from GP surgeries and by means of self-referral (including existing studies and Joint Dementia Research platform). For all other recruitment, all AD cases met criteria for either probable (NINCDS-ADRDA, DSM-IV) or definite (CERAD) AD. All elderly controls were screened for dementia using the Mini Mental State Examination (MMSE) or ADAS-cog, were determined to be free from dementia at neuropathological examination or had a Braak score of 2.5 or lower. Control samples were chosen to match case samples for age, gender, ethnicity and country of origin. Informed consent was obtained for all study participants, and the relevant independent ethical committees approved study protocols.

*SOTON*. All AD cases collected at the University of Southampton, Southampton, UK met criteria for either probable (NINCDS-ADRDA, DSM-IV) or definite (CERAD) AD. All elderly controls were screened for dementia using the MMSE or ADAS-cog, were determined to be free from dementia at neuropathological examination or had a Braak score of 2.5 or lower.

*Nottingham and Manchester*, University of Nottingham, Nottingham, UK and Manchester Brain Bank. All AD cases met criteria for either probable (NINCDS-ADRDA, DSM-IV) or definite (CERAD) AD. All elderly controls were screened for dementia using the MMSE or ADAS-cog, were determined to be free from dementia at neuropathological examination or had a Braak score of 2.5 or lower.

*King's College London (KCL).* For the KCL London Neurodegenerative Diseases Brain Bank, all AD cases met criteria for either probable (NINCDS-ADRDA, DSM-IV) or definite (CERAD) AD. All elderly controls were screened for dementia using the MMSE or ADAS-cog, were determined to be free from dementia at neuropathological examination or had a Braak score of 2.5 or lower.

*PRION.* All AD cases met criteria for either probable (NINCDS-ADRDA, DSM-IV) or definite (CERAD) AD. All elderly controls were screened for dementia using the MMSE or ADAS-cog, were determined to be free from dementia at neuropathological examination or had a Braak score of 2.5 or lower.

*The Cognitive Function and Ageing Study Wales (CFAS-Wales).* CFAS-Wales is a longitudinal population-based study of people aged 65 years and over in rural and urban areas of Wales that aims to investigate physical and cognitive health in older age and examine the interactions between health, social networks, activity, and participation. Individuals aged 65 years and over were randomly sampled from general medical practice lists between 2011 and 2013, stratified by age to ensure equal numbers in two age groups, 65-74 years and 75 and over. The baseline sample included 3593 older people and included those living in care homes as well as those living at home. Those who provided written consent to join the study were interviewed in their own homes by trained interviewers and could choose to have the interview conducted through the medium of either English or Welsh. Participants were followed up 2 years later. All AD cases met criteria for either probable (NINCDS-ADRDA, DSM-IV) or definite (CERAD) AD. All elderly controls were screened for dementia using the MMSE or ADAS-cog, were determined to be free from dementia at neuropathological examination or had a Braak score of 2.5 or lower.

*The UCL EOAD cohort of the Dementia Research Centre (UCL-DRC).* The UCL-DRC cohort included patients seen at the Cognitive Disorders Clinics at The National Hospital for Neurology and Neurosurgery (Queen Square), or affiliated hospitals. Individuals were assessed clinically and diagnosed as having probable Alzheimer's disease based on contemporary clinical criteria in use at the time, including imaging and neuropsychological testing where appropriate.

***EADB-Germany.*** In the German node, samples were collected from seven countries (11 centers/studies) and after QCs, we obtained 4,159 AD cases and 4,545 controls. All these samples were genotyped at Life&brain (Bonn, Germany).

***Germany.*** *The VOGEL study*. The VOGEL study is a prospective, observational, long-term follow-up study with three time points of investigation within 6–8 years. This cohort includes dementia and healthy subjects. Residents of the city of Würzburg born between 1936 and 1941 were recruited. Every participant underwent physical, psychiatric, and laboratory examinations and performed intense neuropsychological testing as well as VSEP and NIRS according to the published procedures. A total of 604 subjects were included.

*The Heidelberg/Mannheim memory clinic sample.* This cohort includes 61 subjects from whom 40 MCI patients were recruited and assessed between 2012 and 2016. Some of those patients converted to dementia by AD or other dementias.

*The PAGES study.* This study includes 301 subjects. AD patients were recruited at the memory clinic of the Department of Psychiatry, University of Munich, Germany. Participants in whom dementia associated with AD was diagnosed fulfilled the criteria for probable AD according to the NINCDS–ADRDA. The control group included participants who were randomly selected from the general population of Munich. Controls who had central nervous system diseases or psychotic disorders or who had first-degree relatives with psychotic disorders were excluded.

*The Technische Universität München study.* This cohort includes 359 healthy, AD, and other dementias patients recruited from the Centre for Cognitive Disorders. All the participants provided written informed consent. A biobank was submitted to the ethics committee of the Technical University of Munich, School of Medicine (Munich, Germany), which raised no objections and approved the biobank (reference number 347-14).

*The Göttingen Universität study.* This study includes 111 in- and outpatients with a healthy or AD dementia status from the Department of Psychiatry of the University of Göttingen. The study's ethical statement was provided locally at the Göttingen University Medical Centre.

*The German Dementia Competence Network (DCN) cohort.* Individuals from the DCN cohort were recruited from 14 university hospital memory clinics across Germany between 2003 and 2005[76]. The study was approved by the respective ethics committees, and written informed consent was obtained from all the participants prior to inclusion.

*The German Study on Aging, Cognition, and Dementia (AgeCoDe).* The AgeCoDe study is a general practice (GP) registry-based longitudinal study in elderly individuals that recruited patients aged 75 years and above in six German cities from 2003 to 2004[77]. The study was approved by the respective ethics committees, and written informed consent was obtained from all the participants prior to inclusion.

***Greece.*** *The HELIAD study*, comprising 49 AD cases and 1,150 controls. HELIAD is a population-based, multidisciplinary, collaborative study designed to estimate, in the Greek population over the age of 64 years, the prevalence and incidence of MCI, AD, other forms of dementia, and other neuropsychiatric conditions of aging and to investigate associations between nutrition and cognitive dysfunction or age-related neuropsychiatric diseases. The participants were selected through random sampling from the records of two Greek municipalities, Larissa and Marousi. All the participants signed informed consent in Greek.

***Portugal***. *The Lisbon study.* This Portuguese cohort totaling 78 AD cases and 74 controls was recruited in 2008–2009 to investigate the connections between oxidative stress and lipid dyshomeostasis in AD. The project includes 190 subjects and was approved by the local ethics committee, and all the participants provided written informed consent. This study includes healthy and dementia-by-AD subjects.

***Spain.*** *DEGESCO.* DEGESCO Centers from whom DNA samples were genotyped in the German node (1,778 cases and 470 controls) were the Alzheimer Research Center and Memory Clinic, Fundació ACE, Institut Català de Neurociències Aplicades (Barcelona), the Neurology Service at University Hospital Marqués de Valdecilla (Santander), the Alzheimer's disease and other cognitive disorders, Neurology Department, at Hospital Clínic, IDIBAPS (Barcelona), the Molecular Genetics Laboratory, at the Hospital Universitario Central de Asturias (Oviedo), and Fundació Docència i Recerca Mútua de Terrassa and Movement Disorders Unit, Department of Neurology, University Hospital Mútua de Terrassa (Barcelona).

***Switzerland.*** Two datasets from Switzerland and Austria were combined, totaling 182 AD cases and 388 controls.

*The Lausanne study*. This study includes 137 community-dwelling participants aged 55+ years with cognitive impairment (memory clinic patients with MCI, dementia) or normal cognition (recruited by advertisement, word of mouth). The study's ethical statement was provided locally at the Department of Psychiatry, Geneva University Centre, Switzerland.

*The VITA study*. This is a longitudinal study of 606 individuals (Vienna, Austria) who were 75 years old in 2000, followed up every 30–90 months. This cohort includes dementia and healthy subjects. All the participants gave written informed consent. The study conformed to the latest version of the Declaration of Helsinki and was approved by the ethics committee of the City of Vienna, Austria

***EADB-Netherlands.*** In the Dutch node, samples were collected from six organizations in the Netherlands and after QCs, we obtained 2,438 AD cases and 2,389 controls. All these samples were genotyped at the Erasmus Medical University (Rotterdam, The Netherlands). The Medical Ethics Committee (METC) of the local institutes approved the studies. All the participants and/or their legal guardians gave written informed consent for participation in the clinical and genetic studies. Samples from the following institutes were included.

*Erasmus Medical Center.* Most individuals were selected from population studies from the epidemiology department and accounted for most of the control, while a smaller subset of samples originated from the neurology department, where AD was diagnosed according to the NINCDS-ADRDA/NIA-AA criteria for AD[11,12].

*The Amsterdam Dementia Cohort (ADC)*. This cohort[78] comprises patients who visit the memory clinic of the VU University Medical Centre, the Netherlands. The diagnosis of probable

AD is based on the clinical criteria formulated by the NINCDS–ADRDA and based on the NIA–AA. Diagnosis of MCI was made according to Petersen et al.[79] and NIA-AA criteria. Controls presented with subjective cognitive decline at the memory clinic but performed within normal limits on all clinical investigations.

*The 100-Plus study.* This study includes Dutch-speaking individuals who (i) can provide official evidence for being aged 100 years or older, (ii) self-report to be cognitively healthy, which is confirmed by a proxy, (iii) consent to the donation of a blood sample, (iv) consent to (at least) two home visits from a researcher, and (v) consent to undergo an interview and neuropsychological test battery[80].

*Parelsnoer Institute.* A collaboration between eight Dutch University Medical Centers in which clinical data and biomaterials from patients suffering from chronic diseases (so called "Pearls") are collected according to harmonized protocols. The Pearl Neurodegenerative Diseases study[81] includes individuals diagnosed with dementia, mild cognitive impairment, and controls with subjective memory complaints.

*Netherlands Brain Bank:* a non-profit organization that collects human brain tissue of donors with a variety of neurological and psychiatric disorders, but also of non-diseased donors. A clinical diagnosis of AD is based on the clinical criteria of probable AD[11,82]. The selected AD patients for this study all received a definitive diagnosis which was based on autopsy.

*Maastricht University Medical Center*: a subset of individuals that were referred to the memory clinic for cognitive complaints were included if they participated in the BioBank-Alzheimer Centrum Limburg (BB-ACL)[83]. For these patients, blood and cerebrospinal fluid is withdrawn and collected in a biobank for scientific research. Diagnosis of MCI was made according to the criteria of Peterson, and diagnosis of AD-type dementia was made according to the criteria of the DSM-IV[84] and the NINCDS-ADRDA[11].

***EADB-Australia.*** *The Sydney MAS study.* MAS is a longitudinal study investigating MCI, related syndromes, and age-related cognitive change. Older adults (70–90 years old) were randomly recruited from the community in Sydney, Australia (n = 1,037). An extensive interview was undertaken and questionnaire data collected, including demographics, cognitive performance, and medical history. The majority of participants provided blood samples for genetic analysis. Neuroimaging was performed on a subset of participants. Ethics approval for the study was provided by the ethics committee of the University of New South Wales and the Illawarra Area Health Service Human Research Ethics Committee. All the participants provided written informed consent to join the study. More information is provided elsewhere[85]. In our study, there were 43 AD cases and 215 controls. Due to the low sample size, the study was not considered in the meta-analysis.

**UK Biobank**

*Dataset Description.* The AD cases (subjects with one or both parents with a history of AD) were 66.1% female and had an average age of 81.9 (± 5.49 SD). The AD proxy controls (no history of AD in either parent) were 53.1% female and had an average age of 78.4 (± 6.0 SD). All subjects selected for analyses were of British ancestry to facilitate comparison to IGAP and EADB. Additional descriptive characteristics are included in **Supplementary Table 1**. The UK Biobank genotypes were imputed using both the HRC reference panel r1.1 and a merged UK10K and 1000 Genomes phase 3 reference panels to produce 93,095,623 autosomal SNPs, short indels, and large structural variants in 487,442 individuals. A detailed description of the chip arrays and methods is available elsewhere[86].

***Methods for EADB and UKBB Replication Datasets***

**EADB**

A detailed description of the EADB genotyping, QC, imputation and association analyses is available in Bellenguez et al.[68], and a summary is provided below.

*Genotyping.* Samples were genotyped with the Illumina Infinium Global Screening Array (GSA, GSAsharedCUSTOM_24+v1.0) in 3 genotyping centers and called using the same cluster file.

*Chip assessment.* Positions and alleles of variants were assessed using the Illumina support files. Variants which are part of the removed markers list provided by Illumina were excluded. Finally, only variants for which the full-length probe(s) aligned uniquely on the GRCh38 genome without any mismatch were kept.

*Variant Intensity Quality Control (QC).* A QC on the intensity metrics were performed on all autosomal variants. The metrics and their thresholds were adapted from the HumanExome BeadChip quality control paper[87].

*Sample QC.* Samples showing missingness >0.05 on autosomal variants or having a heterozygosity metric (Method-of-moments *F* coefficient estimate from PLINK v1.9[88-90] outside the interval mean ±6 standard deviation (SD) were excluded. Samples showing a discordant genetic and clinical sex using the sex-check function of PLINK v1.9 with the default parameters were also removed. Population outliers were identified using the 1000 Genome Phase3 data (1000GP3). Principal components (PCs) were computed on 1000GP3 samples and the EADB samples were then projected onto these PCs using the flashPCA2 software[91]. EADB samples outside the interval median ±12 median absolute deviation (mad), computed using only the EADB samples, on PC1 or on PC2 were flagged as population outliers. The relatedness between samples was estimated using the GENESIS package[92] to take into account the EADB population structure. The variants used were the same than the ones included in the population outlier analysis and 21 PCs were used in the PC-Relate step. All pairs with a kinship >0.09375 (representing the mean between 2$^{nd}$ and 3$^{rd}$ degrees) were processed to select the unrelated samples, preferring cases over controls or controls over missing status samples. Moreover, duplicates with sex or status mismatch were discarded.

*Variant QC.* Autosomal variants showing a missingness >0.05 in at least one genotyping center or having a differential missingness test $P<10^{-10}$ were excluded. The Hardy-Weinberg equilibrium test was performed only in controls, excluding flagged population outliers, and for each genotyping center/country separately. A variant was excluded if at least one center/country test showed $P<5\times10^{-8}$. The remaining variants were then assessed by comparing their frequencies against two reference panels (i.e. the Haplotype Reference Consortium r1.1 (HRC)[93] and the Genome Aggregation Database v3 (gnomAD)[94]). For HRC, the allele counts excluding 1000 Genome samples were used, and for gnomAD, Finnish and non-Finnish allele counts were used. Allele counts were then compared to EADB allele counts by performing a chi-square test statistics ($\chi^2$). After graphical review, variants showing a $\chi^2$ >3,000 in both HRC and gnomAD or a $\chi^2$>3,000 in one reference panel and not present in the other were excluded. Finally, genome-wide association studies (GWAS) were performed between controls across genotyping centers, excluding population outliers and related samples, to assess frequency differences between genotyping centers. The analyses were performed for each genotyping centers pairs in parallel (3 analyses), converting the center into a binary variable and used as the analysis phenotype. The analyses were performed using SNPTEST[95] using an additive model with the new model-fitting functionality ('newml'), adjusting on the associated PCs. All variants having a Likelihood Ratio Test of $P<10^{-5}$ were excluded. Finally, all ambiguous variants having a MAF>0.4 were excluded and only one copy of any duplicated variants of the chip were included, considering the one with the lowest missingness.

*Imputations.* All samples and variants passing the QC were used as the input of the imputation process. The imputation was performed by the Michigan Imputation Server[96] where the TOPMed Freeze 5 reference panel was granted to the EADB consortium. The server version used was the 1.2.4 with EAGLE v2.4[97] as the phasing software and Minimac4 v4-1.0.2 as the imputation software. Due to the limitation in terms of maximum number of samples per job (20,000), the samples were split into 5 batches for imputation. After the imputation process of all batches, in order to have a global imputation quality, a merged imputation quality was recomputed including all samples using the *bcftools impute_info* plugin.

*Association Analyses.* 20,301 AD cases and 21,839 controls with TopMed Freeze 5 imputed data were finally available for analysis. Association tests between AD status and each genetic variant of autosomal chromosomes were conducted using an additive genetic model with a logistic regression as implemented in SNPTEST[95]. Analyses were performed on the genotype probabilities with the 'newml' method and were adjusted on 20 principal components and genotyping centers. We filtered (i) duplicated variants, (ii) variants with missing effect size, standard error or *P*-value, (iii) variants with a mac-info score below 20 (the 'mac-info' score is defined as the product of the minor allele count (MAC) and the imputation quality). Prior to meta-analysis, additional filtering was done for two other criteria, (iv) variants with absolute value of effect size above 5 and iv) variants with imputation quality of $R^2$<0.3.

**UK Biobank**

*AD Parental History GWAS.* Association analysis of parental history of AD were performed in the UKBB by identifying subjects with available genotyping and non-missing information on parental history of AD/dementia. Parental history was ascertained by particpants' responses to two questions, "Has/did your father ever suffer from Alzheimer's disease/dementia?" and "Has/did your mother ever suffer from Alzheimer'sdisease/dementia?" Individuals reporting at least one parent with a history of AD/dementia were classified as cases while those reporting no parents with AD/dementia were classified as controls. Single variant-based case-control association testing was performed similarly to IGAP and EADB association analyses, in SNPTEST[95] using score-based logistic regression under an additive model with covariate adjustment for the first three PCs only from EIGENSTRAT. Standard QC was implemented prior to imputation and analysis, and following analysis, variants were additional filtered for extreme departure from HWE ($P<10^{-6}$) among controls and $R^2$<0.8. Overall GWAS results are presented for common variants (MAF>0.01) in **Supplementary Table 4** and **Supplementary Figure 3a** and rare variants (MAF≤0.01) in **Supplementary Table 7** and **Supplementary Figure 3b**. Modest inflation of genome-wide association was observed among common variants (**Supplementary Figure 3a**) and rare variants (**Supplementary Figure 3b**).

**Supplementary Table 3.** Strongest common variant (MAF≥0.01) association results by genomic region in UKBB. Variants depicted had $P<10^{-5}$ in UKBB AD-proxy analysis (n=1,472 variants). Also shown are associations for those variants in IGAP and findings from meta-analysis of UKBB and IGAP for those variants.

| Variant RSID | Chr | Basepair (hg19) | Genomic Region | EFF | REF | UKBB | | | | IGAP | | | UKBB + IGAP Meta-Analysis | | | |
|---|---|---|---|---|---|---|---|---|---|---|---|---|---|---|---|---|
| | | | | | | EAF | OR (95% CI) | *P* | $R^2$ | EAF | OR (95% CI) | *P* | EAF | OR (95% CI) | *P* | $P_{HET}$ |
| rs12972156 | 19 | 45387459 | *APOE* Region | G | C | 0.1514 | 1.51 (1.48, 1.54) | <1E-320 | 0.99 | 0.1566 | 2.42 (2.35, 2.50) | 3.36E-625 | 0.1445 | 1.74 (1.71, 1.77) | 5.32E-817 | 1.04E-127 |
| rs6733839 | 2 | 127892810 | *BIN1* | T | C | 0.3921 | 1.07 (1.05, 1.09) | 1.97E-16 | 0.95 | 0.3836 | 1.18 (1.15, 1.21) | 2.36E-35 | 0.3876 | 1.10 (1.09, 1.12) | 2.10E-42 | 1.59E-9 |
| rs7384878 | 7 | 99932049 | *PILRB* | T | C | 0.3191 | 0.95 (0.93, 0.97) | 1.43E-8 | 1.01 | 0.3117 | 0.93 (0.90, 0.96) | 2.00E-7 | 0.3182 | 0.94 (0.93, 0.96) | 3.50E-14 | 0.178 |
| rs35653258 | 6 | 32568783 | *HLA-DRB1* | A | C | 0.1879 | 0.94 (0.92, 0.96) | 1.78E-8 | 0.96 | 0.1560* | 0.92 (0.88, 0.95)* | 4.11E-5* | 0.1865* | 0.94 (0.92, 0.96)* | 3.05E-11* | 0.193* |
| rs375452507 | 16 | 31115259 | *VKORC1/ BCKDK* | A | T | 0.3758 | 1.05 (1.03, 1.07) | 3.82E-8 | 1.01 | 0.3890* | 1.03 (1.00, 1.05)* | 0.0438* | 0.3704* | 1.04 (1.02, 1.05)* | 9.19E-8* | 0.268 |
| rs7759000 | 6 | 22291959 | *PRL/CASC15* | C | T | 0.2467 | 1.05 (1.03, 1.07) | 5.99E-8 | 0.99 | 0.2513 | 1.01 (0.98, 1.04) | 0.3595 | 0.2470 | 1.04 (1.02, 1.06) | 4.793E-07 | 0.0277 |
| rs7341557 | 8 | 27402777 | *EPHX2* | A | G | 0.1120 | 0.93 (0.91, 0.96) | 1.10E-7 | 0.99 | 0.0984 | 0.92 (0.88, 0.96) | 1.69E-4 | 0.1091 | 0.93 (0.91, 0.95) | 8.13E-11 | 0.665 |
| rs12642946 | 4 | 16266117 | *TAPT1-AS1* | G | A | 0.0569 | 1.10 (1.06, 1.14) | 2.47E-7 | 0.9 | 0.3890* | 1.03 (1.00, 1.05)* | 0.0438* | 0.3704* | 1.04 (1.02, 1.05)* | 9.19E-8* | 0.268* |
| rs9282562 | 19 | 1051137 | *ABCA7* | C | CTG | 0.1111 | 1.07 (1.04, 1.10) | 2.89E-7 | 0.99 | 0.1151* | 1.07 (1.03, 1.12)* | 9.04E-4* | 0.1141* | 1.06 (1.04, 1.09)* | 3.31E-8* | 0.575 |
| rs139920711 | 8 | 139353780 | *FAM135B* | GTA CTT | G | 0.3253 | 1.05 (1.03, 1.06) | 4.82E-7 | 0.98 | 0.3518* | 0.99 (0.96, 1.01)* | 0.293* | 0.3358* | 1.03 (1.01, 1.04)* | 4.13E-4* | 3.23E-4* |
| rs1332263 | 10 | 110247810 | *LINC02661* | A | C | 0.3929 | 0.96 (0.94, 0.97) | 5.12E-7 | 1 | 0.4066 | 1.00 (0.97, 1.02) | 0.818 | 0.3982 | 0.97 (0.96, 0.98) | 1.54E-5 | 0.0103 |
| rs527162 | 11 | 85715736 | *PICALM* | T | C | 0.1959 | 0.95 (0.93, 0.97) | 8.12E-7 | 1 | 0.2146 | 0.91 (0.88, 0.94) | 1.45E-9 | 0.2025 | 0.94 (0.92, 0.95) | 9.00E-14 | 0.0215 |
| rs373377575 | 11 | 59999757 | *MS4A* Region | A | AAC | 0.3792 | 0.96 (0.94, 0.97) | 9.29E-7 | 0.94 | 0.3830* | 0.89 (0.87, 0.92)* | 3.63E-18* | 0.3824* | 0.94 (0.93, 0.95)* | 3.10E-18* | 2.54E-6* |
| rs593742 | 15 | 59045774 | *ADAM10/ MINDY2* | G | A | 0.3071 | 0.96 (0.94, 0.97) | 1.51E-6 | 0.99 | 0.2999 | 0.95 (0.92, 0.98) | 1.84E-4 | 0.3059 | 0.96 (0.94, 0.97) | 1.27E-9 | 0.617 |
| rs17269688 | 2 | 106469267 | *NCK2* | G | A | 0.0225 | 0.87 (0.82, 0.92) | 1.52E-6 | 0.99 | 0.0185 | 0.92 (0.84, 1.01) | 0.0771 | 0.0217 | 0.88 (0.84, 0.93) | 5.02E-7 | 0.319 |
| rs7810606 | 7 | 143108158 | *EPHA1* | C | T | 0.4825 | 0.96 (0.95, 0.98) | 1.90E-6 | 1.26 | 0.4838 | 0.96 (0.93, 0.98) | 8.78E-4 | 0.4840 | 0.96 (0.95, 0.97) | 6.51E-9 | 0.844 |
| rs35413307 | 13 | 112191778 | *TEX29/SOX1* | G | T | 0.4036 | 1.04 (1.02, 1.06) | 1.94E-6 | 1.01 | 0.4146 | 1.01 (0.98, 1.03) | 0.619 | 0.4058 | 1.03 (1.02, 1.04) | 2.03E-5 | 0.0296 |
| rs6656401 | 1 | 207692049 | *CR1* | G | A | 0.1774 | 1.05 (1.03, 1.07) | 2.34E-6 | 1 | 0.1975 | 1.14 (1.11, 1.18) | 4.18E-17 | 0.1827 | 1.08 (1.06, 1.10) | 9.32E-18 | 1.08E-5 |
| rs79007298 | 8 | 146062391 | *ZNF7* | T | C | 0.0161 | 0.85 (0.79, 0.91) | 2.85E-6 | 1 | 0.0158 | 0.95 (0.84, 1.08) | 0.438 | 0.0163 | 0.87 (0.82, 0.93) | 7.44E-6 | 0.118 |
| rs6125602 | 20 | 36240070 | *LINC01746/ LINC00489* | A | G | 0.2374 | 1.05 (1.03, 1.07) | 3.23E-6 | 0.99 | 0.2530 | 1.00 (0.97, 1.03) | 0.945 | 0.2411 | 1.03 (1.02, 1.05) | 7.81E-5 | 0.0137 |
| rs62378972 | 5 | 172734106 | *STC2* | G | C | 0.1321 | 1.06 (1.04, 1.09) | 3.97E-6 | 0.89 | -- | -- | -- | 0.1310 | 1.06 (1.04, 1.09) | 3.97E-6 | 1 |
| rs61871136 | 10 | 104349697 | *SUFU* | A | G | 0.0120 | 1.19 (1.10, 1.27) | 4.09E-6 | 0.97 | 0.0145 | 1.06 (0.94, 1.18) | 0.337 | 0.0124 | 1.15 (1.08, 1.22) | 1.11E-5 | 0.0928 |
| rs115553581 | 4 | 152935279 | *AC097375.5* | T | C | 0.0200 | 1.14 (1.08, 1.21) | 4.31E-6 | 0.99 | 0.0214 | 0.94 (0.85, 1.03) | 0.160 | 0.0200 | 1.08 (1.03, 1.13) | 0.00146 | 3.18E-4 |
| rs7136907 | 12 | 129655318 | *TMEM132D* | A | C | 0.3168 | 0.96 (0.94, 0.98) | 4.64E-6 | 1.01 | 0.3355 | 1.00 (0.97, 1.03) | 0.979 | 0.3234 | 0.97 (0.96, 0.99) | 1.32E-4 | 0.0116 |
| rs75479668 | 3 | 54288332 | *CACNA2D3* | T | C | 0.1768 | 0.95 (0.93, 0.97) | 6.71E-6 | 0.99 | 0.1883 | 1.03 (1.00, 1.06) | 0.0807 | 0.1812 | 0.98 (0.96, 0.99) | 0.00621 | 6.84E-5 |
| rs11711675 | 3 | 193411197 | *OPA1* | T | G | 0.4834 | 0.96 (0.95, 0.98) | 6.89E-6 | 1.01 | 0.4819 | 1.01 (0.98, 1.03) | 0.531 | 0.4841 | 0.98 (0.96, 0.99) | 6.30E-4 | 0.00280 |
| rs4575098 | 1 | 161155392 | *ADAMTS4* | A | G | 0.2358 | 1.04 (1.02, 1.06) | 9.04E-6 | 1 | 0.2373 | 1.04 (1.01, 1.07) | 0.00792 | 0.2353 | 1.04 (1.03, 1.06) | 2.31E-7 | 0.844 |

Abbreviations: Chr, chromosome; EFF, Effect allele; REF, reference allele; EAF, effect allele frequency; OR, Odds ratio; CI, Confidence interval; *P*, P-value

Shaded box indicates genome-wide significance within analysis stage

* Proxy variants used in IGAP where index variant not available:

rs35653258: used rs34039593 ($D'$=1, $r^2$=0.9262), with minor allele G (MAF=0.1924) and UKBB association of $P=7.33\times10^{-8}$ [OR (95% CI): 0.94 (0.92, 0.96)]
rs375452507: used rs56314408 ($D'$=1, $r^2$=0.9713), with minor allele G (MAF=0.3639) and UKBB association of $P=3.92\times10^{-7}$ [OR (95% CI): 1.04 (1.02, 1.06)]
rs12642946: used rs56314408 ($D'$=1, $r^2$=0.0291), with minor allele G (MAF=0.1205) and UKBB association of $P=9.26\times10^{-7}$ [OR (95% CI): 1.06 (1.04, 1.09)]
rs9282562: used rs72973584 ($D'$=0.9512, $r^2$=0.8961), with minor allele T (MAF=0.1147) and UKBB association of $P=8.63\times10^{-6}$ [OR (95% CI): 1.06 (1.03, 1.09)]
rs139920711: used rs4410912 ($D'$=1, $r^2$= 0.9957), with minor allele T (MAF=0.3304) and UKBB association of $P=8.27\times10^{-7}$ [OR (95% CI): 1.04 (1.03, 1.06)]
rs373377575: used rs1582763 ($D'$=0.8790, $r^2$=0.6920), with minor allele A (MAF=0.3805) and UKBB association of $P=2.25\times10^{-6}$ [OR (95% CI): 0.96 (0.94, 0.98)]
rs62378972: No LD-based proxy with complete IGAP association results was available

**Supplementary Table 6.** Strongest rare variant (MAF<0.01) association results by genomic region in UKBB. Variants depicted had $P<10^{-5}$ in UKBB AD-proxy analysis (n=153 variants). Also shown are associations for those variants in IGAP and findings from meta-analysis of UKBB and IGAP for those variants.

| Variant RSID | Chr | Basepair (hg19) | Genomic Region | EFF | REF | UKBB | | | | IGAP | | | UKBB + IGAP Meta-Analysis | | | |
|---|---|---|---|---|---|---|---|---|---|---|---|---|---|---|---|---|
| | | | | | | EAF | OR (95% CI) | *P* | $R^2$ | EAF | OR (95% CI) | *P* | EAF | OR (95% CI) | *P* | $P_{HET}$ |
| rs138607350 | 19 | 45363820 | *APOE* Region | G | T | 0.0091 | 1.56 (1.44, 1.69) | 2.02E-27 | 0.95 | 0.0113 | 2.80 (2.45, 3.19) | 5.72E-52 | 0.0104 | 2.65 (2.39, 2.93) | 2.90E-77 | 0.205 |
| rs149416930 | 16 | 14222206 | *MRTFB* | T | C | 0.0015 | 1.75 (1.44, 2.13) | 1.94E-8 | 0.94 | 0.0017 | 1.04 (0.72, 1.51) | 0.839 | 0.0015 | 1.56 (1.32, 1.86) | 3.92E-7 | 0.0155 |
| rs148024771 | 12 | 25995521 | *AC019209.2/ MIR4302/RASSF8* | T | G | 0.0019 | 1.61 (1.36, 1.90) | 3.89E-8 | 0.96 | 0.0018 | 0.89 (0.64, 1.23) | 0.479 | 0.0019 | 1.42 (1.22, 1.65) | 4.85E-6 | 0.00174 |
| rs187370608 | 6 | 40942196 | *TREM2* | A | G | 0.0031 | 1.45 (1.26, 1.67) | 1.65E-7 | 0.96 | 0.0042 | 2.19 (1.78, 2.70) | 2.33E-13 | 0.0034 | 1.65 (1.47, 1.85) | 3.82E-17 | 0.00138 |
| rs191971784 | 6 | 2194461 | *GMDS* | A | G | 0.0013 | 1.67 (1.36, 2.04) | 8.59E-7 | 0.99 | -- | -- | -- | 0.0013 | 1.67 (1.36, 2.04) | 8.59E-7 | 1 |
| rs554191882 | 11 | 113654268 | *CLDN25/ ZW10/USP28* | G | A | 0.0011 | 1.73 (1.39, 2.16) | 9.11E-7 | 0.93 | 0.0011 | 1.01 (0.64, 1.58) | 0.973 | 0.8065 | 0.64 (0.53, 0.78) | 1.10E-5 | 0.0288 |
| rs542529520 | 5 | 56961754 | *AC008780.2* | T | C | 0.0014 | 1.64 (1.34, 2.00) | 1.08E-6 | 0.84 | -- | -- | -- | 0.0014 | 1.64 (1.34, 2.00) | 1.08E-6 | 1 |
| rs540776099 | 5 | 139349958 | *NRG2* | C | T | 0.0033 | 1.41 (1.23, 1.62) | 1.22E-6 | 0.93 | 0.0044 | 1.13 (0.91, 1.40) | 0.269 | 0.7021 | 0.81 (0.72, 0.91) | 5.28E-4 | 3.57E-4 |
| rs192467905 | 11 | 64019292 | *PLCB3* | G | C | 0.0013 | 1.62 (1.33, 1.98) | 1.67E-6 | 0.99 | 0.0051 | 1.01 (0.84, 1.21) | 0.927 | 0.4672 | 0.80 (0.70, 0.92) | 0.00138 | 3.63E-4 |
| rs117321942 | 7 | 136814593 | *AC009264.1* | G | A | 0.0071 | 0.77 (0.69, 0.85) | 1.73E-6 | 0.97 | 0.0073 | 1.01 (0.86, 1.19) | 0.884 | 0.6767 | 1.20 (1.10, 1.32) | 5.73E-5 | 0.00964 |
| rs75575390 | 8 | 142650468 | *AC025839.1* | A | G | 0.0081 | 1.23 (1.12, 1.34) | 4.08E-6 | 0.97 | 0.0102 | 1.10 (0.97, 1.25) | 0.137 | 0.0088 | 1.19 (1.10, 1.28) | 3.33E-6 | 0.176 |
| rs551543924 | 12 | 120639806 | *PXN-AS1* | C | T | 0.0073 | 1.25 (1.13, 1.37) | 4.40E-6 | 0.95 | 0.0081 | 1.11 (0.94, 1.31) | 0.218 | 0.7472 | 0.87 (0.80, 0.94) | 7.71E-4 | 7.80E-4 |
| rs575683378 | 4 | 90745093 | *SNCA* | T | C | 0.0014 | 1.59 (1.30, 1.94) | 4.85E-6 | 0.94 | -- | -- | -- | 0.0014 | 1.59 (1.30, 1.94) | 4.85E-6 | 1 |
| rs541044282 | 10 | 112359045 | *SMC3* | A | G | 0.0018 | 1.57 (1.29, 1.91) | 5.22E-6 | 0.90 | 0.0015 | 1.03 (0.69, 1.54) | 0.874 | 0.0018 | 1.45 (1.22, 1.73) | 3.19E-5 | 0.0621 |
| rs534010278 | 4 | 74175325 | *ALB/ANKRD17* | A | G | 0.0011 | 1.65 (1.32, 2.05) | 7.23E-6 | 0.97 | -- | -- | -- | 0.0011 | 1.65 (1.32, 2.05) | 7.23E-6 | 1 |

Abbreviations: Chr, chromosome; EFF, Effect allele; REF, reference allele; EAF, effect allele frequency; OR, Odds ratio; CI, Confidence interval; *P*, P-value

Shaded box indicates genome-wide significance within analysis stage

**Supplementary Table 8.** Rare variant (MAF<0.01) conditional association in ADGC for *APOE*, *TREM2*, and *NCK2* variants

**(a)** Conditional analyses of *APOE* region variants on *APOE* ε2 and ε4 haplotypes

| Variant RSID | Chrom. | Basepair (hg19) | Genomic Region | EFF | REF | ADGC Meta-Analysis | | | Adjustment for *APOE* ε2 dosage | | | Adjustment for *APOE* ε2 and ε4 dosages | | |
|---|---|---|---|---|---|---|---|---|---|---|---|---|---|---|
| | | | | | | EAF | OR (95% CI) | *P* | EAF | OR (95% CI) | *P* | EAF | OR (95% CI) | *P* |
| rs547509922 | 19 | 45331261 | *APOE* Region (intergenic) | T | C | 0.0050 | 2.57 (2.00, 3.30) | 1.88E-13 | 0.0048 | 2.80 (2.13, 3.67) | 1.22E-13 | 0.0050 | 1.36 (1.02, 1.82) | 0.037 |
| rs559118614 | 19 | 45554058 | *APOE* Region (*CLASRP*) | G | T | 0.0106 | 1.97 (1.65, 2.35) | 5.09E-14 | 0.0105 | 2.06 (1.71, 2.48) | 1.65E-14 | 0.0106 | 1.13 (0.92, 1.38) | 0.243 |
| rs112952132 | 19 | 45198060 | *APOE* Region (intergenic) | T | C | 0.0081 | 0.50 (0.41, 0.62) | 2.80E-11 | 0.0082 | 0.56 (0.46, 0.70) | 1.30E-7 | 0.0080 | 0.82 (0.66, 1.03) | 0.0937 |
| rs78905796 | 19 | 45132679 | *APOE* Region (*IGSF23*) | A | G | 0.0055 | 1.70 (1.34, 2.15) | 1.12E-5 | 0.0053 | 1.73 (1.34, 2.23) | 2.45E-5 | 0.0055 | 1.11 (0.85, 1.45) | 0.429 |

Abbreviations: Chr, chromosome; EFF, Effect allele; REF, reference allele; EAF, effect allele frequency; OR, Odds ratio; CI, Confidence interval; *P*, P-value

**(b)** Conditional analyses of *TREM2* variants on R47H and R62H*

| Variant RSID | Chrom. | Basepair (hg19) | Genomic Region | EFF | REF | ADGC Meta-Analysis | | | Adjustment for *TREM2* R47H and R62H | | |
|---|---|---|---|---|---|---|---|---|---|---|---|
| | | | | | | EAF | OR (95% CI) | *P* | EAF | OR (95% CI) | *P* |
| rs143202663 | 6 | 40865240 | *TREM2* (upstream) | C | T | 0.0049 | 1.90 (1.48, 2.44) | 4.67E-7 | 0.0049 | 1.3 (0.74, 2.28) | 0.368 |
| rs145342536 | 6 | 40911888 | *TREM2* (upstream) | A | T | 0.0056 | 1.80 (1.43, 2.27) | 5.883E-7 | 0.0056 | 1.26 (0.66, 2.41) | 0.487 |
| rs12664332 | 6 | 40904030 | *TREM2* (upstream) | A | G | 0.0056 | 1.79 (1.42, 2.25) | 8.55E-7 | 0.0055 | 1.21 (0.63, 2.33) | 0.564 |
| rs192675224 | 6 | 40706366 | *TREM2* (upstream) | A | G | 0.0041 | 1.73 (1.31, 2.28) | 1.19E-4 | 0.0041 | 1.06 (0.69, 1.62) | 0.785 |
| rs150085726 | 6 | 40729544 | *TREM2* (upstream) | A | G | 0.0039 | 1.75 (1.32, 2.34) | 1.19E-4 | 0.0039\ | 0.89 (0.56, 1.41) | 0.615 |

Abbreviations: Chr, chromosome; EFF, Effect allele; REF, reference allele; EAF, effect allele frequency; OR, Odds ratio; CI, Confidence interval; *P*, P-value

*R62H was not significantly associated with LOAD in primary analyses

**(c)** Conditional analyses of *NCK2* variant rs143080277 on *APOE* ε2 and *TREM2* R47H, and interaction analysis; ORs, 95% confidence intervals, and *P*-values are for rs143080277 main effects

| Variant RSID | Chrom. | Basepair (hg19) | Adjustment for/ Interaction with | EFF | REF | ADGC Meta-Analysis | | | Adjusted | | | Interaction (Main Effects) | | |
|---|---|---|---|---|---|---|---|---|---|---|---|---|---|---|
| | | | | | | EAF | OR (95% CI) | *P* | EAF | OR (95% CI) | *P* | EAF | OR (95% CI) | *P* |
| rs143080277 | 2 | 106366056 | *APOE* ε2 | C | T | 0.0070 | 1.41 (1.10, 1.80) | 0.00699 | 0.0055 | 1.53 (1.18, 1.98) | 0.00139 | 0.0056 | 1.71 (1.34, 2.20) | 2.36E-5 |
| rs143080277 | 2 | 106366056 | *TREM2* R47H | C | T | 0.0070 | 1.41 (1.10, 1.80) | 0.00699 | 0.0055 | 1.48 (1.16, 1.90) | 0.00191 | 0.0056 | 1.44 (1.13, 1.84) | 0.00315 |

Abbreviations: Chr, chromosome; EFF, Effect allele; REF, reference allele; EAF, effect allele frequency; OR, Odds ratio; CI, Confidence interval; *P*, P-value

**Supplementary Table 10.** Rare Variants (MAF<0.01) Gene-Based Association using variants filtered at two weighted-average imputation quality thresholds, $R^2$>0.8 and $R^2$>0.3, to exclude poorly imputed rare variants.

(a) SKAT-O

| Gene | CHR | BP (5' SNP) | Rho | cMAF | # SNPs | $P_{min}$ | $P$ |
|---|---|---|---|---|---|---|---|
| **Genes containing only variants with $R^2$>0.8 (Bonferroni α=1.36E-6)** | | | | | | | |
| *CLASRP* | 19 | 45542471 | 0 | 0.0094 | 1 | 1.96E-20 | 1.96E-20 |
| *TREM2* | 6 | 41126268 | 0 | 0.0261 | 20 | 5.51E-8 | 9.67E-13 |
| *PSMC3* | 11 | 47440758 | 0 | 0.0133 | 3 | 5.71E-5 | 6.59E-5 |
| **Genes containing only variants with $R^2$>0.3 (Bonferroni α=1.36E-6)** | | | | | | | |
| *CLASRP* | 19 | 45542471 | 0 | 0.1422 | 94 | 6.63E-62 | 3.69E-62 |
| *TREM2* | 6 | 41126268 | 0 | 0.0414 | 42 | 3.68E-8 | 1.82E-10 |
| *PSMC3* | 11 | 47440758 | 0 | 0.0435 | 31 | 2.71E-5 | 4.52E-5 |
| *GCA* | 2 | 163200678 | 0 | 0.2737 | 142 | 4.76E-5 | 6.51E-5 |
| *KRT16P1* | 17 | 18345569 | 0 | 0.0004 | 1 | 7.05E-5 | 7.05E-5 |
| *FLG* | 1 | 152274685 | 0 | 0.2136 | 201 | 6.06E-5 | 7.57E-5 |

Abbreviations: Chr, chromosome; BP, basepair; SNP, single nucleotide polymorphism; cMAF, cumulative minor allele frequency; $P$, P-value; $P_{min}$, minimal P-value (P-value for most significantly associated SNP)

(b) Burden Tests

| Gene | CHR | BP (5' SNP) | cMAF (Used/Total) | # SNPs (Used/Total) | OR (95% CI) | *P* |
|---|---|---|---|---|---|---|
| **Genes containing only variants with $R^2$>0.8 (Bonferroni α=1.30E-6)** | | | | | | |
| *CLASRP* | 19 | 45542471 | 0.0094/11.1051 | 1/42 | 2.05 (1.76, 2.38) | 1.96E-20 |
| *OXNAD1* | 3 | 16306721 | 0.1514/42.9974 | 48/277 | 1.03 (1.02, 1.05) | 9.14E-5 |
| **Genes containing only variants with $R^2$>0.3 (Bonferroni α=1.30E-6)** | | | | | | |
| *CLASRP* | 19 | 45542471 | 0.1422/11.706 | 94/154 | 1.23 (1.18, 1.28) | 7.12E-22 |
| *LRFN2* | 6 | 40359473 | 1.6142/146.8429 | 1182/1947 | 1.01 (1.00, 1.01) | 1.30E-5 |
| *ANO6* | 12 | 45609776 | 1.6861/33.1218 | 1279/1765 | 1.01 (1.00, 1.01) | 2.25E-5 |
| *UGT2B4* | 4 | 70345904 | 0.3007/45.3293 | 250/423 | 1.03 (1.02, 1.05) | 2.42E-5 |
| *TREM2* | 6 | 41126268 | 0.0414/0.1128 | 42/46 | 1.08 (1.04, 1.12) | 4.16E-5 |
| *RAET1E-AS1* | 6 | 150184722 | 0.3707/39.6613 | 395/562 | 1.02 (1.01, 1.03) | 4.88E-5 |
| *KRT16P1* | 17 | 18345569 | 0.0004/0.0004 | 1/1 | 6.69 (2.62, 17.09) | 7.05E-5 |
| *KCNH7* | 2 | 163227985 | 3.6758/122.4768 | 1818/2702 | 1.003 (1.001, 1.004) | 8.09E-5 |
| *FLG* | 1 | 152274685 | 0.2136/7.6337 | 201/270 | 1.03 (1.02, 1.05) | 8.57E-5 |

Abbreviations: Chr, chromosome; BP, basepair; SNP, single nucleotide polymorphism; cMAF, cumulative minor allele frequency; OR, Odds ratio; CI, Confidence interval; *P*, P-value

Shaded box indicates statistical significance for a Bonferroni-corrected α threshold based on the number of separate genes/regions in each analysis.

**Supplementary Table 13.** Rare Variants (MAF<0.01) Gene-Based Association using variants filtered at multiple predicted deleteriousness thresholds (CADD≥5, ≥10, and ≥15) using a weighted-average imputation quality threshold of $R^2$>0.3 to exclude poorly imputed rare variants.

(a) SKAT-O

| Gene | CHR | BP (5' SNP) | Rho | cMAF | # SNPs | $P_{min}$ | $P$ |
|---|---|---|---|---|---|---|---|
| **Genes containing only variants with CADD>5 and $R^2$>0.3 (Bonferroni α=1.32E-6)** | | | | | | | |
| *TREM2* | 6 | 41126268 | 0 | 0.0135 | 16 | <E-120 | 6.23E-17 |
| *PPP1R37* | 19 | 45596453 | 0 | 0.0767 | 53 | 4.93E-8 | 4.328E-7 |
| *LOC149086* | 1 | 31971908 | 1 | 0.0013 | 3 | 2.03E-5 | 2.33E-5 |
| *GCA* | 2 | 163200678 | 0 | 0.0927 | 47 | 1.71E-5 | 2.43E-5 |
| *ORC2* | 2 | 201774974 | 0 | 0.0890 | 76 | 1.76E-5 | 3.08E-5 |
| *KRT16P1* | 17 | 18345569 | 0 | 0.0004 | 1 | 7.05E-5 | 7.05E-5 |
| **Genes containing only variants with CADD>10 and $R^2$>0.3 (Bonferroni α=1.42E-6)** | | | | | | | |
| *TREM2* | 6 | 41126268 | 0 | 0.0108 | 9 | <E-120 | 6.24E-17 |
| *BCAM* | 19 | 45312738 | 0 | 0.0171 | 13 | <E-120 | 1.36E-12 |
| *TMEM72* | 10 | 45406996 | 0 | 0.0013 | 7 | 7.40E-6 | 9.88E-6 |
| *DENND1C* | 19 | 6467374 | 0 | 0.0025 | 5 | 1.17E-5 | 1.29E-5 |
| *FBL* | 19 | 40325254 | 1 | 0.0080 | 5 | 1.91E-5 | 2.24E-5 |
| *LOC149086* | 1 | 31971908 | 0 | 0.0012 | 2 | 3.28E-5 | 3.68E-5 |
| *KRT16P1* | 17 | 18345569 | 0 | 0.0004 | 1 | 7.05E-5 | 7.05E-5 |
| **Genes containing only variants with CADD>15 and $R^2$>0.3 (Bonferroni α=1.76E-6)** | | | | | | | |
| *TREM2* | 6 | 41126268 | 0 | 0.0082 | 7 | <E-120 | 6.89E-17 |
| *TOMM40* | 19 | 45394601 | 0 | 0.0023 | 3 | 4.09E-7 | 6.97E-8 |
| *DENND1C* | 19 | 6467374 | 0 | 0.0024 | 5 | 1.17E-5 | 1.29E-5 |
| *LOC149086* | 1 | 31971908 | 0 | 0.0007 | 1 | 2.35E-5 | 2.35E-5 |
| *TMEM72* | 10 | 45406996 | 0 | 0.0009 | 6 | 4.57E-5 | 5.81E-5 |
| *FBXW2* | 9 | 123519826 | 1 | 0.0042 | 5 | 4.70E-5 | 5.81E-5 |

Abbreviations: Chr, chromosome; BP, basepair; SNP, single nucleotide polymorphism; cMAF, cumulative minor allele frequency; *P*, P-value; $P_{min}$, minimal P-value (P-value for most significantly associated SNP)

Shaded box indicates statistical significance for a Bonferroni-corrected α threshold based on the number of separate genes/regions in each analysis.

(b) Burden Tests

| Gene | CHR | BP (5' SNP) | cMAF (Used/Total) | # SNPs (Used/Total) | OR (95% CI) | *P* |
|---|---|---|---|---|---|---|
| **Genes containing only variants with CADD>5 and $R^2$>0.3 (Bonferroni α=1.33E-6)** | | | | | | |
| *TREM2* | 6 | 41126268 | 0.0135/0.0368 | 16/18 | 1.34 (1.22, 1.48) | 4.11E-9 |
| *CLASRP* | 19 | 45542471 | 0.0228/1.6005 | 18/30 | 1.32 (1.18, 1.47) | 7.98E-7 |
| *LOC149086* | 1 | 31971908 | 0.0013/3.1372 | 3/12 | 2.97 (1.8, 4.91) | 2.03E-5 |
| *KRT16P1* | 17 | 18345569 | 0.0004/0.0004 | 1/1 | 6.69 (2.62, 17.09) | 7.05E-5 |
| *FBXL17* | 5 | 107194762 | 1.2051/81.4585 | 1137/1531 | 1.01 (1.01, 1.02) | 7.70E-5 |
| *RAB11FIP5* | 2 | 73300560 | 0.0687/4.0197 | 68/93 | 1.1 (1.05, 1.16) | 8.51E-5 |
| **Genes containing only variants with CADD>10 and $R^2$>0.3 (Bonferroni α=1.42E-6)** | | | | | | |
| *TREM2* | 6 | 41126268 | 0.0108/0.021 | 9/10 | 1.51 (1.34, 1.70) | 4.04E-12 |
| *BCAM* | 19 | 45312738 | 0.0171/0.7742 | 13/25 | 1.56 (1.37, 1.78) | 1.53E-11 |
| *FBL* | 19 | 40325254 | 0.008/0.3108 | 5/8 | 1.48 (1.24, 1.78) | 1.917E-5 |
| *DENND1C* | 19 | 6467374 | 0.0025/0.4211 | 5/9 | 0.46 (0.32, 0.67) | 3.17E-5 |
| *FLG* | 1 | 152274685 | 0.0868/2.9552 | 84/112 | 1.07 (1.04, 1.11) | 6.30E-5 |
| *LOC149086* | 1 | 31971908 | 0.0012/0.8383 | 2/4 | 2.86 (1.71, 4.78) | 6.66E-5 |
| *KRT16P1* | 17 | 18345569 | 0.0004/0.0004 | 1/1 | 6.69 (2.62, 17.09) | 7.05E-5 |
| **Genes containing only variants with CADD>15 and $R^2$>0.3 (Bonferroni α=1.76E-6)** | | | | | | |
| *TREM2* | 6 | 41126268 | 0.0082/0.0184 | 7/8 | 1.82 (1.57, 2.12) | 1.52E-15 |
| *BCAM* | 19 | 45312738 | 0.0113/0.3063 | 9/16 | 1.57 (1.34, 1.84) | 1.63E-8 |
| *LOC149086* | 1 | 31971908 | 0.0007/0.0007 | 1/1 | 4.49 (2.24, 9.02) | 2.35E-5 |
| *DENND1C* | 19 | 6467374 | 0.0025/0.0502 | 5/7 | 0.46 (0.32, 0.67) | 3.17E-5 |
| *FBXW2* | 9 | 123519826 | 0.0042/0.0174 | 5/6 | 0.62 (0.50, 0.78) | 4.70E-5 |

Abbreviations: Chr, chromosome; BP, basepair; SNP, single nucleotide polymorphism; cMAF, cumulative minor allele frequency; OR, Odds ratio; CI, Confidence interval; *P*, P-value

Shaded box indicates statistical significance for a Bonferroni-corrected α threshold based on the number of separate genes/regions in each analysis.

**Supplementary Table 19.** Descriptive Characteristics of Modules used in MAGMA Co-expression Analysis

| Study | PubMed ID | Tissue | Sample size | # Genes | # Modules | Module Size Range |
|---|---|---|---|---|---|---|
| BRAINEAC | 25174004 | 10 brain tissues | 134 controls | 15,595 | 219 | 34-8,417 |
| Common Mind | 27668389 | Prefrontal cortex | 258 SCZ + 279 controls | 14,729 | 60 | 27-8,636 |
| Gandal et al . | 29439242 | Cerebral Cortex | 407 ASD/SCZ/MDD, 293 controls | 11,245 | 14 | 110-5,034 |
| Gibbs et al. | 20485568 | Cerebellum, temporal cortex, frontal cortex, pons | 150 controls | 12,733 | 117 | 22-3,443 |
| Neueder et al. | 25358814 | Cerebellum, frontal cortex, caudate nucleus | 44 HD, 36 control | 12,506 | 124 | 7-3,610 |
| Mostafavi et al. | 29802388 | Dorsolateral prefrontal cortex | 478 control + MCI + dementia | 13,136 | 47 | 21-553 |
| PsychEncode | 30545856 | Frontal and temporal cortex | 1,695 ASD + SCZ + BPD + control | 25,774 | 34 | 54-2,645 |
| Zhang et al. | 23622250 | Prefrontal cortex, visual cortex, cerebellum | 376 AD patients + 173 controls | 8,792 | 62 | 41-1,192 |
| Fairfax et al. | 24604202 | Monocytes | 432 controls | 11,364 | 62 | 25-7,402 |

**Supplementary Figure 1.** Genome-wide Manhattan plot and Q-Q Plot for common variant (MAF>0.01) associations in Stage 1 IGAP analysis. Names of loci in red indicate known loci with genome-wide significance (GWS; $P<5\times10^{-8}$) after Stage 3, in blue indicate novel loci with GWS after Stage 3, and in green indicate loci with discovery-level significance ($P<10^{-5}$) in Stage 1 that did not attain GWS in Stage 3 analysis.

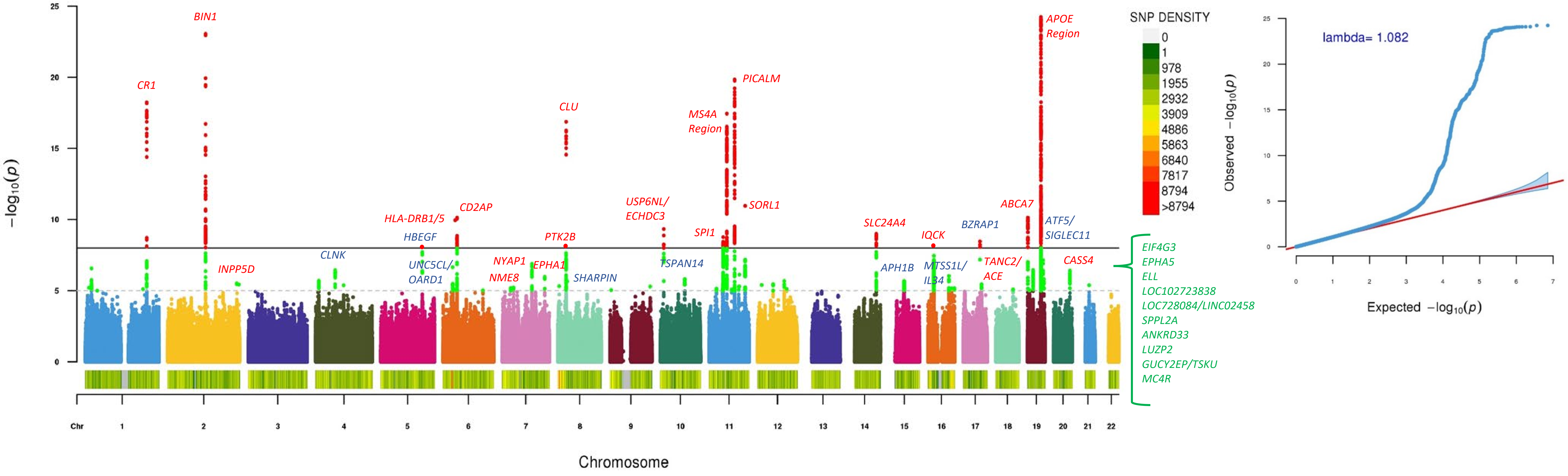

**Supplementary Figure 2.** Genome-wide Manhattan plot and Q-Q Plot for rare variant (MAF≤0.01) associations in Stage 1 IGAP analysis. Names of loci in red indicate known loci with genome-wide significance (GWS; $P<5\times10^{-8}$) after Stage 3, in blue indicate novel loci with GWS after Stage 3, and in green indicate loci with discovery-level significance ($P<10^{-5}$) in Stage 1 that did not attain GWS in Stage 3 analysis.

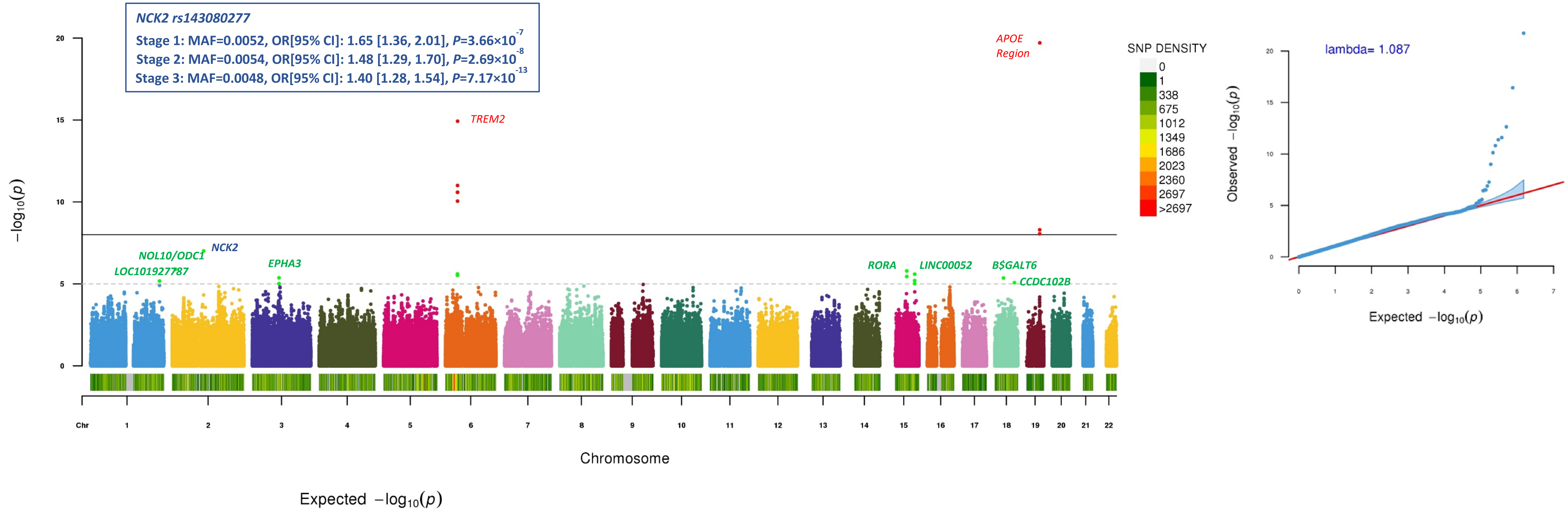

**Supplementary Figure 3.** Genome-wide Manhattan plots and Q-Q plots for (a) common variant (MAF>0.01) and (b) rare variant (MAF≤0.01) associations in Stage 1 analysis of the UK Biobank GWAS of AD/dementia parental history. Names of loci in red indicate known loci with genome-wide significance (GWS; $P<5\times10^{-8}$) after Stage 1, while names in blue indicate novel loci with GWS after Stage 3.

(a) Common variants (MAF>0.01)

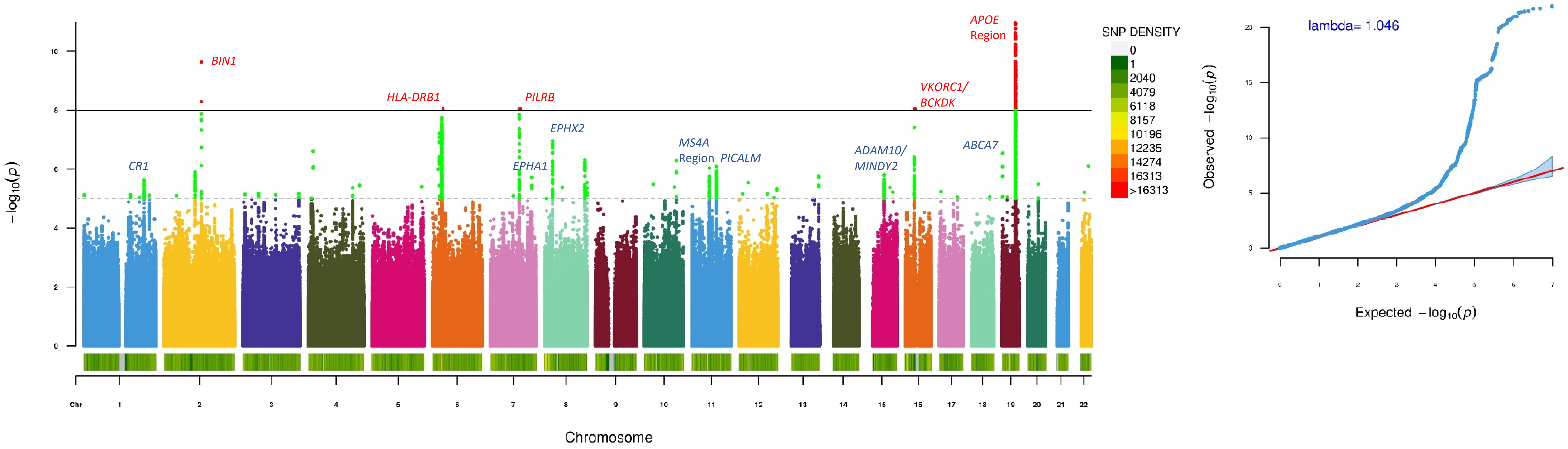

(b) Rare variants (MAF≤0.01)

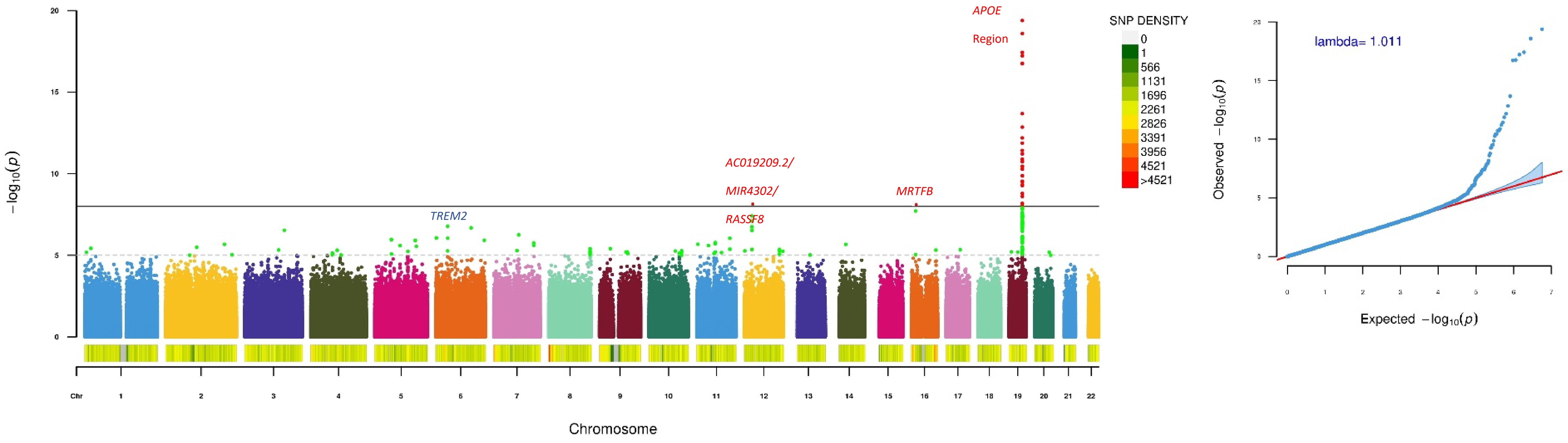

**Supplementary Figure 4.** Individual regional association plots from LocusZoom and Forest plots of Stage 1-3 associations for most strongly associated variants in each common variant (MAF>0.01) locus.

(a) LocusZoom and Forest plot of rs7259620 (*APOE*) from IGAP meta-analysis (Stage1)

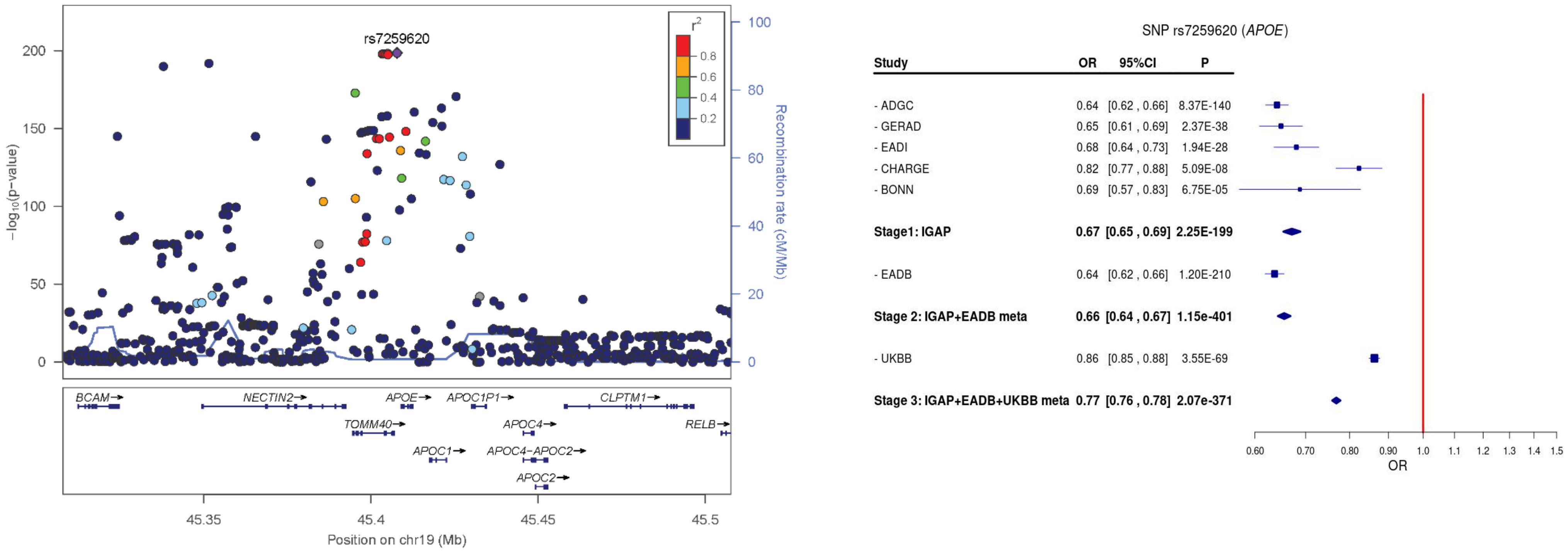

Abbreviations: OR = odds ratio, CI = confidence interval

(b) LocusZoom and Forest plot of rs6733839 (*BIN1*) from IGAP meta-analysis (Stage1)

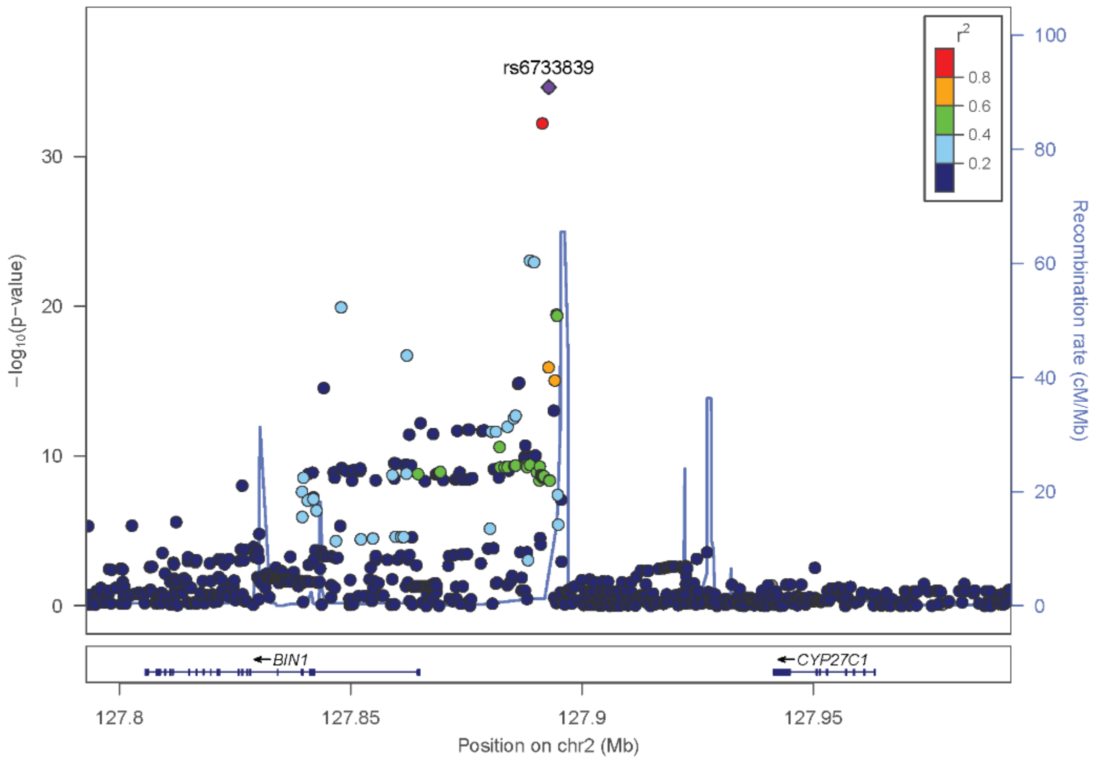

SNP rs6733839 (*BIN1*)

| Study | OR | 95%CI | P |
|---|---|---|---|
| - ADGC | 1.20 | [1.16 , 1.24] | 3.25E-24 |
| - GERAD | 1.22 | [1.14 , 1.31] | 3.13E-09 |
| - EADI | 1.19 | [1.11 , 1.28] | 1.45E-06 |
| - CHARGE | 1.10 | [1.02 , 1.17] | 8.81E-03 |
| - BONN | 1.05 | [0.86 , 1.28] | 6.21E-01 |
| **Stage1: IGAP** | **1.18** | **[1.15 , 1.21]** | **2.36E-35** |
| - EADB | 1.21 | [1.18 , 1.25] | 5.12E-40 |
| **Stage 2: IGAP+EADB meta** | **1.19** | **[1.17 , 1.22]** | **3.06E-73** |
| - UKBB | 1.07 | [1.05 , 1.09] | 1.97E-16 |
| **Stage 3: IGAP+EADB+UKBB meta** | **1.12** | **[1.11 , 1.14]** | **4.66E-73** |

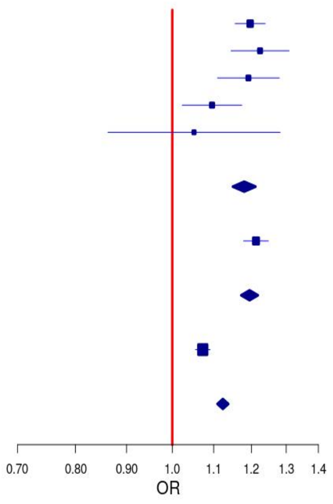

(c) LocusZoom and Forest plot of rs711061 (*PICALM*) from IGAP meta-analysis (Stage1)

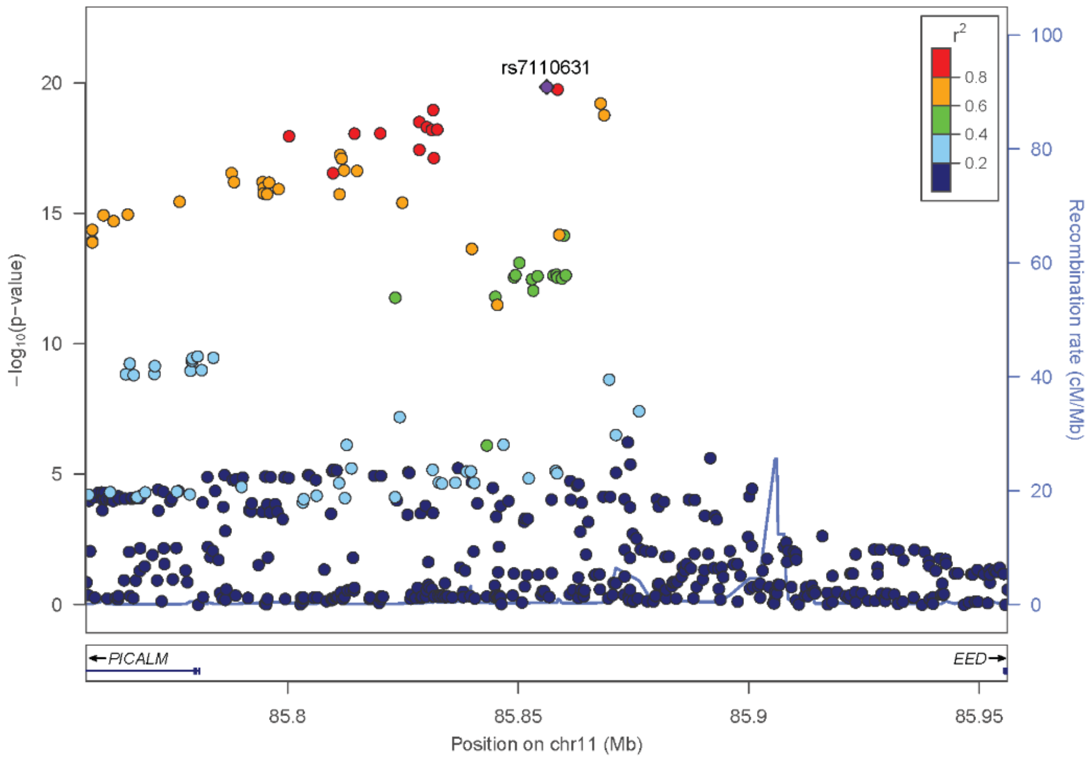

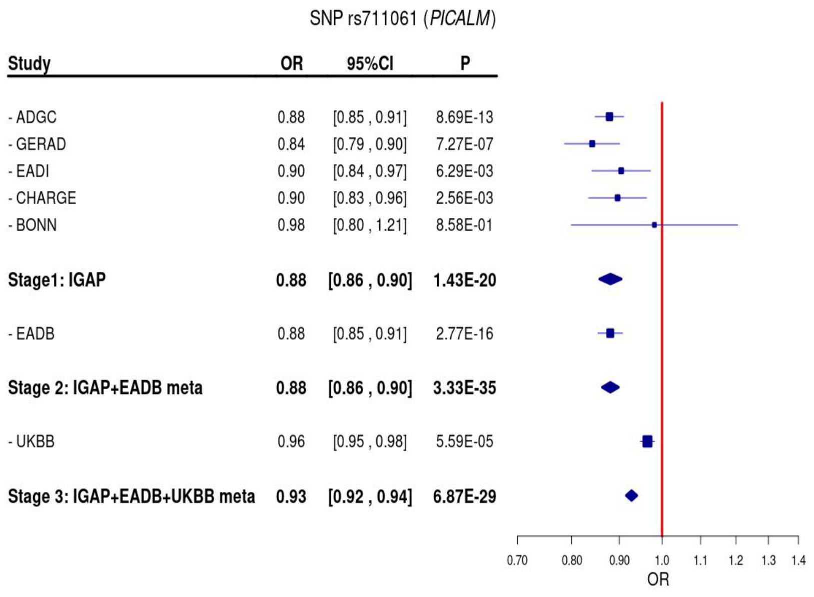

(d) LocusZoom and Forest plot of rs679515 (*CR1*) from IGAP meta-analysis (Stage1)

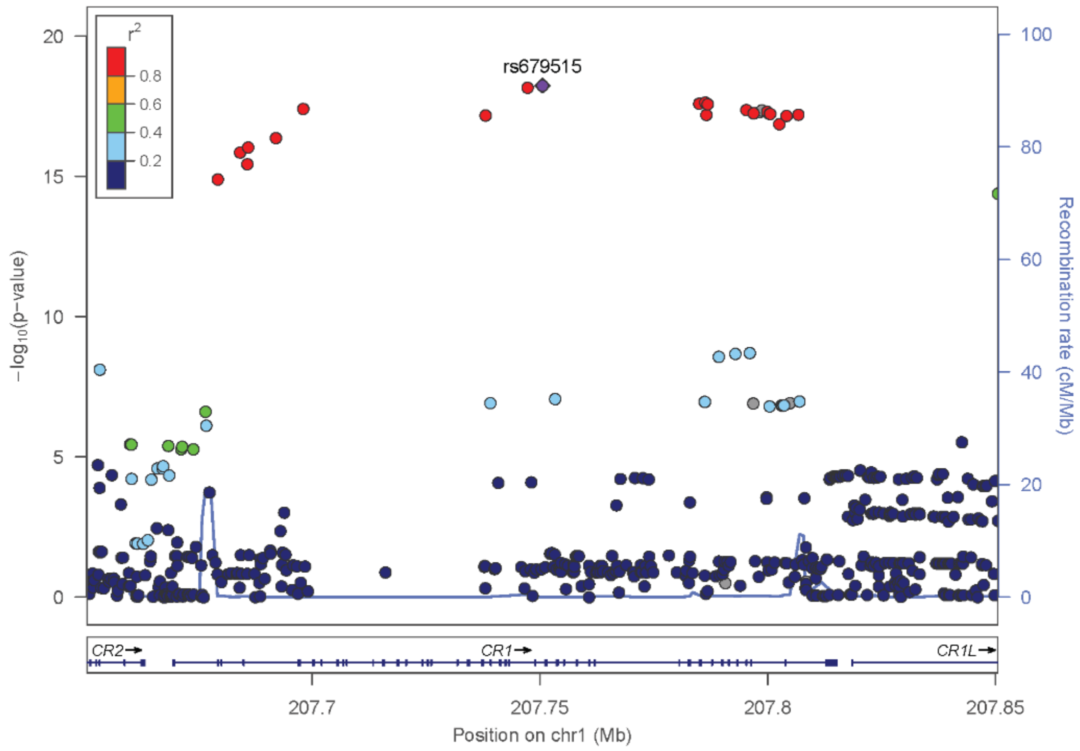

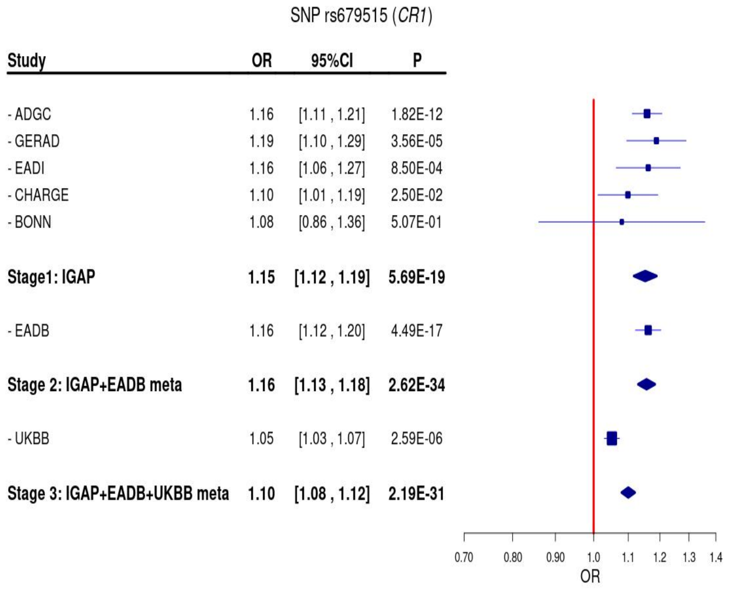

(e) LocusZoom and Forest plot of rs1582763 (*MS4A* Region) from IGAP meta-analysis (Stage1)

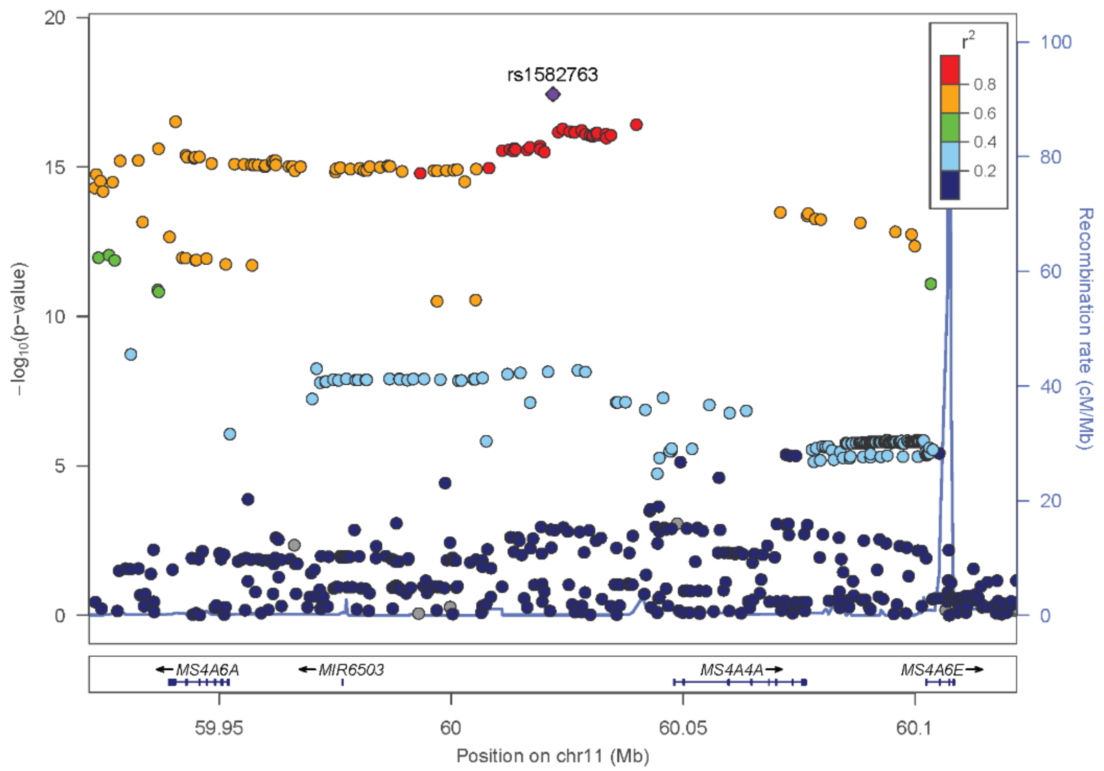

SNP rs1582763 (*MS4A*)

| Study | OR | 95%CI | P |
|---|---|---|---|
| - ADGC | 0.87 | [0.84 , 0.90] | 7.23E-15 |
| - GERAD | 0.87 | [0.81 , 0.92] | 1.86E-05 |
| - EADI | 0.93 | [0.86 , 0.99] | 3.00E-02 |
| - CHARGE | 0.96 | [0.90 , 1.02] | 2.02E-01 |
| - BONN | 0.96 | [0.79 , 1.17] | 6.83E-01 |
| **Stage1: IGAP** | **0.89** | **[0.87 , 0.92]** | **3.63E-18** |
| - EADB | 0.90 | [0.87 , 0.92] | 1.52E-13 |
| **Stage 2: IGAP+EADB meta** | **0.89** | **[0.88 , 0.91]** | **4.08E-30** |
| - UKBB | 0.96 | [0.94 , 0.98] | 2.25E-06 |
| **Stage 3: IGAP+EADB+UKBB meta** | **0.93** | **[0.92 , 0.94]** | **2.66E-28** |

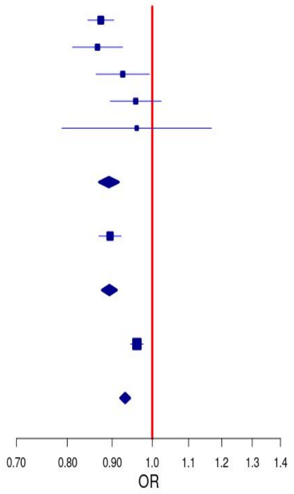

(f) LocusZoom and Forest plot of rs867230 (*CLU*) from IGAP meta-analysis (Stage1)

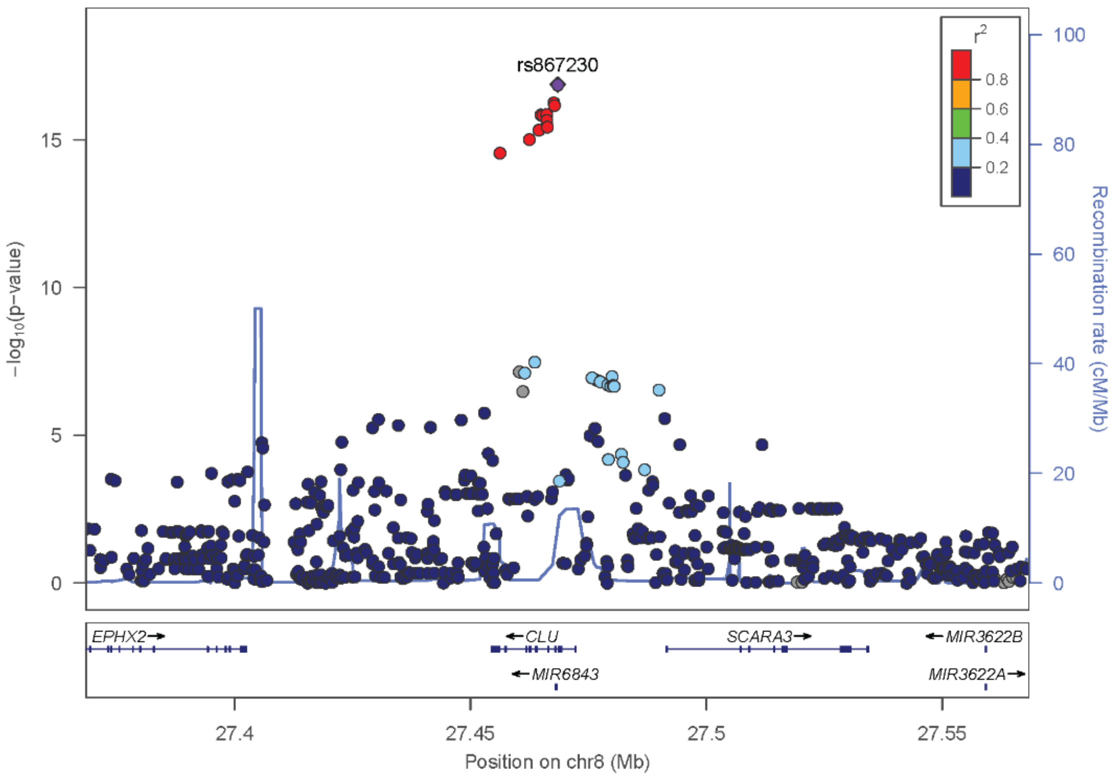

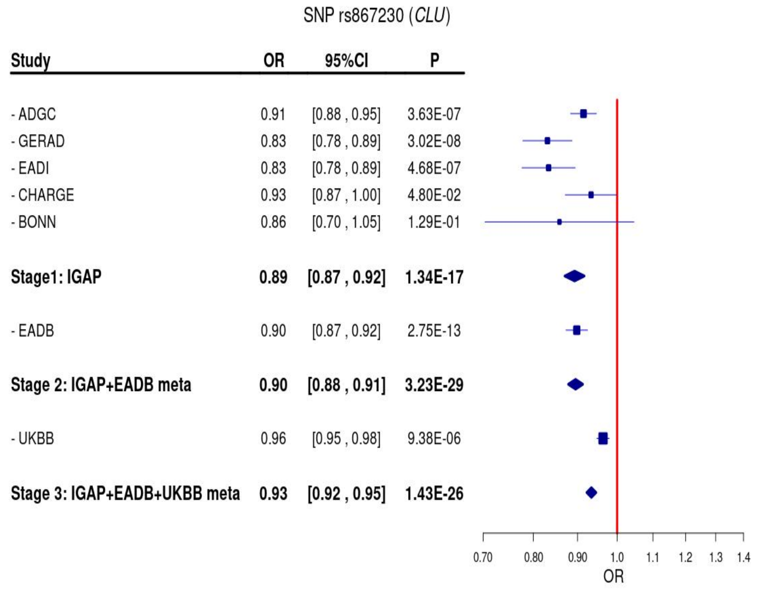

(g) LocusZoom and Forest plot of rs11218343 (*SORL1*) from IGAP meta-analysis (Stage1)

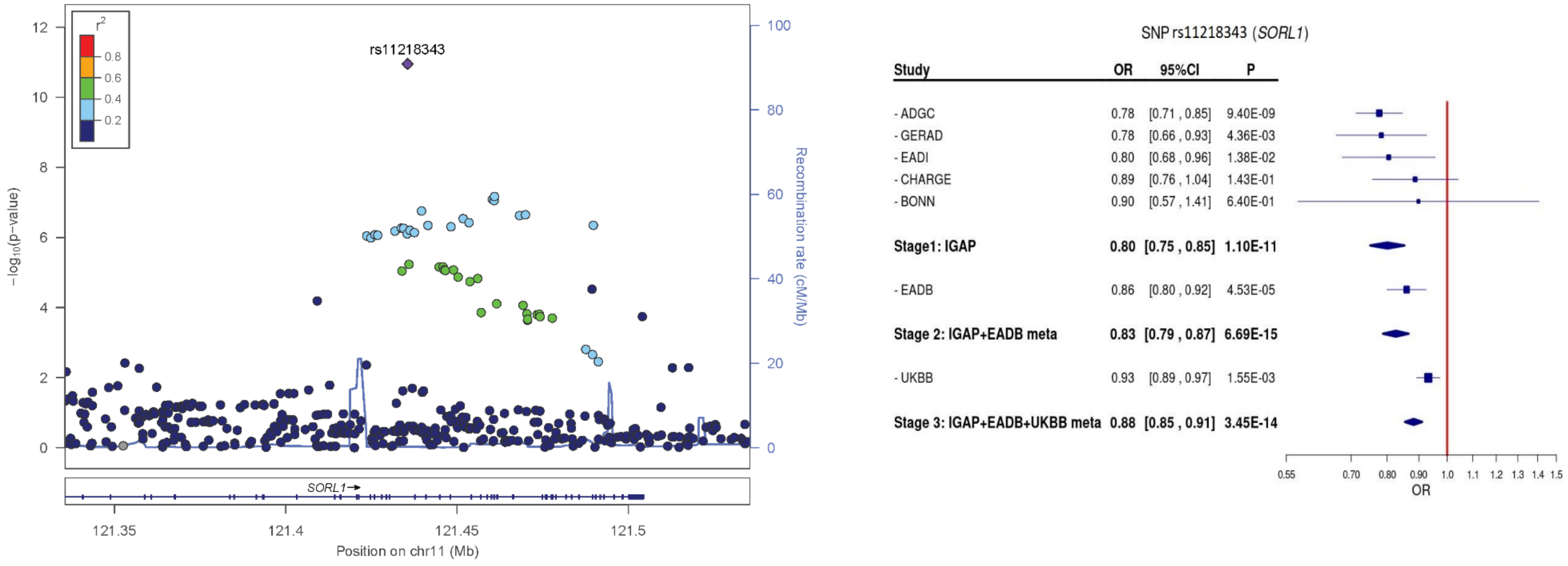

(h) LocusZoom and Forest plot of rs1385742 (*CD2AP*) from IGAP meta-analysis (Stage1)

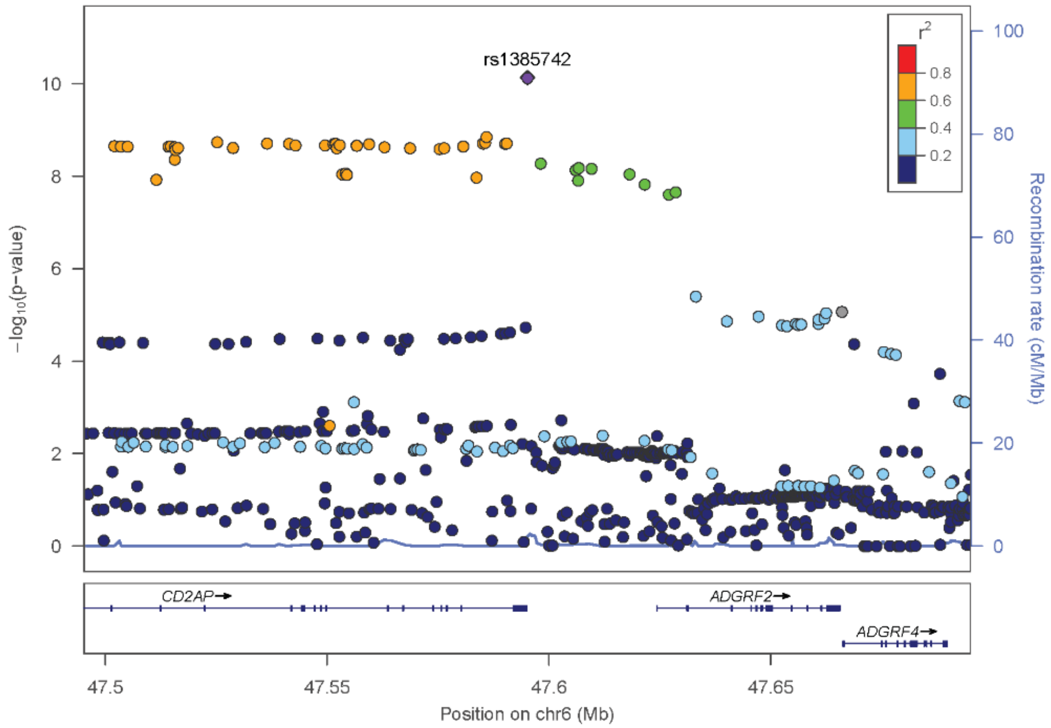

SNP rs1385742 (*CD2AP*)

| Study | OR | 95%CI | P |
|---|---|---|---|
| - ADGC | 1.10 | [1.06 , 1.14] | 3.76E-08 |
| - GERAD | 1.11 | [1.03 , 1.18] | 3.60E-03 |
| - EADI | 1.07 | [0.99 , 1.15] | 7.73E-02 |
| - CHARGE | 1.07 | [1.00 , 1.14] | 6.70E-02 |
| - BONN | 1.00 | [0.82 , 1.22] | 9.81E-01 |
| **Stage1: IGAP** | **1.09** | **[1.06 , 1.12]** | **7.41E-11** |
| - EADB | 1.06 | [1.03 , 1.10] | 7.27E-05 |
| **Stage 2: IGAP+EADB meta** | **1.08** | **[1.06 , 1.10]** | **5.37E-14** |
| - UKBB | 1.03 | [1.01 , 1.05] | 1.65E-03 |
| **Stage 3: IGAP+EADB+UKBB meta** | **1.05** | **[1.04 , 1.06]** | **2.99E-13** |

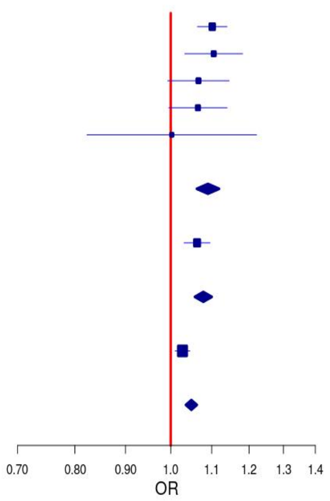

(i) LocusZoom and Forest plot of rs12151021 (*ABCA7*) from IGAP meta-analysis (Stage1)

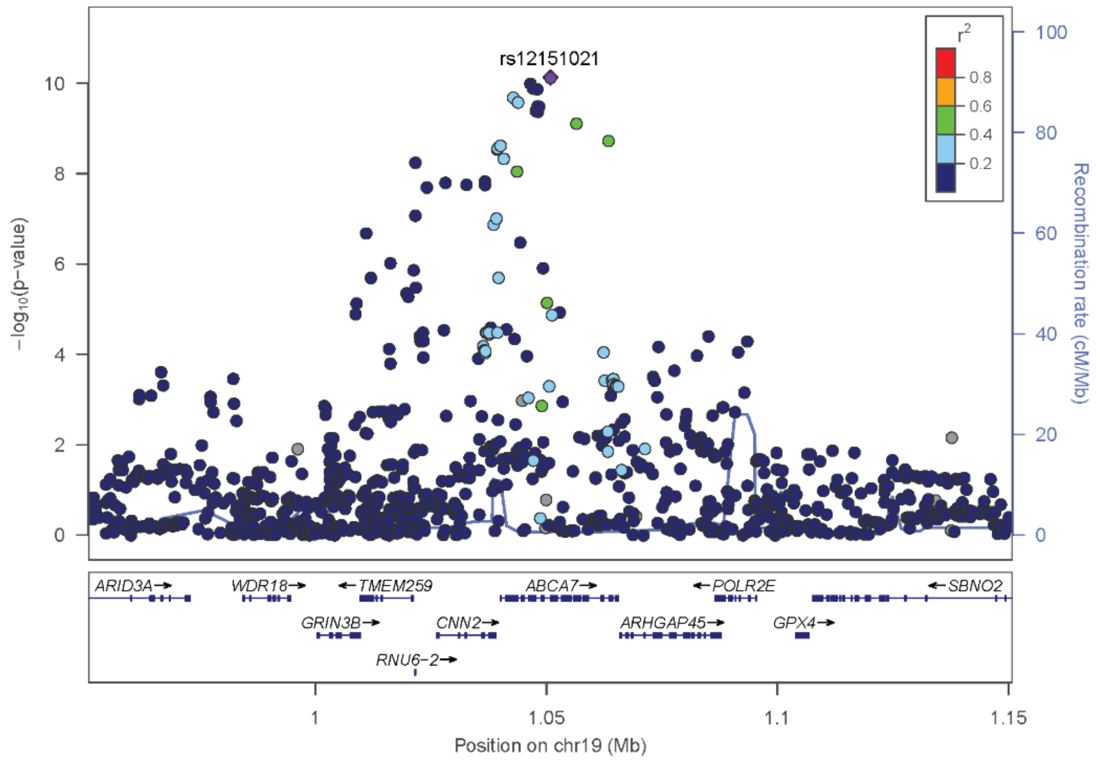

SNP rs12151021 (*ABCA7*)

| Study | OR | 95%CI | P |
|---|---|---|---|
| - ADGC | 1.10 | [1.06 , 1.14] | 1.62E-06 |
| - GERAD | 1.13 | [1.05 , 1.21] | 8.07E-04 |
| - EADI | 1.15 | [1.07 , 1.24] | 1.12E-04 |
| - CHARGE | 1.01 | [0.94 , 1.10] | 7.39E-01 |
| - BONN | 1.10 | [0.90 , 1.35] | 3.48E-01 |
| **Stage1: IGAP** | **1.10** | **[1.07 , 1.13]** | **7.46E-11** |
| - EADB | 1.15 | [1.12 , 1.18] | 8.98E-20 |
| **Stage 2: IGAP+EADB meta** | **1.12** | **[1.10 , 1.14]** | **5.24E-28** |
| - UKBB | 1.03 | [1.01 , 1.05] | 1.74E-03 |
| **Stage 3: IGAP+EADB+UKBB meta** | **1.07** | **[1.05 , 1.08]** | **3.06E-21** |

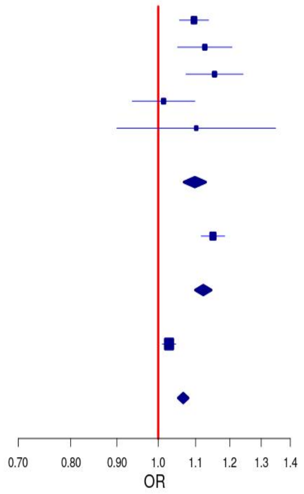

(j) LocusZoom and Forest plot of rs7920721 (*USP6NL/ECHDC3*) from IGAP meta-analysis (Stage1)

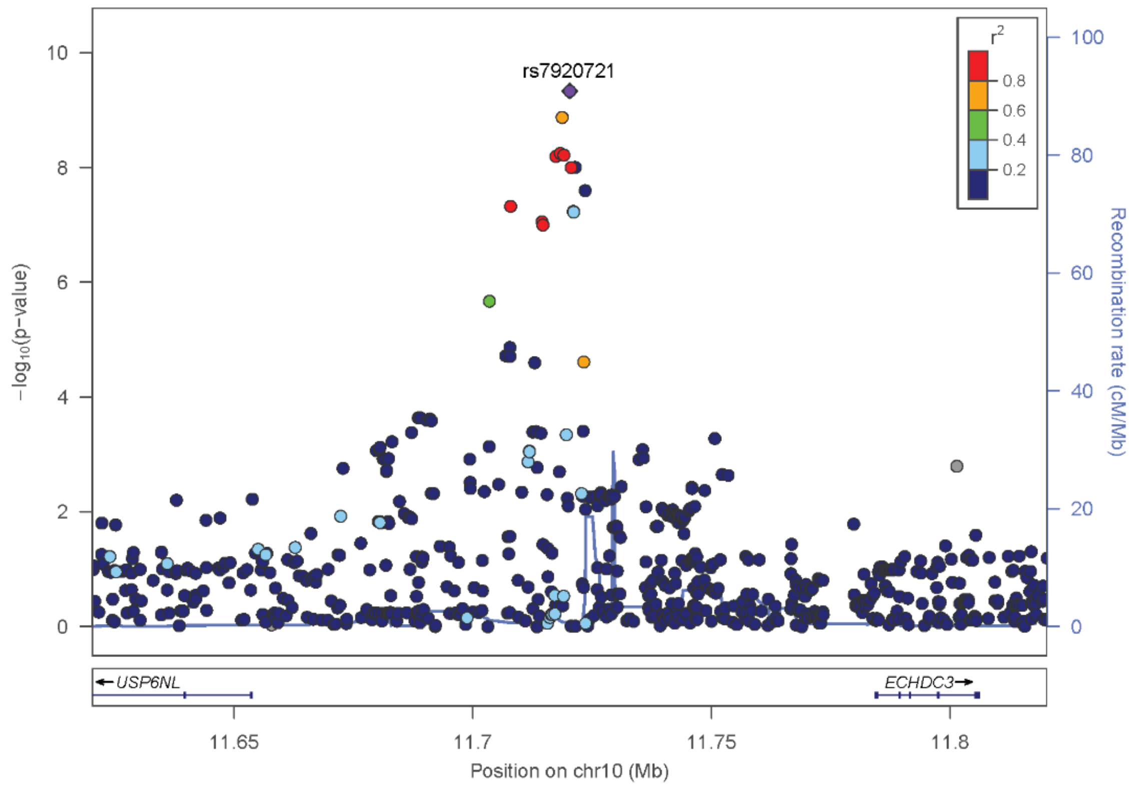

SNP rs7920721 (*USP6NL/ECHDC3*)

| Study | OR | 95%CI | P |
|---|---|---|---|
| - ADGC | 1.09 | [1.06 , 1.13] | 2.99E-07 |
| - GERAD | 1.09 | [1.02 , 1.17] | 7.99E-03 |
| - EADI | 1.07 | [0.99 , 1.14] | 6.96E-02 |
| - CHARGE | 1.08 | [1.01 , 1.16] | 2.28E-02 |
| - BONN | 1.07 | [0.88 , 1.29] | 5.13E-01 |
| **Stage1: IGAP** | **1.09** | **[1.06 , 1.11]** | **4.59E-10** |
| - EADB | 1.07 | [1.03 , 1.10] | 1.90E-05 |
| **Stage 2: IGAP+EADB meta** | **1.08** | **[1.06 , 1.10]** | **7.11E-14** |
| - UKBB | 1.03 | [1.01 , 1.05] | 6.09E-04 |
| **Stage 3: IGAP+EADB+UKBB meta** | **1.05** | **[1.04 , 1.06]** | **7.14E-14** |

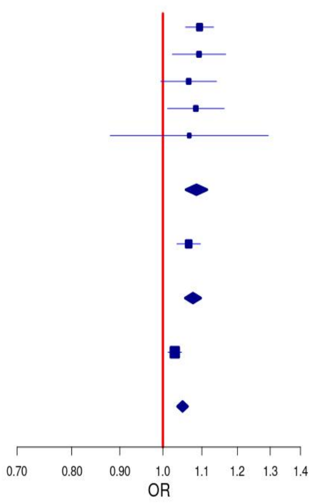

(k) LocusZoom and Forest plot of rs9323877 (*SLC24A4*) from IGAP meta-analysis (Stage1)

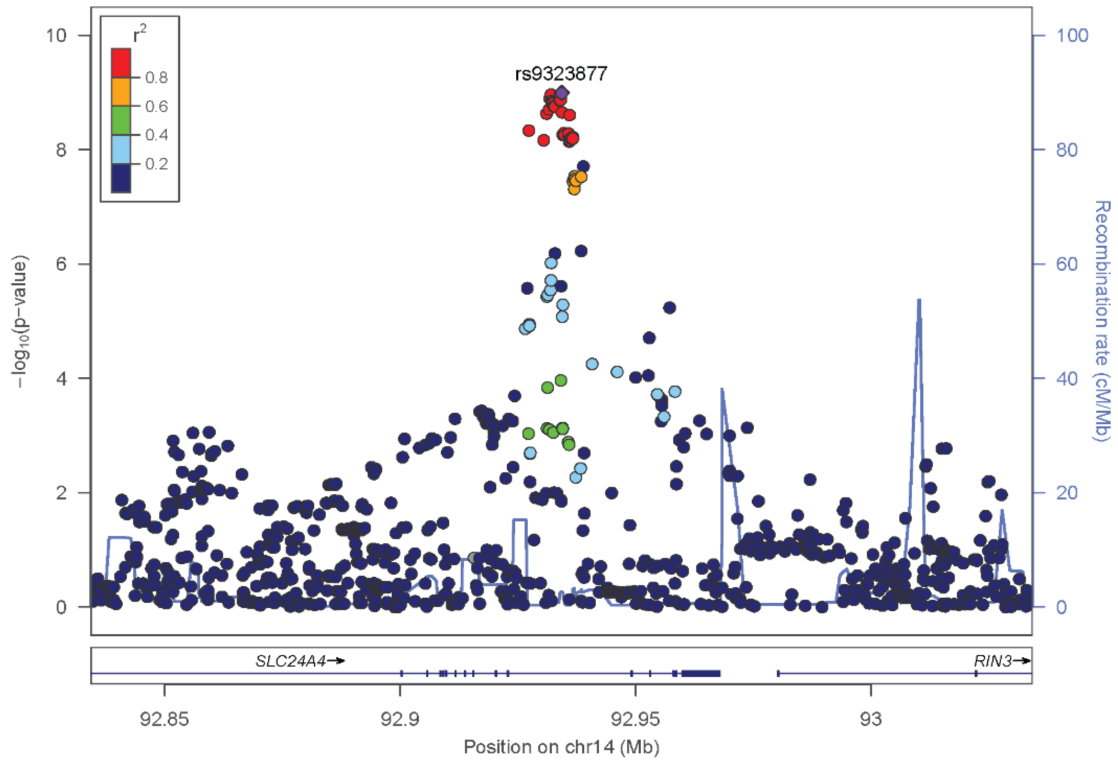

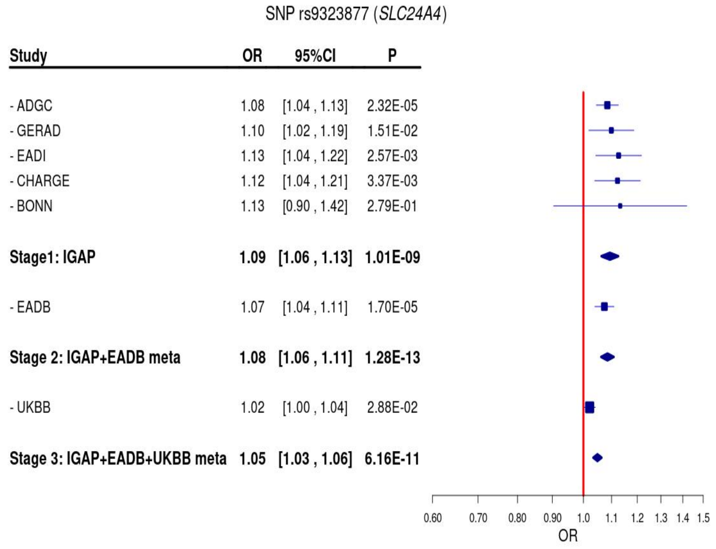

(l) LocusZoom and Forest plot of rs7928419 (*SPI1*) from IGAP meta-analysis (Stage1)

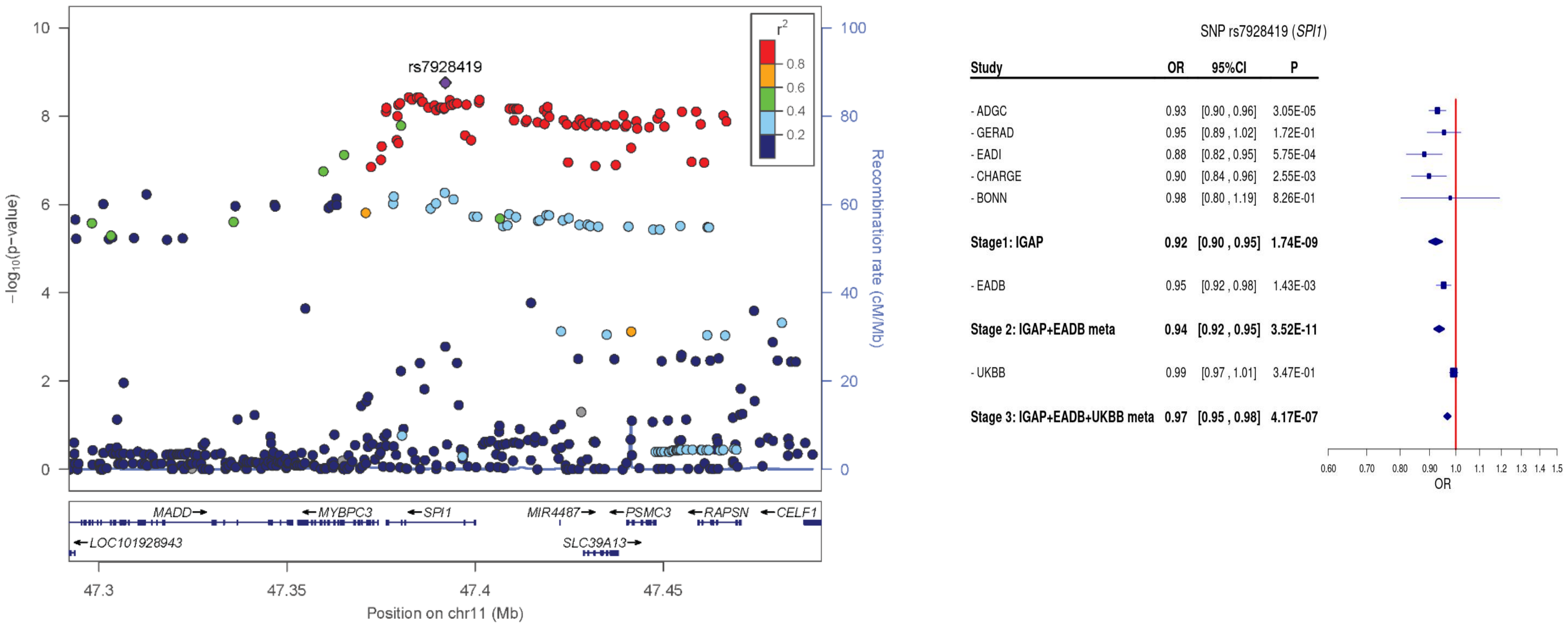

(m)LocusZoom and Forest plot of rs2526377 (*TSPOAP1-AS1/BZRAP1-AS1*) from IGAP meta-analysis (Stage1)

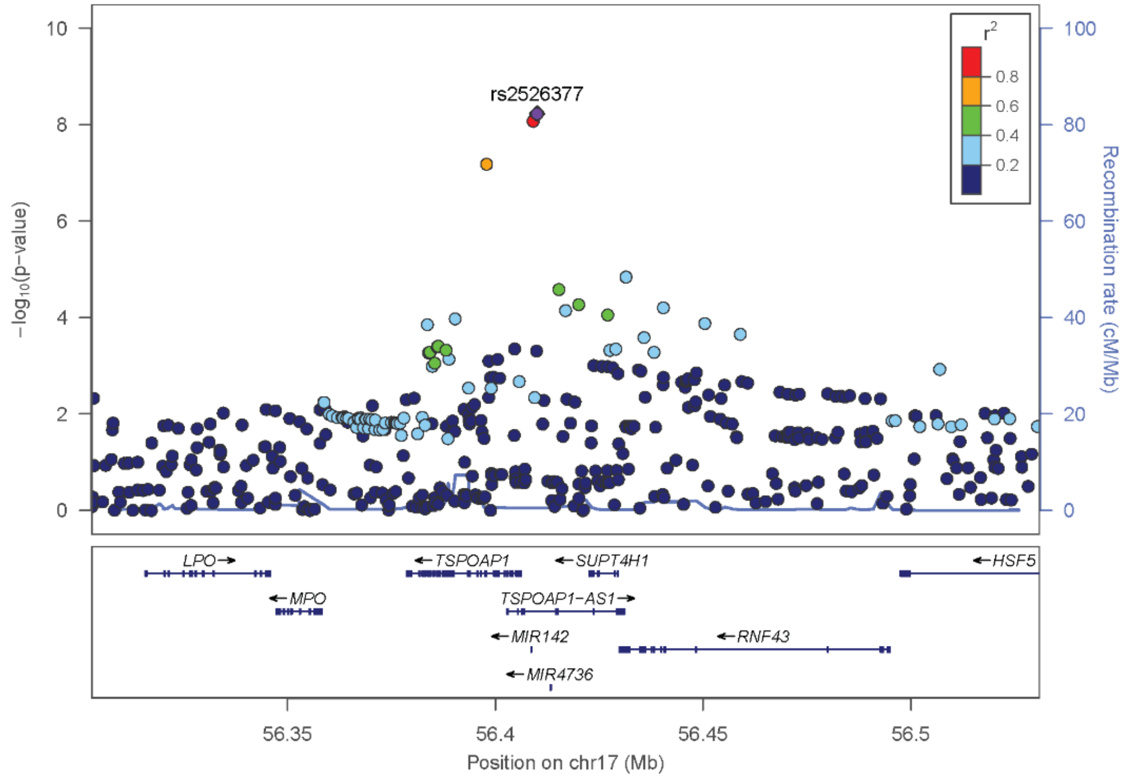

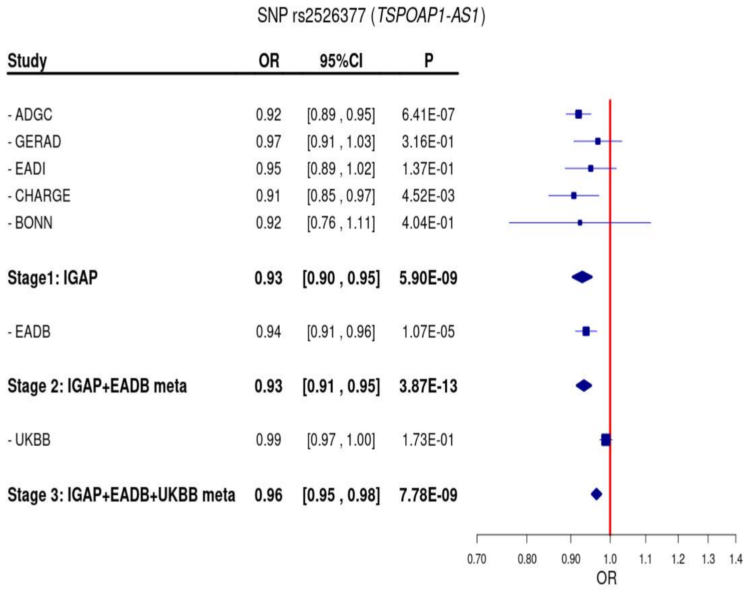

(n) LocusZoom and Forest plot of rs11168036 (*HBEGF*) from IGAP meta-analysis (Stage1)

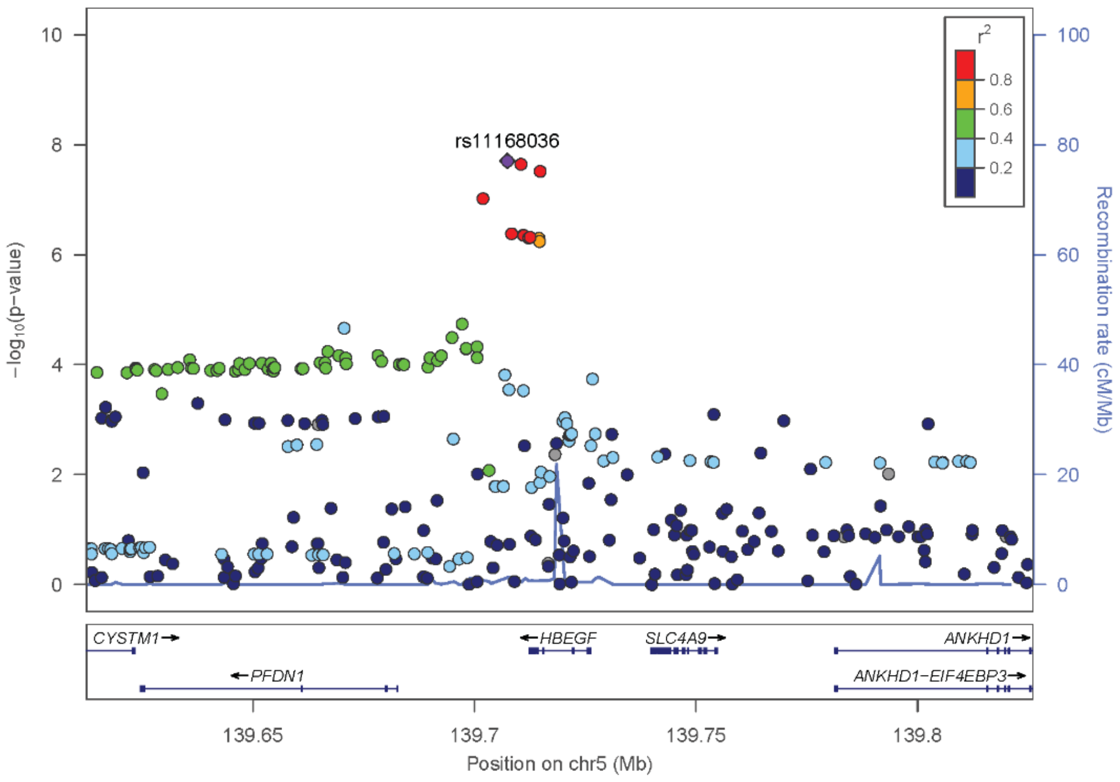

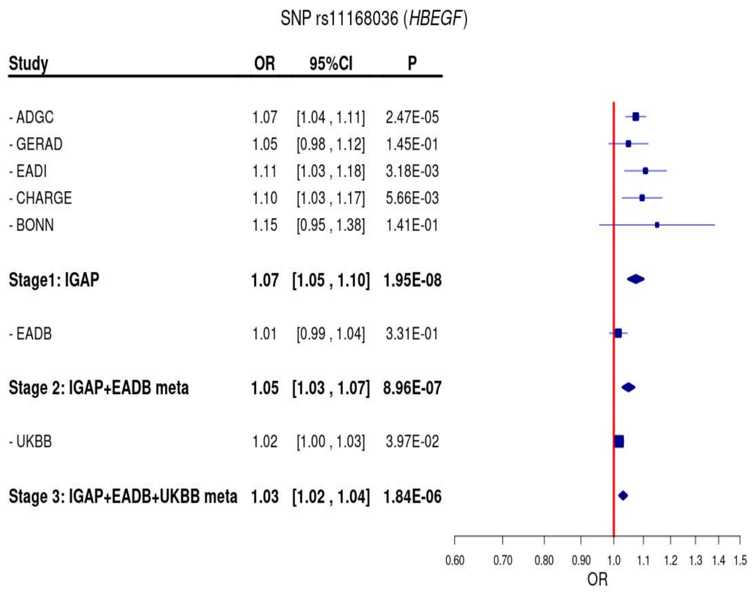

(o) LocusZoom and Forest plot of rs72332431 (*PTK2B*) from IGAP meta-analysis (Stage1)

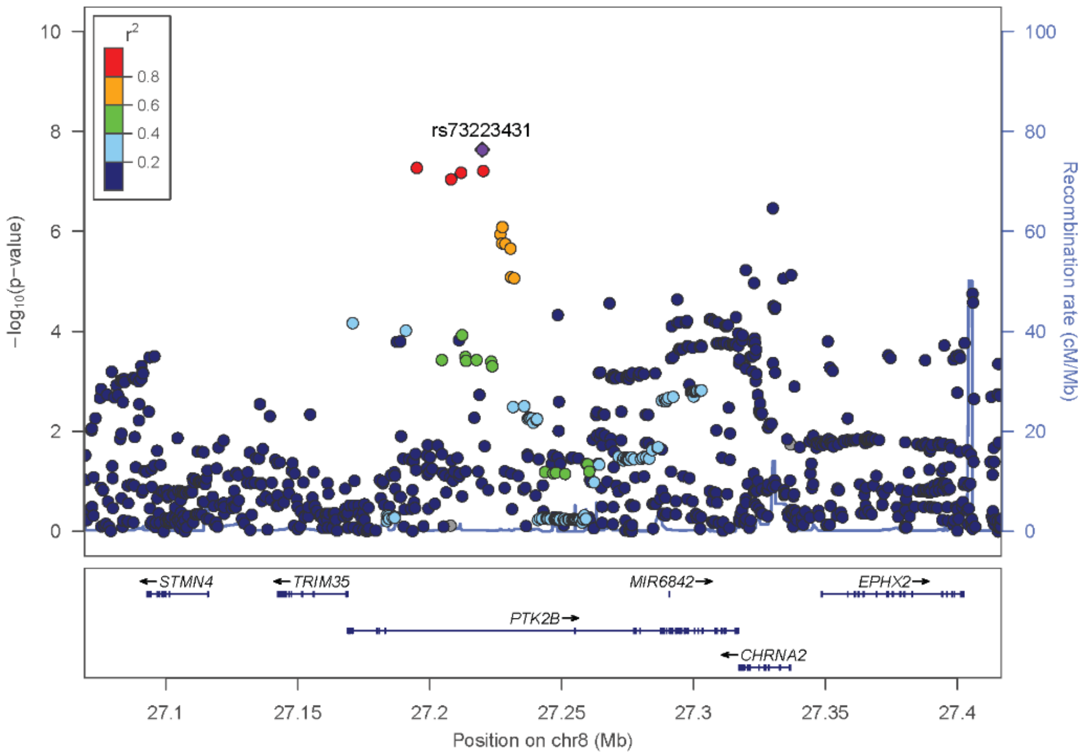

SNP rs72332431 (*PTK2B*)

| Study | OR | 95%CI | P |
|---|---|---|---|
| - ADGC | 1.08 | [1.05 , 1.12] | 4.55E-06 |
| - GERAD | 1.09 | [1.02 , 1.16] | 1.26E-02 |
| - EADI | 1.07 | [1.00 , 1.15] | 5.94E-02 |
| - CHARGE | 1.05 | [0.98 , 1.12] | 1.79E-01 |
| - BONN | 1.05 | [0.87 , 1.28] | 6.06E-01 |
| **Stage1: IGAP** | **1.08** | **[1.05 , 1.10]** | **2.31E-08** |
| - EADB | 1.07 | [1.04 , 1.11] | 1.20E-06 |
| **Stage 2: IGAP+EADB meta** | **1.08** | **[1.06 , 1.10]** | **1.44E-13** |
| - UKBB | 1.04 | [1.02 , 1.05] | 2.67E-05 |
| **Stage 3: IGAP+EADB+UKBB meta** | **1.05** | **[1.04 , 1.07]** | **1.12E-15** |

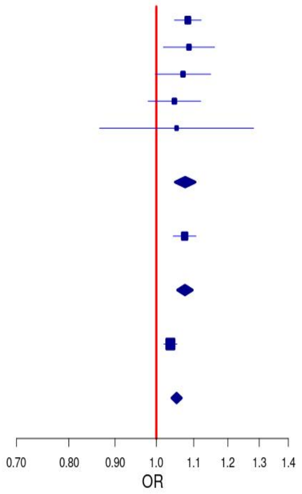

(p) LocusZoom and Forest plot of rs114285994 (*IQCK*) from IGAP meta-analysis (Stage1)

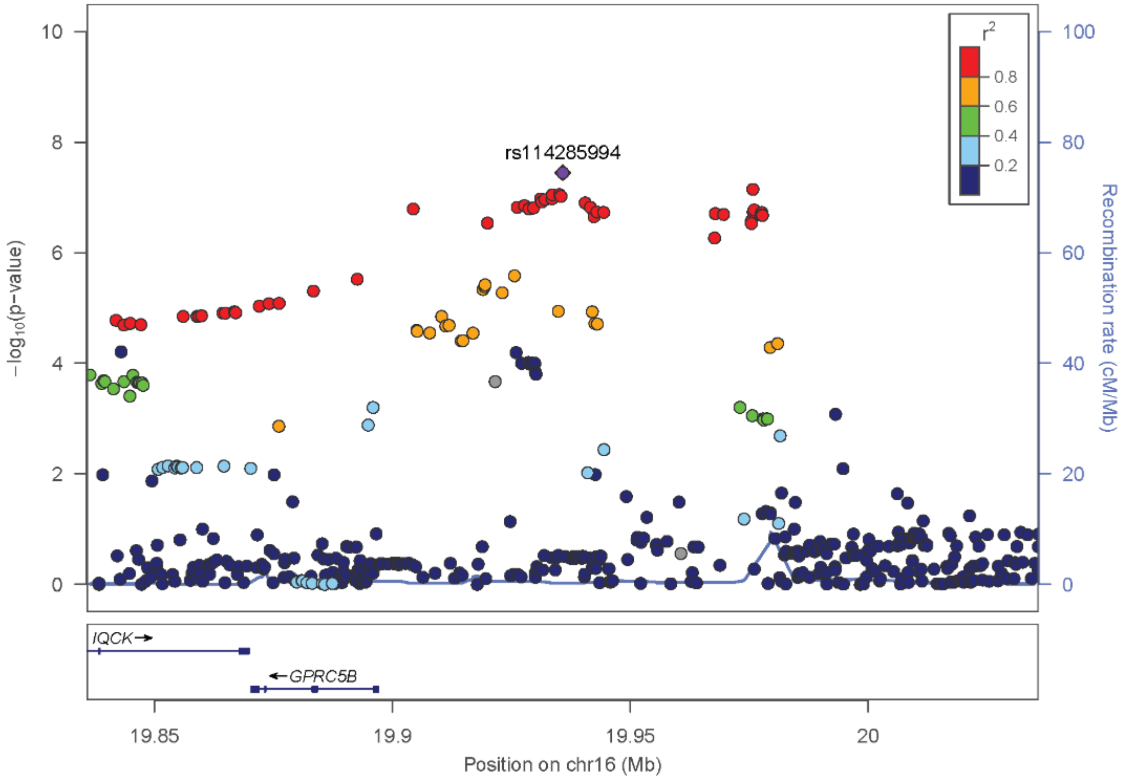

SNP rs114285994 (*IQCK*)

| Study | OR | 95%CI | P |
|---|---|---|---|
| - ADGC | 0.90 | [0.86 , 0.95] | 2.87E-05 |
| - GERAD | 0.92 | [0.84 , 1.01] | 6.46E-02 |
| - EADI | 0.90 | [0.82 , 1.00] | 4.01E-02 |
| - CHARGE | 0.88 | [0.80 , 0.97] | 9.89E-03 |
| - BONN | 0.99 | [0.76 , 1.30] | 9.56E-01 |
| **Stage1: IGAP** | **0.90** | **[0.87 , 0.94]** | **3.53E-08** |
| - EADB | 0.98 | [0.94 , 1.02] | 3.53E-01 |
| **Stage 2: IGAP+EADB meta** | **0.93** | **[0.91 , 0.96]** | **1.54E-06** |
| - UKBB | 1.00 | [0.97 , 1.02] | 8.44E-01 |
| **Stage 3: IGAP+EADB+UKBB meta** | **0.97** | **[0.96 , 0.99]** | **3.05E-03** |

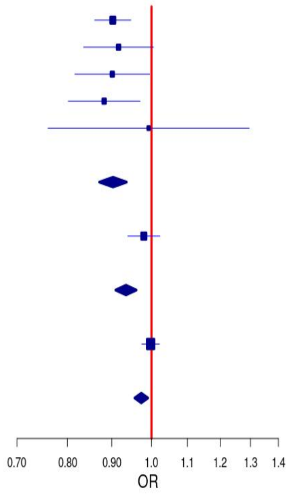

(q) LocusZoom and Forest plot of rs12539172 (*NYAP1*) from IGAP meta-analysis (Stage1)

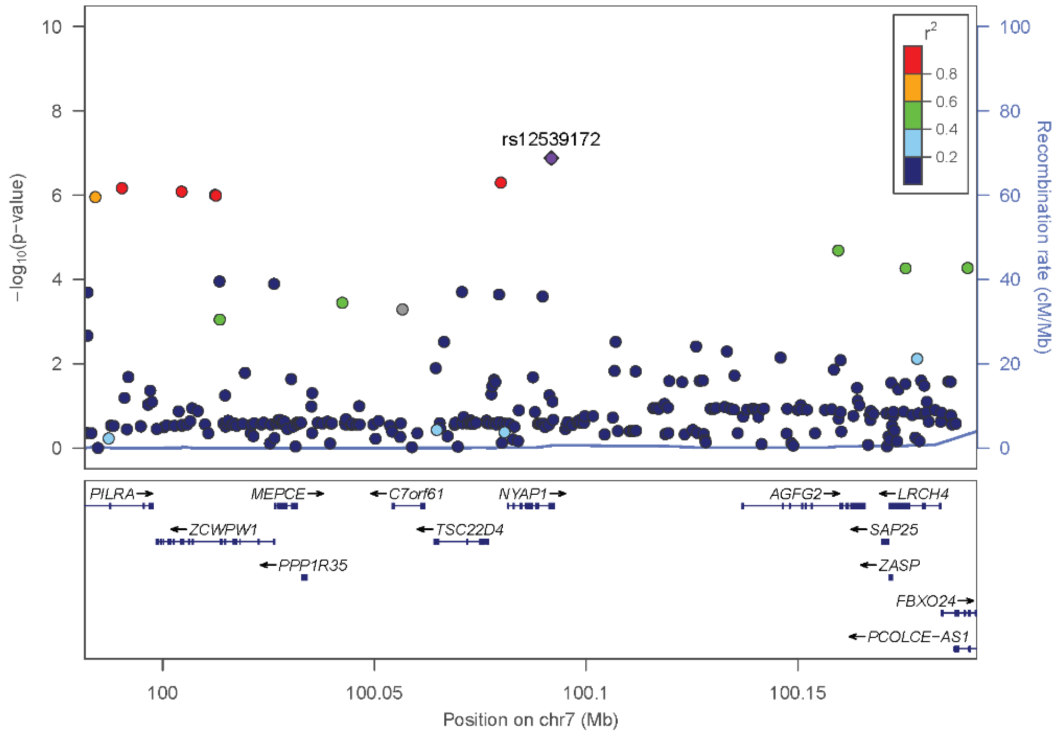

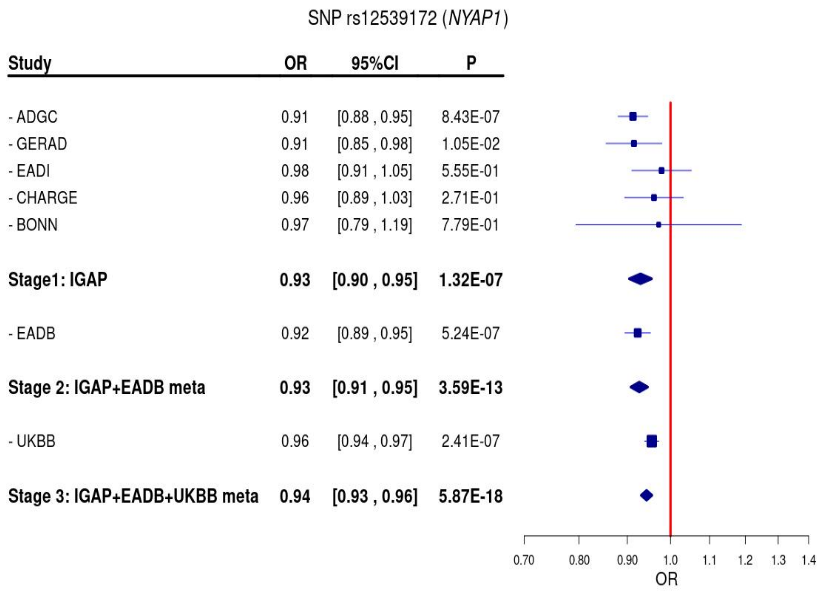

(r) LocusZoom and Forest plot of rs17462136 (*CASS4*) from IGAP meta-analysis (Stage1)

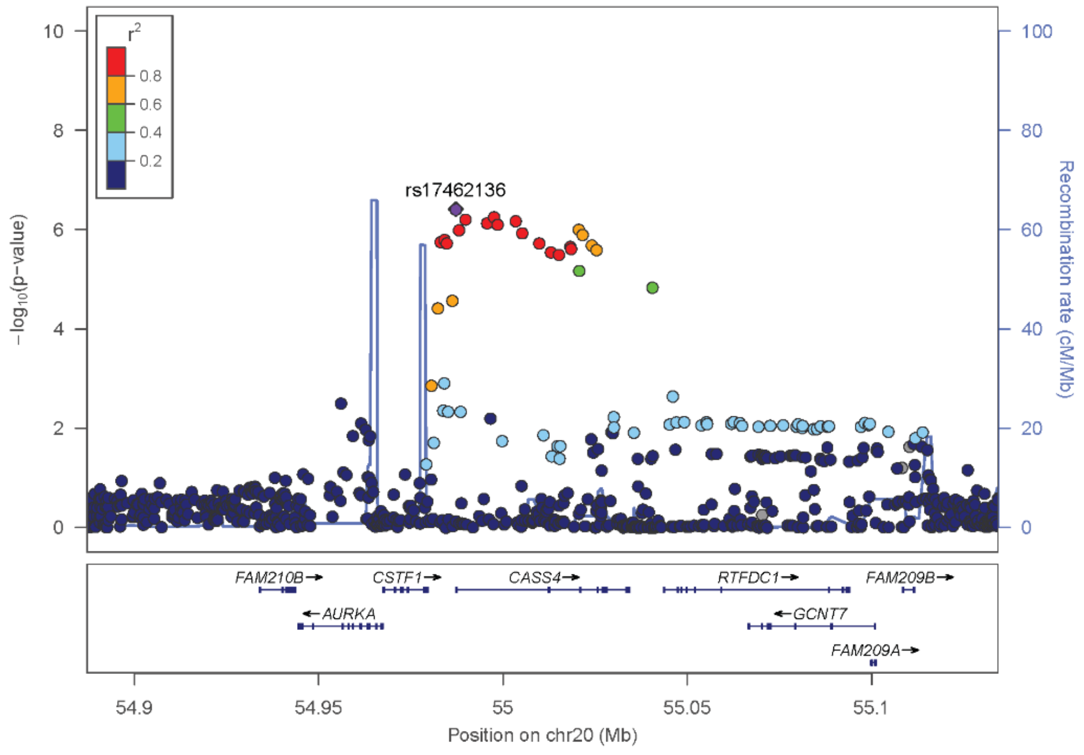

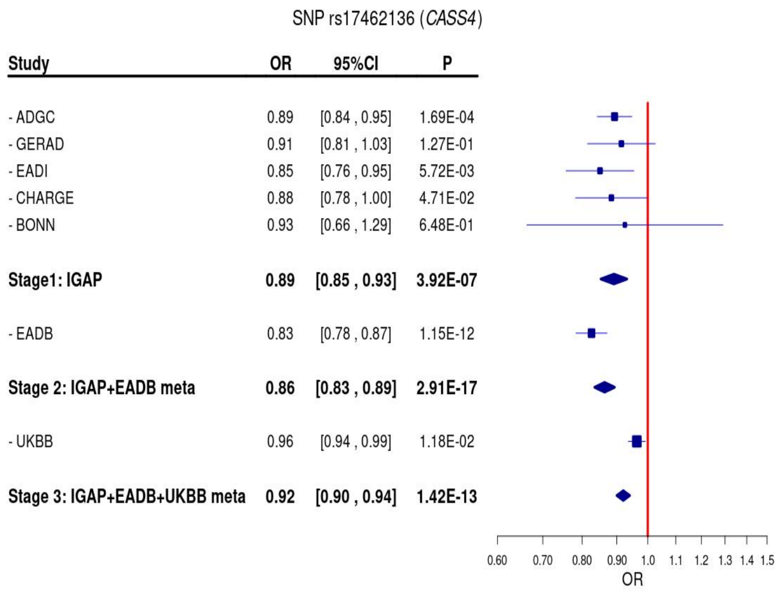

(s) LocusZoom and Forest plot of rs140016620 (*MTSS2/1L*) from IGAP meta-analysis (Stage1)

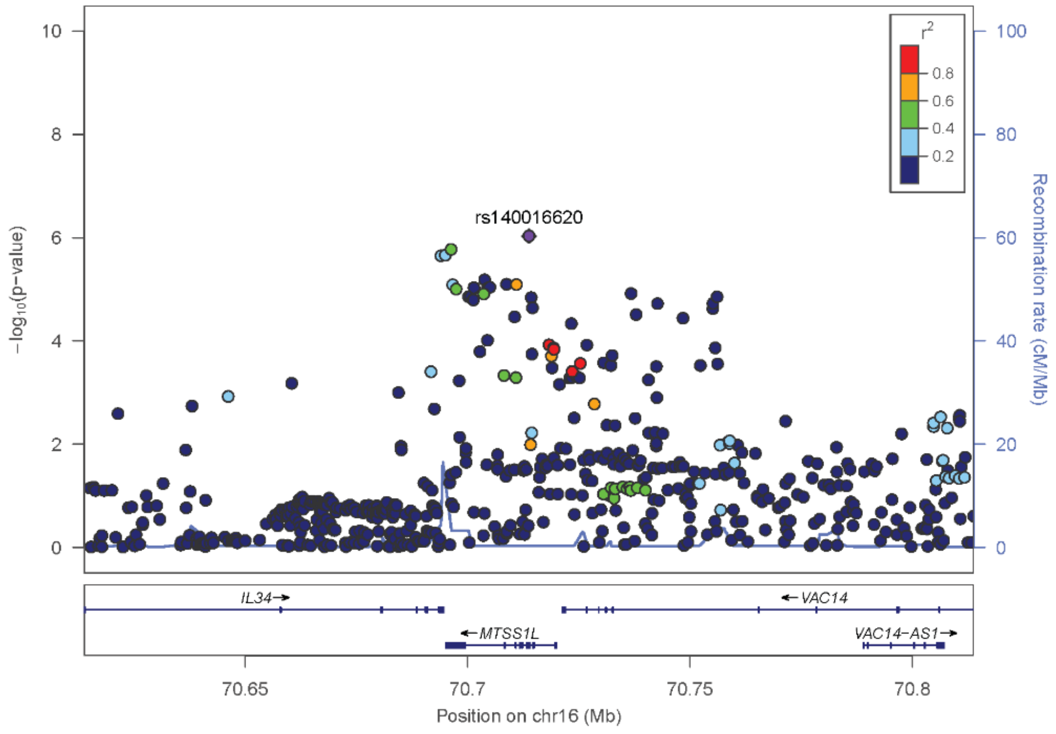

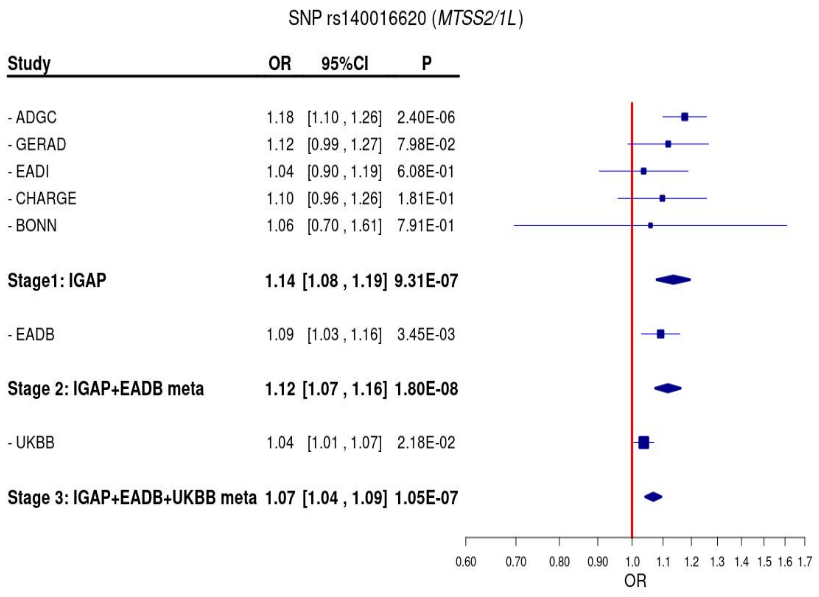

(t) LocusZoom and Forest plot of rs56402156 (*EPHA1*) from IGAP meta-analysis (Stage1)

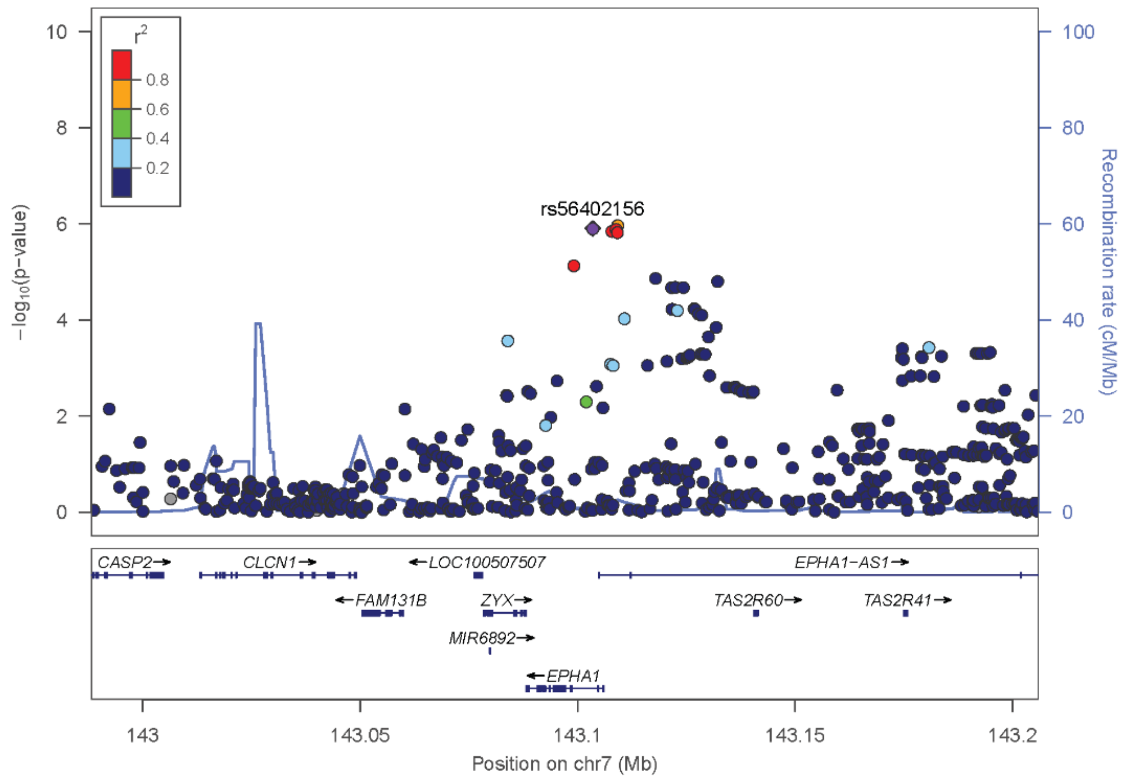

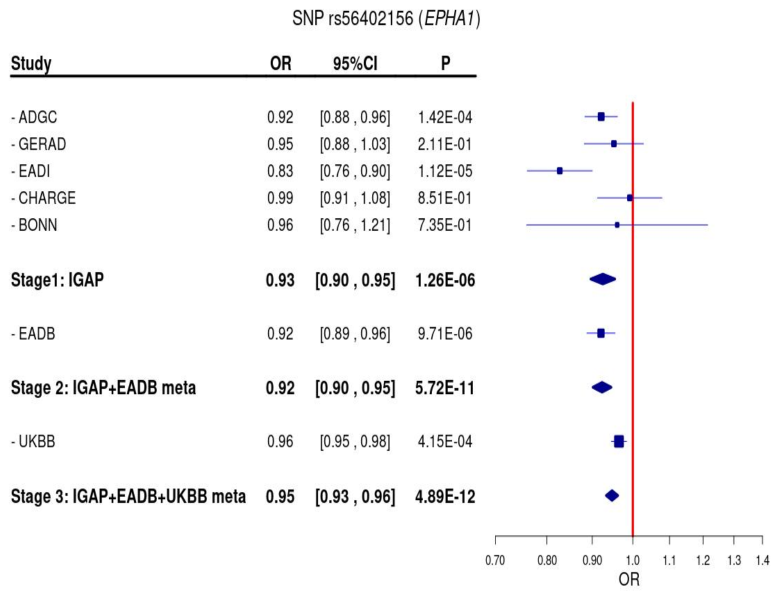

(u) LocusZoom and Forest plot of rs1001158 (*CLNK*) from IGAP meta-analysis (Stage1)

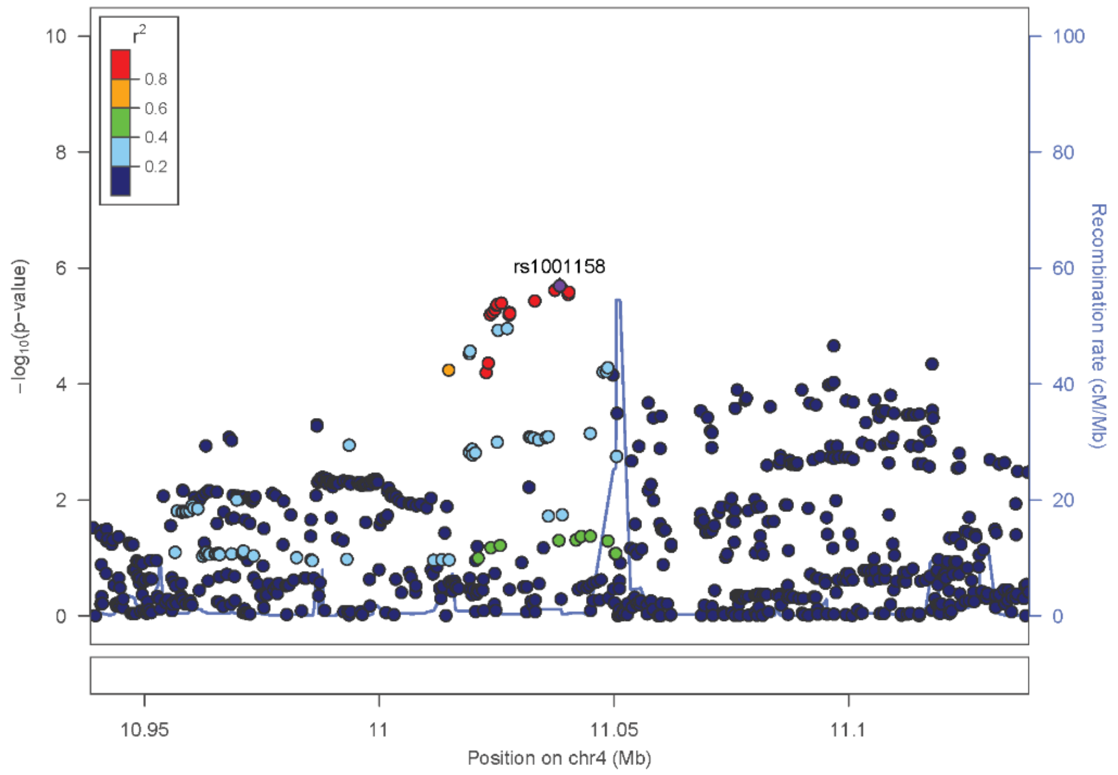

SNP rs1001158 (*CLNK*)

| Study | OR | 95%CI | P |
|---|---|---|---|
| - ADGC | 1.08 | [1.04 , 1.12] | 1.04E-04 |
| - GERAD | 1.12 | [1.04 , 1.20] | 2.45E-03 |
| - EADI | 1.07 | [0.99 , 1.15] | 8.42E-02 |
| - CHARGE | 1.03 | [0.96 , 1.11] | 3.85E-01 |
| - BONN | 1.18 | [0.96 , 1.46] | 1.15E-01 |
| **Stage1: IGAP** | **1.07** | **[1.04 , 1.10]** | **2.03E-06** |
| - EADB | 1.07 | [1.04 , 1.10] | 2.63E-05 |
| **Stage 2: IGAP+EADB meta** | **1.07** | **[1.05 , 1.09]** | **2.24E-10** |
| - UKBB | 1.04 | [1.02 , 1.06] | 2.50E-05 |
| **Stage 3: IGAP+EADB+UKBB meta** | **1.05** | **[1.04 , 1.07]** | **2.05E-13** |

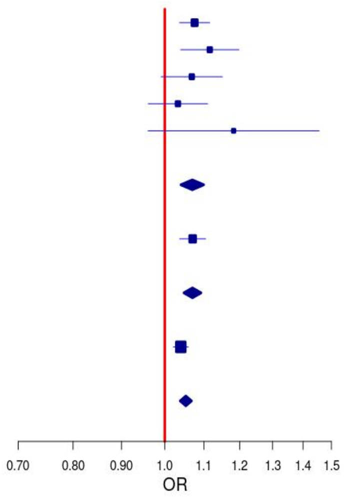

(v) LocusZoom and Forest plot of rs6966331 (*NME8*) from IGAP meta-analysis (Stage1)

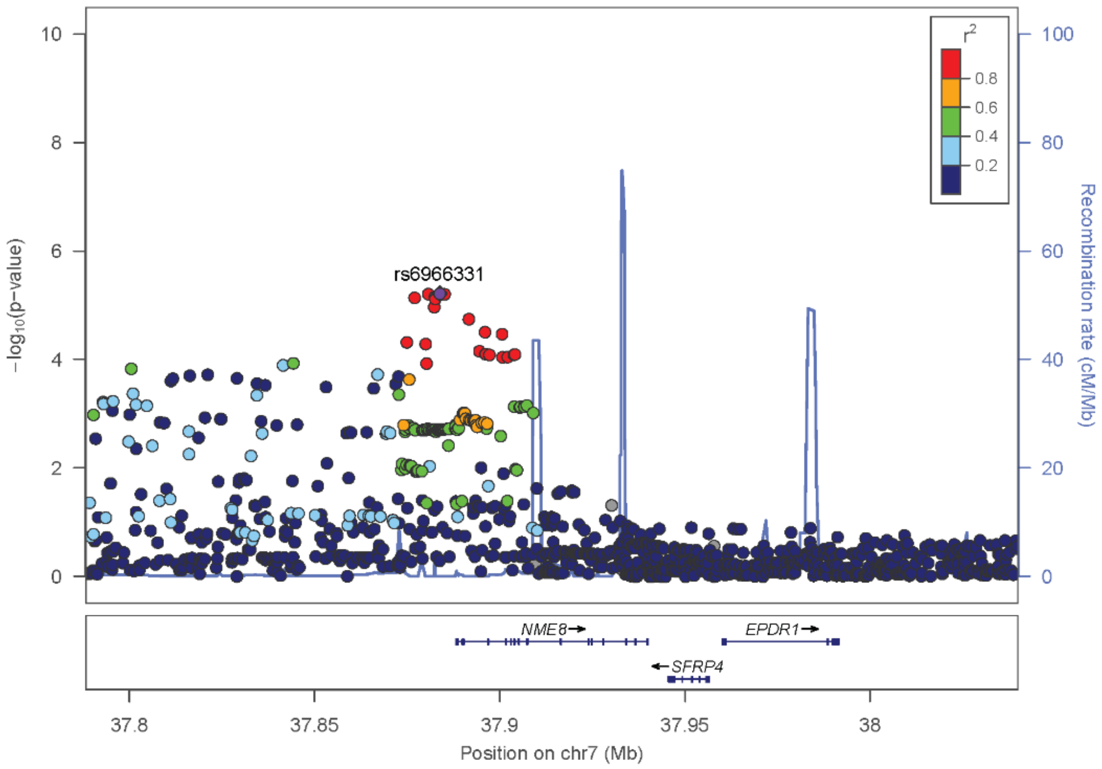

SNP rs6966331 (*NME8*)

| Study | OR | 95%CI | P |
|---|---|---|---|
| - ADGC | 0.93 | [0.89 , 0.96] | 9.39E-06 |
| - GERAD | 0.99 | [0.93 , 1.06] | 8.33E-01 |
| - EADI | 0.92 | [0.86 , 0.99] | 2.59E-02 |
| - CHARGE | 0.96 | [0.89 , 1.02] | 1.87E-01 |
| - BONN | 0.86 | [0.71 , 1.04] | 1.18E-01 |
| **Stage1: IGAP** | **0.94** | **[0.92 , 0.97]** | **6.09E-06** |
| - EADB | 0.94 | [0.91 , 0.97] | 2.88E-05 |
| **Stage 2: IGAP+EADB meta** | **0.94** | **[0.92 , 0.96]** | **7.14E-10** |
| - UKBB | 0.98 | [0.97 , 1.00] | 6.45E-02 |
| **Stage 3: IGAP+EADB+UKBB meta** | **0.96** | **[0.95 , 0.98]** | **4.81E-08** |

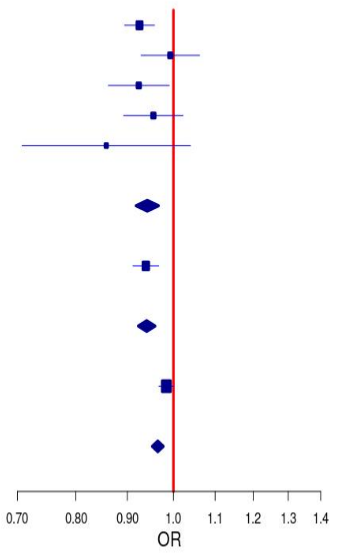

**Supplementary Figure 5.** Individual (a) regional association plots from LocusZoom and (b) Forest plots of Stage 1-3 associations for most strongly associated variants in each rare variant (MAF≤0.01) locus.

(a) LocusZoom and Forest plot of rs547509922 (*APOE* Region) from IGAP meta-analysis (Stage1)

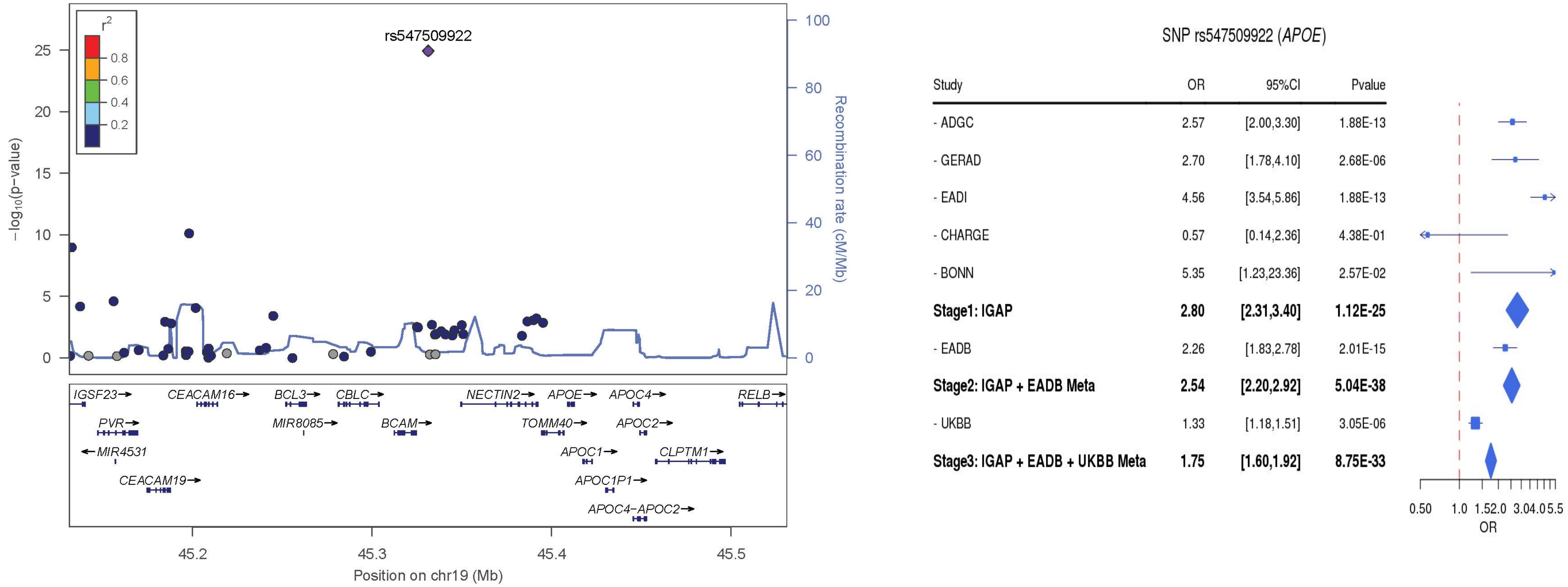

Abbreviations: OR = odds ratio, CI = confidence interval

(b) LocusZoom and Forest plot of rs559118614 (*APOE* Region/*CLASRP*) from IGAP meta-analysis (Stage1)

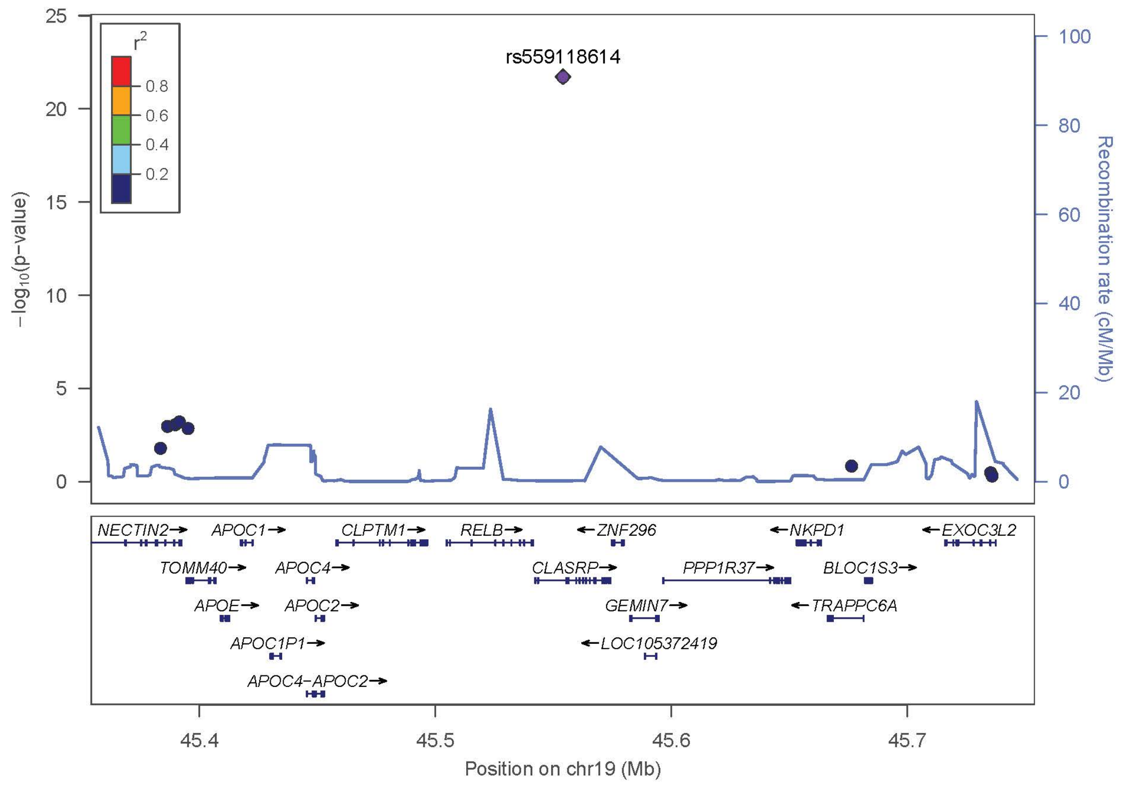

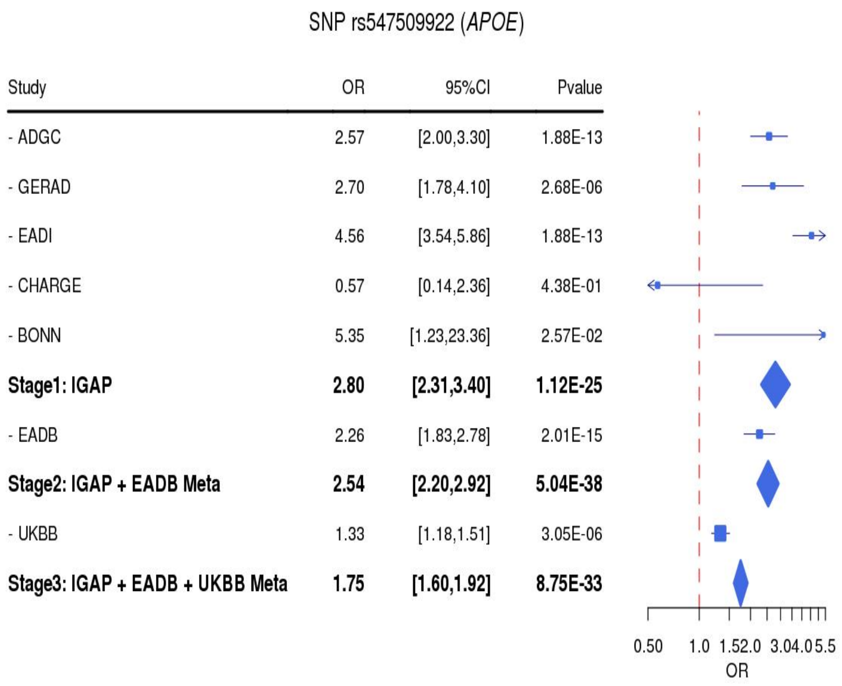

(c) LocusZoom and Forest plot of rs75932628 (*TREM2*) from IGAP meta-analysis (Stage1)

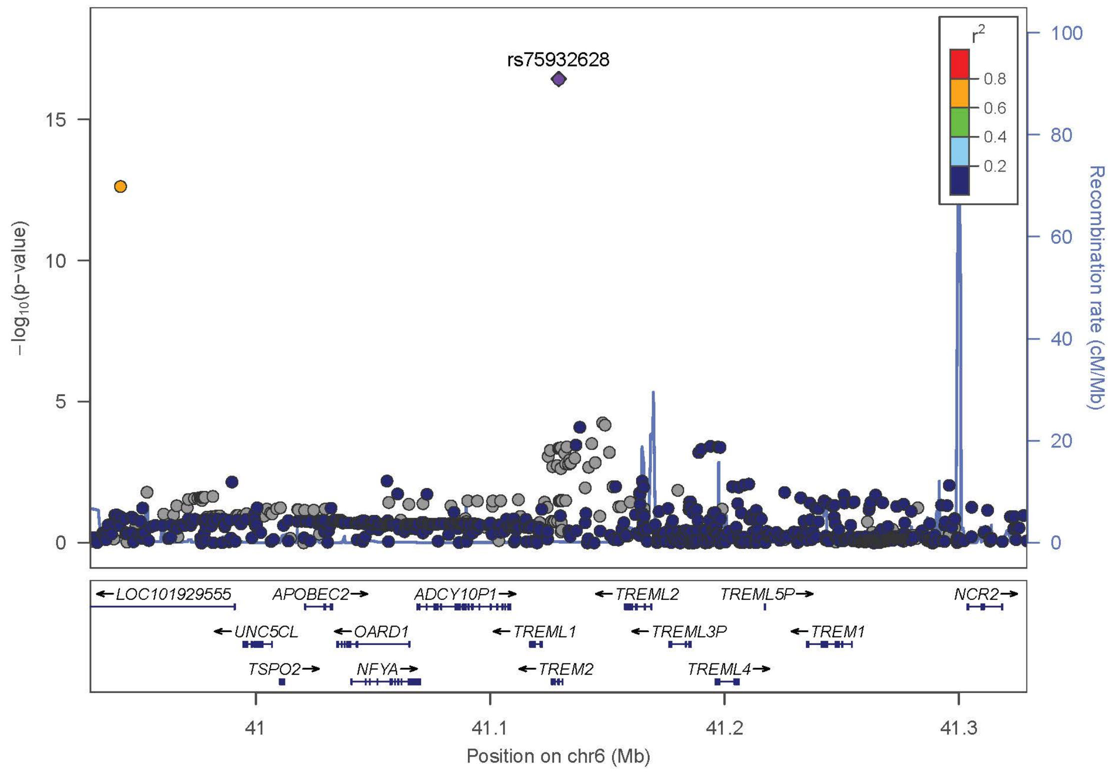

SNP rs75932628 (*TREM2*)

| Study | OR | 95%CI | Pvalue |
|---|---|---|---|
| - ADGC | 2.29 | [1.78,2.94] | 1.00E-10 |
| - GERAD | 2.50 | [1.48,4.21] | 6.09E-04 |
| - EADI | 3.96 | [3.08,5.09] | 1.00E-10 |
| - CHARGE | 1.80 | [0.60,5.38] | 2.91E-01 |
| - BONN | 4.24 | [0.47,38.32] | 1.98E-01 |
| **Stage1: IGAP** | **2.45** | **[1.99,3.02]** | **3.72E-17** |
| - EADB | 2.14 | [1.58,2.89] | 2.97E-07 |
| **Stage2: IGAP + EADB Meta** | **2.34** | **[1.97,2.78]** | **2.15E-22** |
| - UKBB | 1.45 | [1.25,1.69] | 9.00E-07 |
| **Stage3: IGAP + EADB + UKBB Meta** | **1.79** | **[1.60,2.00]** | **5.59E-24** |

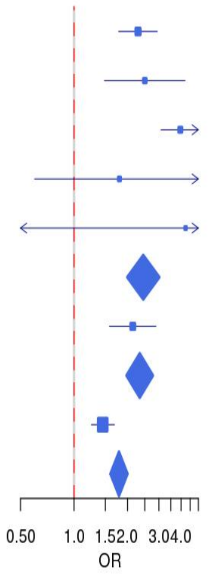

(d) LocusZoom and Forest plot of rs187370608 (*TREM2*) from IGAP meta-analysis (Stage1)

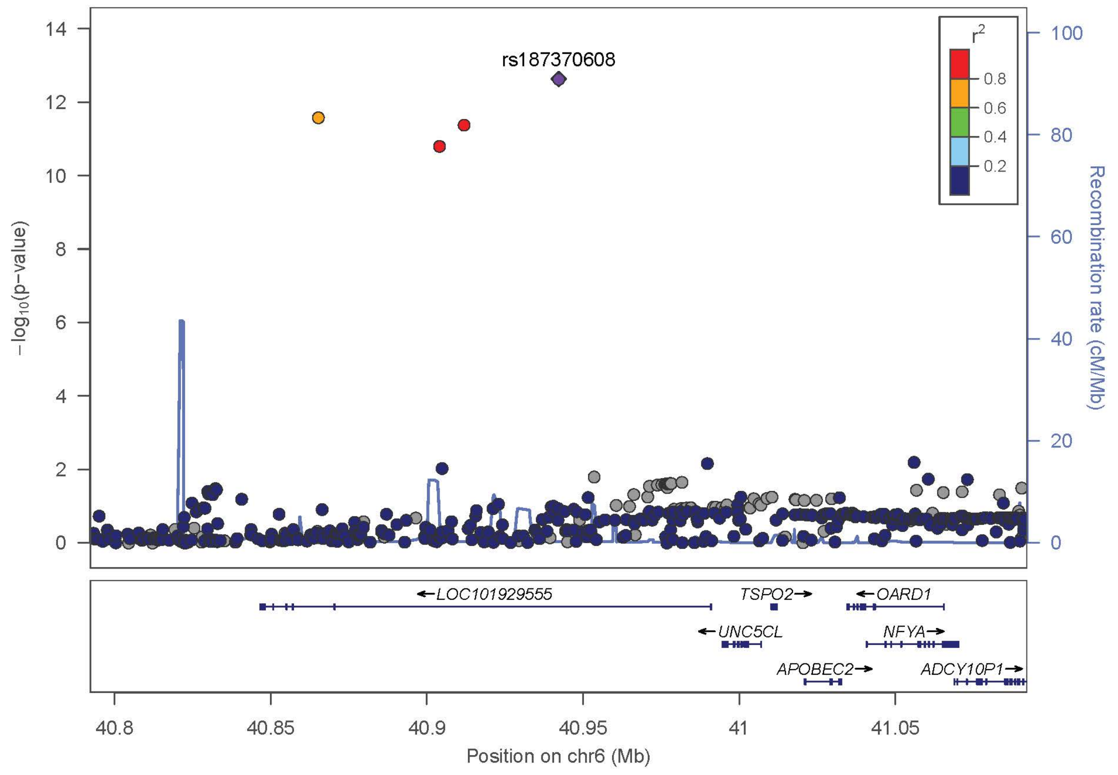

SNP rs187370608 (*TREM2*)

| Study | OR | 95%CI | Pvalue |
|---|---|---|---|
| - ADGC | 2.01 | [1.56,2.59] | 6.74E-08 |
| - GERAD | 2.24 | [1.35,3.74] | 1.89E-03 |
| - EADI | 3.53 | [2.74,4.55] | 6.74E-08 |
| - CHARGE | 1.77 | [0.57,5.50] | 3.21E-01 |
| - BONN | 9.05 | [0.84,97.10] | 6.88E-02 |
| **Stage1: IGAP** | **2.19** | **[1.78,2.70]** | **2.34E-13** |
| - EADB | 2.10 | [1.57,2.81] | 2.77E-07 |
| **Stage2: IGAP + EADB Meta** | **2.16** | **[1.82,2.56]** | **8.70E-19** |
| - UKBB | 1.45 | [1.26,1.67] | 1.65E-07 |
| **Stage3: IGAP + EADB + UKBB Meta** | **1.70** | **[1.53,1.90]** | **4.57E-22** |

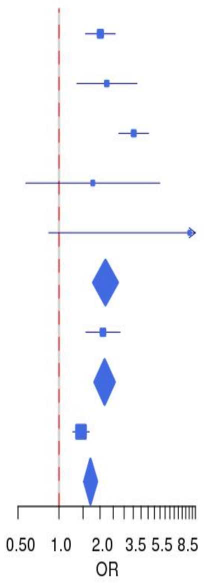

(e) LocusZoom and Forest plot of rs143202663 (*TREM2*) from IGAP meta-analysis (Stage1)

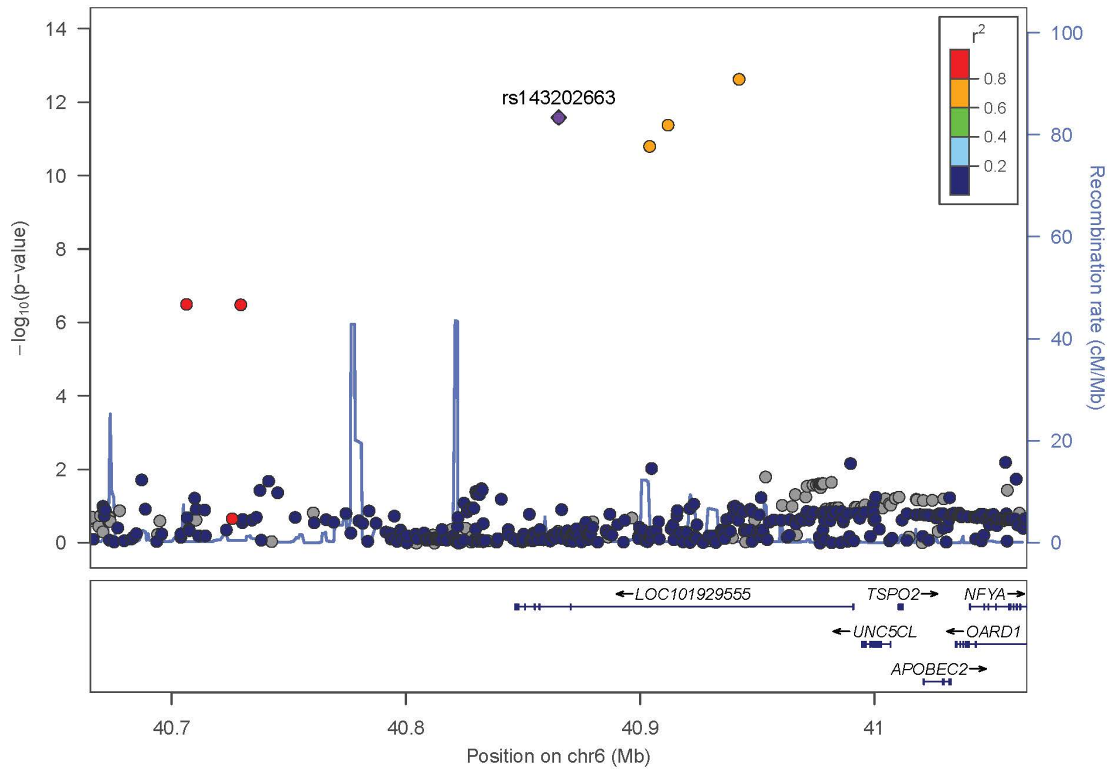

SNP rs143202663 (*TREM2*)

| Study | OR | 95%CI | Pvalue |
|---|---|---|---|
| - ADGC | 1.90 | [1.48,2.44] | 4.67E-07 |
| - GERAD | 2.12 | [1.24,3.63] | 6.35E-03 |
| - EADI | 3.94 | [3.07,5.06] | 4.67E-07 |
| - CHARGE | 2.40 | [0.90,6.41] | 8.13E-02 |
| - BONN | 9.38 | [0.78,112.26] | 7.72E-02 |
| **Stage1: IGAP** | **2.11** | **[1.71,2.61]** | **2.59E-12** |
| - EADB | 2.18 | [1.63,2.90] | 4.41E-08 |
| **Stage2: IGAP + EADB Meta** | **0.47** | **[0.40,0.55]** | **1.83E-18** |
| - UKBB | 1.34 | [1.15,1.55] | 1.19E-04 |
| **Stage3: IGAP + EADB + UKBB Meta** | **0.61** | **[0.55,0.68]** | **4.54E-18** |

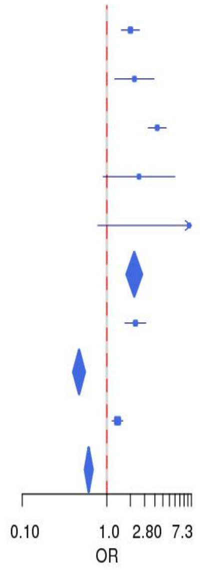

(f) LocusZoom and Forest plot of rs145342536 (*TREM2*) from IGAP meta-analysis (Stage1)

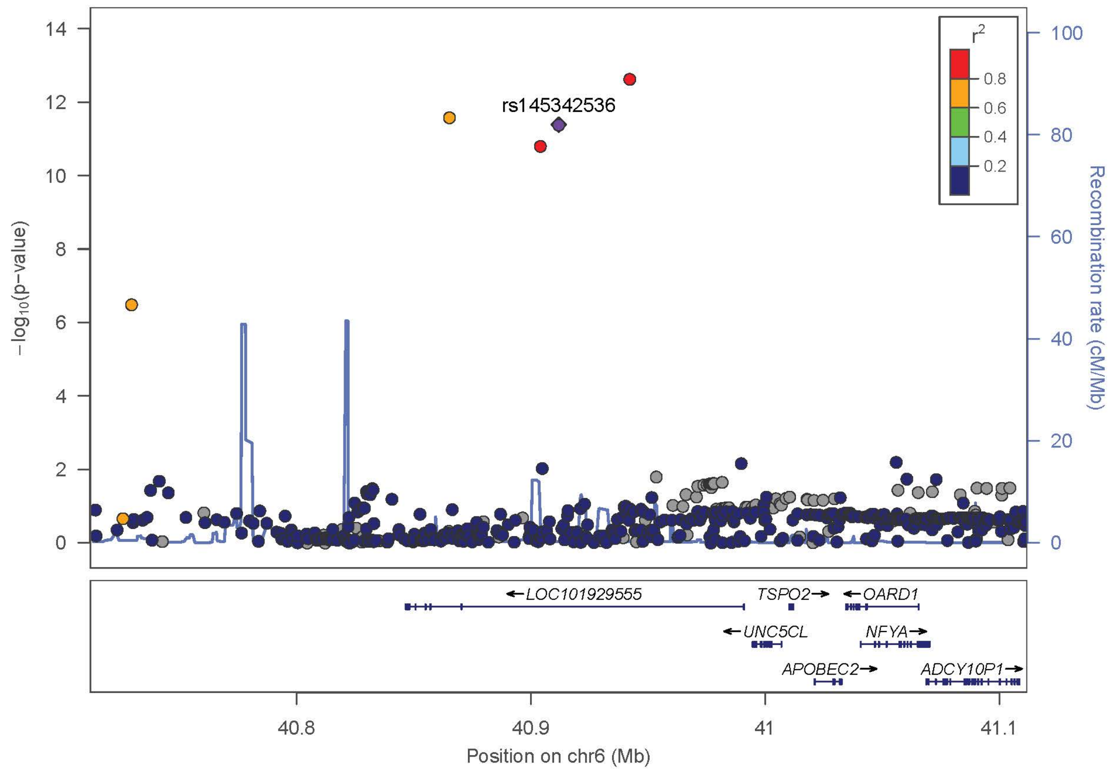

SNP rs145342536 (*TREM2*)

| Study | OR | 95%CI | Pvalue |
|---|---|---|---|
| - ADGC | 1.80 | [1.43,2.27] | 5.88E-07 |
| - GERAD | 2.03 | [1.24,3.32] | 4.59E-03 |
| - EADI | 3.70 | [2.93,4.66] | 5.88E-07 |
| - CHARGE | 1.86 | [0.68,5.05] | 2.24E-01 |
| - BONN | 7.28 | [0.74,71.54] | 8.87E-02 |
| **Stage1: IGAP** | **1.99** | **[1.64,2.41]** | **4.13E-12** |
| - EADB | 2.07 | [1.56,2.75] | 2.14E-07 |
| **Stage2: IGAP + EADB Meta** | **2.01** | **[1.71,2.36]** | **1.14E-17** |
| - UKBB | 1.37 | [1.20,1.57] | 5.37E-06 |
| **Stage3: IGAP + EADB + UKBB Meta** | **1.61** | **[1.45,1.78]** | **2.39E-19** |

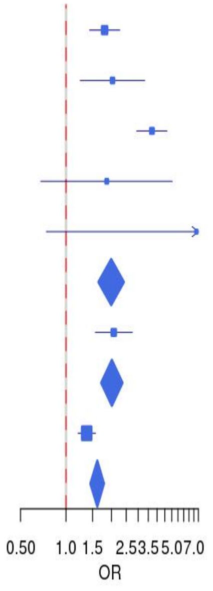

(g) LocusZoom and Forest plot of rs12664332 (*TREM2*) from IGAP meta-analysis (Stage1)

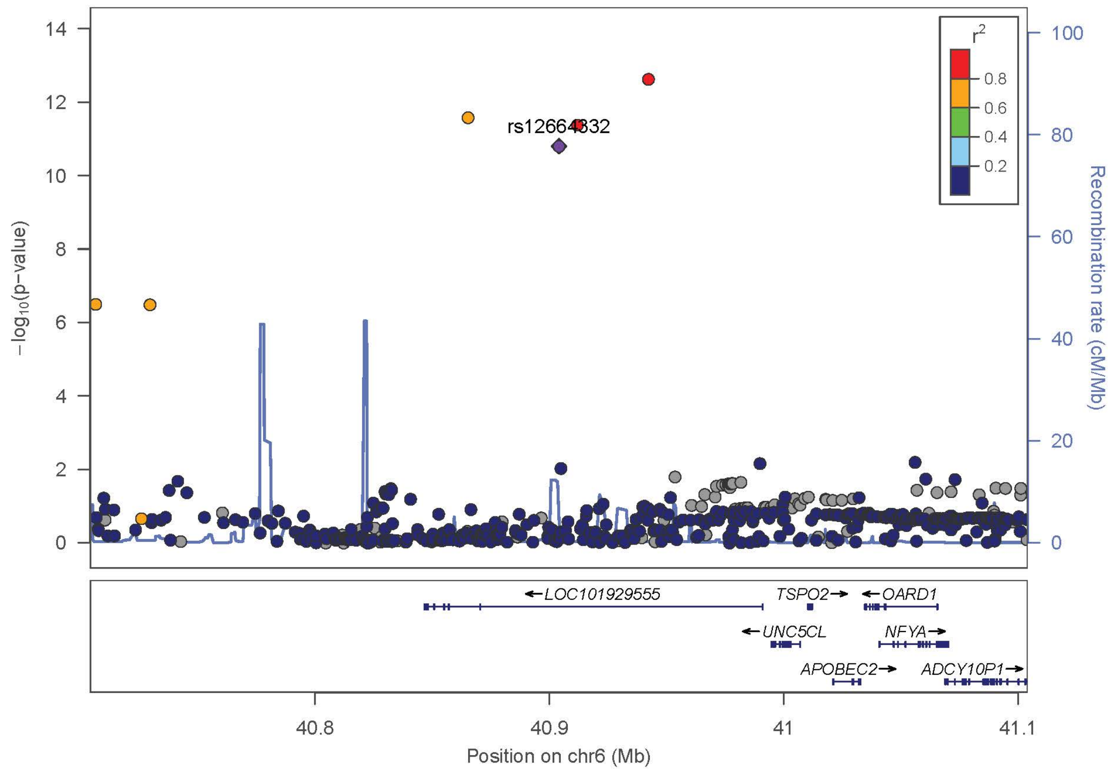

SNP rs12664332 (*TREM2*)

| Study | OR | 95%CI | Pvalue |
|---|---|---|---|
| - ADGC | 1.79 | [1.42,2.25] | 8.55E-07 |
| - GERAD | 1.86 | [1.14,3.05] | 1.37E-02 |
| - EADI | 3.71 | [2.95,4.68] | 8.55E-07 |
| - CHARGE | 1.88 | [0.7,5.05] | 2.10E-01 |
| - BONN | 7.45 | [0.77,71.98] | 8.25E-02 |
| **Stage1: IGAP** | **1.95** | **[1.61,2.37]** | **1.57E-11** |
| - EADB | 2.10 | [1.58,2.79] | 1.31E-07 |
| **Stage2: IGAP + EADB Meta** | **2.00** | **[1.7,2.34]** | **2.98E-17** |
| - UKBB | 1.35 | [1.18,1.55] | 1.25E-05 |
| **Stage3: IGAP + EADB + UKBB Meta** | **1.59** | **[1.44,1.77]** | **1.43E-18** |

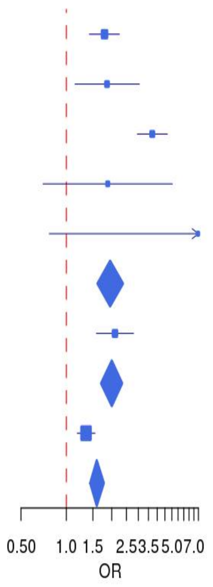

(h) LocusZoom and Forest plot of rs112952132 (*APOE* Region) from IGAP meta-analysis (Stage1)

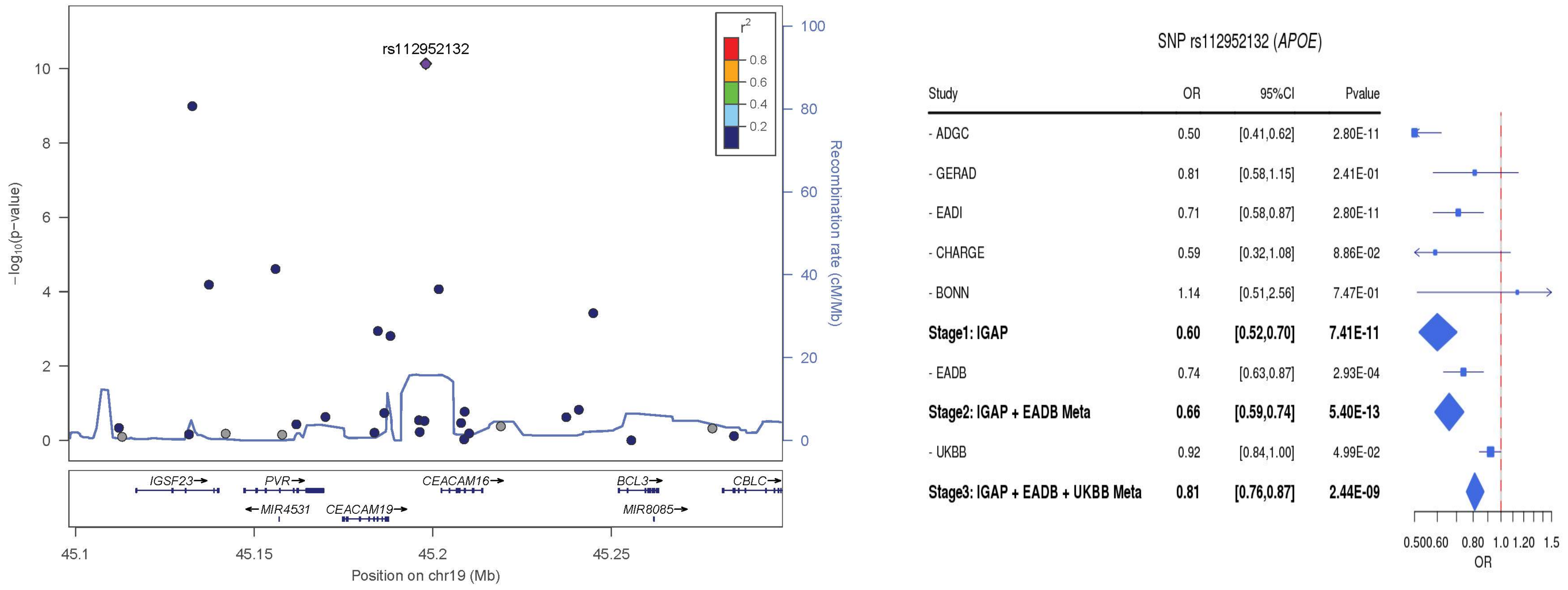

(i) LocusZoom and Forest plot of rs78905796 (*APOE* Region/*IGSF23*) from IGAP meta-analysis (Stage1)

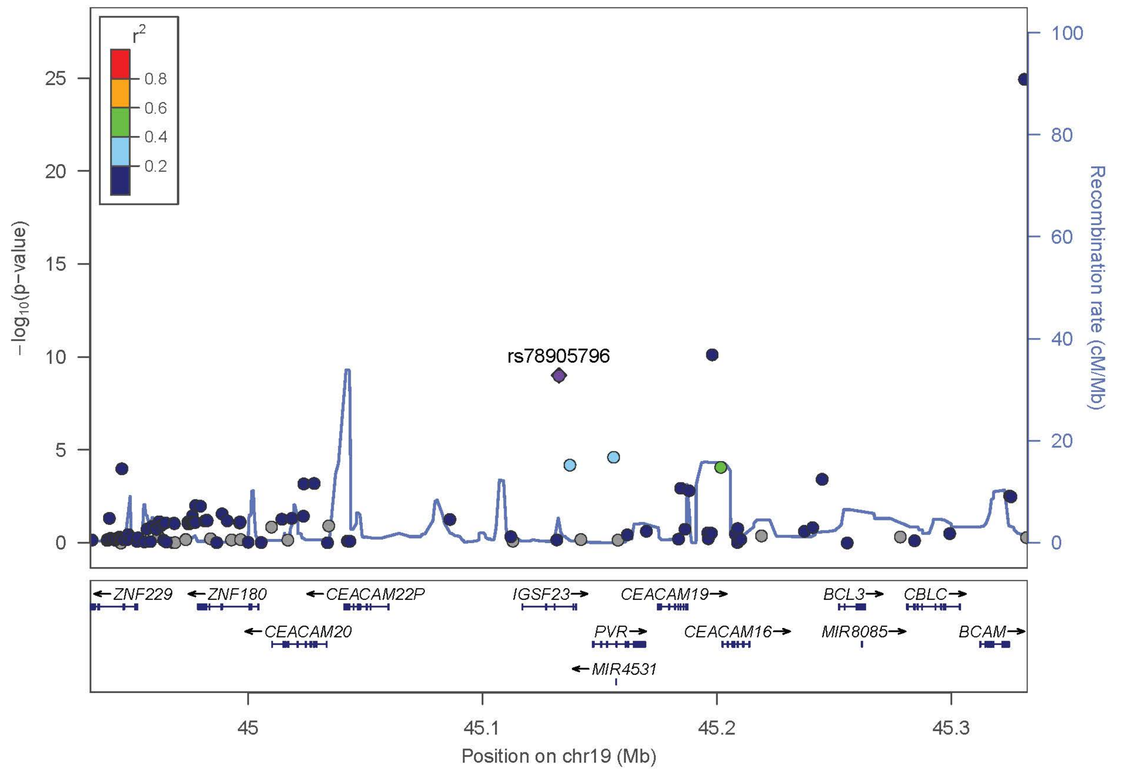

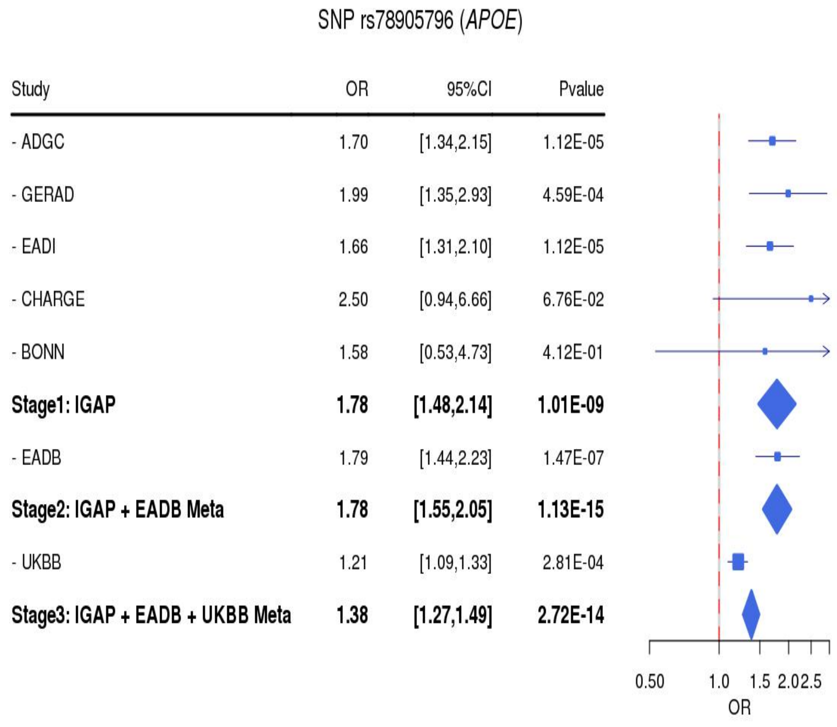

(j) LocusZoom and Forest plot of rs540038005 (*RORA*) from IGAP meta-analysis (Stage1)

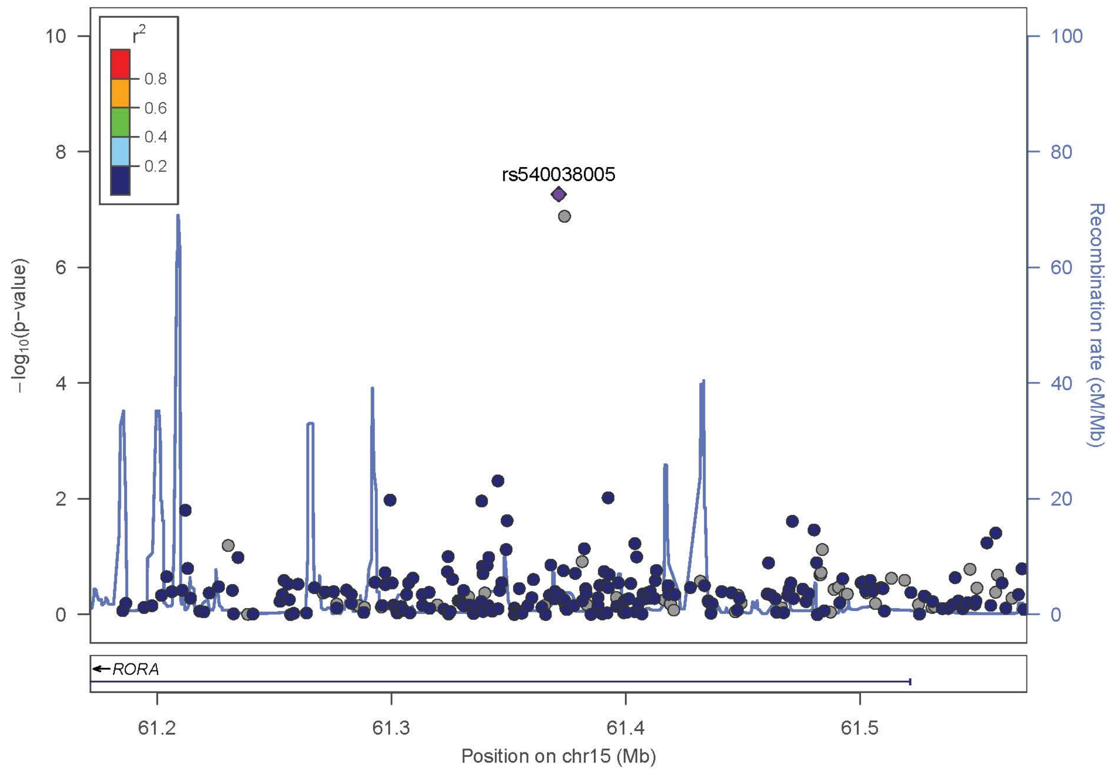

SNP rs540038005 (*RORA*)

| Study | OR | 95%CI | Pvalue |
|---|---|---|---|
| - ADGC | 3.09 | [1.73,5.52] | 1.43E-04 |
| - GERAD | 6.32 | [2.16,18.49] | 7.52E-04 |
| - EADI | 1.33 | [0.74,2.37] | 1.43E-04 |
| - CHARGE | 11.50 | [1.67,79.32] | 1.32E-02 |
| - BONN | 6.09 | [0.50,74.07] | 1.56E-01 |
| **Stage1: IGAP** | **3.60** | **[2.27,5.71]** | **5.40E-08** |
| - EADB | 0.80 | [0.49,1.31] | 3.70E-01 |
| **Stage2: IGAP + EADB Meta** | **1.79** | **[1.28,2.51]** | **7.25E-04** |
| - UKBB | NA | [NA,NA] | NA |
| **Stage3: IGAP + EADB + UKBB Meta** | **1.79** | **[1.28,2.51]** | **7.25E-04** |

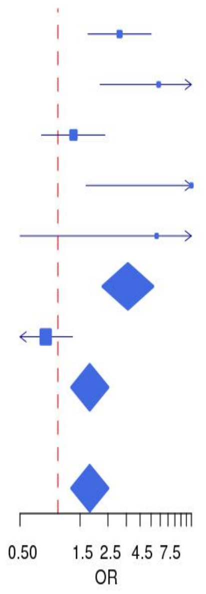

(k) LocusZoom and Forest plot of rs551336410 (*RORA*) from IGAP meta-analysis (Stage1)

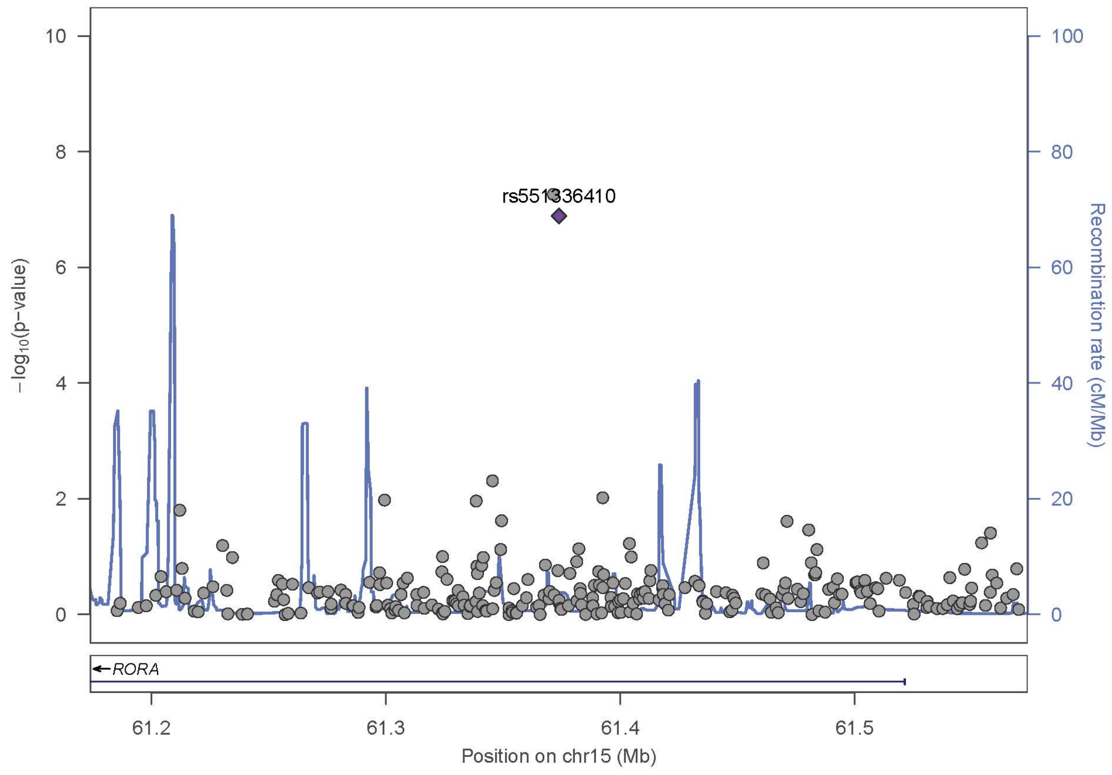

SNP rs551336410 (*RORA*)

| Study | OR | 95%CI | Pvalue |
|---|---|---|---|
| - ADGC | 3.14 | [1.78,5.57] | 8.59E-05 |
| - GERAD | 3.61 | [1.35,9.66] | 1.07E-02 |
| - EADI | 1.58 | [0.89,2.79] | 8.59E-05 |
| - CHARGE | 13.44 | [1.91,94.41] | 9.01E-03 |
| - BONN | 6.07 | [0.50,73.84] | 1.57E-01 |
| **Stage1: IGAP** | **3.34** | **[2.14,5.23]** | **1.29E-07** |
| - EADB | 0.77 | [0.47,1.26] | 2.97E-01 |
| **Stage2: IGAP + EADB Meta** | **1.71** | **[1.23,2.38]** | **1.41E-03** |
| - UKBB | NA | [NA,NA] | NA |
| **Stage3: IGAP + EADB + UKBB Meta** | **1.71** | **[1.23,2.38]** | **1.41E-03** |

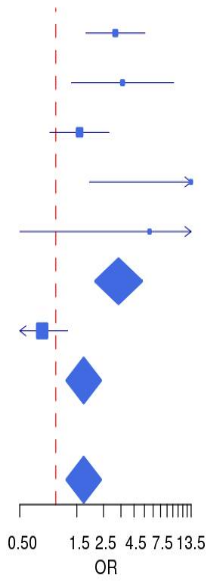

(l) LocusZoom and Forest plot of rs192675224 (*TREM2*) from IGAP meta-analysis (Stage1)

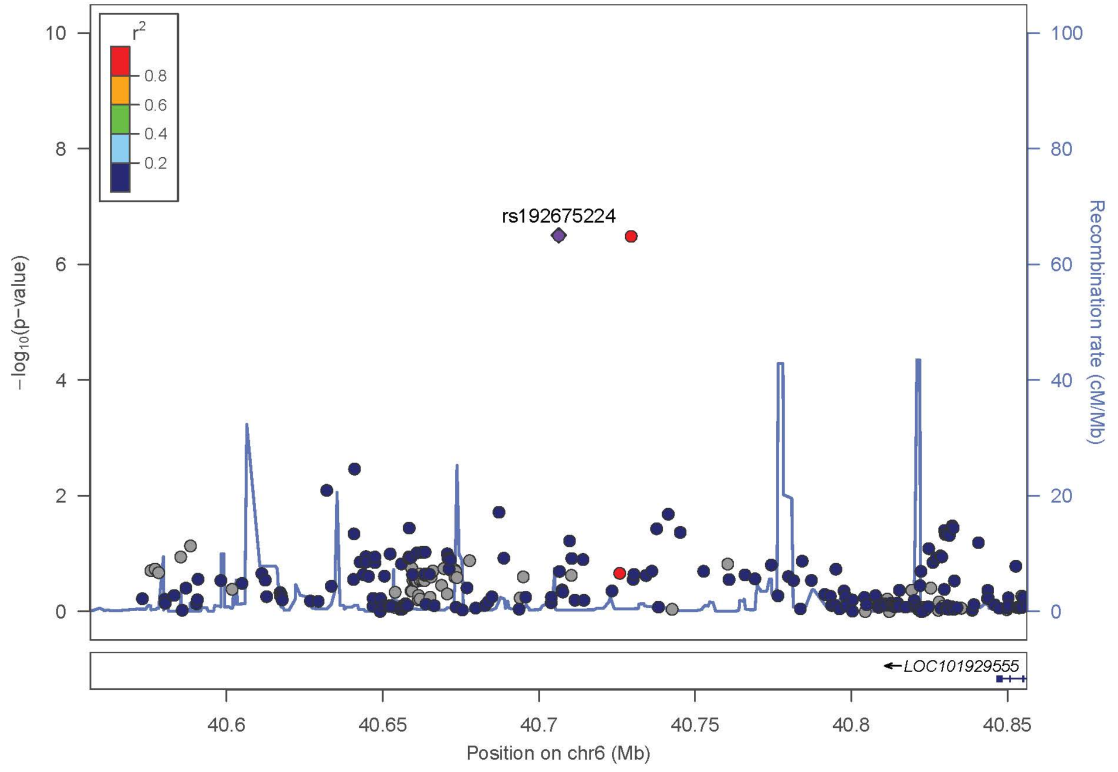

SNP rs192675224 (*TREM2*)

| Study | OR | 95%CI | Pvalue |
|---|---|---|---|
| - ADGC | 1.73 | [1.31,2.28] | 1.19E-04 |
| - GERAD | 1.37 | [0.71,2.65] | 3.53E-01 |
| - EADI | 4.50 | [3.41,5.94] | 1.19E-04 |
| - CHARGE | 2.34 | [0.94,5.86] | 6.90E-02 |
| - BONN | 8.01 | [0.53,120.67] | 1.33E-01 |
| **Stage1: IGAP** | **1.85** | **[1.46,2.35]** | **3.15E-07** |
| - EADB | 1.70 | [1.26,2.29] | 4.65E-04 |
| **Stage2: IGAP + EADB Meta** | **1.79** | **[1.49,2.16]** | **7.57E-10** |
| - UKBB | 1.22 | [1.02,1.45] | 2.71E-02 |
| **Stage3: IGAP + EADB + UKBB Meta** | **1.46** | **[1.29,1.66]** | **5.71E-09** |

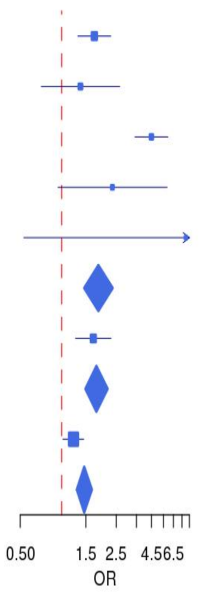

(m)LocusZoom and Forest plot of rs150085726 (*TREM2*) from IGAP meta-analysis (Stage1)

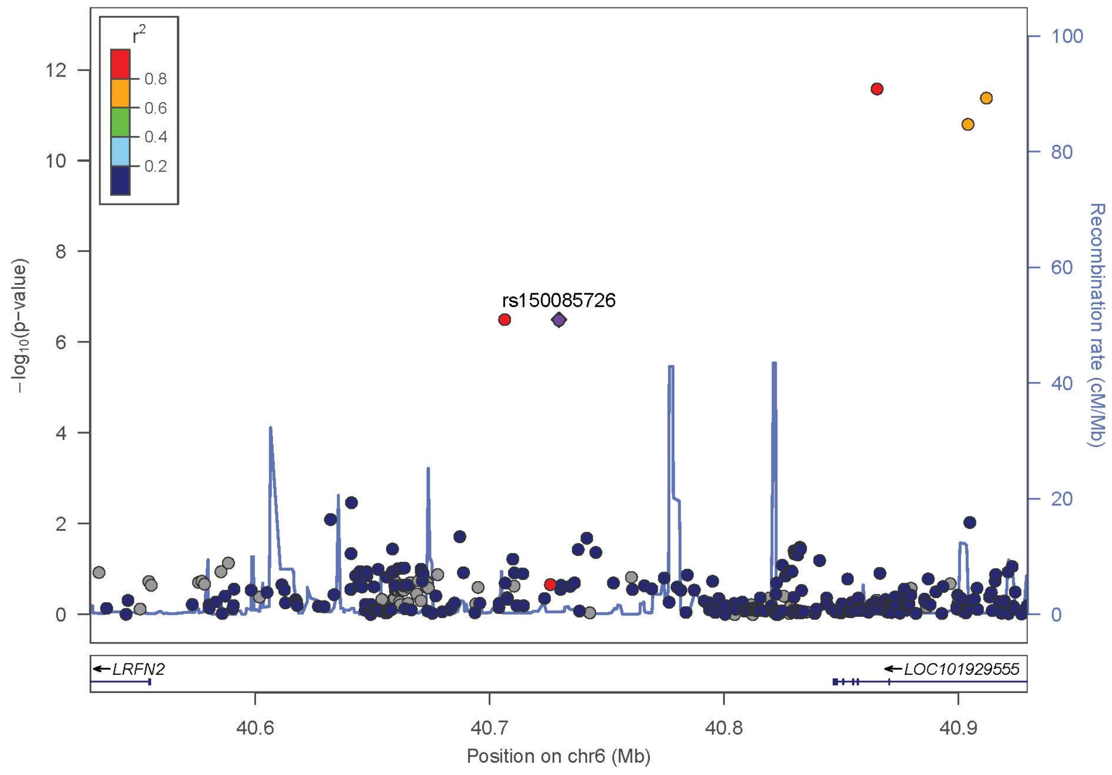

SNP rs150085726 (*TREM2*)

| Study | OR | 95%CI | Pvalue |
|---|---|---|---|
| - ADGC | 1.75 | [1.32,2.34] | 1.19E-04 |
| - GERAD | 1.29 | [0.71,2.36] | 4.00E-01 |
| - EADI | 3.71 | [2.79,4.94] | 1.19E-04 |
| - CHARGE | 3.33 | [1.16,9.56] | 2.51E-02 |
| - BONN | 7.03 | [0.52,95.19] | 1.42E-01 |
| **Stage1: IGAP** | **1.86** | **[1.47,2.37]** | **3.24E-07** |
| - EADB | 1.76 | [1.29,2.40] | 2.91E-04 |
| **Stage2: IGAP + EADB Meta** | **1.82** | **[1.51,2.20]** | **4.84E-10** |
| - UKBB | 1.27 | [1.08,1.48] | 3.48E-03 |
| **Stage3: IGAP + EADB + UKBB Meta** | **1.47** | **[1.30,1.66]** | **4.68E-10** |

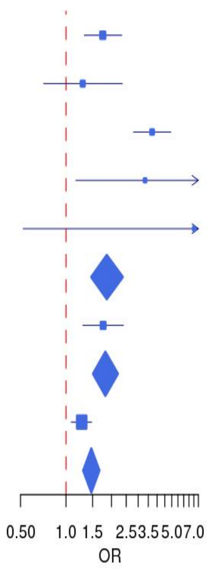

(n) LocusZoom and Forest plot of rs143080277 (*NCK2*) from IGAP meta-analysis (Stage1)

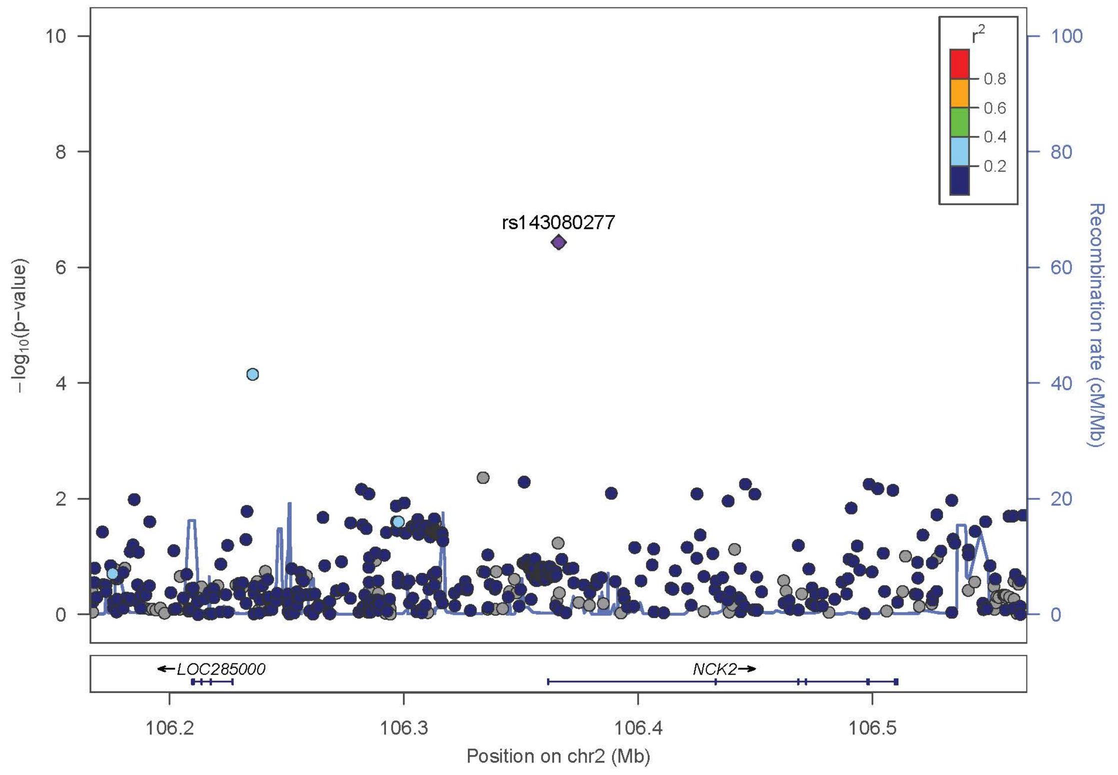

SNP rs143080277 (*NCK2*)

| Study | OR | 95%CI | Pvalue |
|---|---|---|---|
| - ADGC | 1.41 | [1.10,1.80] | 6.99E-03 |
| - GERAD | 2.22 | [1.44,3.42] | 3.33E-04 |
| - EADI | 2.36 | [1.84,3.02] | 6.99E-03 |
| - CHARGE | 1.50 | [0.68,3.30] | 3.11E-01 |
| - BONN | 1.98 | [0.41,9.49] | 3.95E-01 |
| **Stage1: IGAP** | **1.65** | **[1.36,2.01]** | **3.66E-07** |
| - EADB | 1.32 | [1.08,1.61] | 5.76E-03 |
| **Stage2: IGAP + EADB Meta** | **0.68** | **[0.59,0.78]** | **2.69E-08** |
| - UKBB | 1.34 | [1.18,1.52] | 3.22E-06 |
| **Stage3: IGAP + EADB + UKBB Meta** | **0.71** | **[0.65,0.78]** | **7.17E-13** |

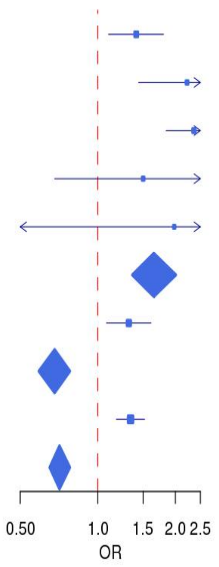

(o) LocusZoom and Forest plot of rs17035636 (*LINC02033/TRANK1*) from IGAP meta-analysis (Stage1)

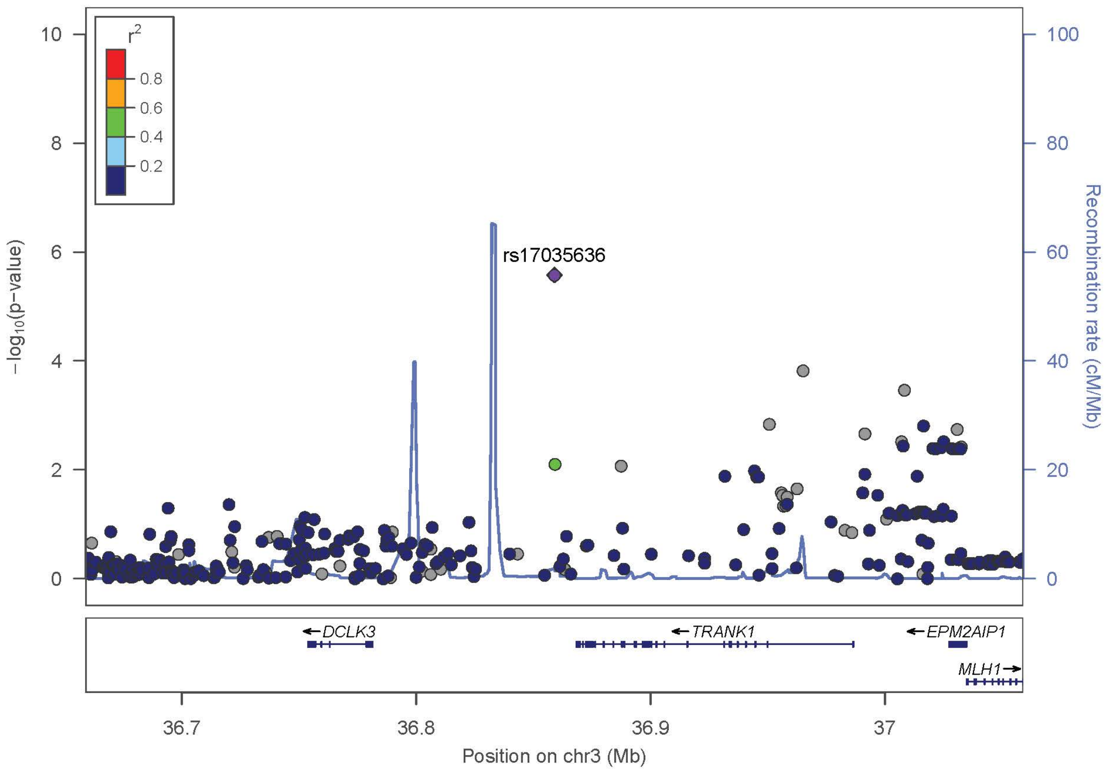

SNP rs17035636 (*LINC02033/TRANK1*)

| Study | OR | 95%CI | Pvalue |
|---|---|---|---|
| - ADGC | 0.36 | [0.23,0.57] | 1.12E-05 |
| - GERAD | 1.09 | [0.30,4.02] | 8.95E-01 |
| - EADI | 0.65 | [0.41,1.02] | 1.12E-05 |
| - CHARGE | 0.38 | [0.13,1.05] | 6.23E-02 |
| - BONN | 0.16 | [0.02,1.64] | 1.23E-01 |
| **Stage1: IGAP** | **0.43** | **[0.31,0.62]** | **2.64E-06** |
| - EADB | 0.85 | [0.62,1.16] | 3.02E-01 |
| **Stage2: IGAP + EADB Meta** | **1.60** | **[1.26,2.02]** | **8.62E-05** |
| - UKBB | NA | [NA,NA] | NA |
| **Stage3: IGAP + EADB + UKBB Meta** | **1.60** | **[1.26,2.02]** | **8.62E-05** |

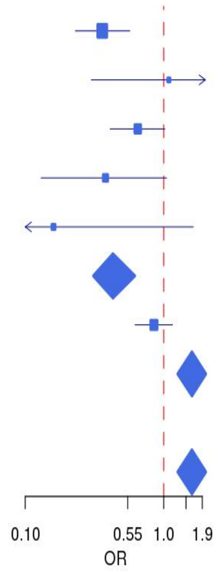

(p) LocusZoom and Forest plot of rs182464045 (*DYTN/MDH1B*) from IGAP meta-analysis (Stage1)

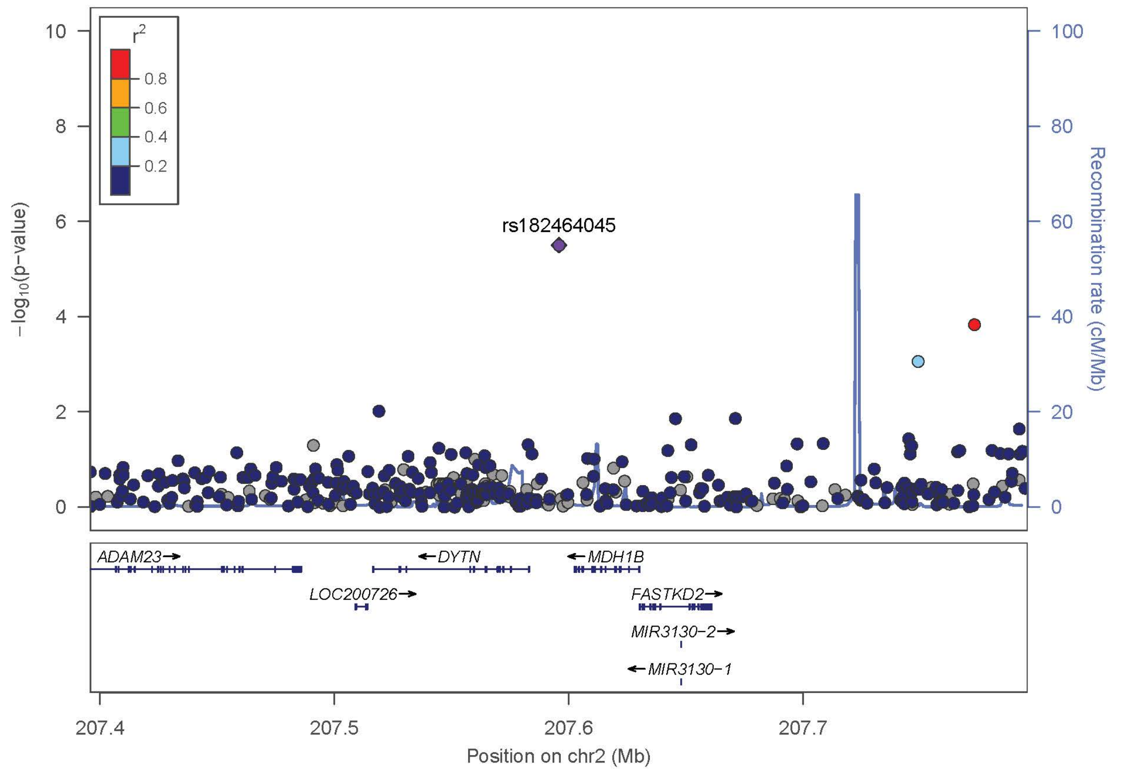

SNP rs182464045 (*DYTN/MDH1B*)

| Study | OR | 95%CI | Pvalue |
|---|---|---|---|
| - ADGC | 2.24 | [1.35,3.72] | 1.84E-03 |
| - GERAD | 3.20 | [1.12,9.13] | 2.94E-02 |
| - EADI | 2.47 | [1.49,4.10] | 1.84E-03 |
| - CHARGE | 3.29 | [0.50,21.54] | 2.14E-01 |
| - BONN | 1.03 | [0.08,12.57] | 9.82E-01 |
| **Stage1: IGAP** | **2.40** | **[1.66,3.47]** | **3.14E-06** |
| - EADB | 1.08 | [0.79,1.46] | 6.35E-01 |
| **Stage2: IGAP + EADB Meta** | **1.49** | **[1.18,1.89]** | **8.55E-04** |
| - UKBB | NA | [NA,NA] | NA |
| **Stage3: IGAP + EADB + UKBB Meta** | **1.49** | **[1.18,1.89]** | **8.55E-04** |

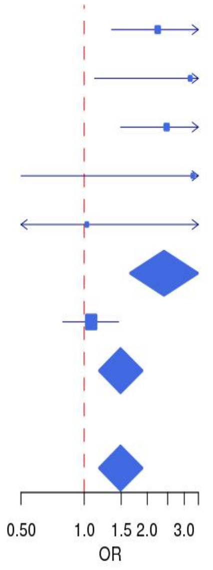

(q) LocusZoom and Forest plot of rs78774825 (*ZBTB10/ZNF704*) from IGAP meta-analysis (Stage1)

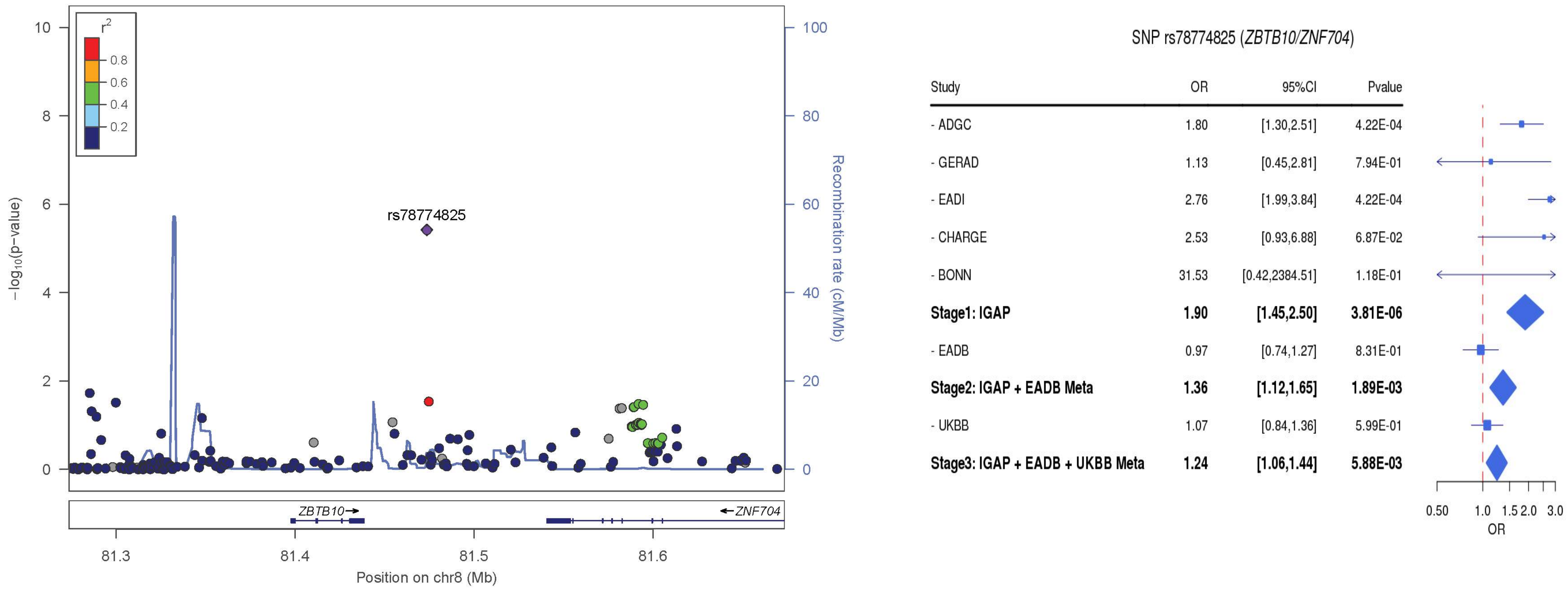

(r) LocusZoom and Forest plot of rs114999466 (*CNTNAP4*) from IGAP meta-analysis (Stage1)

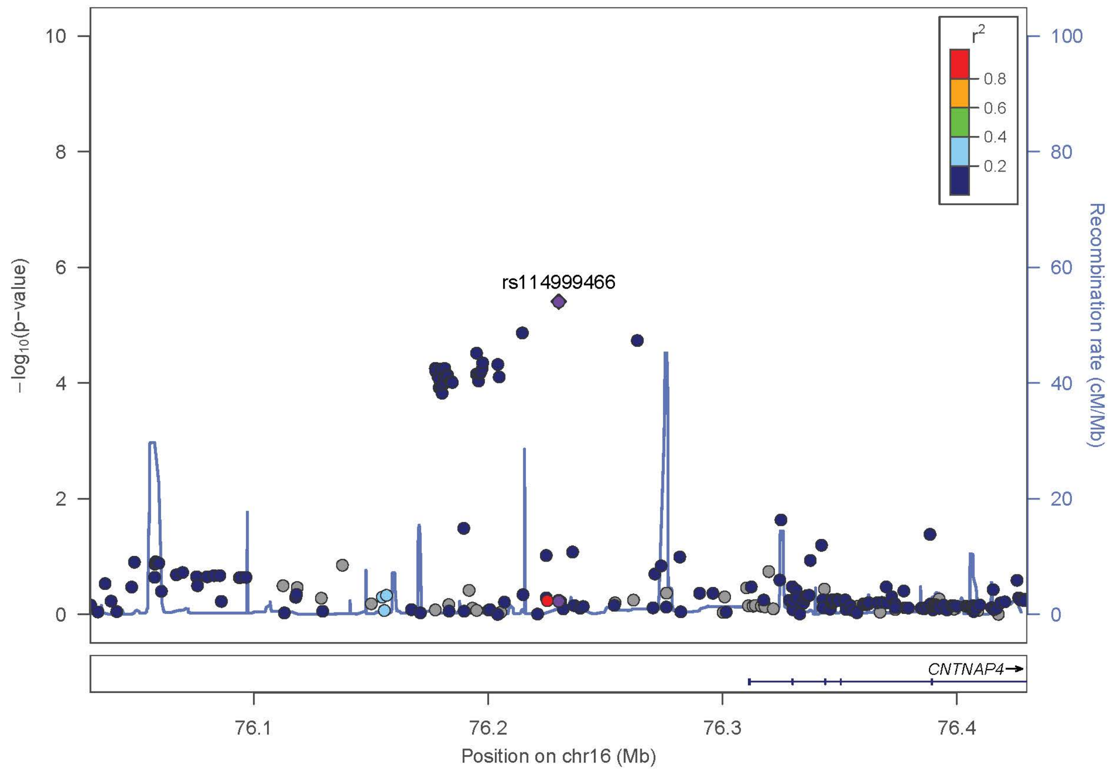

SNP rs114999466 (*CNTNAP4*)

| Study | OR | 95%CI | Pvalue |
|---|---|---|---|
| - ADGC | 0.68 | [0.56,0.84] | 2.09E-04 |
| - GERAD | 0.66 | [0.46,0.94] | 2.28E-02 |
| - EADI | 0.76 | [0.62,0.93] | 2.09E-04 |
| - CHARGE | 0.63 | [0.33,1.21] | 1.65E-01 |
| - BONN | 0.92 | [0.42,1.99] | 8.34E-01 |
| **Stage1: IGAP** | **0.69** | **[0.59,0.81]** | **3.91E-06** |
| - EADB | 0.82 | [0.70,0.96] | 1.14E-02 |
| **Stage2: IGAP + EADB Meta** | **1.33** | **[1.19,1.48]** | **4.51E-07** |
| - UKBB | 1.02 | [0.93,1.13] | 6.51E-01 |
| **Stage3: IGAP + EADB + UKBB Meta** | **1.12** | **[1.04,1.21]** | **2.52E-03** |

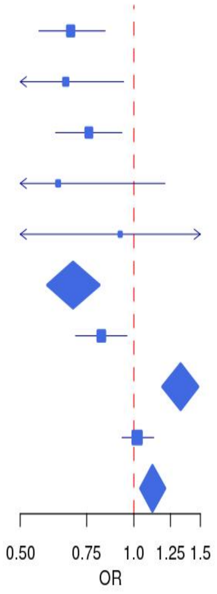

(s) LocusZoom and Forest plot of rs552075630 (*ODC1/NOL10*) from IGAP meta-analysis (Stage1)

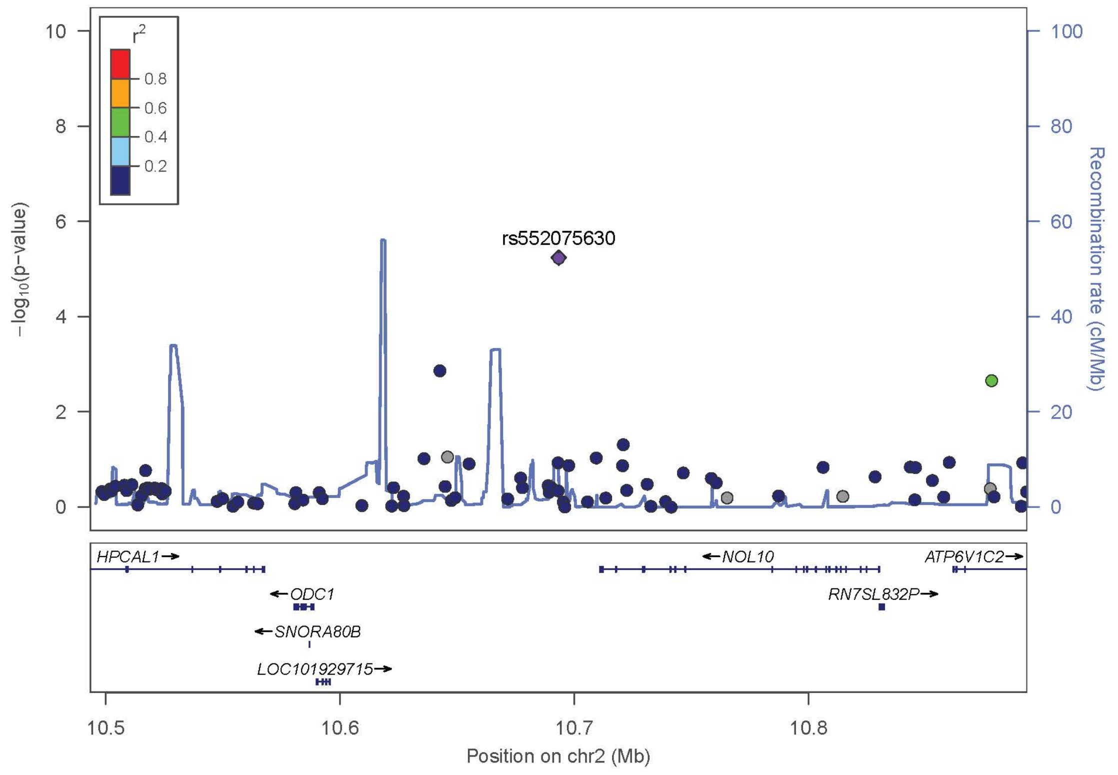

SNP rs552075630 (*ODC1/NOL10*)

| Study | OR | 95%CI | Pvalue |
|---|---|---|---|
| - ADGC | 1.44 | [1.10,1.89] | 7.67E-03 |
| - GERAD | 2.09 | [1.40,3.11] | 3.06E-04 |
| - EADI | 2.12 | [1.62,2.77] | 7.67E-03 |
| - CHARGE | 0.81 | [0.35,1.90] | 6.28E-01 |
| - BONN | 0.81 | [0.32,2.06] | 6.63E-01 |
| **Stage1: IGAP** | **1.57** | **[1.29,1.91]** | **5.82E-06** |
| - EADB | 1.07 | [0.86,1.33] | 5.25E-01 |
| **Stage2: IGAP + EADB Meta** | **1.33** | **[1.15,1.53]** | **1.44E-04** |
| - UKBB | 0.96 | [0.86,1.07] | 4.79E-01 |
| **Stage3: IGAP + EADB + UKBB Meta** | **1.08** | **[0.99,1.18]** | **8.36E-02** |

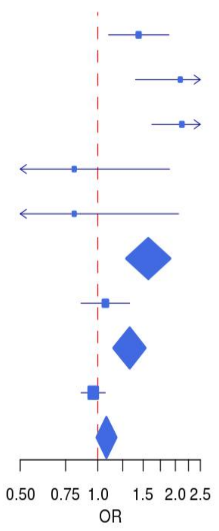

(t) LocusZoom and Forest plot of rs112120395 (*LINC00320*) from IGAP meta-analysis (Stage1)

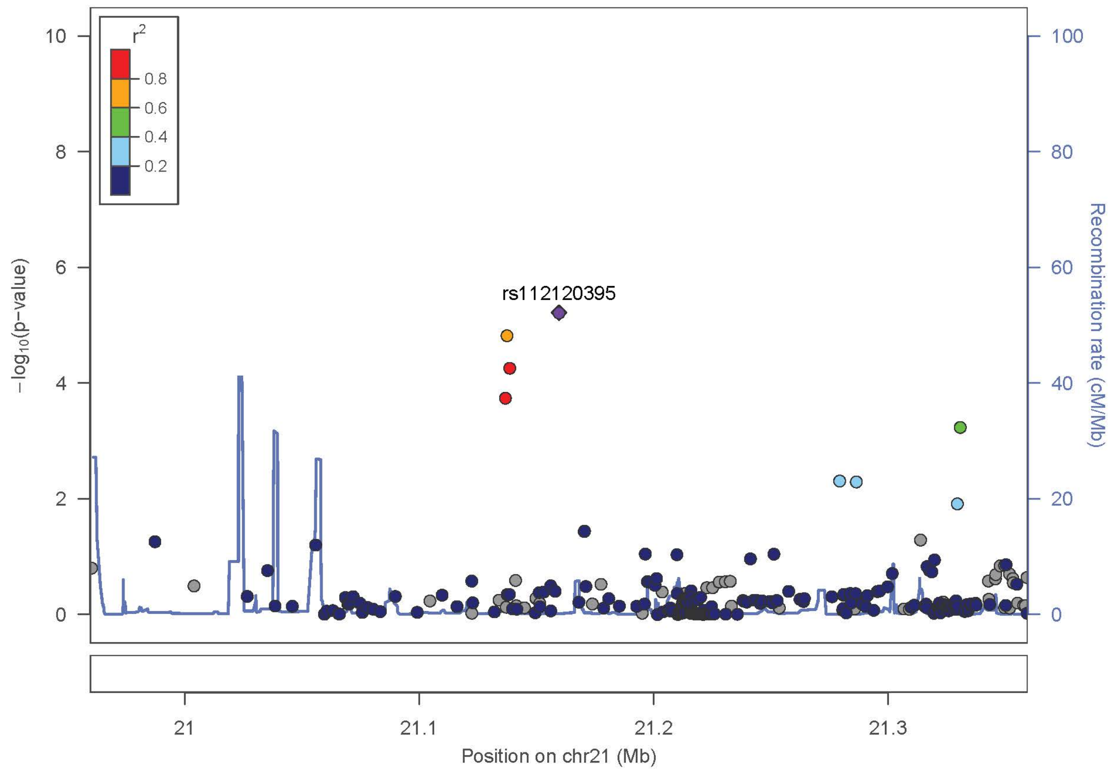

SNP rs112120395 (*LINC00320*)

| Study | OR | 95%CI | Pvalue |
|---|---|---|---|
| - ADGC | 1.38 | [1.09,1.73] | 7.10E-03 |
| - GERAD | 1.71 | [1.09,2.66] | 1.87E-02 |
| - EADI | 1.57 | [1.25,1.99] | 7.10E-03 |
| - CHARGE | 2.51 | [1.26,4.98] | 8.60E-03 |
| - BONN | 2.31 | [0.36,14.62] | 3.75E-01 |
| **Stage1: IGAP** | **1.52** | **[1.27,1.83]** | **6.09E-06** |
| - EADB | 0.90 | [0.74,1.09] | 2.67E-01 |
| **Stage2: IGAP + EADB Meta** | **0.84** | **[0.74,0.96]** | **1.07E-02** |
| - UKBB | 1.13 | [1.01,1.27] | 3.42E-02 |
| **Stage3: IGAP + EADB + UKBB Meta** | **0.87** | **[0.79,0.94]** | **1.08E-03** |

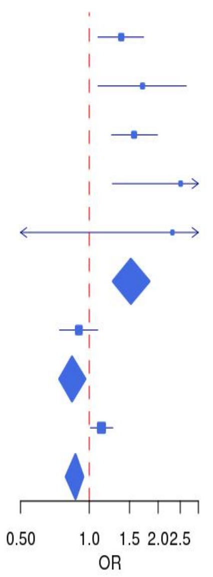

(u) LocusZoom and Forest plot of rs527488596 (*EPHA3*) from IGAP meta-analysis (Stage1)

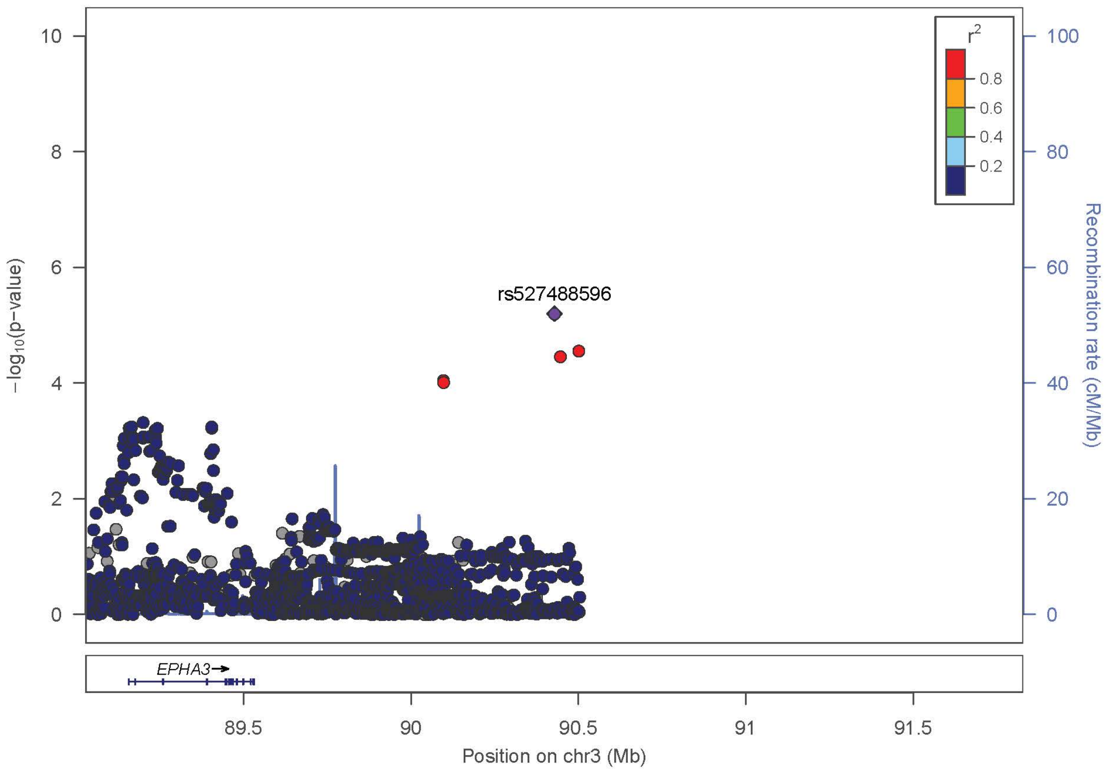

SNP rs527488596 (*EPHA3*)

| Study | OR | 95%CI | Pvalue |
|---|---|---|---|
| - ADGC | 0.67 | [0.52,0.87] | 2.91E-03 |
| - GERAD | 0.53 | [0.35,0.80] | 2.67E-03 |
| - EADI | 0.61 | [0.47,0.80] | 2.91E-03 |
| - CHARGE | 0.93 | [0.42,2.09] | 8.61E-01 |
| - BONN | 0.92 | [0.29,2.92] | 8.93E-01 |
| **Stage1: IGAP** | **0.65** | **[0.53,0.78]** | **6.32E-06** |
| - EADB | NA | [NA,NA] | NA |
| **Stage2: IGAP + EADB Meta** | **0.65** | **[0.53,0.78]** | **6.32E-06** |
| - UKBB | 0.98 | [0.88,1.10] | 7.84E-01 |
| **Stage3: IGAP + EADB + UKBB Meta** | **0.88** | **[0.80,0.97]** | **1.13E-02** |

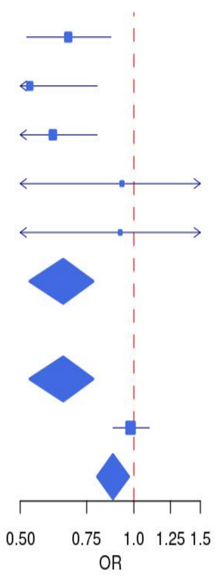

(v) LocusZoom and Forest plot of rs145472714 (*CCDC102B*) from IGAP meta-analysis (Stage1)

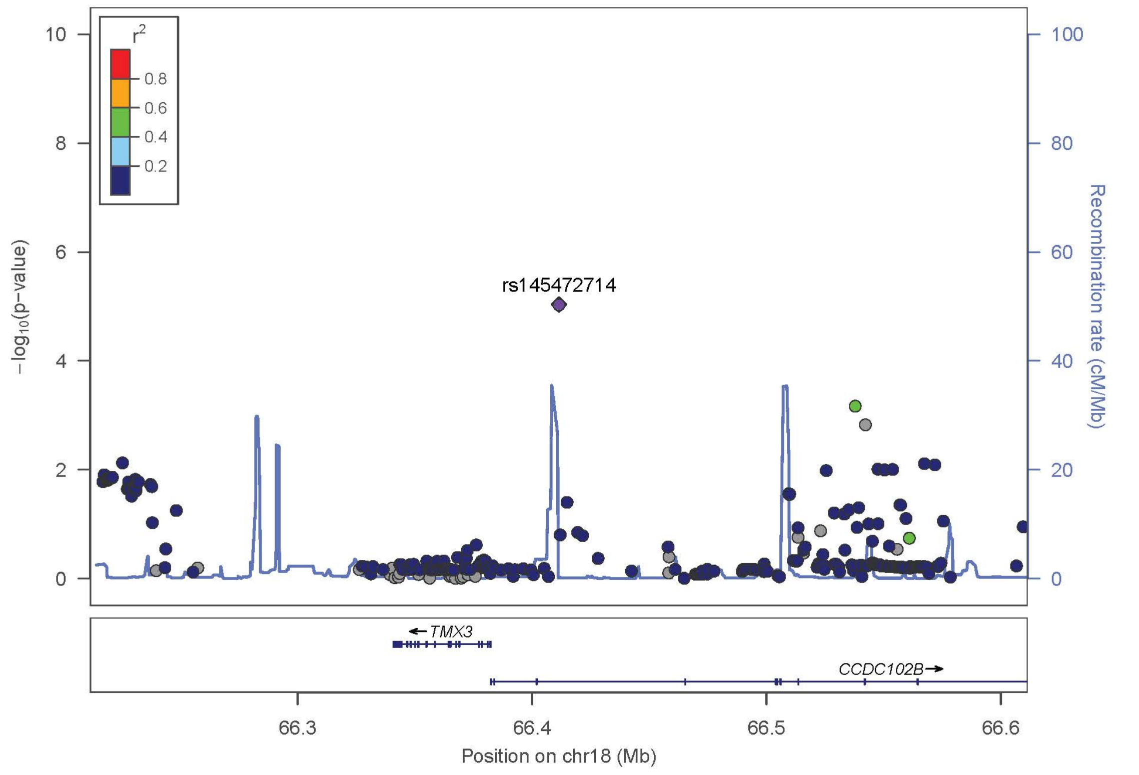

SNP rs145472714 (*CCDC102B*)

| Study | OR | 95%CI | Pvalue |
|---|---|---|---|
| - ADGC | 0.71 | [0.59,0.85] | 3.19E-04 |
| - GERAD | 0.82 | [0.48,1.40] | 4.73E-01 |
| - EADI | 0.64 | [0.53,0.78] | 3.19E-04 |
| - CHARGE | 0.56 | [0.33,0.98] | 4.17E-02 |
| - BONN | 0.86 | [0.15,4.82] | 8.65E-01 |
| **Stage1: IGAP** | **0.70** | **[0.60,0.82]** | **9.24E-06** |
| - EADB | 0.92 | [0.73,1.17] | 5.06E-01 |
| **Stage2: IGAP + EADB Meta** | **1.31** | **[1.15,1.49]** | **5.46E-05** |
| - UKBB | 1.03 | [0.89,1.19] | 7.05E-01 |
| **Stage3: IGAP + EADB + UKBB Meta** | **1.14** | **[1.04,1.26]** | **6.44E-03** |

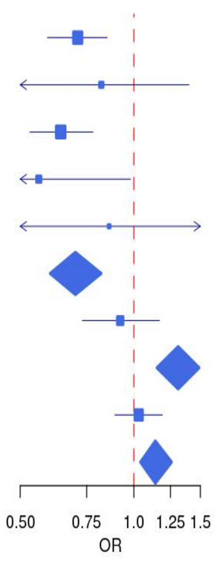

**Supplementary Figure 6.** Tissue-/cell-type-specific expression patterns of *NCK2* taken from the BioGPS Portal (http://biogps.org/#goto=genereport&id=8440).

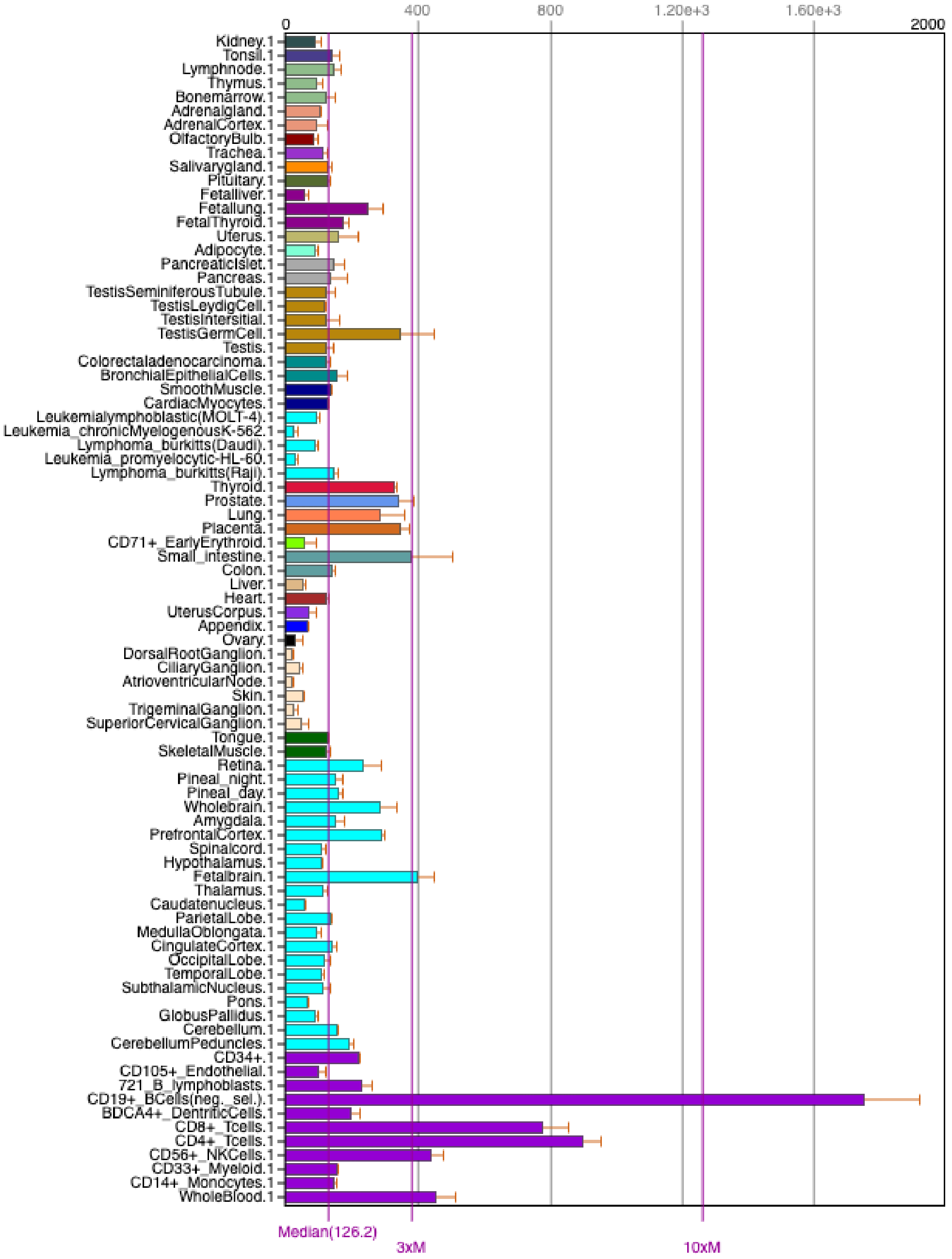

**Supplementary Figure 7.** Diagram of chromatin interactions ±50kb of the *NCK2* SNP rs143080277 on chromosome 2q12.2. The position of rs143080277 is indicated with the blue line and falls within physically-interacting regions only in the replicates of the GM12878 cell line.

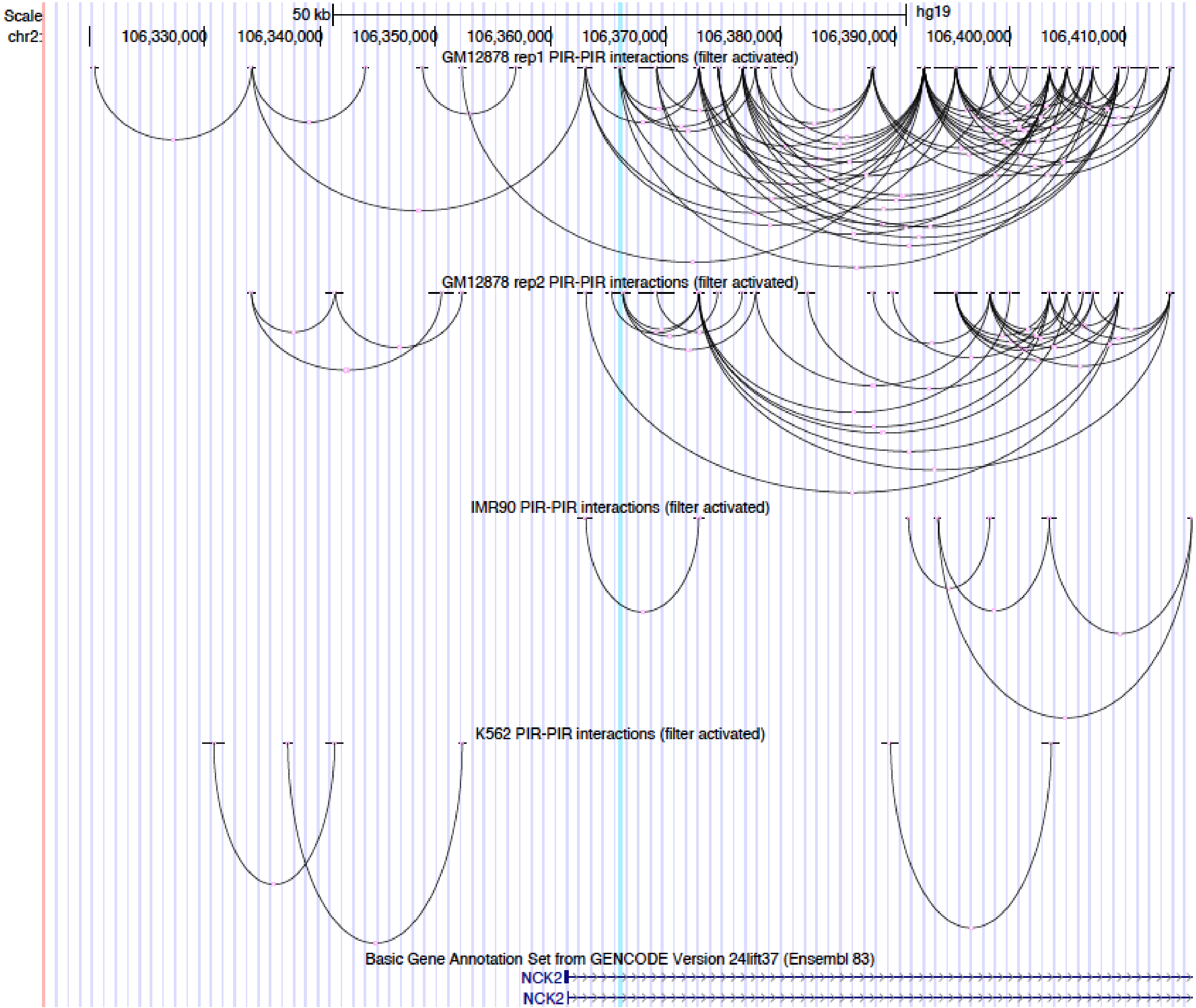

**Supplementary Figure 8.** UCSC Genome Browser session showing ATAC-seq OCRs (black) and Capture C chromatin loops (magenta) near variants of interest (blue) in TFH cells[98]. (a) rs143080277 resides in an OCR and contacts the promoters a short isoform of *UXS1* and a short isoform of *NCK2* (both in red). (b) rs144636993 contacts an alternative promoter of the same short isoform of *NCK2* (red) in ESC-derived hypothalamic neurons[99], however neither the SNP nor the promoter reside in an OCR.

**(a)** Interactions with rs143080277

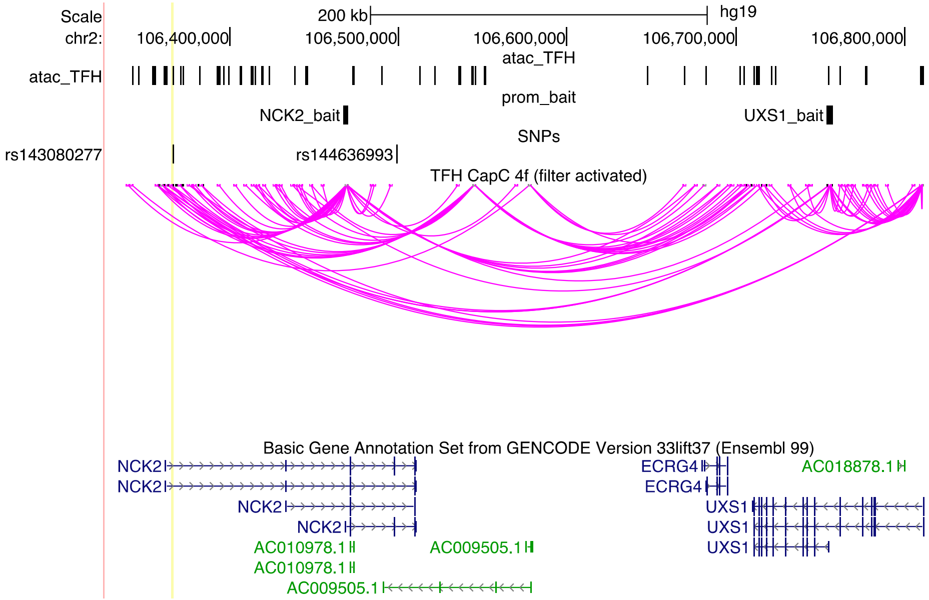

**(b)** Interactions with rs144636993

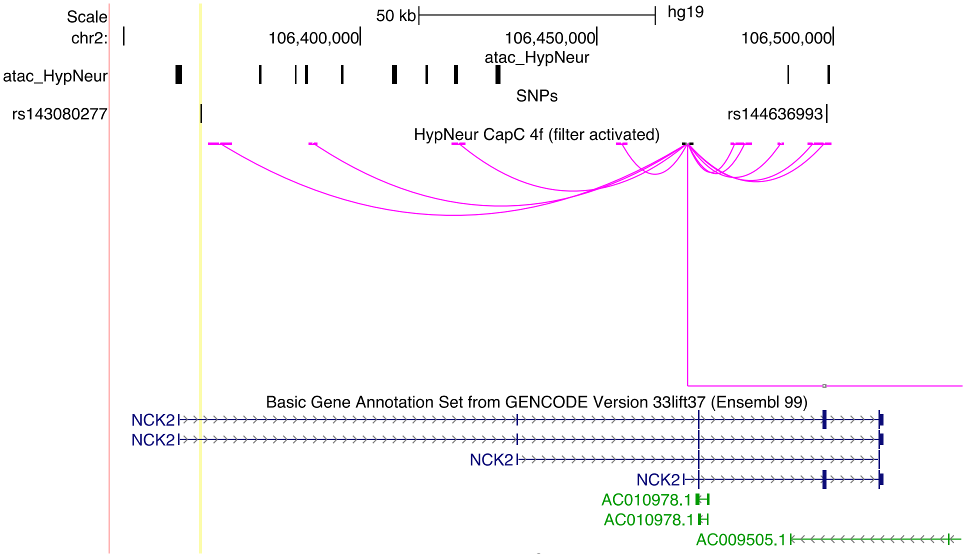

**Supplementary Figure 9.** GeneMANIA[100,101] network diagram showing the predicted functional association network of *NCK2*.

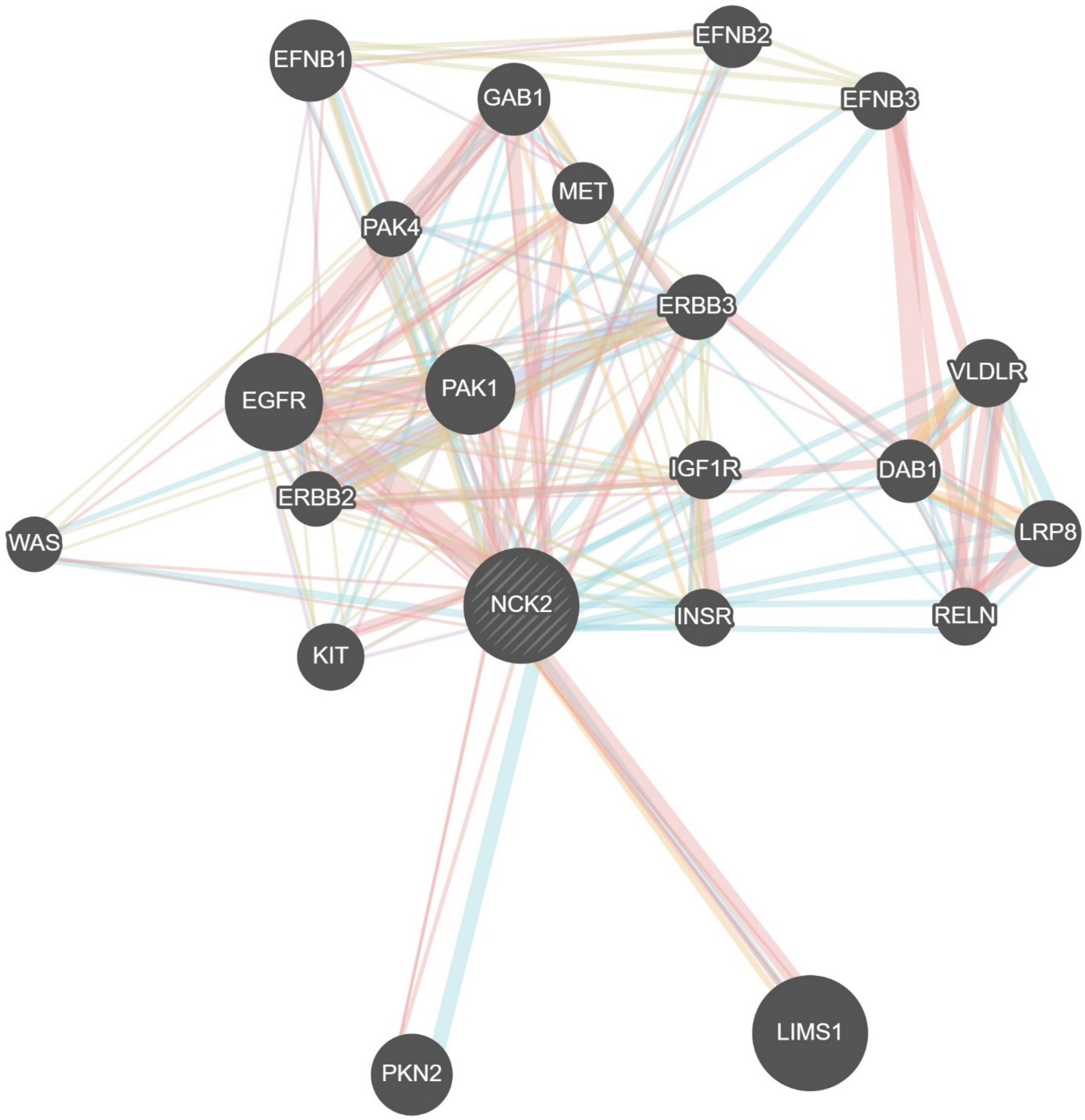

### References

1. Naj, A.C. *et al.* Common variants at MS4A4/MS4A6E, CD2AP, CD33 and EPHA1 are associated with late-onset Alzheimer's disease. *Nat Genet* **43**, 436-41 (2011).
2. Lambert, J.C. *et al.* Meta-analysis of 74,046 individuals identifies 11 new susceptibility loci for Alzheimer's disease. *Nat Genet* **45**, 1452-8 (2013).
3. Sims, R. *et al.* Rare coding variants in PLCG2, ABI3, and TREM2 implicate microglial-mediated innate immunity in Alzheimer's disease. *Nat Genet* **49**, 1373-1384 (2017).
4. Jun, G. *et al.* A novel Alzheimer disease locus located near the gene encoding tau protein. *Mol Psychiatry* **21**, 108-17 (2016).
5. Jun, G. *et al.* Meta-analysis confirms CR1, CLU, and PICALM as alzheimer disease risk loci and reveals interactions with APOE genotypes. *Arch Neurol* **67**, 1473-84 (2010).
6. Kunkle, B.W. *et al.* Genetic meta-analysis of diagnosed Alzheimer's disease identifies new risk loci and implicates Abeta, tau, immunity and lipid processing. *Nat Genet* **51**, 414-430 (2019).
7. Kukull, W.A. *et al.* Dementia and Alzheimer disease incidence: a prospective cohort study. *Arch Neurol* **59**, 1737-46 (2002).
8. Larson, E.B. *et al.* Exercise is associated with reduced risk for incident dementia among persons 65 years of age and older. *Ann Intern Med* **144**, 73-81 (2006).
9. Beekly, D.L. *et al.* The National Alzheimer's Coordinating Center (NACC) database: the Uniform Data Set. *Alzheimer Dis Assoc Disord* **21**, 249-58 (2007).
10. Morris, J.C. *et al.* The Uniform Data Set (UDS): clinical and cognitive variables and descriptive data from Alzheimer Disease Centers. *Alzheimer Dis Assoc Disord* **20**, 210-6 (2006).
11. McKhann, G. *et al.* Clinical diagnosis of Alzheimer's disease: report of the NINCDS-ADRDA Work Group under the auspices of Department of Health and Human Services Task Force on Alzheimer's Disease. *Neurology* **34**, 939-44 (1984).
12. McKhann, G.M. *et al.* The diagnosis of dementia due to Alzheimer's disease: recommendations from the National Institute on Aging-Alzheimer's Association workgroups on diagnostic guidelines for Alzheimer's disease. *Alzheimers Dement* **7**, 263-9 (2011).
13. Mirra, S.S., Hart, M.N. & Terry, R.D. Making the diagnosis of Alzheimer's disease. A primer for practicing pathologists. *Arch Pathol Lab Med* **117**, 132-44 (1993).
14. Nagy, Z. *et al.* Assessment of the pathological stages of Alzheimer's disease in thin paraffin sections: a comparative study. *Dement Geriatr Cogn Disord* **9**, 140-4 (1998).
15. Braak, H. & Braak, E. Neuropathological stageing of Alzheimer-related changes. *Acta Neuropathol* **82**, 239-59 (1991).
16. Petersen, R.C. *et al.* Alzheimer's Disease Neuroimaging Initiative (ADNI): clinical characterization. *Neurology* **74**, 201-9 (2010).
17. Saykin, A.J. *et al.* Alzheimer's Disease Neuroimaging Initiative biomarkers as quantitative phenotypes: Genetics core aims, progress, and plans. *Alzheimers Dement* **6**, 265-73 (2010).
18. Albert, M. *et al.* Cognitive changes preceding clinical symptom onset of mild cognitive impairment and relationship to ApoE genotype. *Curr Alzheimer Res* **11**, 773-84 (2014).
19. Bienias, J.L., Beckett, L.A., Bennett, D.A., Wilson, R.S. & Evans, D.A. Design of the Chicago Health and Aging Project (CHAP). *J Alzheimers Dis* **5**, 349-55 (2003).

20. Barzilai, N., Rossetti, L. & Lipton, R.B. Einstein's Institute for Aging Research: collaborative and programmatic approaches in the search for successful aging. *Exp Gerontol* **39**, 151-7 (2004).
21. Katz, M.J. *et al.* Age-specific and sex-specific prevalence and incidence of mild cognitive impairment, dementia, and Alzheimer dementia in blacks and whites: a report from the Einstein Aging Study. *Alzheimer Dis Assoc Disord* **26**, 335-43 (2012).
22. Li, H. *et al.* Candidate single-nucleotide polymorphisms from a genomewide association study of Alzheimer disease. *Arch Neurol* **65**, 45-53 (2008).
23. Green, R.C. *et al.* Risk of dementia among white and African American relatives of patients with Alzheimer disease. *JAMA* **287**, 329-36 (2002).
24. Roccaforte, W.H., Burke, W.J., Bayer, B.L. & Wengel, S.P. Validation of a telephone version of the mini-mental state examination. *J Am Geriatr Soc* **40**, 697-702 (1992).
25. Lee, J.H. *et al.* Analyses of the National Institute on Aging Late-Onset Alzheimer's Disease Family Study: implication of additional loci. *Arch Neurol* **65**, 1518-26 (2008).
26. Ravid, R. & Swaab, D.F. The Netherlands brain bank--a clinico-pathological link in aging and dementia research. *J Neural Transm Suppl* **39**, 143-53 (1993).
27. Kramer, P.L. *et al.* Alzheimer disease pathology in cognitively healthy elderly: a genome-wide study. *Neurobiol Aging* **32**, 2113-22 (2011).
28. Bennett, D.A. *et al.* The Rush Memory and Aging Project: study design and baseline characteristics of the study cohort. *Neuroepidemiology* **25**, 163-75 (2005).
29. Bennett, D.A. *et al.* Natural history of mild cognitive impairment in older persons. *Neurology* **59**, 198-205 (2002).
30. Bennett, D.A., Schneider, J.A., Bienias, J.L., Evans, D.A. & Wilson, R.S. Mild cognitive impairment is related to Alzheimer disease pathology and cerebral infarctions. *Neurology* **64**, 834-41 (2005).
31. Hall, J.R. *et al.* Biomarkers of vascular risk, systemic inflammation, and microvascular pathology and neuropsychiatric symptoms in Alzheimer's disease. *J Alzheimers Dis* **35**, 363-71 (2013).
32. Reiman, E.M. *et al.* GAB2 alleles modify Alzheimer's risk in APOE epsilon4 carriers. *Neuron* **54**, 713-20 (2007).
33. Caselli, R.J. *et al.* Cognitive domain decline in healthy apolipoprotein E epsilon4 homozygotes before the diagnosis of mild cognitive impairment. *Arch Neurol* **64**, 1306-11 (2007).
34. Webster, J.A. *et al.* Genetic control of human brain transcript expression in Alzheimer disease. *Am J Hum Genet* **84**, 445-58 (2009).
35. Petyuk, V.A. *et al.* The human brainome: network analysis identifies HSPA2 as a novel Alzheimer’s disease target. *Brain* **141**, 2721-2739 (2018).
36. Scott, W.K. *et al.* Complete genomic screen in Parkinson disease: evidence for multiple genes. *JAMA* **286**, 2239-44 (2001).
37. Beecham, G.W. *et al.* Genome-wide association study implicates a chromosome 12 risk locus for late-onset Alzheimer disease. *Am J Hum Genet* **84**, 35-43 (2009).
38. Edwards, T.L. *et al.* Genome-wide association study confirms SNPs in SNCA and the MAPT region as common risk factors for Parkinson disease. *Ann Hum Genet* **74**, 97-109 (2010).

39. Naj, A.C. *et al.* Dementia revealed: novel chromosome 6 locus for late-onset Alzheimer disease provides genetic evidence for folate-pathway abnormalities. *PLoS Genet* **6**, e1001130 (2010).
40. Haroutunian, V. *et al.* Regional distribution of neuritic plaques in the nondemented elderly and subjects with very mild Alzheimer disease. *Arch Neurol* **55**, 1185-91 (1998).
41. Kamboh, M.I. *et al.* Association of CLU and PICALM variants with Alzheimer's disease. *Neurobiol Aging* **33**, 518-21 (2012).
42. Hughes, C.P., Berg, L., Danziger, W.L., Coben, L.A. & Martin, R.L. A new clinical scale for the staging of dementia. *Br J Psychiatry* **140**, 566-72 (1982).
43. Tang, M.X. *et al.* Incidence of AD in African-Americans, Caribbean Hispanics, and Caucasians in northern Manhattan. *Neurology* **56**, 49-56 (2001).
44. Mayeux, R., Small, S.A., Tang, M., Tycko, B. & Stern, Y. Memory performance in healthy elderly without Alzheimer's disease: effects of time and apolipoprotein-E. *Neurobiol Aging* **22**, 683-9 (2001).
45. Price, A.L. *et al.* Principal components analysis corrects for stratification in genome-wide association studies. *Nat Genet* **38**, 904-9 (2006).
46. Patterson, N., Price, A.L. & Reich, D. Population structure and eigenanalysis. *PLoS Genet* **2**, e190 (2006).
47. Psaty, B.M. *et al.* Cohorts for Heart and Aging Research in Genomic Epidemiology (CHARGE) Consortium: Design of prospective meta-analyses of genome-wide association studies from 5 cohorts. *Circ Cardiovasc Genet* **2**, 73-80 (2009).
48. Ikram, M.A. *et al.* Genomewide association studies of stroke. *N Engl J Med* **360**, 1718-28 (2009).
49. The Atherosclerosis Risk in Communities (ARIC) Study: design and objectives. The ARIC investigators. *Am J Epidemiol* **129**, 687-702 (1989).
50. Fried, L.P. *et al.* The Cardiovascular Health Study: design and rationale. *Ann Epidemiol* **1**, 263-76 (1991).
51. Fitzpatrick, A.L. *et al.* Incidence and prevalence of dementia in the Cardiovascular Health Study. *J Am Geriatr Soc* **52**, 195-204 (2004).
52. Lopez, O.L. *et al.* Evaluation of dementia in the cardiovascular health cognition study. *Neuroepidemiology* **22**, 1-12 (2003).
53. Dawber, T.R. & Kannel, W.B. The Framingham study. An epidemiological approach to coronary heart disease. *Circulation* **34**, 553-5 (1966).
54. Feinleib, M., Kannel, W.B., Garrison, R.J., McNamara, P.M. & Castelli, W.P. The Framingham Offspring Study. Design and preliminary data. *Prev Med* **4**, 518-25 (1975).
55. Splansky, G.L. *et al.* The Third Generation Cohort of the National Heart, Lung, and Blood Institute's Framingham Heart Study: design, recruitment, and initial examination. *Am J Epidemiol* **165**, 1328-35 (2007).
56. Beiser, A., D'Agostino, R.B., Sr., Seshadri, S., Sullivan, L.M. & Wolf, P.A. Computing estimates of incidence, including lifetime risk: Alzheimer's disease in the Framingham Study. The Practical Incidence Estimators (PIE) macro. *Stat Med* **19**, 1495-522 (2000).
57. Bachman, D.L. *et al.* Incidence of dementia and probable Alzheimer's disease in a general population: the Framingham Study. *Neurology* **43**, 515-9 (1993).
58. Farmer, M.E. *et al.* Neuropsychological test performance in Framingham: a descriptive study. *Psychol Rep* **60**, 1023-40 (1987).

59. DeCarli, C. *et al.* Measures of brain morphology and infarction in the framingham heart study: establishing what is normal. *Neurobiol Aging* **26**, 491-510 (2005).
60. Au, R. *et al.* New norms for a new generation: cognitive performance in the framingham offspring cohort. *Exp Aging Res* **30**, 333-58 (2004).
61. Seshadri, S. *et al.* Lifetime risk of dementia and Alzheimer's disease. The impact of mortality on risk estimates in the Framingham Study. *Neurology* **49**, 1498-504 (1997).
62. Hofman, A. *et al.* The Rotterdam Study: 2010 objectives and design update. *Eur J Epidemiol* **24**, 553-72 (2009).
63. Hofman, A. *et al.* The Rotterdam Study: objectives and design update. *Eur J Epidemiol* **22**, 819-29 (2007).
64. Ott, A., Breteler, M.M., van Harskamp, F., Stijnen, T. & Hofman, A. Incidence and risk of dementia. The Rotterdam Study. *Am J Epidemiol* **147**, 574-80 (1998).
65. Dreses-Werringloer, U. *et al.* A polymorphism in CALHM1 influences Ca2+ homeostasis, Abeta levels, and Alzheimer's disease risk. *Cell* **133**, 1149-61 (2008).
66. 3C Study Group. Vascular factors and risk of dementia: design of the Three-City Study and baseline characteristics of the study population. *Neuroepidemiology* **22**, 316-25 (2003).
67. Harold, D. *et al.* Genome-wide association study identifies variants at CLU and PICALM associated with Alzheimer's disease. *Nat Genet* **41**, 1088-93 (2009).
68. Bellenguez, C. *et al.* New insights on the genetic etiology of Alzheimer's and related dementia. *medRxiv*, 2020.10.01.20200659 (2020).
69. De Roeck, A. *et al.* An intronic VNTR affects splicing of ABCA7 and increases risk of Alzheimer's disease. *Acta Neuropathol* **135**, 827-837 (2018).
70. Sheardova, K. *et al.* Czech Brain Aging Study (CBAS): prospective multicentre cohort study on risk and protective factors for dementia in the Czech Republic. *BMJ Open* **9**, e030379 (2019).
71. Steinberg, S. *et al.* Loss-of-function variants in ABCA7 confer risk of Alzheimer's disease. *Nat Genet* **47**, 445-7 (2015).
72. Ngandu, T. *et al.* A 2 year multidomain intervention of diet, exercise, cognitive training, and vascular risk monitoring versus control to prevent cognitive decline in at-risk elderly people (FINGER): a randomised controlled trial. *Lancet* **385**, 2255-63 (2015).
73. Hanon, O. *et al.* Plasma amyloid levels within the Alzheimer's process and correlations with central biomarkers. *Alzheimers Dement* **14**, 858-868 (2018).
74. Dufouil, C. *et al.* Cognitive and imaging markers in non-demented subjects attending a memory clinic: study design and baseline findings of the MEMENTO cohort. *Alzheimers Res Ther* **9**, 67 (2017).
75. Nicolas, G. *et al.* Screening of dementia genes by whole-exome sequencing in early-onset Alzheimer disease: input and lessons. *Eur J Hum Genet* **24**, 710-6 (2016).
76. Kornhuber, J. *et al.* Early and differential diagnosis of dementia and mild cognitive impairment: design and cohort baseline characteristics of the German Dementia Competence Network. *Dement Geriatr Cogn Disord* **27**, 404-17 (2009).
77. Luck, T. *et al.* Mild cognitive impairment in general practice: age-specific prevalence and correlate results from the German study on ageing, cognition and dementia in primary care patients (AgeCoDe). *Dement Geriatr Cogn Disord* **24**, 307-16 (2007).
78. van der Flier, W.M. & Scheltens, P. Amsterdam Dementia Cohort: Performing Research to Optimize Care. *J Alzheimers Dis* **62**, 1091-1111 (2018).

79. Petersen, R.C. *et al.* Mild cognitive impairment: clinical characterization and outcome. *Arch Neurol* **56**, 303-8 (1999).
80. Holstege, H. *et al.* The 100-plus Study of cognitively healthy centenarians: rationale, design and cohort description. *Eur J Epidemiol* **33**, 1229-1249 (2018).
81. Aalten, P. *et al.* The Dutch Parelsnoer Institute--Neurodegenerative diseases; methods, design and baseline results. *BMC Neurol* **14**, 254 (2014).
82. Dubois, B. *et al.* Research criteria for the diagnosis of Alzheimer's disease: revising the NINCDS-ADRDA criteria. *Lancet Neurol* **6**, 734-46 (2007).
83. Ramakers, I.H.G.B. *et al.* Biobank Alzheimer Center Limburg cohort: design and cohort characteristics. *(In Preparation)*.
84. American Psychiatric Association. *Diagnostic and statistical manual of mental disorders: DSM-IV-TR*, (Washington, DC, 2000).
85. Sachdev, P.S. *et al.* The Sydney Memory and Ageing Study (MAS): methodology and baseline medical and neuropsychiatric characteristics of an elderly epidemiological non-demented cohort of Australians aged 70-90 years. *Int Psychogeriatr* **22**, 1248-64 (2010).
86. Bycroft, C. *et al.* The UK Biobank resource with deep phenotyping and genomic data. *Nature* **562**, 203-209 (2018).
87. Grove, M.L. *et al.* Best practices and joint calling of the HumanExome BeadChip: the CHARGE Consortium. *PLoS One* **8**, e68095 (2013).
88. Purcell, S. *et al.* PLINK: a tool set for whole-genome association and population-based linkage analyses. *Am J Hum Genet* **81**, 559-75 (2007).
89. Chang, C.C. *et al.* Second-generation PLINK: rising to the challenge of larger and richer datasets. *Gigascience* **4**, 7 (2015).
90. Chang, C.C. Data Management and Summary Statistics with PLINK. *Methods Mol Biol* **2090**, 49-65 (2020).
91. Abraham, G., Qiu, Y. & Inouye, M. FlashPCA2: principal component analysis of Biobank-scale genotype datasets. *Bioinformatics* **33**, 2776-2778 (2017).
92. Gogarten, S.M. *et al.* Genetic association testing using the GENESIS R/Bioconductor package. *Bioinformatics* **35**, 5346-5348 (2019).
93. McCarthy, S. *et al.* A reference panel of 64,976 haplotypes for genotype imputation. *Nat Genet* **48**, 1279-83 (2016).
94. Karczewski, K.J. *et al.* The mutational constraint spectrum quantified from variation in 141,456 humans. *Nature* **581**, 434-443 (2020).
95. Marchini, J., Howie, B., Myers, S., McVean, G. & Donnelly, P. A new multipoint method for genome-wide association studies by imputation of genotypes. *Nat Genet* **39**, 906-13 (2007).
96. Das, S. *et al.* Next-generation genotype imputation service and methods. *Nat Genet* **48**, 1284-1287 (2016).
97. Loh, P.R. *et al.* Reference-based phasing using the Haplotype Reference Consortium panel. *Nat Genet* **48**, 1443-1448 (2016).
98. Su, C. *et al.* Mapping effector genes at lupus GWAS loci using promoter Capture-C in follicular helper T cells. *Nat Commun* **11**, 3294 (2020).
99. Lasconi, C. *et al.* Variant-to-Gene-Mapping Analyses Reveal a Role for the Hypothalamus in Genetic Susceptibility to Inflammatory Bowel Disease. *Cell Mol Gastroenterol Hepatol* (2020).

100. Mostafavi, S., Ray, D., Warde-Farley, D., Grouios, C. & Morris, Q. GeneMANIA: a real-time multiple association network integration algorithm for predicting gene function. *Genome Biol* **9** **Suppl 1**, S4 (2008).
101. Warde-Farley, D. *et al.* The GeneMANIA prediction server: biological network integration for gene prioritization and predicting gene function. *Nucleic Acids Res* **38**, W214-20 (2010).
